# Effects of prenatal small-quantity lipid-based nutrient supplements on pregnancy, birth and infant outcomes: a systematic review and meta-analysis of individual participant data from randomized controlled trials in low- and middle-income countries

**DOI:** 10.1101/2024.05.17.24307546

**Authors:** Kathryn G. Dewey, K. Ryan Wessells, Charles D. Arnold, Seth Adu-Afarwuah, Benjamin F. Arnold, Ulla Ashorn, Per Ashorn, Ana Garcés, Lieven Huybregts, Nancy F. Krebs, Anna Lartey, Jef L. Leroy, Kenneth Maleta, Susana L. Matias, Sophie E. Moore, Malay K. Mridha, Harriet Okronipa, Christine P. Stewart

**Author notes:** Corresponding Author: Kathryn G. Dewey, Department of Nutrition, University of California, One Shields Ave., Davis, CA 95616. Sources of Support: Bill & Melinda Gates Foundation OPP49817. The funder had no role in the design, implementation, analysis or interpretation of the data. Registry and registry number for systematic reviews or meta-analyses: Registered at www.crd.york.ac.uk/PROSPERO as CRD42021283391 on 11 April 2021. Data described in the manuscript, code book, and analytic code will not be made available because they are compiled from 4 different trials, and access is under the control of the investigators of each of those trials. Dr. Nancy Krebs is on the Editorial Board of the American Journal of Clinical Nutrition and played no role in the Journal’s evaluation of the manuscript.

## Abstract

**Background:** Undernutrition during pregnancy increases the risk of giving birth to a small vulnerable newborn. Small-quantity lipid-based nutrient supplements (SQ-LNS) contain both macro- and micronutrients and can help prevent nutritional deficiencies during pregnancy and lactation.

**Objectives:** We examined effects of SQ-LNS provided to pregnant women, compared to a) iron and folic acid or standard of care (IFA/SOC) or b) multiple micronutrient supplements (MMS), and identified characteristics that modified effects of SQ-LNS on birth outcomes.

**Methods:** We conducted a 2-stage meta-analysis of individual participant data from 4 randomized controlled trials of SQ-LNS provided to pregnant women (n = 5,273). We generated study-specific and subgroup estimates of SQ-LNS compared with IFA/SOC or MMS and pooled the estimates. In sensitivity analyses, we examined whether results differed depending on methods for gestational age dating, birth anthropometry, or study design.

**Results:** SQ-LNS (vs IFA/SOC) increased birth weight (mean difference: +49g; 95% CI: 26, 71g), duration of gestation (+0.12 wk; 95% CI: 0.01, 0.24 wk), and all birth anthropometric z-scores (+0.10-0.13 SD); it reduced risk of low birthweight by 11%, newborn stunting by 17%, newborn wasting by 11%, and small head size by 11%. Only 2 trials compared SQ-LNS and MMS; birth outcomes did not differ except for a marginal increase in head circumference for gestational age (+0.11; 95% CI: -0.01, 0.23). Effect estimates for SQ-LNS vs IFA/SOC were greater among female infants and among women with body mass index < 20 kg/m^2^, inflammation, malaria, or household food insecurity. Effect estimates for SQ-LNS vs MMS were greater among female infants, first-born infants, and women < 25 y.

**Conclusions:** SQ-LNS had positive impacts on multiple outcomes compared to IFA/SOC, but further research directly comparing SQ-LNS and MMS is needed. Targeting SQ-LNS to vulnerable subgroups may be worth considering. Analysis registered at www.crd.york.ac.uk/PROSPERO (CRD42021283391).

## Introduction

Undernutrition is prevalent among women of reproductive age globally, with an estimated 1.2 billion deficient in one or more micronutrients (1), 571 million (30%) with anemia (2–4), and 170 million (∼10%) being underweight (3, 4). As a result, many women and girls enter pregnancy with nutritional deficits. The risk of undernutrition during pregnancy is exacerbated by the elevated nutrient needs to support gestation, particularly in low- and middle-income countries (LMICs) where diets are often inadequate in multiple nutrients (5). This situation contributes to poor maternal health and the risk of giving birth to a small vulnerable newborn (SVN), an umbrella term that encompasses small-for-gestational age (SGA), preterm birth, and low birthweight (LBW) (6). In 2020, 26.2% of all livebirths globally were SVNs, with 16.3% SGA, 8.8% preterm, and 1.1% both SGA and preterm (7). Although poor nutrition is not the only cause of these outcomes (8), interventions to improve maternal nutrition, such as multiple micronutrient supplements (MMS) and balanced energy-protein supplementation (BEP) for undernourished mothers, should be considered critical elements of antenatal care packages aimed at reducing SVN births (9).

Lipid-based nutrient supplements (LNS) provide multiple micronutrients embedded in a food base that also provides energy, protein and fat. LNS for pregnant women are a type of BEP, as they meet the criterion that protein contributes < 25% of the energy content (10). The intended daily ration of LNS can be small-medium- or large-quantity (SQ-, MQ- or LQ-LNS) (11). Maternal SQ-LNS was designed to fill nutrient gaps during pregnancy and the first 6 mo postpartum with a daily ration of only 20 g (118 kcal/d) (11), which minimizes cost and potential displacement of home-prepared foods. SQ-LNS is not primarily designed to fill energy gaps, as such gaps can be filled by more affordable local foods (11). SQ-LNS provides several key nutrients not provided by iron and folic acid (IFA) supplements or MMS, including essential fatty acids (EFAs) as well as calcium, magnesium, potassium, and phosphorus (11). This combination of macro- and micro-nutrients addresses multiple potential nutritional deficiencies and thus can reduce maternal undernutrition. Maternal SQ-LNS is currently being distributed by USAID in selected food aid programs (12), but its use is not yet widespread.

A previous meta-analysis of maternal LNS (13) demonstrated that LNS given during pregnancy, compared to IFA, had positive effects on birth weight, length, duration of gestation, SGA, and newborn stunting. That meta-analysis included 3 trials, all of which used SQ-LNS. In a separate comparison of LNS vs MMS that also included 3 trials (2 SQ-LNS, 1 MQ-LNS), there were no significant differences in birth outcomes (13). More recently, Hunter et al. (14) conducted a meta-analysis of maternal LNS vs MMS that included 4 trials (2 SQ-LNS, 1 MQ-LNS, 1 LQ-LNS); they found a significant reduction in LBW but not preterm birth or SGA. The authors of these previous meta-analyses did not have individual participant data (IPD) and thus were not able to examine individual-level effect modification.

Effect modification analysis can provide important insights regarding the potential for participants to benefit from an intervention, as well as their potential to respond (15). These 2 concepts reflect different attributes, with potential to benefit most likely related to greater deficits at baseline, and potential to respond related to lack of constraints on exhibiting an improvement in the outcome due to factors such as infection or inflammation. In some cases, individuals with greater potential to benefit may also have a lower potential to respond, which can limit the beneficial impact of an intervention. Identification of subgroups of pregnant women with the greatest potential to benefit from or respond to LNS can help inform decisions regarding targeting. We conducted an IPD meta-analysis of randomized controlled trials (RCTs) of SQ-LNS provided to pregnant women that had 2 objectives: 1) to compare overall effects of SQ-LNS with provision of a) IFA or standard of care (IFA/SOC) or b) MMS, and 2) examine potential effect modifiers of the impact of SQ-LNS (as compared to either IFA/SOC or MMS) on birth outcomes.

## Methods

The protocol for this systematic review and IPD meta-analysis was prospectively registered on PROSPERO (CRD42021283391) (16). The statistical analysis plan (SAP) is available on Open Science Framework (https://osf.io/nj5f9/) (17) and was posted prior to analysis. The protocol was reviewed by the institutional review board (IRB) of the University of California, Davis and determined to be exempt from IRB approval given that protocols for each individual trial had been previously approved by their respective ethical committees.

### Inclusion and exclusion criteria for this IPD meta-analysis

We included prospective randomized controlled trials of maternal SQ-LNS that met the following study-level inclusion criteria: 1) trial was conducted in a low- or middle-income country (18), 2) maternal SQ-LNS (∼125 kcal/d) was provided for at least part of pregnancy to intervention group participants, 3) comparison group(s) received iron and folic acid (IFA), multiple micronutrient supplements (MMS) (defined below) or standard of care (SOC), 4) trial reported at least one outcome of interest (defined below), and 5) trial used an individual or cluster randomized design in which the same participants were measured at baseline and endline (longitudinal follow-up) or different participants were measured at baseline and endline (repeated cross-sectional data collection). Trials were excluded if: 1) severe or moderate malnutrition was an inclusion criterion for pregnant women to be eligible to participate, 2) study was conducted with sick or hospitalized populations, 3) the only available comparison group received other types of non-LNS maternal food supplementation, or 4) SQ-LNS provision was combined with an additional nutrition-specific intervention within a single arm (e.g. SQ-LNS + food rations vs. control), and there was no appropriate comparison group that would allow isolation of the SQ-LNS effect (e.g., food rations alone).

For comparisons of SQ-LNS with MMS, MMS was defined as including at least 3 micronutrients (19), and similar in form (e.g. tablet or capsule) to globally used MMS formulations (20). We used the same exclusion criterion as Smith et al. (19), i.e., excluding micronutrient powders because they provide additional nutrients compared to MMS tablets that might have independent effects on the outcomes of interest.

### Search methods and identification of studies

We began the search by considering the studies identified by and included in the 2018 Cochrane systematic review and meta-analysis of the provision of preventive LNS to pregnant women (13). Then we repeated the database search methods employed in that review to capture any studies published or registered as a randomized trial between January 2018 and September 2021. There were no language restrictions. We searched the following international electronic bibliographic databases: The Cochrane Library (Cochrane Central Register of Controlled Trials, Cochrane Database of Systematic Reviews), MEDLINE (Ovid, In-Process and Other Non-Indexed Citations Ovid, Epub ahead of print Ovid), EMBASE (Ovid), CINAHL Complete (EBSCOhost), Web of Science (Social Sciences Citation Index, Science Citation Index, Conference Proceedings Citation Index-Science, Conference Proceedings Citation Index-Social Sciences), Epistemonikos (current issue), ClinicalTrials.gov, and WHO International Clinical Trials Registry Platform. In addition, we searched nine regional databases: IBECS, SciELO (Scientific Electronic Library Online), AIM (Africa Index Medicus), IMEMR (Index Medicus for the Eastern Mediterranean Region), LILACS (Latin American and Caribbean Health Sciences Literature), IRIS (PAHO/WHO Institutional Repository for Information Sharing) WPRIM (Western Pacific Index Medicus), IMSEAR (Index Medicus for the South-East Asian Region), and Native Health Research Database.

After reviewing the titles and abstracts of all studies included in the previous review, as well as the additional studies identified by the database searches, we selected all potentially relevant studies for full text review and screened them based on the inclusion and exclusion criteria. After completing the search, we communicated with investigators of all potentially eligible studies in progress, regardless of whether the results had been published.

### Data collection and harmonization

We invited the principal investigators of eligible studies published or in progress to participate in this IPD meta-analysis, and we provided a data dictionary listing definitions of variables requested. Each contacted investigator provided de-identified individual participant datasets with those variables to the IPD analyst (CDA), who communicated with investigators to request any missing variables or other clarifications, as needed.

### IPD integrity

We conducted a complete-case intention-to-treat analysis (21). We evaluated whether the study sample sizes in our pooled data set were the same as in study protocols and publications. To address missing outcome data, we tabulated the percentage of participants lost to follow-up between enrollment and the assessment of the outcomes for each study. We also assessed whether missing data were differential with respect to intervention group by comparing rates of missingness across randomized arms.

We flagged biologically implausible values. For anthropometric outcomes, we calculated z-scores using the 2006 WHO child growth standards (22, 23) and the INTERGROWTH standards (24), checked the values for acceptable SDs, and flagged implausible values if they were outside of published WHO acceptable ranges (22, 23). Implausible values were inspected for errors and either winsorized (25) if within 2 SD of the WHO acceptable ranges or removed from analysis if clearly impossible on an outcome-by-outcome basis. Such cleaning was necessary for <0.2% of participants. There was a consistently low rate of implausibility across outcomes and studies. We also checked summary statistics from the harmonized data-set (e.g., means and SDs) against each trial’s published values.

### Assessment of risk of bias in each study and quality of evidence across studies

Two independent reviewers (KRW and CDA) assessed risk of bias in each trial against the following criteria: random sequence generation and allocation concealment (selection bias), blinding of participants and personnel (performance bias), blinding of outcome assessment (detection bias), incomplete outcome data (attrition bias), selective reporting (reporting bias), and other sources of bias (26). Any discrepancies were resolved by discussion or consultation with the core working group, as needed. KRW, CDA and CPS assessed the quality of evidence for anthropometric outcomes across all trials based on the 5 Grading of Recommendations Assessment, Development and Evaluation (GRADE) criteria: risk of bias, inconsistency of effect, imprecision, indirectness, and publication bias (27).

### Specification of outcomes and effect measures

We prespecified all outcomes in the SAP (17). As shown in **Box 1**, there were 3 categories of outcomes: 1) birth size and duration of gestation, 2) anthropometric outcomes at 6 mo of age, and 3) adverse outcomes (miscarriage, stillbirth, Cesarean-section, early neonatal mortality, neonatal mortality, and 0-6 mo mortality). Continuous birth size outcomes included values in units of measurement (e.g., birth weight in g; birth length, head circumference and mid-upper arm circumference (MUAC) in cm), as z-scores (length-for-age z-score (LAZ), weight-for-age z-score (WAZ), weight-for-length z-score (WLZ), BMI-for-age z-score (BMIZ), and head circumference-for-age z-score (HCZ)) based on WHO Child Growth Standards at birth (22, 23)), and as z-scores for gestational age (weight-for-gestational age z-score (WGAZ), length-for-gestational age z-score (LGAZ) and head circumference-for-gestational age z-score (HCGAZ) based on INTERGROWTH-21st standards (24)). At birth, BMIZ was used instead of WLZ because the latter cannot be calculated for children with lengths <45 cm and exclusion of those infants would create bias; when WLZ can be calculated these 2 variables are highly correlated (r>0.90). For continuous outcomes based on INTERGROWTH-21st standards, we excluded participants without ultrasound data for calculation of gestational age. Binary birth size outcomes included low birth weight (LBW, < 2500 g), birth weight < 2000 g, small-for gestational age (SGA, < 10th percentile based on INTERGROWTH-21st), large-for-gestational age (LGA, >90th percentile based on INTERGROWTH-21st), newborn stunting (LAZ < -2 SD), low LGAZ (< -2 SD), low BMIZ (< -2 SD), low HCZ (< -2 SD), and low HCGAZ (< -2 SD). Duration of gestation was expressed in wk, and preterm birth was defined as delivery < 37 wk. For binary outcomes based on gestational age, we retained participants without ultrasound dating but conducted a sensitivity analysis in which they were excluded.

For continuous outcomes, the principal measure of effect was the mean difference (MD) between intervention and comparison groups; for binary outcomes it was the relative risk (RR) for birth outcomes and adverse outcomes, and the prevalence ratio (PR; relative difference in proportions between groups) for anthropometric outcomes at 6 mo of age. We also estimated the effect for binary outcomes as the absolute differences between intervention groups in events per 1000 individuals. The absolute differences are useful for assessing public health impact, but we considered them secondary outcomes because they can be less consistent than RRs or PRs across studies (26).

The comparisons of interest were 1) SQ-LNS vs IFA/SOC and 2) SQ-LNS vs MMS. Study arms in which SQ-LNS began prior to conception were excluded from the main analyses because the objectives were to evaluate effects of SQ-LNS when given during pregnancy; however, the pre-conception study arms were included in sensitivity analyses.

### Synthesis methods and exploration of variation in effects

We followed the same procedures for analysis as described previously (28), which included evaluation of full sample main effects of the intervention as well as effect modification by individual-level characteristics. We had also planned to evaluate effect modification by study level characteristics (17) but only 4 eligible trials were identified, so there was inadequate statistical power for study-level effect modification; thus those analyses are not described herein.

Briefly, we followed a two-stage meta-analysis approach (29). In the first stage, intervention effect estimates (or effect modification interaction term estimates) were generated within each individual study according to its study design. In the second stage, the first-stage estimates were pooled using both inverse-variance fixed effects and random effects.

The individual-level characteristics considered as potential effect modifiers were similar to those included in our previous IPD meta-analysis of trials using child SQ-LNS (28). The potential effect modifiers for this analysis are shown in **Box 2**.

We assessed heterogeneity of effect estimates using I^2^ and Tau^2^ statistics, within strata when relevant (30). We used a P value <0.05 for main effects and a p-for-interaction < 0.10 for effect modification by individual-level characteristics. Given that the birth outcomes are highly correlated and the effect modification analyses are inherently exploratory, we did not adjust for multiple hypothesis testing because doing so may be unnecessary and counterproductive (31).

### Additional sensitivity analyses

We pre-specified several sensitivity analyses:

- Exclusion of trials with a high level of missingness (>20%) of outcome data relative to enrollment for pregnancy outcomes and relative to live births for later outcomes.
- For outcomes that rely on gestational age, exclusion of individuals for whom there is no ultrasound dating of the gestational age of the fetus.
- For birth outcomes (e.g., infant anthropometry), exclusion of individuals for whom data were collected >72 hours after birth.
- Exclusion of studies in which additional LNS was provided to certain mothers (e.g., those with low BMI or gestational weight gain).
- Inclusion of trials/arms that provided SQ-LNS prior to conception (pooled main effects only).

## Results

### Literature search and study characteristics

We identified 4 trials that met our selection criteria and were completed in time to be included in our analyses, all of which provided IPD (**Figure 1**, **Table 1**). The trials were conducted in Bangladesh (32), Ghana (33), Malawi (34), and Guatemala (35) between 2009 and 2017. The Guatemala study was one of 4 sites in the Women First trial (35). In that trial, an unfortified LNS (in addition to SQ-LNS) supplying 300 kcal/d was provided to intervention group women who were either underweight or who had inadequate gestational weight gain. In 3 of the 4 study sites (Democratic Republic of the Congo, India, and Pakistan), >90% of enrolled women in the intervention groups received this supplement in addition to SQ-LNS, so the total amount of LNS provided to them was ∼418 kcal/d, which would be categorized as MQ-LNS rather than SQ-LNS. Therefore, only the Guatemala site is included in these analyses (where < 10% of enrolled women received the unfortified LNS in addition to SQ-LNS).

**Figure 1:**
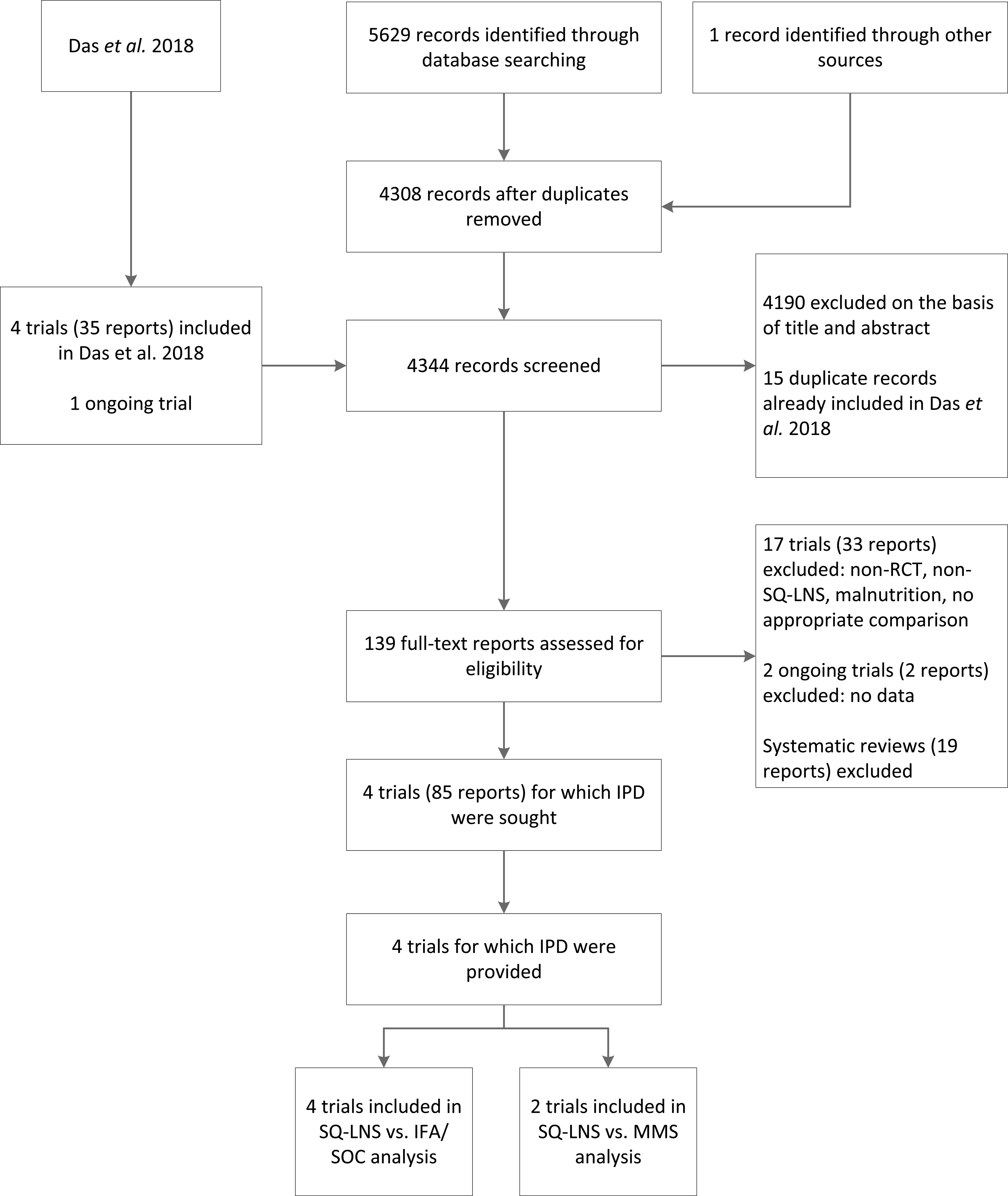
Study flow diagram. IFA, iron and folic acid supplements; IPD, individual participant data; MMS, multiple micronutrient supplements; RCT, randomized controlled trial; SOC, standard of care; SQ-LNS, small-quantity lipid-based nutrient supplements.

**Table 1.**
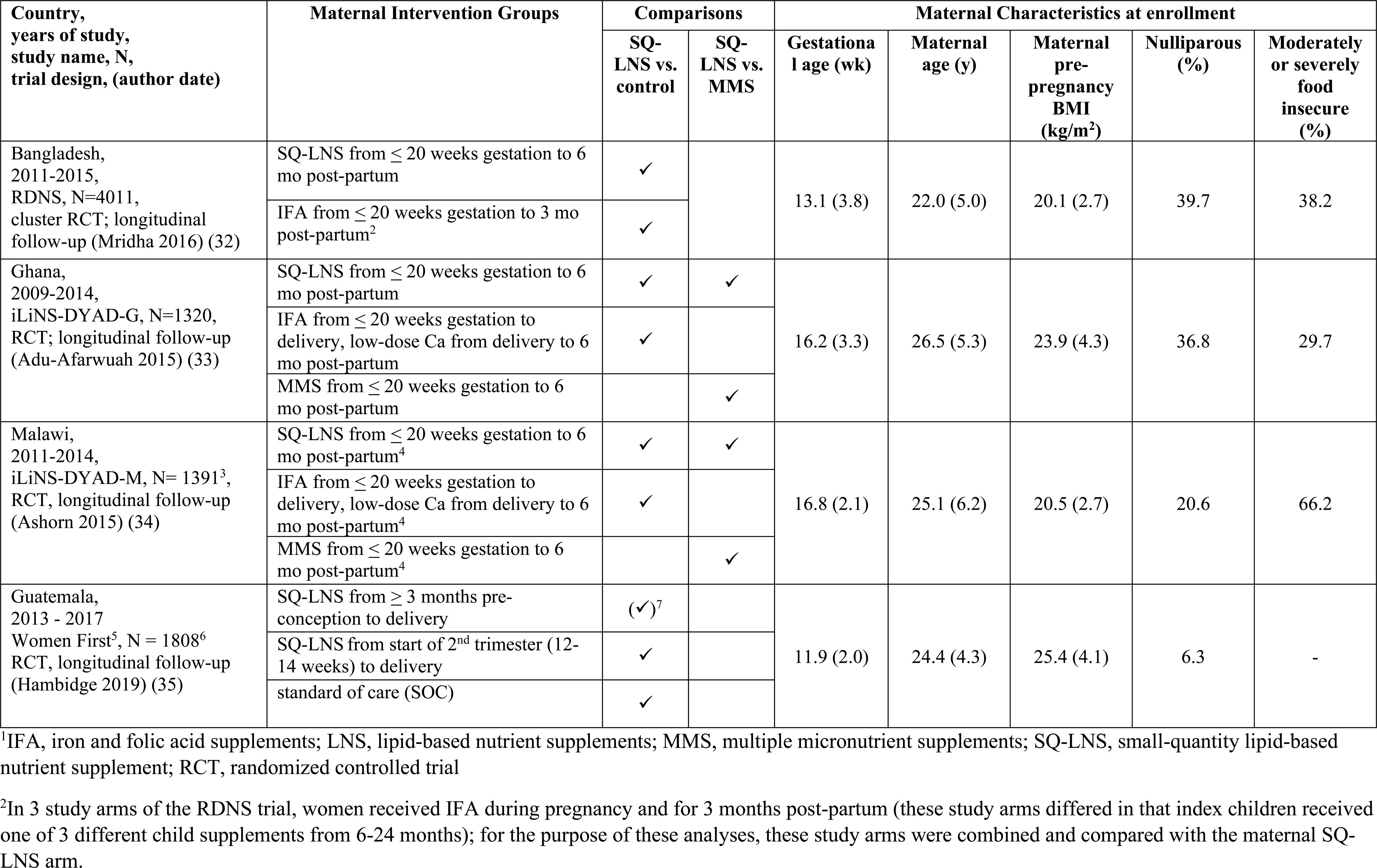
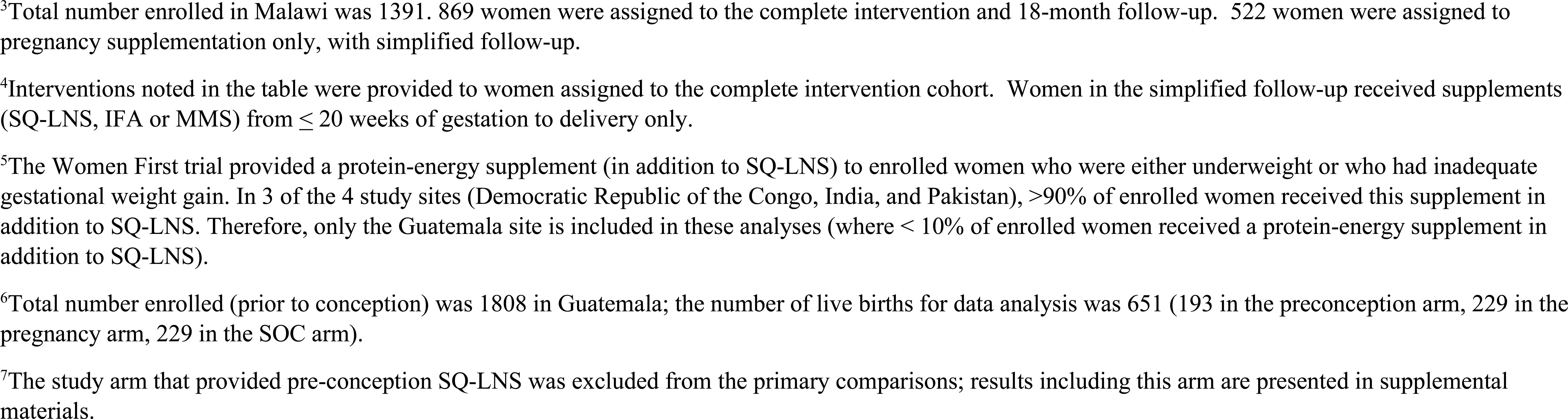
Characteristics of trials included in the individual participant data analysis^1^.

| Country, years of study, study name, N, trial design, (author date) | Maternal Intervention Groups | Comparisons |  | Maternal Characteristics at enrollment |  |  |  |  |
| --- | --- | --- | --- | --- | --- | --- | --- | --- |
|  |  | SQ-LNS vs. control | SQ-LNS vs. MMS | Gestational age (wk) | Maternal age (y) | Maternal pre-pregnancy BMI (kg/m<sup>2</sup>) | Nulliparous (%) | Moderately or severely food insecure (%) |
| Bangladesh, 2011-2015, RDNS, N=4011, cluster RCT; longitudinal follow-up (Mridha 2016) (32) | SQ-LNS from ≤ 20 weeks gestation to 6 mo post-partum | ✓ |  | 13.1 (3.8) | 22.0 (5.0) | 20.1 (2.7) | 39.7 | 38.2 |
|  | IFA from ≤ 20 weeks gestation to 3 mo post-partum<sup>2</sup> | ✓ |  |  |  |  |  |  |
| Ghana, 2009-2014, iLiNS-DYAD-G, N=1320, RCT; longitudinal follow-up (Adu-Afarwuah 2015) (33) | SQ-LNS from ≤ 20 weeks gestation to 6 mo post-partum | ✓ | ✓ | 16.2 (3.3) | 26.5 (5.3) | 23.9 (4.3) | 36.8 | 29.7 |
|  | IFA from ≤ 20 weeks gestation to delivery, low-dose Ca from delivery to 6 mo post-partum | ✓ |  |  |  |  |  |  |
|  | MMS from ≤ 20 weeks gestation to 6 mo post-partum |  | ✓ |  |  |  |  |  |
| Malawi, 2011-2014, iLiNS-DYAD-M, N= 1391<sup>3</sup>, RCT, longitudinal follow-up (Ashorn 2015) (34) | SQ-LNS from ≤ 20 weeks gestation to 6 mo post-partum<sup>4</sup> | ✓ | ✓ | 16.8 (2.1) | 25.1 (6.2) | 20.5 (2.7) | 20.6 | 66.2 |
|  | IFA from ≤ 20 weeks gestation to delivery, low-dose Ca from delivery to 6 mo post-partum<sup>4</sup> | ✓ |  |  |  |  |  |  |
|  | MMS from ≤ 20 weeks gestation to 6 mo post-partum<sup>4</sup> |  | ✓ |  |  |  |  |  |
| Guatemala, 2013 - 2017 Women First<sup>5</sup>, N = 1808<sup>6</sup> RCT, longitudinal follow-up (Hambidge 2019) (35) | SQ-LNS from ≥ 3 months pre-conception to delivery | (✓)<sup>7</sup> |  | 11.9 (2.0) | 24.4 (4.3) | 25.4 (4.1) | 6.3 | - |
|  | SQ-LNS from start of 2<sup>nd</sup> trimester (12-14 weeks) to delivery | ✓ |  |  |  |  |  |  |
|  | standard of care (SOC) | ✓ |  |  |  |  |  |  |

<sup>1</sup>IFA, iron and folic acid supplements; LNS, lipid-based nutrient supplements; MMS, multiple micronutrient supplements; SQ-LNS, small-quantity lipid-based nutrient supplement; RCT, randomized controlled trial
<sup>2</sup>In 3 study arms of the RDNS trial, women received IFA during pregnancy and for 3 months post-partum (these study arms differed in that index children received one of 3 different child supplements from 6-24 months); for the purpose of these analyses, these study arms were combined and compared with the maternal SQ-LNS arm.
<sup>3</sup>Total number enrolled in Malawi was 1391. 869 women were assigned to the complete intervention and 18-month follow-up. 522 women were assigned to pregnancy supplementation only, with simplified follow-up.
<sup>4</sup>Interventions noted in the table were provided to women assigned to the complete intervention cohort. Women in the simplified follow-up received supplements (SQ-LNS, IFA or MMS) from $\leq 20$ weeks of gestation to delivery only.
<sup>5</sup>The Women First trial provided a protein-energy supplement (in addition to SQ-LNS) to enrolled women who were either underweight or who had inadequate gestational weight gain. In 3 of the 4 study sites (Democratic Republic of the Congo, India, and Pakistan), >90% of enrolled women received this supplement in addition to SQ-LNS. Therefore, only the Guatemala site is included in these analyses (where < 10% of enrolled women received a protein-energy supplement in addition to SQ-LNS).
<sup>6</sup>Total number enrolled (prior to conception) was 1808 in Guatemala; the number of live births for data analysis was 651 (193 in the preconception arm, 229 in the pregnancy arm, 229 in the SOC arm).
<sup>7</sup>The study arm that provided pre-conception SQ-LNS was excluded from the primary comparisons; results including this arm are presented in supplemental materials.

All 4 trials provided data for the comparison of SQ-LNS vs IFA/SOC; in 3 trials IFA was provided to this comparison group (32–34), and in 1 trial (Guatemala) the comparison group received SOC (biweekly visits to monitor pregnancy status) (35). Two of the 4 trials (Ghana and Malawi) also included an arm that received MMS and thus provided data for the comparison of SQ-LNS vs MMS (33, 34). In Ghana and in the “complete follow-up” cohort in Malawi (who received supplementation beyond pregnancy, see Table 1), SQ-LNS and MMS were provided until 6 mo postpartum and IFA was provided until delivery followed by placebo (low dose calcium) until 6 mo postpartum; in the “simplified follow-up” cohort in Malawi (supplementation only during pregnancy), SQ-LNS, MMS or IFA were provided only until delivery (33, 34). In Bangladesh, SQ-LNS was provided until 6 mo postpartum and IFA until 3 mo postpartum (32), and in Guatemala, SQ-LNS was provided only until delivery (35). The nutrient composition of SQ-LNS was nearly identical across trials (**Supplemental Table 1)**, except for more vitamin D and less zinc in the version used in Guatemala. The MMS used in Ghana and Malawi was a formulation designed to match the micronutrient content of the SQ-LNS (11), which was based in part on results of a trial in Guinea-Bissau (36) that demonstrated a greater impact on birth weight when the MMS contained 2X (vs 1X) the recommended dietary allowance of most of the nutrients (except for iron, folic acid and vitamin A). Thus, the MMS used in Ghana and Malawi had higher levels of most of the micronutrients compared to the United Nations International Multiple Micronutrient Antenatal Preparation (UNIMMAP) (37).

Mean gestational age when prenatal supplementation began was ∼12 wk in Guatemala, ∼13 wk in Bangladesh, and 16-17 wk in Ghana and Malawi (**Table 1**). Mean maternal age was lowest in Bangladesh (∼22 y) and 24-27 y in the other sites. Mean maternal BMI was lowest in Bangladesh (20.1 kg/m^2^) and Malawi (20.5 kg/m^2^), and was considerably higher in Ghana and Guatemala (∼24-25 kg/m^2^). The proportion of women who were nulliparous at enrollment was highest in Bangladesh and lowest in Guatemala. In the 3 sites in which household food insecurity was assessed (Ghana, Malawi and Bangladesh), the percentage reporting moderate or severe food insecurity ranged from 30% in Ghana to 66% in Malawi. Additional information on maternal, child and household characteristics is available in **Supplemental Table 2**.

The study populations differed with regard to risk of adverse birth outcomes, with Bangladesh having the highest incidence (in the IFA/SOC group) of LBW (37%), SGA (59%), newborn stunting (23%), low BMIZ (34%) and preterm birth (14%) (**Supplemental Table 3**). The incidence of these outcomes in the IFA/SOC groups in the other 3 sites was 13-14% for LBW, 22-27% for SGA, 10-17% for newborn stunting, 6-11% for low BMIZ, and 8-12% for preterm birth. Study-level data on anthropometric outcomes at 6 mo of age and adverse outcomes in the IFA/SOC groups are available in **Supplemental Tables 4-5**.

All trials were judged to have low risk of bias for 6 of the 7 categories: random sequence generation, allocation concealment, outcome assessment (except for 1 trial labeled “unclear”), incomplete outcome, selective reporting, and “other” (**Supplemental Table 6** and **Supplemental Figure 1).** All trials had a high risk of bias for blinding of participants, as blinding was not possible given the physical difference in the supplements provided.

#### SQ-LNS vs IFA/SOC

##### Main effects

For most birth outcomes, all 4 trials contributed to the pooled effect estimates and the total sample size was 4,922-5,348 (**Table 2**). For continuous birth outcomes based on INTERGROWTH-21st standards, we excluded participants without ultrasound data, as mentioned above, and as a result only 3 trials contributed to the estimates for WGAZ, LGAZ and HCGAZ (total n ∼1,460-1,717).

**Table 2.** Main effects of maternal SQ-LNS vs IFA/SOC on birth outcomes^1^.

| **Birth outcome** | **Number of participants (trials)** | **SQ-LNS events per 1000** | **IFA/SOC events per 1000** | **MD or RR plus difference in events per 1000 (95% CI)<sup>2</sup>** | **P value<sup>3</sup>** | **Heterogeneity *I*<sup>2</sup>** | **Quality of the evidence (GRADE)** |
| --- | --- | --- | --- | --- | --- | --- | --- |
| Birth weight (g) | 5273 (4) | - | - | 48.7 (26.1, 71.2) | <0.001 | 0.00 | Moderate |
| Weight-for-age z score (WAZ) | 5273 (4) | - | - | 0.12 (0.06, 0.17) | <0.001 | 0.00 | Moderate |
| Weight-for-gestational age z-score (WGAZ)<sup>4</sup> | 1717 (3) | - | - | 0.13 (0.05, 0.21) | 0.001 | 0.31 | Moderate |
| Low birth weight (LBW) | 5273 (4) | 168 | 198 | 0.89 (0.80, 0.99)<br>-28.6 (-51.3, -5.9) | 0.033<br>0.013 | 0.00<br>0.00 | Moderate |
| Birth weight < 2 kg | 5273 (4) | 25 | 35 | 0.78 (0.60, 1.01)<br>-9.1 (-18.5, 0.2) | 0.062<br>0.054 | 0.00<br>0.00 | Moderate |
| Small-for-gestational age (SGA) | 5181 (4) | 303 | 335 | 0.96 (0.92, 1.01)<br>-23.0 (-46.7, 0.7) | 0.133<br>0.057 | 0.00<br>0.00 | Moderate |
| Large-for-gestational age (LGA) | 4834 (3) | 20 | 23 | 1.00 (0.61, 1.65)<br>1.1 (-3.1, 5.3) | 0.984<br>0.611 | 0.00<br>0.00 | Moderate |
| Birth length (cm) | 5014 (4) | - | - | 0.20 (0.09, 0.31) | <0.001 | 0.00 | Moderate |
| Length-for-age z score (LAZ) | 5014 (4) | - | - | 0.11 (0.06, 0.17) | <0.001 | 0.00 | Moderate |
| Length-for-gestational age z-score (LGAZ)<sup>4</sup> | 1460 (3) | - | - | 0.13 (0.05, 0.21) | 0.002 | 0.00 | Moderate |
| Newborn stunting | 5014 (4) | 138 | 164 | 0.83 (0.74, 0.93)<br>-32.2 (-51.5, -12.9) | 0.001<br>0.001 | 0.00<br>0.00 | Moderate |
| Low LGAZ | 4922 (4) | 118 | 127 | 0.90 (0.80, 1.01)<br>-12.0 (-29.6, 5.6) | 0.082<br>0.182 | 0.00<br>0.00 | Moderate |
| BMI-for-age z-score (BMIZ) | 5002 (4) | - | - | 0.10 (0.04, 0.16) | 0.002 | 0.00 | Moderate |
| Low BMIZ | 5002 (4) | 123 | 145 | 0.89 (0.81, 0.98)<br>-23.6 (-42.7, -4.5) | 0.022<br>0.015 | 0.26<br>0.20 | Moderate |
| Head circumference (cm) | 5016 (4) | - | - | 0.11 (0.04, 0.18) | 0.002 | 0.00 | Moderate |
| Head circumference-for-age z-score (HCZ) | 5016 (4) | - | - | 0.10 (0.04, 0.16) | 0.001 | 0.00 | Moderate |
| Head circumference-for-gestational age z-score (HCGAZ)<sup>4</sup> | 1461 (3) | - | - | 0.11 (0.02, 0.20) | 0.019 | 0.00 | Moderate |
| Low HCZ | 5016 (4) | 100 | 118 | 0.85 (0.75, 0.96)<br>-22.1 (-40.5, -3.7) | 0.009<br>0.019 | 0.00<br>0.00 | Moderate |
| Low HCGAZ | 4924 (4) | 54 | 64 | 0.88 (0.76, 1.01)<br>-6.4 (-18.4, 5.5) | 0.072<br>0.291 | 0.00<br>0.32 | Moderate |
| Mid-upper arm circumference (MUAC) (cm) | 4581 (3) | - | - | 0.08 (0.02, 0.14) | 0.009 | 0.00 | Moderate |
| Duration of gestation (wk) | 5348 (4) | - | - | 0.12 (0.01, 0.24) | 0.040 | 0.00 | Moderate |
| Preterm birth | 5348 (4) | 106 | 111 | 0.94 (0.80, 1.10)<br>-7.1 (-25.8, 11.5) | 0.445<br>0.454 | 0.00<br>0.00 | Moderate |

<sup>1</sup>BMIZ, BMI-for-age z-score; GRADE, Grading of Recommendations Assessment, Development and Evaluation; HCGAZ, head circumference-for-gestational age z-score; HCZ, head circumference-for-age z-score; IFA, iron and folic acid supplement; LAZ, length-for-age z-score; LBW, low birth weight; LGA, large-for-gestational age; LGAZ, length-for-gestational age z-score; MD, mean difference; MUAC, mid-upper arm circumference; RR, relative risk; SGA, small-for-gestational age; SOC, standard of care; SQ-LNS, small-quantity lipid-based nutrient supplement; WAZ, weight-for-age z-score; WGAZ, weight-for-gestational age z-score
<sup>2</sup>For continuous outcomes, values are MDs: LNS – IFA/SOC (95% CIs). For binary outcomes, values are RRs: LNS compared with IFA/SOC (95% CIs). Difference in events per 1000 was calculated using the prevalence difference (95% CI) and multiplying by 1000.
<sup>3</sup>The P value column corresponds to the pooled main effect 2-sided superiority testing of the intervention effect estimate and 95% CI presented in the preceding column. $I^2$ describes the percentage of variability in effect estimates that may be due to heterogeneity rather than chance. Roughly, 0.3 – 0.6 may be considered moderate heterogeneity.
<sup>4</sup> For continuous outcomes based on INTERGROWTH-21st standards, we excluded participants without ultrasound data for calculation of gestational age. For the corresponding binary outcomes (SGA, LGA, Low LGAZ, Low HCGAZ), we retained participants without ultrasound dating but conducted a sensitivity analysis in which they were excluded.

Maternal SQ-LNS had a significant positive effect on all of the continuous birth outcomes, with a MD compared to IFA/SOC of +49 g for birth weight, +0.2 cm for birth length, +0.10-0.13 z-scores for WAZ, WGAZ, LAZ, LGAZ, BMIZ, HCZ, and HCGAZ, +0.08 cm for MUAC, and +0.12 wk for duration of gestation. Maternal SQ-LNS reduced the risk of LBW by 11%, newborn stunting by 17%, low BMIZ by 11%, and low HCZ by 11%. Effect estimates for the other binary birth outcomes were not statistically significant but were in the same direction (except for LGA, RR 1.00 (0.61, 1.65)).

We rated the quality of the evidence for all birth outcomes as moderate based on the GRADE criteria listed in the Methods: 3-4 RCTs were available for all outcomes, risk of bias was low except for blinding of participants, heterogeneity was low, precision was rated as high because all trials had sample sizes > 400, all trials were directly aimed at evaluating SQ-LNSs, and funnel plots revealed no indication of publication bias.

In general, there was consistency across studies in the direction of effects, with very low heterogeneity based on I^2^ values (**Figures 2A-F** for birth weight, LBW, newborn LAZ, newborn HCZ, newborn BMIZ and duration of gestation; **Supplemental Figures 2A-BM** for all outcomes). Low I^2^ values may be attributable to relatively wide CIs for some of the point estimates rather than low variability in the RRs. For nearly all outcomes, fixed effects and random-effects models generated identical estimates.

**Figures 2 A-F:**
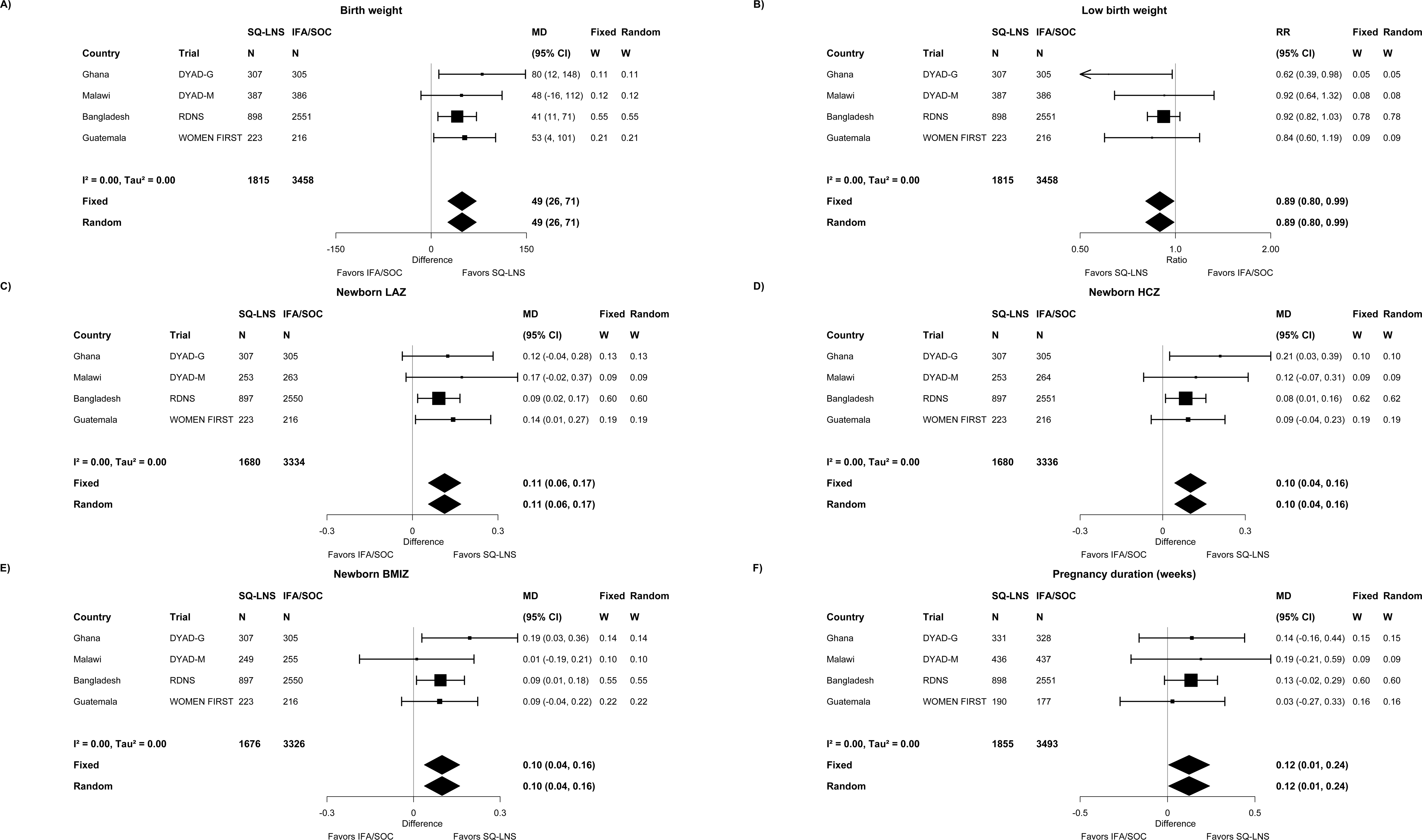
Forest plots of effect of SQ-LNS vs IFA/SOC on birth weight, LBW, newborn LAZ, newborn HCZ, newborn BMIZ and duration of gestation. BMIZ, body mass index z-score; HCZ, head circumference-for-age z-score; IFA, iron and folic acid; LAZ, length-for-age z-score; LNS, lipid-based nutrient supplements; LBW, low birthweight; MD, mean difference; RR, relative risk; SOC, standard of care; SQ-LNS, small-quantity lipid-based nutrient supplements. Individual study estimates were generated from log-binomial regression for binary outcomes and linear regression for continuous outcomes controlling for baseline measure when available and using robust standard errors for cluster-randomized trials. Pooled estimates were generated using inverse-variance weighting with both fixed and random effects.

With regard to the sensitivity analyses, no trials had a high level of missingness of outcome data, so no sensitivity analyses for that reason were needed. When including only ultrasound dating for outcomes dependent on gestational age (which excluded the Bangladesh trial), the magnitude of the effect estimates increased but effects remained non-significant: RR=0.86 (0.73, 1.01) for SGA (n=1,717), MD=0.15 (-0.03, 0.33) for duration of gestation (n=1,880) and RR=0.90 (0.68, 1.18) for preterm birth (n=1,880) (**Supplemental Table 7A**). For the other sensitivity analyses, restricting birth size outcomes to data collected within 72 h of birth, or excluding the Guatemala trial (in which some women received additional LNS), or including the pre-conception arm of the Guatemala trial, there was very little change in the findings.

For most of the effects of SQ-LNS vs IFA/SOC on anthropometric outcomes at 6 mo of age, all 4 trials contributed to the pooled estimates and the total sample size was ∼5,030; outcomes based on MUACZ were not available for Guatemala (**Table 3)**. Maternal SQ-LNS reduced underweight at 6 mo (PR 0.85 (0.73, 0.99)), but effects on the other outcomes were not statistically significant. In sensitivity analyses, the PR for underweight became non-significant when Guatemala was excluded (0.96 (0.80, 1.16)) (with or without exclusion of the “simplified follow-up” cohort in Malawi; in this cohort and in Guatemala, maternal supplementation did not continue after delivery). The PR for underweight was not strengthened when the pre-conception arm was included for Guatemala (0.90 (0.78, 1.13)). For other outcomes, these sensitivity analyses did not substantively alter the findings (**Supplemental Table 7B**).

**Table 3.** Main effects of maternal SQ-LNS vs IFA/SOC on infant anthropometric outcomes at 6 mo of age^1^.

| Infant anthropometric outcomes at 6 mo | Number of participants (trials) | SQ-LNS events per 1000 | IFA/SOC events per 1000 | MD or PR plus difference in events per 1000 (95% CI)<sup>2</sup> | P value<sup>3</sup> | Heterogeneity *I*<sup>2</sup> | Quality of the evidence (GRADE) |
| --- | --- | --- | --- | --- | --- | --- | --- |
| Weight-for-age z-score (WAZ) | 5029 (4) | - | - | 0.05 (-0.02, 0.13) | 0.173 | 0.00 | Moderate |
| Underweight | 5029 (4) | 114 | 132 | 0.85 (0.73, 0.99)<br>-22.1 (-41.8, -2.5) | 0.041<br>0.027 | 0.41<br>0.37 | Moderate |
| Length-for-age z-score (LAZ) | 5032 (4) | - | - | 0.06 (-0.01, 0.13) | 0.080 | 0.02 | Moderate |
| Stunted | 5032 (4) | 198 | 219 | 0.90 (0.80, 1.02)<br>-17.4 (-43.3, 8.5) | 0.102<br>0.187 | 0.00<br>0.00 | Moderate |
| Weight-for-length z-score (WLZ) | 5028 (4) | - | - | 0.00 (-0.07, 0.07) | 0.969 | 0.00 | Moderate |
| Wasted | 5028 (4) | 24 | 32 | 0.91 (0.66, 1.25)<br>-6.3 (-15.8, 3.1) | 0.558<br>0.191 | 0.39<br>0.36 | Moderate |
| Head circumference-for-age z-score (HCZ) | 5028 (4) | - | - | 0.01 (-0.05, 0.07) | 0.679 | 0.00 | Moderate |
| Low HCZ | 5028 (4) | 110 | 125 | 0.90 (0.80, 1.02)<br>-13.2 (-31.9, 5.4) | 0.098<br>0.164 | 0.00<br>0.00 | Moderate |
| MUAC-for-age z-score (MUACZ) | 4600 (3) | - | - | 0.00 (-0.07, 0.08) | 0.949 | 0.00 | Moderate |
| Low MUAC [MUACZ < -2 SD or MUAC < 125 mm] | 4600 (3) | 68 | 64 | 1.00 (0.78, 1.29)<br>-0.1 (-17.9, 17.7) | 0.970<br>0.993 | 0.00<br>0.00 | Moderate |
| Acute malnutrition [WLZ < -2 SD or MUAC < 125 mm] | 4595 (3) | 80 | 82 | 1.00 (0.79, 1.25)<br>-0.7 (-19.4, 17.9) | 0.970<br>0.938 | 0.00<br>0.00 | Moderate |

<sup>1</sup>GRADE, Grading of Recommendations Assessment, Development and Evaluation; HCZ, head circumference-for-age z-score; IFA, iron and folic acid supplement; LAZ, length-for-age z-score; MD, mean difference; MUAC, mid-upper arm circumference; MUACZ, mid-upper arm circumference-for-age z-score; PR, prevalence ratio; SOC, standard of care; SQ-LNS, small-quantity lipid-based nutrient supplement; WAZ, weight-for-age z-score; WLZ, weight-for-length z-score<sup>2</sup>For continuous outcomes, values are MDs: LNS – IFA/SOC (95% CIs). For binary outcomes, values are PRs: LNS compared with IFA/SOC (95% CIs). Difference in events per 1000 was calculated using the prevalence difference (95% CI) and multiplying by 1000.
<sup>3</sup>The P value column corresponds to the pooled main effect 2-sided superiority testing of the intervention effect estimate and 95% CI presented in the preceding column. *I*<sup>2</sup> describes the percentage of variability in effect estimates that may be due to heterogeneity rather than chance. Roughly, 0.3 – 0.6 may be considered moderate heterogeneity.

There were no significant differences in any of the adverse outcomes for the SQ-LNS vs IFA/SOC comparison, and the RR was generally < 1 except for incidence of stillbirths (**Table 4**).

**Table 4.** Main effects of maternal SQ-LNS vs IFA/SOC on adverse outcomes^1^.

| Adverse outcomes | Number of participants (trials) | SQ-LNS events per 1000 | IFA/SOC events per 1000 | RR and difference in events per 1000 (95% CI)<sup>2</sup> | P value<sup>3</sup> | Heterogeneity *I*<sup>2</sup> | Quality of the evidence (GRADE) |
| --- | --- | --- | --- | --- | --- | --- | --- |
| Cesarian-Section | 5693 (4) | 167 | 156 | 1.04 (0.90, 1.19)<br>18.6 (-3.6, 40.9) | 0.595<br>0.101 | 0.42<br>0.01 | Moderate |
| Miscarriage | 6102 (4) | 49 | 59 | 0.87 (0.70, 1.08)<br>0.7 (-7.1, 8.6) | 0.213<br>0.852 | 0.00<br>0.27 | Moderate |
| Stillbirth | 6102 (4) | 24 | 18 | 1.31 (0.90, 1.91)<br>7.1 (-1.7, 15.9) | 0.153<br>0.116 | 0.49<br>0.53 | Moderate |
| Miscarriage or stillbirth | 6102 (4) | 75 | 77 | 0.98 (0.81, 1.19)<br>1.4 (-12.3, 15.1) | 0.870<br>0.844 | 0.39<br>0.40 | Moderate |
| Early neonatal mortality | 5506 (4) | 14 | 20 | 0.74 (0.46, 1.19)<br>-4.8 (-13.2, 3.7) | 0.208<br>0.272 | 0.04<br>0.22 | Moderate |
| Miscarriage or stillbirth or early neonatal mortality | 6102 (4) | 89 | 97 | 0.93 (0.77, 1.12)<br>-6.6 (-23.4, 10.3) | 0.452<br>0.444 | 0.00<br>0.00 | Moderate |
| Neonatal mortality | 5528 (4) | 21 | 25 | 0.83 (0.56, 1.24)<br>-2.9 (-12.3, 6.6) | 0.360<br>0.551 | 0.00<br>0.10 | Moderate |
| Mortality 0-6 mo | 5566 (4) | 30 | 32 | 0.95 (0.69, 1.30)<br>-0.6 (-11.0, 9.8) | 0.739<br>0.913 | 0.00<br>0.00 | Moderate |

<sup>1</sup>GRADE, Grading of Recommendations Assessment, Development and Evaluation; IFA, iron and folic acid supplement; RR, relative risk; SOC, standard of care; SQ-LNS, small-quantity lipid-based nutrient supplement
<sup>2</sup>Values are RRs: LNS compared with IFA/SOC (95% CIs). Difference in events per 1000 was calculated using the prevalence difference (95% CI) and multiplying by 1000.
<sup>3</sup>The P value column corresponds to the pooled main effect 2-sided superiority testing of the intervention effect estimate and 95% CI presented in the preceding column. *I*<sup>2</sup> describes the percentage of variability in effect estimates that may be due to heterogeneity rather than chance. Roughly, 0.3 – 0.6 may be considered moderate heterogeneity.

Because of uncertainty in gestational age dating, it is sometimes difficult to distinguish between miscarriage and stillbirths (i.e., before vs. after 28 wk gestation), and in low resource settings it can also be difficult to differentiate between stillbirth and early neonatal death (34). For these reasons, Table 4 includes comparisons for 2 composite variables for these outcomes (miscarriage or stillbirth; miscarriage or stillbirth or early neonatal mortality). The RRs for those composite adverse outcomes were both < 1.0. Adverse outcome findings were similar in the sensitivity analyses (**Supplemental Table 7C**).

##### Effect modification by individual-level characteristics

For several characteristics, the p-for-interaction was > 0.10 for all or almost all infant birth outcomes, i.e., effect modification was generally not evident for child birth order, maternal height, education, or anemia at baseline, gestational age at start of supplementation, compliance with supplementation, household socio-economic status or sanitation (see **Supplemental Figures 3A-N** for results stratified by all characteristics). For 5 characteristics, the p-for-interaction was < 0.10 for at least 2 birth outcomes, and data from at least 3 trials were available (**Figures 3A-J** show 12 selected birth outcomes for each of these characteristics, though the number of outcomes for which p-for-interaction was < 0.10 varied). The estimated effects of maternal SQ-LNS on birth outcomes were greater among a) female (vs. male) infants, for birth weight, WAZ, LBW, birth weight < 2 kg, BMIZ, low BMIZ, HCZ, duration of gestation, and preterm birth; b) women with lower BMI (< 20 vs. ≥ 20 kg/m^2^), for MUAC and low infant HCZ; c) younger women (< 25 vs. ≥ 25 y), for duration of gestation and birth weight <2 kg (although a greater effect on SGA was seen among women ≥ 25 y); d) women with inflammation at baseline (vs. without inflammation), for WAZ (Supplemental Figure 3H1), duration of gestation and low infant HCZ; and e) women with greater household food insecurity (moderate to severe vs. mild or secure), for birth length (Supplemental Figure 3M1), LAZ, head circumference (Supplemental Figure 3M1) and newborn stunting. Only 2 trials had information on maternal malaria at baseline, but in those trials the effect estimates for maternal SQ-LNS were greater among women with a positive rapid test for malaria at baseline (vs. those with a negative test) for birth weight, WAZ, LBW and SGA (Supplemental Figures 3I1-2). In sensitivity analyses including only ultrasound dating for outcomes dependent on gestational age (**Supplemental Table 8**), the interaction with child sex was still evident for duration of gestation but not preterm birth; the interaction with maternal age was still significant (and stronger) for duration of gestation but not for SGA; the interaction with maternal inflammation became weaker for duration of gestation; and there was no change in the interaction with maternal malaria for SGA. In sensitivity analyses restricted to anthropometric outcomes assessed within 72 h of birth, results were generally stronger for the interactions with child sex and maternal inflammation, similar for maternal BMI, age, and household food insecurity, and weaker for maternal malaria (for birth weight and LBW, but not SGA). The other sensitivity analyses (e.g., exclusion of the Guatemala study) did not substantively alter the results of the effect modification analyses (available at https://osf.io/nj5f9/ (17)).

**Figures 3 A-J:**
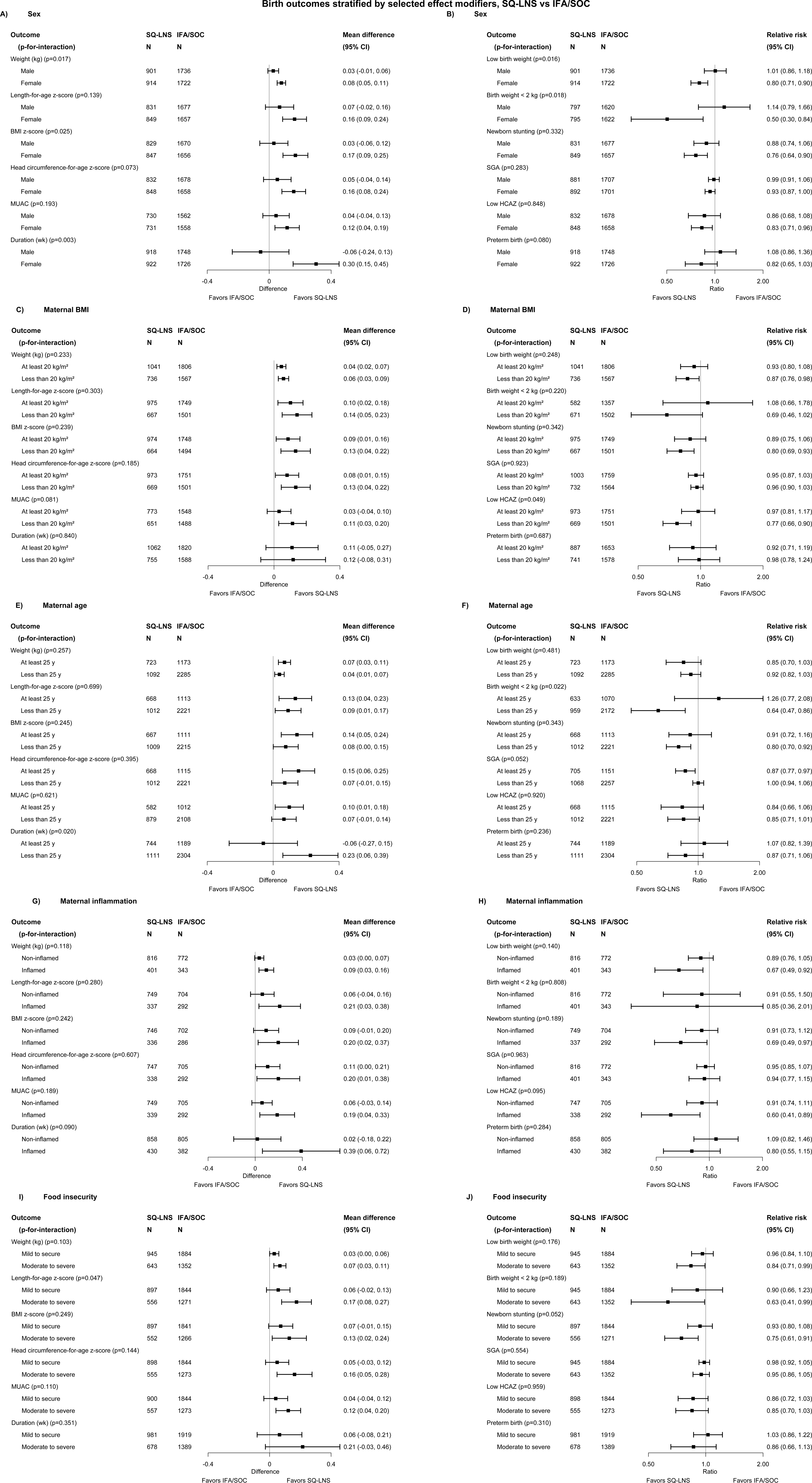
Pooled effects of SQ-LNS vs IFA/SOC on birth outcomes, stratified by selected effect modifiers. LNS, lipid-based nutrient supplements; P-for-interaction, p-value for the interaction indicating the difference in effects of SQ-LNS between the two levels of the effect modifier; PR, prevalence ratio; SQ-LNS, small-quantity lipid-based nutrient supplements. Individual study estimates (not shown) were generated from log-binomial regression for binary outcomes and linear regression for continuous outcomes controlling for baseline measure when available and using robust standard errors for cluster-randomized trials. Pooled estimates (shown here) were generated using inverse-variance weighting with fixed effects.

We explored whether the greater effect of SQ-LNS on birth size outcomes among females (vs. males) could be attributed mainly to the mediating effects on duration of gestation: when including duration of gestation in the models, the p-for-interaction was > 0.10 for birth weight, WAZ, BMIZ and HCZ but remained < 0.10 for LBW, birth weight < 2 kg, and low BMIZ (available at https://osf.io/nj5f9/(17)).

Some of the above characteristics also modified the effect estimates for maternal SQ-LNS vs. IFA/SOC with respect to infant anthropometric outcomes at 6 mo, particularly infant sex. Effect estimates were larger for females than for males, as was the case for the birth outcomes (**Figure 4**). Among females, maternal SQ-LNS was associated with a 26% reduction in underweight and a 30% reduction in low HCZ, whereas there was no reduction among males. These findings were very similar in the sensitivity analyses excluding Guatemala (with or without exclusion of the “simplified follow-up” cohort in Malawi) (available at https://osf.io/nj5f9/ (17)). For 3 other characteristics, the p-for-interaction was < 0.10 for at least 2 distinct infant outcomes at 6 mo.

**Figures 4 A,B:**
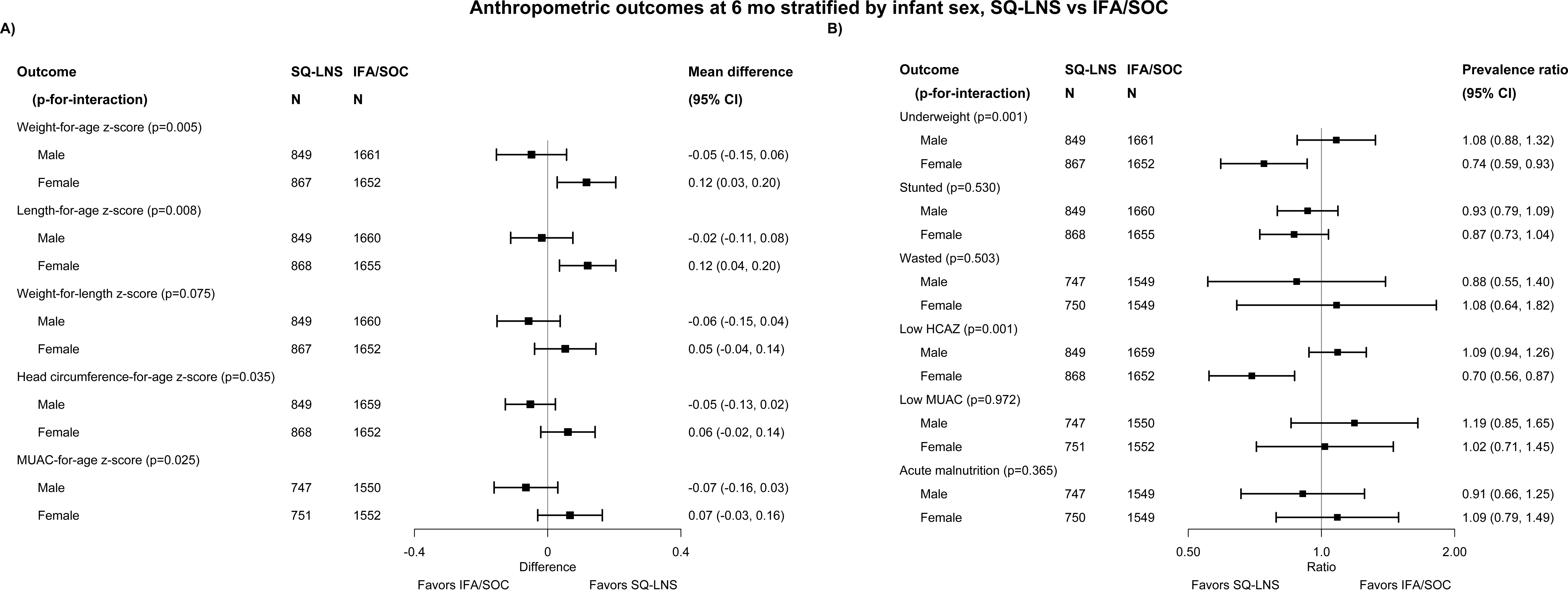
Pooled effects of SQ-LNS vs IFA/SOC on anthropometric outcomes at 6 mo of age, stratified by infant sex. LNS, lipid-based nutrient supplements; P-for-interaction, p-value for the interaction indicating the difference in effects of SQ-LNS between the two levels of the effect modifier; PR, prevalence ratio; SQ-LNS, small-quantity lipid-based nutrient supplements. Individual study estimates (not shown) were generated from log-binomial regression for binary outcomes and linear regression for continuous outcomes controlling for baseline measure when available and using robust standard errors for cluster-randomized trials. Pooled estimates (shown here) were generated using inverse-variance weighting with fixed effects.

Effect estimates for maternal SQ-LNS were greater among a) women with greater food insecurity, for LAZ and underweight at 6 mo (**Supplemental Figures 3M3-4**); b) women with improved sanitation, for WAZ and WLZ at 6 mo (**Supplemental Figure 3N3**); and c) later-born children, for underweight and stunting at 6 mo (**Supplemental Figure 3B4**).

**Supplemental Figures 4A-N** present pooled estimates for adverse outcomes, stratified by potential individual-level effect modifiers. For many of the potential effect modifiers, the p-for-interaction was > 0.10 for all of the adverse outcomes; for several other potential effect modifiers, the p-for-interaction was < 0.10 for only one adverse outcome, and for that outcome the difference between SQ-LNS and IFA/SOC groups was not significant in either subgroup. For birth order, the p-for-interaction was < 0.10 for 2 outcomes: neonatal mortality and mortality 0-6 mo. In both cases, the stratum-specific estimates were not significant: neonatal and 0-6 mo mortality RRs were respectively 0.54 (0.27, 1.06) and 0.60 (0.32, 1.15) among first-born infants, and 1.08 (0.69, 1.70) and 1.22 (0.86, 1.73) among later-born infants. These results were similar in the sensitivity analyses (available at https://osf.io/nj5f9/ (17)).

#### SQ-LNS vs MMS

##### Main effects

Effects of SQ-LNS vs MMS on birth outcomes are based on 2 trials and a total sample size of 1,121-1,539 (**Table 5**). There were marginally significant differences in head circumference (+0.14 (-0.02, 0.31) cm) and HCGAZ (+0.11 (-0.01, 0.23)) favoring the SQ-LNS group, but p-values for all other outcomes were > 0.10. There was low heterogeneity, and fixed effects and random-effects models generated nearly identical estimates (**Figures 5A-C** and **Supplemental Figures 5A-AF**). We rated the quality of the evidence for all outcomes as low because data were available for only 2 trials, both conducted in Africa.

**Figures 5 A-C:**
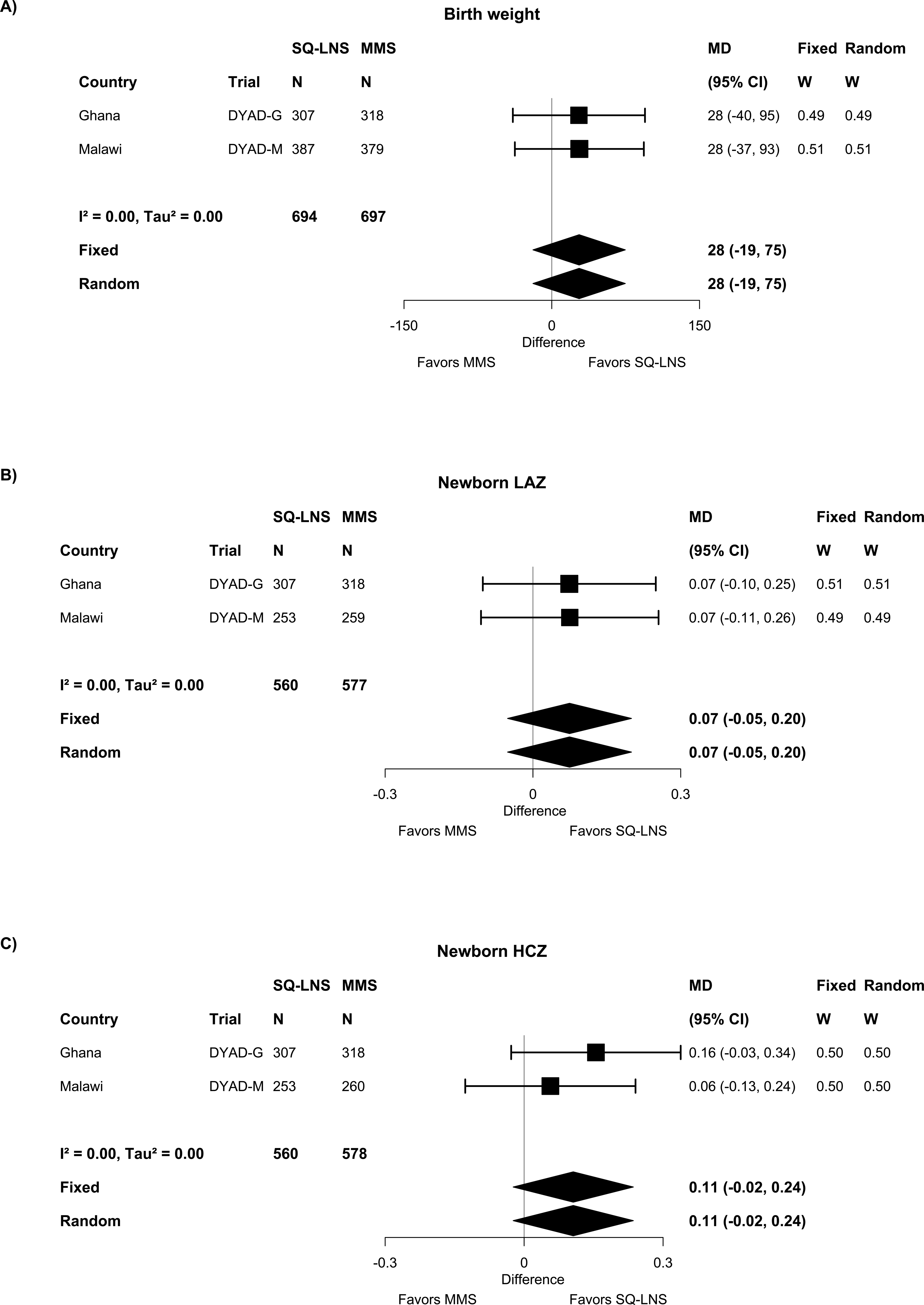
Forest plots of effect of SQ-LNS vs MMS on birth weight, newborn LAZ, and newborn HCZ. HCZ, head circumference-for-age z-score; LAZ, length-for-age z-score; LNS, lipid-based nutrient supplements; MD, mean difference; MMS, multiple micronutrient supplements; SQ-LNS, small-quantity lipid-based nutrient supplements. Individual study estimates were generated from log-binomial regression for binary outcomes and linear regression for continuous outcomes controlling for baseline measure when available and using robust standard errors for cluster-randomized trials. Pooled estimates were generated using inverse-variance weighting with both fixed and random effects.

**Table 5.** Main effects of maternal SQ-LNS vs MMS on birth outcomes^1^.

| **Birth outcome** | **Number of participants (trials)** | **SQ-LNS events per 1000** | **MMS events per 1000** | **MD or RR plus difference in events per 1000 (95% CI)<sup>2</sup>** | **P value<sup>3</sup>** | **Heterogeneity *I*<sup>2</sup>** | **Quality of the evidence (GRADE)** |
| --- | --- | --- | --- | --- | --- | --- | --- |
| Birth weight (g) | 1391 (2) | - | - | 27.8 (-19.1, 74.6) | 0.245 | 0.00 | Low |
| Weight-for-age z score (WAZ) | 1391 (2) | - | - | 0.06 (-0.05, 0.17) | 0.263 | 0.00 | Low |
| Weight-for-gestational age z-score (WGAZ) | 1377 (2) | - | - | 0.06 (-0.05, 0.16) | 0.294 | 0.00 | Low |
| Low birth weight (LBW) | 1391 (2) | 105 | 119 | 0.88 (0.66, 1.18)<br>-14.2 (-47.3, 18.8) | 0.397<br>0.399 | 0.00<br>0.00 | Low |
| Birth weight < 2 kg | 1391 (2) | 19 | 19 | 1.07 (0.52, 2.23)<br>-1.1 (-15.1, 12.9) | 0.850<br>0.877 | 0.00<br>0.00 | Low |
| Small-for-gestational age (SGA) | 1391 (2) | 214 | 224 | 0.96 (0.79, 1.17)<br>-9.3 (-52.7, 34.2) | 0.717<br>0.677 | 0.00<br>0.00 | Low |
| Large-for-gestational age (LGA) | 1391 (2) | 32 | 28 | 1.16 (0.64, 2.10)<br>5.2 (-12.6, 23.1) | 0.622<br>0.564 | 0.00<br>0.00 | Low |
| Birth length (cm) | 1137 (2) | - | - | 0.16 (-0.09, 0.40) | 0.209 | 0.00 | Low |
| Length-for-age z score (LAZ) | 1137 (2) | - | - | 0.07 (-0.05, 0.20) | 0.247 | 0.00 | Low |
| Length-for-gestational age z-score (LGAZ) | 1123 (2) | - | - | 0.08 (-0.04, 0.20) | 0.214 | 0.00 | Low |
| Newborn stunting | 1137 (2) | 118 | 110 | 1.06 (0.77, 1.47)<br>5.5 (-31.3, 42.3) | 0.710<br>0.770 | 0.00<br>0.00 | Low |
| Low LGAZ | 1137 (2) | 109 | 88 | 1.22 (0.85, 1.74)<br>11.8 (-19.6, 43.2) | 0.276<br>0.463 | 0.00<br>0.01 | Low |
| BMI-for-age z-score (BMIZ) | 1121 (2) | - | - | 0.01 (-0.12, 0.14) | 0.830 | 0.00 | Low |
| Low BMIZ | 1121 (2) | 67 | 68 | 0.99 (0.64, 1.53)<br>-1.2 (-30.8, 28.5) | 0.964<br>0.939 | 0.30<br>0.30 | Low |
| Head circumference (cm) | 1138 (2) | - | - | 0.14 (-0.02, 0.31) | 0.088 | 0.08 | Low |
| Head circumference-for-age z-score (HCZ) | 1138 (2) | - | - | 0.11 (-0.02, 0.24) | 0.109 | 0.00 | Low |
| Head circumference-for-gestational age z-score (HCGAZ) | 1124 (2) | - | - | 0.11 (-0.01, 0.23) | 0.066 | 0.00 | Low |
| Low HCZ | 1138 (2) | 56 | 43 | 1.27 (0.77, 2.10)<br>12.3 (-13.0, 37.6) | 0.352<br>0.341 | 0.00<br>0.00 | Low |
| Low HCGAZ | 1138 (2) | 16 | 17 | 0.85 (0.35, 2.05)<br>-1.6 (-16.6, 13.4) | 0.711<br>0.831 | 0.00<br>0.00 | Low |
| Mid-upper arm circumference (MUAC) (cm) | 1142 (2) | - | - | 0.08 (-0.02, 0.18) | 0.127 | 0.00 | Low |
| Duration of gestation (wk) | 1539 (2) | - | - | -0.04 (-0.27, 0.20) | 0.771 | 0.00 | Low |
| Preterm birth | 1539 (2) | 99 | 79 | 1.21 (0.88, 1.66)<br>18.4 (-10.2, 47.0) | 0.244<br>0.206 | 0.00<br>0.00 | Low |

<sup>1</sup>BMIZ, BMI-for-age z-score; GRADE, Grading of Recommendations Assessment, Development and Evaluation; HCGAZ, head circumference-for-gestational age z-score; HCZ, head circumference-for-age z-score; LAZ, length-for-age z-score; LBW, low birth weight; LGA, large-for-gestational age; LGAZ, length-for-gestational age z-score; MD, mean difference; MMS, multiple micronutrient supplements; MUAC, mid-upper arm circumference; RR, relative risk; SGA, small-for-gestational age; SQ-LNS, small-quantity lipid-based nutrient supplement; WAZ, weight-for-age z-score; WGAZ, weight-for-gestational age z-score
<sup>2</sup>For continuous outcomes, values are MDs: LNS – MMS (95% CIs). For binary outcomes, values are RRs: LNS compared with MMS (95% CIs). Difference in events per 1000 was calculated using the prevalence difference (95% CI) and multiplying by 1000.
<sup>3</sup>The P value column corresponds to the pooled main effect 2-sided superiority testing of the intervention effect estimate and 95% CI presented in the preceding column. $I^2$ describes the percentage of variability in effect estimates that may be due to heterogeneity rather than chance. Roughly, 0.3 – 0.6 may be considered moderate heterogeneity.

Most of the pre-planned sensitivity analyses were not applicable to these 2 trials, except for the one restricting birth size outcomes to data collected within 72 h of birth. In that sensitivity analysis, there was little change in the results except that the MDs for head circumference (+0.21 (-0.03, 0.45)) and HCGAZ (+0.16 (-0.01, 0.34)) became larger (though still marginally significant).

**Table 6** shows that there were no significant effects of SQ-LNS vs MMS on anthropometric outcomes at 6 mo of age. Findings were not altered when excluding the “simplified cohort follow-up” in Malawi. **Table 7** shows no significant differences in adverse outcomes for the SQ-LNS vs MMS comparison.

**Table 6.** Main effects of maternal SQ-LNS vs MMS on infant anthropometric outcomes at 6 mo of age^1^.

| Infant anthropometric outcomes at 6 mo | Number of participants (trials) | SQ-LNS events per 1000 | MMS events per 1000 | MD or PR plus difference in events per 1000 (95% CI)<sup>2</sup> | P value<sup>3</sup> | Heterogeneity *I*<sup>2</sup> | Quality of the evidence (GRADE) |
| --- | --- | --- | --- | --- | --- | --- | --- |
| Weight-for-age z-score (WAZ) | 1292 (2) | - | - | -0.01 (-0.14, 0.11) | 0.827 | 0.00 | Low |
| Underweight | 1292 (2) | 85 | 75 | 1.13 (0.78, 1.64)<br>9.5 (-20.2, 39.2) | 0.510<br>0.531 | 0.00<br>0.00 | Low |
| Length-for-age z-score (LAZ) | 1289 (2) | - | - | 0.02 (-0.10, 0.13) | 0.792 | 0.00 | Low |
| Stunted | 1289 (2) | 166 | 155 | 1.05 (0.83, 1.34)<br>13.7 (-25.0, 52.5) | 0.673<br>0.487 | 0.00<br>0.00 | Low |
| Weight-for-length z-score (WLZ) | 1288 (2) | - | - | -0.04 (-0.16, 0.09) | 0.551 | 0.00 | Low |
| Wasted | 1288 (2) | 24 | 23 | 0.97 (0.48, 1.97)<br>0.2 (-16.4, 16.9) | 0.943<br>0.977 | 0.00<br>0.00 | Low |
| Head circumference-for-age z-score (HCZ) | 1293 (2) | - | - | -0.03 (-0.13, 0.08) | 0.636 | 0.00 | Low |
| Low HCZ | 1293 (2) | 63 | 81 | 0.77 (0.52, 1.15)<br>-18.0 (-46.2, 10.2) | 0.203<br>0.211 | 0.00<br>0.00 | Low |
| MUAC-for-age z-score (MUACZ) | 1293 (2) | - | - | -0.02 (-0.14, 0.10) | 0.727 | 0.00 | Low |
| Low MUACZ [MUACZ < -2 SD or MUAC < 125 mm] | 1293 (2) | 66 | 74 | 0.92 (0.62, 1.36)<br>-6.8 (-34.6, 21.1) | 0.672<br>0.633 | 0.00<br>0.00 | Low |
| Acute malnutrition [WLZ < -2 SD or MUAC < 125 mm] | 1288 (2) | 75 | 77 | 0.98 (0.67, 1.43)<br>-1.9 (-31.0, 27.2) | 0.918<br>0.899 | 0.00<br>0.00 | Low |

<sup>1</sup>GRADE, Grading of Recommendations Assessment, Development and Evaluation; HCZ, head circumference-for-age z-score; LAZ, length-for-age z-score; MD, mean difference; MUAC, mid-upper arm circumference; MMS, multiple micronutrient supplements; MUACZ, mid-upper arm circumference-for-age z-score; PR, prevalence ratio; SQ-LNS, small-quantity lipid-based nutrient supplement; WAZ, weight-for-age z-score; WLZ, weight-for-length z-score
<sup>2</sup>For continuous outcomes, values are MDs: LNS – MMS (95% CIs). For binary outcomes, values are PRs: LNS compared with MMS (95% CIs). Difference in events per 1000 was calculated using the prevalence difference (95% CI) and multiplying by 1000.
<sup>3</sup>The P value column corresponds to the pooled main effect 2-sided superiority testing of the intervention effect estimate and 95% CI presented in the preceding column. *I*<sup>2</sup> describes the percentage of variability in effect estimates that may be due to heterogeneity rather than chance. Roughly, 0.3 – 0.6 may be considered moderate heterogeneity.

**Table 7.** Main effects of maternal SQ-LNS vs MMS on adverse outcomes^1^.

| Adverse outcomes | Number of participants (trials) | SQ-LNS events per 1000 | MMS events per 1000 | RR and difference in events per 1000 (95% CI)<sup>2</sup> | P value<sup>3</sup> | Heterogeneity $I^2$ | Quality of the evidence (GRADE) |
| --- | --- | --- | --- | --- | --- | --- | --- |
| Cesarian-Section | 1491 (2) | 129 | 103 | 1.25 (0.94, 1.66)<br>27.0 (-2.1, 56.2) | 0.125<br>0.069 | 0.00<br>0.00 | Low |
| Miscarriage | 1566 (2) | 16 | 16 | 1.00 (0.47, 2.13)<br>-0.1 (-11.3, 11.2) | 0.994<br>0.991 | 0.00<br>0.00 | Low |
| Stillbirth | 1566 (2) | 21 | 11 | 1.57 (0.61, 4.03)<br>9.9 (-2.9, 22.7) | 0.344<br>0.129 | 0.85<br>0.87 | Low |
| Miscarriage or stillbirth | 1566 (2) | 39 | 28 | 1.33 (0.76, 2.34)<br>14.7 (-2.9, 32.2) | 0.320<br>0.102 | 0.79<br>0.73 | Low |
| Early neonatal mortality | 1470 (2) | 19 | 17 | 0.96 (0.44, 2.09)<br>4.0 (-9.1, 17.2) | 0.924<br>0.550 | 0.61<br>0.59 | Low |
| Miscarriage or stillbirth or early neonatal mortality | 1566 (2) | 57 | 46 | 1.24 (0.81, 1.91)<br>11.3 (-10.7, 33.3) | 0.321<br>0.313 | 0.00<br>0.00 | Low |
| Neonatal mortality | 1473 (2) | 23 | 19 | 1.05 (0.52, 2.10)<br>4.5 (-9.9, 19.0) | 0.896<br>0.537 | 0.47<br>0.38 | Low |
| Mortality 0-6 mo | 1483 (2) | 30 | 28 | 1.00 (0.56, 1.78)<br>4.6 (-11.7, 20.9) | 0.988<br>0.584 | 0.43<br>0.33 | Low |

<sup>1</sup>GRADE, Grading of Recommendations Assessment, Development and Evaluation; MMS, multiple micronutrient supplement; RR, relative risk; SQ-LNS, small-quantity lipid-based nutrient supplement
<sup>2</sup>Values are RRs: LNS compared with MMS (95% CIs). Difference in events per 1000 was calculated using the prevalence difference (95% CI) and multiplying by 1000.
<sup>3</sup>The P value column corresponds to the pooled main effect 2-sided superiority testing of the intervention effect estimate and 95% CI presented in the preceding column. $I^2$ describes the percentage of variability in effect estimates that may be due to heterogeneity rather than chance. Roughly, 0.3 – 0.6 may be considered moderate heterogeneity.

##### Effect modification by individual-level characteristics

For most characteristics, the p-for-interaction was > 0.10 for all infant birth outcomes, i.e., effect modification was not evident for maternal height, BMI, education, or anemia at baseline, gestational age at start of supplementation, compliance with supplementation, household socio-economic status, food insecurity, or sanitation (**Supplemental Figures 6A-M**). The p-for-interaction was < 0.10 for at least 1 birth size outcome for 5 characteristics: child sex, birth order, maternal age, inflammation at baseline, and malaria at baseline. Effect modification estimates for maternal SQ-LNS vs MMS were greater (p-for-interaction < 0.10) among a) female (vs. male) infants for head circumference, HCZ, birth weight < 2 kg, SGA, low BMIZ and low HCZ (Supplemental Figures 6A1-2), b) first-born (vs. later-born) infants for birth weight and WAZ (Supplemental Figure 6B1), and c) younger women (< 25 vs. > 25 y) for SGA (Supplemental Figure 6E). Effect modification by maternal inflammation was difficult to interpret: effect estimates for maternal SQ-LNS vs. MMS were greater among women with inflammation for birth weight, but greater among those without inflammation for HCZ and HCGAZ (Supplemental Figure 6H1). Results for effect modification by maternal malaria were not interpretable because they could not be confirmed in the sensitivity analysis restricted to anthropometric outcomes measured within 72 h of birth, due to insufficient sample sizes in the subgroup with malaria at enrollment. Results of the sensitivity analysis restricted to birth size measured within 72 h for effect modification by child sex, birth order and maternal age are available at https://osf.io/nj5f9/ (17); the findings were similar but for some outcomes the sample sizes in certain subgroups were insufficient to generate effect estimates.

Some of the above characteristics also modified the effect estimates for maternal SQ-LNS vs. MMS with regard to a few of the anthropometric outcomes at 6 mo, but the results were not always consistent across outcomes (Supplemental Figures 6A3&4-6M3&4).

For most of the adverse outcomes, statistical power to examine effect modification was low because they are rare events and the total sample size for this comparison, with both trials combined, was < 1,566.

## Discussion

In this IPD meta-analysis, data from 4 trials showed that maternal SQ-LNS, compared to IFA or SOC, increased mean birth weight, length, head circumference, BMIZ, MUAC and duration of gestation, and on average reduced the incidence of LBW by 11%, newborn stunting by 17%, newborn wasting (low BMIZ) by 11%, and small head size (low HCZ) by 15%. Only 2 trials directly compared maternal SQ-LNS and MMS; birth outcomes did not differ significantly between these groups, although there was a marginally significant difference in newborn head circumference. Several individual-level characteristics appeared to modify the impact of maternal SQ-LNS. For the comparison with IFA or SOC, effect estimates for SQ-LNS were greater among female infants and among women with BMI < 20 kg/m^2^, inflammation or malaria at enrollment, or greater household food insecurity. For the comparison with MMS, effect estimates for SQ-LNS were greater among female infants, first-born infants, and women < 25 y of age. Some of these findings may have implications with regard to potential targeting of SQ-LNS to vulnerable women, as discussed below.

### Main effects on birth outcomes and adverse outcomes

For SQ-LNS vs IFA/SOC, our estimated main effects for birth outcomes are similar to those of Das et al. (13), which is expected because their analysis included 3 of the 4 trials in our IPD analysis. We report on several birth outcomes not included in the Das et al. meta-analysis: newborn BMIZ and low BMIZ, and birth size for gestational age outcomes using the INTERGROWTH-21st standard. The MD in birth weight was 49 g, which is similar to the estimated impact of other types of BEP (10). The RR of 0.89 for LBW translates into an estimated absolute difference of ∼29 events per 1000 births. For newborn stunting, wasting, and small head size, the estimated absolute differences are 32, 24 and 22 events per 1000 births, respectively. For SGA (RR 0.96 (0.92, 1.01)), the upper bound of the 95% CI slightly exceeded 1.0; in a sensitivity analysis restricted to results based on ultrasound dating of gestational age (which required exclusion of the largest trial, in Bangladesh), the RR was more substantial but the upper bound of the 95% CI did not change (0.86 (0.73, 1.01)). The RR for preterm birth was not significant in the main analysis (0.94 (0.80, 1.10)) or in the sensitivity analysis restricted to ultrasound dating (0.90 (0.68, 1.18)). The effect estimates for birth size z-scores for gestational age (+0.13 in WGAZ, +0.13 in LGAZ, and +0.11 in HCGAZ) were similar to those for WAZ (+0.12), LAZ (+0.11) and HCZ (+0.10), suggesting that the impact of SQ-LNS on birth size was mainly attributable to improvements in fetal growth.

For SQ-LNS vs MMS, we cannot directly compare our estimated main effects to those of Das et al. (13) because their analysis included 1 MQ-LNS trial in addition to the 2 SQ-LNS trials that are in our IPD meta-analysis. Although neither meta-analysis showed significant differences in birth outcomes between (SQ-)LNS and MMS groups, the MDs for all of the birth size outcomes in our IPD analysis were in the same direction as those for the SQ-LNS vs IFA/SOC analysis, though generally of lower magnitude except for the MD in head circumference (which was almost identical between the SQ-LNS vs MMS and SQ-LNS vs IFA/SOC comparisons, e.g., HCGAZ +0.11 (-0.01, 0.23) and +0.11 (0.02, 0.20), respectively).

Because only 2 trials have directly compared SQ-LNS with MMS, it is useful to examine how the estimated effects of SQ-LNS vs IFA/SOC compare with those of MMS vs IFA in previous meta-analyses, which included a large number of trials (19, 38). Effects of MMS vs IFA on mean birth weight (+48 (40, 57) g in Smith et al. (19)) and RR for LBW (0.86-0.88) (19, 38) are quite similar to those shown for SQ-LNS vs IFA/SOC herein, and the results for SGA and preterm birth are also similar. However, we have demonstrated positive effects of SQ-LNS on newborn LAZ, stunting, HCZ, and small HCZ, whereas these outcomes have not been reported in the MMS vs IFA meta-analyses. The MD for head circumference in the SQ-LNS vs MMS comparison suggests that the differences in nutrient composition between these two supplements (e.g., inclusion of EFAs, calcium in SQ-LNS) may influence certain parameters of fetal growth, even if effects on birth weight are similar.

In both the SQ-LNS vs IFA/SOC and SQ-LNS vs MMS comparisons, we did not find significant differences in incidence of Cesarian-section, miscarriage, stillbirths, early neonatal mortality, neonatal mortality or mortality from birth to 6 mo of age, although it should be noted that the statistical power to detect differences in some of these rare outcomes was particularly limited.

### Effect modification for birth outcomes

#### SQ-LNS vs IFA/SOC

The impact of SQ-LNS was greater among female than among male infants for many of the birth outcomes. Among females, effect estimates for SQ-LNS suggested reductions of 20% for LBW, 24%, for newborn stunting, 22% for newborn wasting and 17% for small head size, and an increase in duration of gestation of +0.3 (0.15, 0.45) wk. In exploratory analyses, we found that the effects on birth size were only partially explained by the increased duration of gestation, suggesting that among female fetuses, SQ-LNS influenced both of the pathways leading to SVNs, i.e., “born too soon” and “born too small” (6). It is noteworthy that child sex did not modify the effects of MMS (vs IFA) on LBW or SGA (19). It is unclear why the effects of SQ-LNS were stronger among female fetuses. In our previous IPD meta-analysis of SQ-LNS provided directly to infants and young children 6-23 mo of age, we also found a greater impact among females than among males for child stunting, wasting and small head size (15), as well as anemia (39). We interpreted this as a greater potential to respond to nutritional interventions among females than among males (15). Males are more vulnerable to environmental stressors (40, 41), and are at higher risk of morbidity and mortality in early life, which could constrain their response to nutrition interventions. Our results suggest that this vulnerability begins prior to birth and is thus likely to be biologically rather than socially driven.

For some outcomes, effects of SQ-LNS were greater among mothers with a low BMI at enrollment than among those with BMI ≥ 20 kg/m^2^. The p-for-interaction was significant only for low HCZ and mean MUAC but effect estimates were also somewhat larger among low BMI mothers for most of the other birth size outcomes (though not for duration of gestation or preterm birth). Among mothers with low BMI, effect estimates for SQ-LNS suggested reductions of 23% for newborn small head size and 20% for newborn stunting. This could be interpreted as a greater potential to benefit among infants of low BMI mothers.

Similarly, effects of SQ-LNS were greater among infants of women with inflammation or malaria at enrolment. For inflammation, the p-for-interaction was significant for WAZ, duration of gestation and small head size and there was a similar pattern for several other birth outcomes. Among women with inflammation, effect estimates for SQ-LNS suggested reductions of 33% for LBW, 31% for newborn stunting, 33% for newborn wasting and 40% for small head size. For malaria, the p-for-interaction was significant for birth weight, WAZ, LBW, and SGA. Among women with malaria, effect estimates for SQ-LNS suggested reductions of 47% for LBW and 40% for SGA. These are large relative reductions in the risk of being “born too small”, and suggest that SQ-LNS may be mitigating the adverse effects of maternal inflammation and possibly malaria on fetal growth (42, 43).

Effects of SQ-LNS were also greater among women in households with moderate-to-severe food insecurity than in those with less food insecurity. The p-for-interaction was significant for birth length, LAZ, newborn stunting, and head circumference (in cm) and there was a similar pattern for several other birth outcomes. Among women with greater food insecurity, effect estimates for SQ-LNS suggested reductions of 16% for LBW and 25% for newborn stunting. These findings suggest a greater potential to benefit among women with food insecurity, perhaps because they are at greater risk of nutrient inadequacy.

Effects of SQ-LNS on birth size outcomes tended to be greater among infants of mothers who consumed supplements ≥ 4 days/wk than in those with lower compliance, although the p-for-interaction was significant only for stunting. Among women with higher compliance, effect estimates for SQ-LNS suggested reductions of 16% for LBW, 22% for newborn stunting, 14% for newborn wasting, and 18% for small head size.

Effect modification was generally not evident for other maternal characteristics. This could be important, as it suggests that a response to maternal SQ-LNS is not constrained by short maternal stature, low education, anemia at baseline, later gestational age at start of supplementation, or low household socio-economic status. In the IPD meta-analysis of MMS vs IFA (19), there was a greater impact of MMS on SGA among women with more education, compared to those with less education. This type of interaction was not observed for SQ-LNS. The lack of interaction of SQ-LNS with maternal anemia is also noteworthy because the iron content of SQ-LNS (20 mg) was much lower than that of IFA (60 mg) in these trials. Although this difference in iron content may influence the risk of maternal iron-deficiency anemia (44–46), it does not appear to compromise the impact of maternal SQ-LNS on infant outcomes (47).

#### SQ-LNS vs MMS

As was found for the comparison of SQ-LNS vs IFA/SOC, effect estimates for SQ-LNS vs MMS were greater among female than among male infants. The p-for-interaction with infant sex was significant for several outcomes (HCZ, SGA, birth weight < 2 kg, low BMIZ, and small head size), and the effect estimates for SQ-LNS were significant among females for LGAZ (+0.17 (0.00, 0.33)), HCZ (+0.21 (0.01, 0.40)) and HCGAZ (+0.19 (0.02, 0.37)). It is noteworthy that these differences between intervention groups among females are for outcomes that reflect linear growth and head size, and may be attributable to the differences in nutrient content between SQ-LNS and MMS, particularly EFAs (important for brain development) (48) and minerals such as calcium, potassium and magnesium (important for linear growth).

Effects of SQ-LNS vs MMS appeared to be greater among first-born than among later-born infants. The p-for-interaction was significant for birth weight and WAZ, and among first-born infants the effect estimates for SQ-LNS were substantial for birth weight (+120 (35, 205) g) and HCZ (+0.31 (0.05, 0.58)). However, there was heterogeneity between the 2 sites for this interaction, with results being driven by greater effects of SQ-LNS among first-born infants in Ghana but not in Malawi.

Effects of SQ-LNS on SGA were more beneficial among younger women than among those ≥ 25 y, and there was a similar pattern for other birth outcomes. Among infants of younger mothers, effect estimates for SQ-LNS vs MMS were significant for birth weight (+69 (4, 135)), WAZ (+0.16 (0.01, 0.31)), and HCZ (+0.20 (0.00, 0.40)). This could reflect a greater potential to benefit among infants of younger mothers (who are also more likely to be first-born), among whom improved intake of EFAs and certain minerals could be more critical.

### Effects on anthropometric status at 6 mo

An important biological and programmatic question is whether prenatal supplementation has a sustained impact on infant growth status after birth. Previous evidence on this question has been mixed (49). In this meta-analysis, maternal SQ-LNS (compared to IFA or SOC) reduced the prevalence of underweight at 6 mo of age by 15%, but effects on the other anthropometric outcomes were not statistically significant. Among female infants, underweight at 6 mo was reduced by 26%, whereas no effect was observed among male infants. In 3 cohorts (Bangladesh, Ghana and half of the Malawi cohort), mothers continued to receive supplements after delivery for up to 6 mo, so it is possible that some of the impact on infant underweight at 6 mo is attributable to postpartum effects, e.g., through breast milk composition or maternal caregiving capacity. However, when we excluded the two cohorts in which mothers did not continue to receive supplements postpartum (Guatemala and half of the Malawi cohort), the effect estimate for underweight was weaker rather than stronger, suggesting that the reduction in underweight is likely related to SQ-LNS received prenatally. This is an important finding because underweight among infants is a key risk factor for mortality (50–52).

There were no significant differences in infant anthropometric status at 6 mo in the comparison of SQ-LNS vs MMS.

### Strengths and limitations

Strengths of this IPD meta-analysis include the high quality of the randomized controlled trials contributing to the estimates, data from diverse settings on 3 different continents, and the consistency in findings between fixed-effects and random-effects models as well as in most of the sensitivity analyses. Limitations include the relatively small number of trials (especially for the SQ-LNS vs MMS comparison), and limited statistical power to detect differences in rare outcomes. Results of the effect modification analyses should be interpreted with caution because many of the potential effect modifiers are interrelated and also may be confounded by other unmeasured factors. In addition, there was variation in the methods used in each study to assess certain potential effect modifiers such as household food insecurity and socio-economic status.

### Conclusions and implications

Maternal SQ-LNS has substantial positive effects on birth outcomes when compared with IFA or SOC, especially among female infants and among vulnerable women such as those with low BMI, inflammation or malaria at enrollment, or greater household food insecurity. Provision of SQ-LNS to pregnant women may also reduce the prevalence of underweight among infants at 6 mo of age. An important programmatic question is whether maternal SQ-LNS is superior to MMS, which is lower in cost and is currently being scaled-up (53, 54). Based on this meta-analysis as well as the meta-analyses of MMS (19, 38), we conclude that SQ-LNS and MMS have similar positive effects on birth weight outcomes when comparing each with IFA.

However, it is not known whether MMS has an impact on newborn LAZ, stunting, BMIZ, wasting, HCZ, or small HCZ, all of which were improved in the SQ-LNS vs IFA/SOC comparisons herein. With only 2 trials directly comparing SQ-LNS and MMS, the statistical power to detect differences between intervention groups was considerably lower than was the case for the SQ-LNS vs IFA/SOC comparison. Moreover, the MMS used in those 2 trials had 1.5-2 times the amounts of most micronutrients compared to the MMS formulation currently being scaled up (UNIMMAP), which may have reduced the likelihood of detecting differences in birth outcomes between the SQ-LNS and MMS groups. Further trials that include this comparison are needed.

It is possible that the impact of SQ-LNS on certain birth outcomes is superior to that of MMS within vulnerable populations, even if the main effects on those outcomes do not differ significantly in the general population. For example, among infants of women < 25 y of age, birth weight and head circumference were greater in the SQ-LNS group, compared to MMS. As mentioned above, effects of SQ-LNS vs IFA/SOC were largest in more vulnerable women, whereas effects of MMS (vs IFA) on birth size (LBW or SGA) have not been shown to differ in such subgroups (i.e., younger women, those with low BMI) (19). The MMS vs IFA IPD meta-analysis did not include household food insecurity or maternal inflammation as potential effect modifiers.

The large and consistent impact of SQ-LNS (vs IFA/SOC) among women with one or more biomarkers of inflammation at enrollment on several different metrics of fetal growth (ponderal, linear and head size) is noteworthy, given that the percentage of women in this subgroup was 40.8% in Ghana, 45.9% in Malawi and 16.6% in Bangladesh (this information was not available for Guatemala). These findings suggest a greater potential to benefit from maternal SQ-LNS among such women. The mechanisms underlying these findings require further investigation.

Further research is also needed to elucidate potential biological explanations for the stronger effects of maternal SQ-LNS observed among female infants, compared to males, and how to overcome the constraints on response among males. Regardless of the mechanism, the substantial reduction in risk of newborn stunting among females may have implications with regard to subsequent height during childhood, adolescence and adulthood (55, 56). Greater maternal stature in females at the time of childbearing may reduce the risk of SGA and thereby help prevent the intergenerational transmission of impaired growth.

From a programmatic perspective, the effects among vulnerable subgroups demonstrated herein suggest that targeting provision of SQ-LNS to younger women, those with low BMI, or those in households with food insecurity may be worth considering. This type of targeting is envisioned for BEP interventions in general (9), and the 2016 WHO guideline on antenatal care (57) already recommends prenatal BEP in populations with a high prevalence of undernutrition among pregnant women. Given that SQ-LNS meets the definition of BEP, our results support the strategy of targeting food-based supplements to pregnant women who have the greatest potential to benefit. SQ-LNS provides less energy than most BEP supplements that have been evaluated, but at present there is no clear dose-response relationship between the quantity of LNS (or BEP of other types) and birth outcomes (58). Further research is needed to identify the optimal energy content of LNS for pregnant women. However, it is noteworthy that the effects of SQ-LNS (vs IFA/SOC) on birth outcomes in this IPD analysis were larger among women with greater food insecurity, despite the small amount of energy provided. This implies that improving intake of essential nutrients during pregnancy in high-risk populations is of paramount importance.

## Data Availability

Data described in the manuscript, code book, and analytic code will not be made available because they are compiled from 4 different trials, and access is under the control of the investigators of each of those trials.

## Abbreviations

BEP: balanced energy protein;
BMIZ: body mass index-for-age z-score;
EFA: essential fatty acids;
GRADE: Grading of Recommendations Assessment, Development and Evaluation;
HCGAZ: head circumference-for-gestational age z-score;
HCZ: head circumference-for-age z-score;
IFA: iron and folic acid supplement;
IPD: individual participant data;
IRB: institutional review board;
LAZ: length-for-age z-score;
LBW: low birth weight;
LMIC: low-and middle-income countries;
LGA: large-for-gestational age;
LGAZ: length-for-gestational age z-score;
LNS: lipid-based nutrient supplements;
LQ-LNS: large-quantity lipid-based nutrient supplements;
MD: mean difference;
MMS: multiple micronutrient supplement;
MQ-LNS: medium-quantity lipid-based nutrient supplements;
MUAC: mid-upper arm circumference;
MUACZ: mid-upper arm circumference z-score;
PR: prevalence ratio;
RCT: randomized controlled trial;
RR: relative risk;
SAP: statistical analysis plan;
SGA: small-for-gestational age;
SOC: standard of care;
SQ-LNS: small-quantity lipid-based nutrient supplements;
SVN: small vulnerable newborn;
WAZ: weight-for-age z-score;
WGAZ: weight-for-gestational age z-score;
WLZ: weight-for-length z-score.

## Acknowledgments

We thank all of the co-investigators, collaborators, study teams, participants and local communities involved in the trials included in these analyses. These trials benefitted from the contributions of many partner organizations, including: icddr,b (Rang-Din Nutrition Study); the International Lipid-based Nutrient Supplements Project Steering Committee (iLiNS Project trials); NICHD-Global Network for Women’s and Children’s Health Research and Institute of Nutrition in Central America and Panama (INCAP) (Women First trial); and Nutriset (for development of SQ-LNS).

The authors’ responsibilities were as follows—KGD: drafted the manuscript with input from KRW, CDA, CPS and other coauthors; KRW, CDA, KGD, and CPS: wrote the statistical analysis plan; BFA, PA, LH, NFK, JL, SM: reviewed, contributed to, and approved the statistical analysis plan; KRW and CDA: compiled the data; CDA: conducted the data analysis; and all authors: read, contributed to, and approved the final manuscript; KGD is responsible for final content.

Supported by Bill & Melinda Gates Foundation grant OPP49817 (to KGD). All authors report no conflicts of interest.

---

**Box 1:** Outcome variables^1^.

Box 1: Outcome variables<sup>1</sup>
| Continuous outcomes | Categorical outcomes |
| --- | --- |
| **Birth size and duration of gestation** |  |
| Birth weight (g), weight-for-age z score (WAZ) | Low birth weight (LBW) < 2500 g (59), Birth weight < 2000 g |
| Weight-for-gestational age z-score (WGAZ) | Small-for-gestational age (SGA) < 10<sup>th</sup> percentile (24) |
|  | Large-for-gestational age (LGA) > 90<sup>th</sup> percentile (24) |
| Birth length (cm), length-for-age z score (LAZ) | Newborn stunting, LAZ < -2 SD (22) |
| Length-for-gestational age z-score (LGAZ) | Low LGAZ (LGAZ < -2 SD) (24) |
| BMI-for-age z-score (BMIZ)<sup>2</sup> | Newborn wasting, BMIZ < -2 SD (22) |
| Birth head circumference (cm), head circumference-for-age z score (HCZ) | Small head circumference, HCZ < -2 SD (23) |
| Head circumference-for-gestational age z score (HGAZ) | Low HGAZ, HGAZ < -2 SD (2) |
| Mid-upper arm circumference (MUAC, cm) |  |
| Duration of gestation (wk) | Preterm birth (duration of gestation < 37 wk) |
| **Anthropometric outcomes at 6 months of age** |  |
| Weight-for-age Z score (WAZ) | Underweight, WAZ < -2 SD (22) |
| Length-for-age Z score (LAZ) | Stunting, LAZ < -2 SD (22) |
| Weight-for-length Z score (WLZ) | Wasting, WLZ < -2 SD (22) |
| Head circumference-for-age Z score (HCZ) | Small head circumference, HCZ < -2 SD (23) |
| MUAC-for-age Z score (MUACZ) | Low MUAC, MUACZ < -2 SD or MUAC < 125 mm (23) |
|  | Acute malnutrition, WLZ < -2 SD or MUAC < 125 mm (22) |
| **Adverse outcomes** |  |
|  | Cesarian-section |
|  | Miscarriage, embryo or fetal death < 28 weeks (59) |
|  | Stillbirth, fetal death ≥ 28 weeks gestation (59) |
|  | Early neonatal mortality ≤ 7 days (59) |
|  | Neonatal mortality ≤ 28 days (59) |
|  | Infant mortality < 6 months |

<sup>1</sup> BMIZ, BMI-for-age z-score; HGAZ, head circumference-for-gestational age z-score; HCZ, head circumference-for-age z-score; LAZ, length-for-age z-score; LBW, low birth weight; LGA, large-for-gestational age; LGAZ, length-for-gestational age z-score; MUAC, mid-upper arm circumference; MUACZ, mid-upper arm circumference-for-age z-score; SGA, small-for-gestational age; WAZ, weight-for-age z-score; WGAZ, weight-for-gestational age z-score; WLZ, weight-for-length z-score
<sup>2</sup> BMIZ is used as a proxy for weight-for-length z-score because the latter is not calculated for children with lengths < 45 cm (22).

**Box 2:** Potential effect modifiers^1^.

Box 2: Potential effect modifiers<sup>1</sup>
| Individual-level child, maternal and household effect modifiers |
| --- |
| Child<br>• Sex (female vs. male)<br>• Birth order (first born vs. later born)<br>Maternal<br>• Maternal height (< 150.1 cm vs. $\geq 150.1$ cm)<sup>2</sup><br>• Maternal BMI (< 20 kg/m<sup>2</sup> vs. $\geq 20$ kg/m<sup>2</sup>)<sup>3</sup><br>• Maternal age (< 25 y vs. $\geq 25$ y)<br>• Maternal education (no formal or incomplete primary vs. complete primary or greater)<br>• Baseline anemia status (Hb $\geq 110$ g/L vs. < 110 g/L)<br>• Baseline inflammation status (CRP $\leq 5$ mg/L and AGP $\leq 1$ g/L vs. not)<br>• Baseline malaria status (positive rapid test for malaria vs. negative test)<br>• Gestational age at start of supplementation (< 14 wk vs. $\geq 14$ wk)<br>• Compliance with supplementation ($\geq 4$ d/wk vs. < 4 d/wk)<sup>4</sup><br>Household<br>• Household socio-economic status (< study median vs. ≥ study median)<sup>5</sup><br>• Household food security (moderate to severe food insecurity vs. mild to secure)<br>• Sanitation (unimproved vs. improved)<sup>6</sup> |

<sup>1</sup> Comparisons follow the format nonreference vs. reference category. AGP, $\alpha$ -1-acid glycoprotein; CRP, C-reactive protein; Hb, hemoglobin. Some potential effect modifiers that were listed in the statistical analysis plan were not included: maternal marital status was dropped because of insufficient variation; maternal depression was dropped because it was evaluated only at 6 mo postpartum; source water quality was not included because there was little variation in two of the eligible trials; season at time of conception was not included because the seasonal factors that may influence pregnancy outcomes (rainy/dry, harvest/hungry, heat stress) do not always coincide and a 9-month pregnancy involves exposure to several seasons at different stages of pregnancy.
<sup>2</sup> Cutoff is -2 SD for height at 19 years of age: https://www.who.int/growthref/hfa_girls_5_19years_z.pdf?ua=1
<sup>3</sup> When a pre-pregnancy weight measurement was not available, we estimated weight back to the 9th week of gestation using a restricted cubic spline model regressing baseline weight on gestational age at enrollment with 4 knots based on quintiles in the study's full dataset.
<sup>4</sup> Cutoff of > 4 d/wk chosen to accommodate the categorical variable for compliance used in the Bangladesh trial (32), which had only these choices: “did not take at all,” “used to take sometimes (1–3 d/wk),” “used to take almost every day (4–6 d/wk),” “used to take regularly every day,” and “other.”
<sup>5</sup> Based on a study-defined, study-specific assets index
<sup>6</sup> Improved sanitation includes flush/pour flush to piped sewer system, septic tanks or pit latrines; ventilated improved pit latrines, composting toilets or pit latrines with slabs (60); see Supplemental Table 2, based on baseline data

