## Supplemental Files for "Effects of prenatal small-quantity lipid-based nutrient supplements on pregnancy, birth and infant outcomes: a systematic review and meta-analysis of individual participant data from randomized controlled trials in low- and middle-income countries"

Dewey *et al.* (2024)

### Supplemental Methods 1: Search strategies

We used the same search terms used by Das et al. (1) to search international and regional databases for studies published since that review was completed. Database of Abstracts of Review of Effect (DARE), Popline and IndMED were not included.

**Cochrane Central Register of Controlled Trials (CENTRAL) in the Cochrane Library**

#1[mh Lipids]

#2(fatty next acid*)

#3((Docosahexaenoic or Eicosapentaenoic) next acid*)

#4(PUFA or PUFAs)

#5lipid*

#6(omega next 3*)

#7(omega next 6*)

#8(soy* or peanut or groundnut or whey or sesame or cashew or chickpea or oil*)

#9{or #1-#8}

#10[mh "Dietary Supplements"]

#11[mh "Food, fortified"]

#12((diet* or food*) near/3 (fortif* or enrich* or supplement*))

#13(complement* near/3 (food* or feed*))

#14"Ready to use"

#15"point of use"

#16(RUSF or RUTF)

#17(home* near/2 fortif*)

#18{or #10-#17}

#19#9 and #18

#20(lipid next based)

#21(lipid* near/3 supplement*)

#22(lipid* near/3 nutrient*)

#23(lipid* near/3 fortif*)

#24(lipid* near/3 formulation*)

#54(lipid* near/3 enrich*)

#26(lipid* near/3 emuls*)

#27(lipid* near/3 powder*)

#28(lipid* near/3 spread*)

#29(lipid* near/3 paste*)

#30(Nutributter* or Plumpy*)

#31(LNS or iLiNS)

#32{or #20-#31}

#33#19 or #32

#34[mh Pregnancy]

#35[mh Pregnant Women]

#36[mh Prenatal Care]

#37[mh Prenatal Care]

#38(perinatal* or peri-natal* or prenatal* or pre-natal* or antenatal* or ante-natal*)

#39pregnan*

#40trimester*

#41[mh Mothers]

#42(mother* or maternal*)

#43{or #34-#42}

#44#33 and #43 in Trials

**MEDLINE Ovid (ALL), Strategy 1**

1 exp Lipids/

2 fatty acid$.tw,kf.

3 Docosahexaenoic acid$.tw,kf.

4 Eicosapentaenoic Acid$.tw,kf.

5 PUFA$.tw,kf.

6 lipid$.tw,kf.

7 (omega 3$ or omega 6$).tw,kf.

8 (soy$ or peanut or groundnut or whey or sesame or cashew or chickpea or oil$).tw,kf.

9 or/1-8

10 Dietary Supplements/

11 Food, fortified/

12 ((diet$ or food$) adj3 (fortif$ or enrich$ or supplement$)).tw,kf.

13 (complement$ adj3 (food$ or feed$)).tw,kf.

14 “Ready to use”.tw,kf.

15 (RUSF or RUTF).tw,kf.

16 “point of use”.tw,kf.

17 (home$ adj2 fortif$).tw,kf.

18 or/10-17

19 9 and 18

20 (lipid$ adj3 nutrient$).tw,kf.

21 (lipid$ adj3 supplement$).tw,kf.

22 lipid based.tw,kf.

23 (lipid$ adj3 fortif$).tw,kf.

24 (lipid$ adj3 enrich$).tw,kf.

25 (lipid$ adj2 emuls$).tw,kf.

26 (lipid$ adj2 formulation$).tw,kf.

27 (lipid$ adj3 powder$).tw,kf.

28 (lipid adj3 spread$).tw,kf.

29 (lipid$ adj3 paste$).tw,kf.

30 (Nutributter$ or Plumpy$).tw,kf.

31 (LNS$1 or iLiNS).tw,kf.

32 or/20-31

33 19 or 32

34 Pregnancy/

35 Pregnant women/

36 Prenatal care/

37 Perinatal care/

38 (perinatal$ or peri-natal$ or prenatal$ or pre-natal$ or antenatal$ or ante-natal$).tw.kf

39 prenan$.tw.kf

40 trimester$.tw.kf

41 Mothers/

42 (mother$ or maternal$).tw.kf

43 or/34-42

44 randomised controlled trial.pt

45 controlled clinical trial.pt

46 randomi#ed.ab

47 placebo$.ab

48 drug therapy.fs

49 randomly.ab

50 trial.ab

51 groups.ab

52 or/44-51

53 exp animals/ not humans.sh.

54 52 not 53

55 33 and 43 and 54

**MEDLINE Ovid (ALL), Strategy 2**

1 fatty acid$.tw,kf.

2 Docosahexaenoic acid$.tw,kf.

3 Eicosapentaenoic Acid$.tw,kf.

4 PUFA$.tw,kf.

5 lipid$.tw,kf.

6 (omega 3$ or omega 6$).tw,kf.

7 (soy$ or peanut or groundnut or whey or sesame or cashew or chickpea or oil$).tw,kf.

8 or/1-7

9 ((diet$ or food$) adj3 (fortif$ or enrich$ or supplement$)).tw,kf.

10 (complement$ adj3 (food$ or feed$)).tw,kf.

11 “Ready to use”.tw,kf.

12 (RUSF or RUTF).tw,kf.

13 “point of use”.tw,kf.

14 (home$ adj2 fortif$).tw,kf.

15 or/9-14

16 (lipid$ adj3 nutrient$).tw,kf.

17 (lipid$ adj3 supplement$).tw,kf.

18 lipid based.tw,kf.

19 (lipid$ adj3 fortif$).tw,kf.

20 (lipid$ adj3 enrich$).tw,kf.

21 (lipid$ adj2 emuls$).tw,kf.

22 (lipid$ adj2 formulation$).tw,kf.

23 (lipid$ adj3 powder$).tw,kf.

24 (lipid adj3 spread$).tw,kf.

25 (lipid$ adj3 paste$).tw,kf.

26 (Nutributter$ or Plumpy$).tw,kf.

27 (LNS$1 or iLiNS).tw,kf.

28 or/16-27

29 8 and (15 or 28)

30 (perinatal$ or peri-natal$ or prenatal$ or pre-natal$ or antenatal$ or ante-natal$).tw.kf

31 prenan$.tw.kf

32 trimester$.tw.kf

33 (mother$ or maternal$).tw.kf

34 or/30-33

35 29 and 34

36 (random$ or control$ or group$ or cluster$ or placebo$ or trial$ or assign$ or prospectiv$ or meta-analysis or systematic review or longitudinal$).tw,kf.

37 35 and 36

**Embase**

1 ‘lipid’/exp or lipid

2 ‘fatty acid*’:ti,ab,kw

3 ‘docosahexaenoic acid*’:ti,ab,kw

4 ‘eicosapentaenoic acid*’:ti,ab,kw

5 PUFA*:ti,ab,kw

6 lipid*:ti,ab,kw

7 ‘omega 3*’:ti,ab,kw or ‘omega 6*’:ti,ab,kw

8 soy*:ti,ab,kw or peanut:ti,ab,kw or groundnut:ti,ab,kw or whey:ti,ab,kw or sesame:ti,ab,kw or cashew:ti,ab,kw or chickpea:ti,ab,kw or oil*:ti,ab,kw

9 #1 or #2 or #3 or #4 or #5 or #6 or #7 or #8

10 ‘dietary supplement’:de

11 ‘fortified food’:de

12 ((diet* or food*) NEAR/3 (fortif* or enrich* or supplement*)):ti,ab,kw

13 (complement* NEAR/3 (food* or feed*)):ti,ab,kw

14 ‘Ready to use’:ti,ab,kw

15 RUSF:ti,ab,kw or RUTF:ti,ab,kw

16 ‘point of use’:ti,ab,kw

17 (home* NEAR/2 fortif*):ti,ab,kw

18 #10 or #11 or #12 or #13 or #14 or #15 or #16 or #17

19 #9 and #18

20 ‘lipid based’:ti,ab,kw

21 (lipid* NEAR/3 nutrient*):ti,ab,kw.

22 (lipid* NEAR/3 supplement*):ti,ab,kw.

23 (lipid* NEAR/3 fortif*):ti,ab,kw.

24 (lipid* NEAR/3 enrich*):ti,ab,kw.

25 (lipid* NEAR/2 emuls*):ti,ab,kw.

26 (lipid* NEAR/2 formulation*):ti,ab,kw.

27 (Lipid* NEAR/3 powder*):ti,ab,kw.

28 (lipid NEAR/3 spread*):ti,ab,kw.

29 (lipid* NEAR/3 paste*):ti,ab,kw.

30 Nutributter*:ti,ab,kw or Plumpy*:ti,ab,kw.

31 LNS:ti,ab,kw or iLiNS:ti,ab,kw.

32 #20 or #21 or #22 or #23 or #24 or #25 or #26 or #27 or #28 or #29 or #30 or #31

33 #19 or #32

34 ‘pregnancy’/exp

35 ‘prenatal care’/exp

36 ‘perinatal care’/exp

37 perinatal*:ti,ab,kw or ‘peri natal*’:ti,ab,kw or prenatal*:ti,ab,kw or ‘pre natal*’:ti,ab,kw or antenatal*:ti,ab,kw or ‘ante natal*’:ti,ab,kw

38 pregnan*:ti,ab,kw

39 trimester*:ti,ab,kw

40 ‘mother’/exp

41 mother*:ti,ab,kw or maternal*:ti,ab,kw

42 #34 or #35 or #36 or #37 or #38 or #39 or #40 or #41

#43 #33 and #42

#44 ‘animals’/exp or ‘invertebrate’/exp or ‘animal experiment’ or ‘animal model’ or ‘animal tissue’ or ‘animal cell’ or ‘nonhuman’

#45 ‘human’ or ‘normal human’ or ‘human cell’

#46 #44 and #45

#47 #44 not #46

#48 #43 not #47

#49 ‘randomized controlled trial’

#50 ‘controlled clinical trial’

#51 ‘single blind procedure’

#52 ‘double blind procedure’

#53 ‘triple blind procedure’

#54 ‘crossover procedure’

#55 crossover:ti,ab,kw or ‘cross over’:ti,ab,kw

#56 ((singl* or doubl* or tripl* or trebl*) NEAR/1 (blind* or mask*)):ti,ab,kw

#57 ‘placebo’

#58 ‘placebo’:ti,ab,kw

#59 ‘prospetive’:ti,ab,kw

#60 ‘factorial*’:ti,ab,kw

#61 ‘random*’:ti,ab,kw

#62 ‘assign*’:ti,ab,kw

#63 ‘allocat*’:ti,ab,kw

#64 ‘volunteer*’:ti,ab,kw

#65 #49 or #50 or #51 or #52 or #53 or #54 or #55 or #56 or #57 or #58 or #59 or #60 or #61 or #62 or #63 or #64

#66 #48 and #65

**CINAHL Plus EBSCOhost (Cumulative Index to Nursing and Allied Health Literature)**

S1 MH “Lipids+”

S2 TI (lipid*) or AB (lipid*)

S3 TI(Docosahexaenoic acid*) OR AB(Docosahexaenoic acid*)

S4 TI(Eicosapentaenoic acid*) OR AB( Eicosapentaenoic acid*)

S5 TI(PUFA*) OR AB(PUFA* )

S6 TI(omega 3* or omega 6*) OR AB(omega 3* or omega 6* )

S7 TI (soy* or peanut or groundnut or whey or sesame or cashew or chickpea or oil*) or AB(soy* or peanut or groundnut or whey or sesame or cashew or chickpea or oil*)

S8 TI(fatty acid*) OR AB(fatty acid* )

S9 S1 OR S2 OR S3 OR S4 OR S5 OR S6 OR S7 OR S8

S10 MH “Dietary Supplements”

S11 MH “Dietary Supplementation”

S12 MH “Food, Fortified”

S13 TI ((diet* or food*) n3 (fortif* or enrich* or supplement*)) OR AB((diet* or food*) n3 (fortif* or enrich* or supplement*))

S14 TI (complement* n3 (food* or feed*)) or AB (complement* n3 (food* or feed*))

S15 “Ready to use”

S16 (RUSF or RUTF)

S17 “point of use”

S18 TI (home* n2 fortif*) OR AB(home* n2 fortif*)

S19 S10 OR S11 OR S12 OR S13 OR S14 OR S15 OR S16 OR S17 OR S18

S20 S9 AND S19

S21 TI (lipid based) or AB (lipid based)

S22 TI(lipid* N3 supplement*) OR AB( lipid* N3 supplement*)

S23 TI(lipid* N3 nutrient*) OR AB(lipid* N3 nutrient*)

S24 TI(lipid* N3 fortif*) OR AB (lipid* N3 fortif*)

S25 TI(lipid* N3 formulation*) OR AB(lipid* N3 formulation*)

S26 TI(lipid* N3 enrich*) OR AB(lipid* N3 enrich* )

S27 TI(lipid* N3 emuls*) OR AB(lipid* N3 emuls*)

S28 TI(lipid* N3 powder*) OR AB(lipid* N3 powder*)

S29 TI(lipid N3 spread*) OR AB(lipid N3 spread*)

S30 TI(lipid* N3 paste*) OR AB(lipid* N3 paste*)

S31 TI(Nutributter* or Plumpy*) or AB(Nutributter* or Plumpy*)

S32 Nutributter* or Plumpy*

S33 TI(LNS or iLiNS) OR AB( LNS or iLiNS)

S34 S21 OR S22 OR S23 OR S24 OR S25 OR S26 OR S27 OR S28 OR S29 OR S30 OR S31 OR S32 OR S33

S35 S20 OR S34

S36 MH "Pregnancy+"

S37 MH "Pregnancy Trimesters+"

S38 MH "Prenatal Care"

S39 MH "Perinatal Care"

S40 TI(perinatal* or peri-natal* or prenatal* or pre-natal* or antenatal* or ante-natal*) OR AB(perinatal* or peri-natal* or prenatal* or pre-natal* or antenatal* or ante-natal*)

S41 pregnan*

S42 trimester*

S43 MH "Mothers"

S44 TI(mother* or maternal*) or AB(mother* or maternal*)

S45 S36 or S37 or S38 or S39 or S40 or S41 or S42 or S43 or S44

S46 S35 and S45

S47 MH "Clinical Trials+"

S48 MH random assignment

S49 MH "meta-analysis"

S50 MH "crossover design"

S51 MH "Quantitative studies"

S52 PT randomized controlled trial

S53 PT clinical trial

S54 (trial* or control* or placebo*)

S55 ("follow-up study" or "follow-up research*)

S56 (prospective* study or prospect* research)

S57 (evaluat* N2 study or evaluat* N2 research)

S58 MH “Program Evaluation”

S59 MH “Treatment Outcomes”

S60 TI(single N2 mask* or single N2 blind*) OR AB(single N2 mask* or single N2 blind*)

S61 TI(doubl* N2 mask* or doubl* N2 blind*) OR AB(doubl* N2 mask* or doubl* N2 blind*)

S62 TI(tripl* N2 mask* or tripl* N2 blind or trebl* N2 mask or trebl* N2 blind) or AB(tripl* N2 mask* or tripl* N2 blind or trebl* N2 mask or trebl* N2 blind)

S63 random*

S64 S47 or S48 or S49 or S50 or S51 or S52 or S53 or S54 or S55 or S56 or S57 or S58 or S59 or S60 or S61 or S62 or S63

S65 S46 and S64

**Web of Science: Science Citation Index (SCI), Social Sciences Citation Index (SSCI), Conference Proceedings Citation Index - Science (CPCI-S) and Conference Proceedings Citation Index -Social Science & Humanities (CPCI-SS&H)**

TS=((lipid* OR "fatty acid*" OR ((Docosahexaenoic or Eicosapentaenoic) next acid*) or PUFA OR PUFAs OR "omega 3*" OR "omega 6*" OR soy* OR peanut* OR groundnut* OR whey* OR sesame* OR cashew* OR chickpea* OR oil*))

TS=(((diet* or food*) near/3 (fortif* or enrich* or supplement*)))

TS=((complement* near/3 (food* or feed*)))

TS=(("Ready to use" or "point of use" or RUSF or RUTF or (home* near/2 fortif*)))

#2 or #3 or #4

#1 and #5

TS=(("lipid based"))

TS=((lipid* near/3 (supplement* or nutrient* or fortif* or formulation* or enrich* or emuls* or powder* or spread* or paste*)))

TS=((Nutributter* or Plumpy* OR LNS or iLiNS))

#7 or #8 or #9

#6 or #10

TS=((perinatal* or peri-natal* or prenatal or pre-natal* or antenatal* or ante-natal* or pregnan* or trimester*))
TS=((mother* or maternal*))

#12 or #13

#11 and #14

TS=((RANDOM* OR TRIAL* OR CONTROL* OR PLACEBO* OR PROSPECTIV* OR LONGITUDINAL OR BLIND* OR GROUP* OR CLUSTER* OR meta-analysis OR systematic review))

#15 and #16

TS=((RATS OR MICE OR SHEEP OR PIGS OR COWS OR CHICKS OR CHICKENS OR DUCK*))

#17 not #18

**Cochrane Database of Systematic Reviews (CDSR), part of the Cochrane Library**

#1[mh Lipids]

#2 (fatty next acid*):ti,ab

#3((Docosahexaenoic or Eicosapentaenoic) next acid*):ti,ab

#4(PUFA or PUFAs):ti,ab

#5lipid*:ti,ab

#6(omega next (3* or 6*)):ti,ab

#7(soy* or peanut or groundnut or whey or sesame or cashew or chickpea or oil*):ti,ab

#8{or #1-#7}

#9[mh "Dietary Supplements"]

#10[mh "Food, fortified"]

#11((diet* or food*) near/3 (fortif* or enrich* or supplement*)):ti,ab

#12(complement* near/3 (food* or feed*)):ti,ab

#13"Ready to use":ti,ab

#14"point of use":ti,ab

#15(RUSF or RUTF):ti,ab

#16(home* near/2 fortif*):ti,ab

#17(2-#16)

#18#8 and #17

#19(lipid next based):ti,ab

#20(lipid* near/3 supplement*):ti,ab

#21(lipid* near/3 nutrient*):ti,ab

#22(lipid* near/3 fortif*):ti,ab

#23(lipid* near/3 formulation*):ti,ab

#24(lipid* near/3 enrich*):ti,ab

#25(lipid* near/3 emuls*):ti,ab

#26(lipid* near/3 powder*):ti,ab

#27(lipid* near/3 spread*):ti,ab

#28(lipid* near/3 paste*):ti,ab

#29(Nutributter* or Plumpy*):ti,ab

#30(LNS* or iLiNS):ti,ab

#31{or #19-#30}

#32#18 or #31

#33[mh “Pregnancy”]

#34[mh “Pregnant Women”]

#35[mh “Prenatal Care”]

#36[mh “Perinatal Care”]

#37(perinatal* or peri-natal* or prenatal* or pre-natal* or antenatal* or ante-natal*):ti,ab

#38pregnan*:ti,ab

#39trimester*:ti,ab

#40[mh “Mothers”]

#41(mother* or maternal*):ti,ab

#42{or #33-#41}

**Epistemonikos (epistemonikos.org)**

(title:((title:(LIPID* OR FATTY ACID* OR OMEGA OR Docosahexaenoic OR Eicosapentaenoic OR soy* OR peanut OR groundnut OR whey OR sesame OR cashew OR chickpea OR oil*) OR abstract:(LIPID* OR

FATTY ACID* OR OMEGA OR Docosahexaenoic OR Eicosapentaenoic OR soy* OR peanut OR groundnut OR whey OR sesame OR cashew OR chickpea OR oil*))) OR abstract:((title:(LIPID* OR FATTY ACID* OR OMEGA OR Docosahexaenoic OR Eicosapentaenoic OR soy* OR peanut OR groundnut OR whey OR sesame OR cashew OR chickpea OR oil*) OR abstract:(LIPID* OR FATTY ACID* OR OMEGA OR Docosahexaenoic OR Eicosapentaenoic OR soy* OR peanut OR groundnut OR whey OR sesame OR cashew OR chickpea OR oil*)))) AND (title:(fortif* OR enrich* OR supplement* OR "Ready to use" OR "point of use" OR RUSF OR RUTF) OR abstract:(fortif* OR enrich* OR supplement* OR "Ready to use" OR "point of use" OR RUSF OR RUTF)) AND (title:(PREGNAN* OR perinatal* OR peri-natal* OR prenatal* OR pre-natal* OR antenatal* OR ante-natal* OR trimester* OR MOTHER* OR MATERNAL*) OR abstract:(PREGNAN* OR perinatal* OR peri-natal* OR prenatal* OR pre-natal* OR antenatal* OR ante-natal* OR trimester* OR MOTHER* OR MATERNAL*))

##### ClinicalTrials.gov (clinicaltrials.gov)

CONDITION| pregnancy OR prenatal OR pre-natal OR antenatal OR ante-natal OR perinatal OR peri-natal OR mothers OR maternal

AND

INTERVENTION| lipid OR lipid-based OR LNS OR iLiNS OR nutrient supplement OR fortification OR fortified OR “ready to use” OR “point of use” OR RUSF OR RUTF OR “therapeutic food” OR paste OR spread OR blend OR Nutributter OR Plumpynut

##### World Health Organization International Clinical Trials Registry Platform (WHO ICTRP; who.int/trialsearch)

CONDITION| pregnancy OR prenatal OR pre-natal OR antenatal OR ante-natal OR perinatal OR peri-natal OR mothers OR maternal

AND

INTERVENTION| lipid OR lipid-based OR LNS OR iLiNS OR nutrient supplement OR fortification OR fortified OR “ready to use” OR “point of use” OR RUSF OR RUTF OR “therapeutic food” OR paste OR spread OR blend OR Nutributter OR Plumpynut

AND

RECRUITMENT STATUS|All

##### IBECS (Índice Bibliográfico Español en Ciencias de la Salud; ibecs.isciii.es)

WORD| lipid OR lipid-based OR LNS OR iLiNS OR nutrient supplement OR fortification OR fortified OR “ready to use” OR “point of use” OR RUSF OR RUTF OR “therapeutic food” OR paste OR spread OR blend OR Nutributter OR Plumpy OR Plumpynut

AND

WORD| pregnancy OR prenatal OR pre-natal OR antenatal OR ante-natal OR perinatal OR peri-natal OR mothers OR maternal

**SciELO (Scientific Electronic Library Online; www.scielo.br)**

(lipid or lipid-based OR LNS OR iLiNS OR nutrient supplement OR fortification OR fortified OR "ready to use" OR "point of use" OR RUSF OR RUTF OR "therapeutic food" OR paste OR spread OR blend OR nutributter OR Plumpy OR PLUMPYNUT) AND (pregnancy OR prenatal OR pre-natal OR antenatal OR ante-natal OR perinatal OR peri-natal OR mothers OR maternal)

**AIM (Africa Index Medicus; search.bvsalud.org/ghl/?lang=en&submit=Search&where=REGIONAL)**

(tw:(lipid OR lipid-based OR LNS OR iLiNS OR nutrient supplement OR fortification OR fortified OR "ready to use" OR "point of use" OR RUSF OR RUTF OR "therapeutic food" OR paste OR spread OR blend OR nutributter OR Plumpy OR PLUMPYNUT)) AND (tw:(pregnancy OR prenatal OR pre-natal OR antenatal OR ante-natal OR perinatal OR peri-natal OR mothers OR maternal))

**IMEMR (Index Medicus for the Eastern Mediterranean Region;**

**search.bvsalud.org/ghl/?lang=en&submit=Search&where=REGIONAL)**

(tw:(lipid OR lipid-based OR LNS OR iLiNS OR nutrient supplement OR fortification OR fortified OR "ready to use" OR "point of use" OR RUSF OR RUTF OR "therapeutic food" OR paste OR spread OR blend OR nutributter OR Plumpy OR PLUMPYNUT)) AND (tw:(pregnancy OR prenatal OR pre-natal OR antenatal OR ante-natal OR perinatal OR peri-natal OR mothers OR maternal))

**LILACS (Latin American and Caribbean Health Sciences Literature; lilacs.bvsalud.org/en)**

(tw:(lipid OR lipid-based OR LNS OR iLiNS OR nutrient supplement OR fortification OR fortified OR "ready to use" OR "point of use" OR RUSF OR RUTF OR "therapeutic food" OR paste OR spread OR blend OR nutributter OR Plumpy OR PLUMPYNUT)) AND (tw:(pregnancy OR prenatal OR pre-natal OR antenatal OR ante-natal OR perinatal OR peri-natal OR mothers OR maternal))

**PAHO/WHO Institutional Repository for Information Sharing (iris.paho.org/xmlui)**

(lipid or lipid-based OR LNS OR iLiNS OR nutrient supplement OR fortification OR fortified OR "ready to use" OR "point of use" OR RUSF OR RUTF OR "therapeutic food" OR paste OR spread OR blend OR nutributter OR Plumpy OR PLUMPYNUT) AND (pregnancy OR prenatal OR pre-natal OR antenatal OR ante-natal OR perinatal OR peri-natal OR mothers OR maternal)

**WPRIM (Western Pacific Index Medicus; search.bvsalud.org/ghl/?lang=en&submit=Search&where=REGIONAL)**

(tw:(lipid OR lipid-based OR LNS OR iLiNS OR nutrient supplement OR fortification OR fortified OR "ready to use" OR "point of use" OR RUSF OR RUTF OR "therapeutic food" OR paste OR spread OR blend OR nutributter OR Plumpy OR PLUMPYNUT)) AND (tw:(pregnancy OR prenatal OR pre-natal OR antenatal OR ante-natal OR perinatal OR peri-natal OR mothers OR maternal))

**IMSEAR (Index Medicus for the South-East Asian Region; search.bvsalud.org/ghl/?lang=en&submit=Search&where=REGIONAL)**

(tw:(lipid OR lipid-based OR LNS OR iLiNS OR nutrient supplement OR fortification OR fortified OR "ready to use" OR "point of use" OR RUSF OR RUTF OR "therapeutic food" OR paste OR spread OR blend OR nutributter OR Plumpy OR PLUMPYNUT)) AND (tw:(pregnancy OR prenatal OR pre-natal OR antenatal OR ante-natal OR perinatal OR peri-natal OR mothers OR maternal))

**Native Health Research Database (nativehealthdatabase.net)**

Keywords: (Supplement AND pregnancy)

**Online Supporting Material**

Effects of prenatal small-quantity lipid-based nutrient supplements on pregnancy, birth and infant outcomes: a systematic review and meta-analysis of individual participant data from randomized controlled trials in low-income and middle-income countries

Dewey *et al.* (2024)

**Supplemental Table 1. Composition of nutrient supplements**

| | iLiNS Project SQ-LNS formulation[1] | Women First SQ-LNS formulation[2] | IFA formulation[1] | MMS formulation[3] |
|---|---|---|---|---|
| Ration (g/day) | 20 | 20 | 1 tablet | 1 tablet |
| Total energy (kcal) | 118 | 118 | - | 0 |
| Protein (g) | 2.6 | 2.6 | - | 0 |
| Fat (g) | 10 | 10 | - | 0 |
| Linoleic acid (g) | 4.59 | 4.59 | - | 0 |
| α-Linolenic acid (g) | 0.59 | 0.59 | - | 0 |
| Vitamin A (µg RE) | 800 | 800 | - | 800 |
| Vitamin C (mg) | 100 | 100 | - | 100 |
| Vitamin B1 (mg) | 2.8 | 2.8 | - | 2.8 |
| Vitamin B2 (mg) | 2.8 | 2.8 | - | 2.8 |
| Niacin (mg) | 36 | 36 | - | 36 |
| Folic acid (µg) | 400 | 400 | 400 | 400 |
| Pantothenic acid (mg) | 7 | 7 | - | 7 |
| Vitamin B6 (mg) | 3.8 | 3.8 | - | 3.8 |
| Vitamin B12 (µg) | 5.2 | 5.2 | - | 5.2 |
| Vitamin D (IU) 1 IU = 0.025 ug | 400 | 1000 | - | 400 |
| Vitamin E (mg) | 20 | 20 | - | 20 |
| Vitamin K (µg) | 45 | 45 | - | 45 |
| Iron (mg) | 20 | 20 | 60 | 20 |
| Zinc (mg) | 30 | 15 | - | 30 |
| Copper (mg) | 4 | 4 | - | 4 |
| Calcium (mg) | 280 | 280 | 0 | 0 |
| Phosphorus (mg) | 190 | 190 | - | 0 |
| Potassium (mg) | 200 | 200 | - | 0 |
| Magnesium (mg) | 65 | 65 | - | 0 |
| Selenium (µg) | 130 | 130 | - | 130 |
| Iodine (µg) | 250 | 250 | - | 250 |
| Manganese (mg) | 2.6 | 2.6 | - | 2.6 |

iLiNS, International Lipid-Based Nutrient Supplements Project; IFA, iron and folic acid; MMS, multiple micronutrient supplements; SQ-LNS, small-quantity lipid-based nutrient supplements

[1]Provided by Mridha 2016 (32), Adu-Afarwuah 2015 (33), Ashorn 2015 (34). iLiNS (International Lipid-based Nutrient Supplements) Project SQ-LNS formulation described in Arimond et al. 2015 (11).

[2]Provided by Hambidge 2019 (35).

[3]Provided by Adu-Afarwuah 2015 (33) and Ashorn 2015 (34).

**Online Supporting Material**

Effects of prenatal small-quantity lipid-based nutrient supplements on pregnancy, birth and infant outcomes: a systematic review and meta-analysis of individual participant data from randomized controlled trials in low-income and middle-income countries

Dewey *et al.* (2024)

**Supplemental Table 2: Descriptive information on maternal, child and household characteristics at baseline, by trial**

| Country | First author, year | Child sex, male (%) | Child birth order, first born (%) | Maternal height < 150.1 cm (%)[1] | Maternal BMI < 20 $kg/m^2$ (%)[2] | Maternal age < 25 y (%) | Maternal education, completed primary (%) | Maternal anemia, Hb < 110 g/L (%) |
|---|---|---|---|---|---|---|---|---|
| Bangladesh | Mridha, 2016 (32) | 50.2% | 39.7% | 46.6% | 53.9% | 72.4% | 72.7% | 29.2% |
| Ghana | Adu Afarwuah, 2015 (33) | 50.9% | 36.8% | 4.6% | 16.3% | 40.4% | 78.5% | 38.8% |
| Malawi | Ashorn, 2015 (34) | 49.5% | 20.6% | 13.9% | 48.2% | 51.1% | 15.2% | 44.1% |
| Guatemala | Hambidge, 2019 (35) | 50.1% | 6.0% | 83.0% | 6.7% | 53.1% | 91.8% | - |

| Country | Maternal inflammation, CRP > 5 mg/dL or AGP > 1 mg/mL (%) | Maternal malaria (%) | Gestational age at start of supplementation < 14 wk (%) | Compliance with supplementation, $\geq$ 4 d/wk | SES index, below median (%)[3] | Moderate to severe food insecurity (%) | Improved sanitation access (%)[4] |
|---|---|---|---|---|---|---|---|
| Bangladesh | 16.6% | - | 61.2% | 84.6% | 50.0% | 38.2% | 70.7% |
| Ghana | 40.8% | 9.4% | 23.6% | 81.0% | 51.7% | 29.7% | 96.5% |
| Malawi | 45.9% | 22.7% | 13.1% | 92.5% | 57.0% | 66.2% | 8.7% |
| Guatemala | - | - | 87.1% | - | 50.3% | - | 44.3% |

AGP, α-1-acid glycoprotein; CRP, C-reactive protein; Hb, hemoglobin; SES, socio-economic status

[1]Cutoff is -2 SD for height at 19 years of age: https://www.who.int/growthref/hfa_girls_5_19years_z.pdf?ua=1

[2]When a pre-pregnancy weight measurement was not available, we estimated weight back to the 9th week of gestation using a restricted cubic spline model regressing baseline weight on gestational age at enrollment with 4 knots based on quintiles in the study's full dataset.

[3]Based on a study-defined, study-specific assets index

.

**Online supporting material**

Effects of prenatal small-quantity lipid-based nutrient supplements on pregnancy, birth and infant outcomes: a systematic review and meta-analysis of individual participant data from randomized controlled trials in low-income and middle-income countries

Dewey *et al.* (2024)

**Supplemental Table 3: Birth outcomes among IFA/SOC groups, by trial and pooled estimates[1]**

| Country | First author, year | Birth weight (g) | WAZ[2] | WGAZ[3] | Low birth weight (%)[4] | Birthweight < 2 kg (%) | SGA (%)[5] |
|---|---|---|---|---|---|---|---|
| Bangladesh | Mridha, 2016 (32) | 2609 (414) | -1.59 (1.02) | - | 37.1 | 7.4 | 59.2 |
| Ghana | Adu Afarwuah, 2015 (33) | 2970 (445) | -0.74 (1.02) | -0.57 (0.99) | 13.8 | 2.3 | 22.3 |
| Malawi | Ashorn, 2015 (34) | 2942 (443) | -0.79 (1.03) | -0.67 (0.96) | 13.7 | 3.1 | 25.6 |
| Guatemala | Hambidge, 2019 (35) | 2871 (332) | -0.96 (0.77) | -0.82 (0.74) | 14.4 | 0.9 | 26.5 |
| Pooled estimate | | 2847 | -1.02 | -0.69 | 19.8 | 3.5 | 33.5 |

| Country | LGA (%)[6] | Birth length (cm) | LAZ[2] | LGAZ[3] | Newborn stunting (%)[7] | Low LGAZ (%)[8] | BMIZ[2] | Low BMIZ (%)[9] |
|---|---|---|---|---|---|---|---|---|
| Bangladesh | 0.3 | 47.1 (2.2) | -1.27 (1.15) | - | 23.3 | 20.7 | -1.64 (1.03) | 33.6 |
| Ghana | 3.9 | 48.1 (1.9) | -0.73 (1.01) | -0.52 (0.97) | 9.8 | 4.9 | -0.67 (1.10) | 10.8 |
| Malawi | 3.4 | 47.5 (2.2) | -1.04 (1.19) | -0.93 (1.16) | 16.7 | 17.5 | -0.27 (1.10) | 7.1 |
| Guatemala | 0.0 | 47.5 (1.7) | -1.08 (0.87) | -0.85 (0.80) | 15.3 | 7.8 | -0.62 (0.83) | 6.5 |
| Pooled estimate | 2.3 | 47.6 | -1.04 | -0.76 | 16.4 | 12.7 | -0.80 | 14.5 |

| Country | Head circumference (cm) | HCZ[2] | HCGAZ[3] | Low HCZ (%)[10] | Low HCGAZ (%)[11] | MUAC (cm) | Duration of gestation (wk) | Preterm birth (%)[12] |
|---|---|---|---|---|---|---|---|---|
| Bangladesh | 32.5 (1.4) | -1.34 (1.12) | - | 24.9 | 18.5 | 9.7 (0.8) | 39.3 (2.3) | 13.8 |
| Ghana | 33.6 (1.4) | -0.45 (1.11) | -0.10 (1.06) | 7.9 | 3.3 | 10.4 (0.9) | 39.2 (1.9) | 9.8 |
| Malawi | 34.0 (1.4) | -0.16 (1.11) | 0.14 (1.02) | 6.1 | 0.8 | 10.4 (1.0) | 39.0 (3.0) | 11.9 |
| Guatemala | 33.3 (1.2) | -0.71 (0.94) | -0.29 (0.9) | 7.9 | 3.0 | - | 38.8 (1.7) | 7.9 |
| Pooled estimate | 33.4 | -0.67 | -0.08 | 11.8 | 6.4 | 10.2 | 39.1 | 11.1 |

BMIZ, BMI-for-age z-score; HCZ, head circumference-for-age z-score; HCGAZ, head circumference-for-gestational age z-score; IFA, iron and folic acid supplements; LAZ, length-for-age z-score; LGA, large for gestational age; LGAZ, length-for-gestational-age z-score; MUAC, mid-upper arm circumference; SGA, small for gestational age; SOC, standard of care; WAZ, weight-for-age z-score; WGAZ, weight-for-gestational-age z-score

[1]Values are mean (SD) or prevalence (%)

[2]WAZ, LAZ, BMIZ, HCAZ are based on WHO Growth Standards (22,23). BMIZ is used as a proxy for weight-for-length z-score because the latter is not calculated for children with lengths < 45 cm (22)

[3]WGAZ, LGAZ, HCGAZ are based on INTERGROWTH-21st Standards (24)

[4]Low birth weight defined as weight < 2.5 kg at birth (59)

[5]Small-for-gestational age defined as < 10th percentile weight-for-gestational age (INTERGROWTH-21st Standards) (24)

[6]Large-for-gestaional age defined as > 90th percentile weight-for-gestational age (INTERGROWTH -21st Standards) (24)

[7]Newborn stunting defined as LAZ < -2 SD (WHO Growth Standards) (22)

[8]Low length-for-gestational age z-score defined as LGAZ < -2 SD (INTERGROWTH-21st Standards)(24)

[9']Low BMI z-score defined as BMIZ < - 2SD (WHO growth standards) (22)

[10]Low head circumference-for-age z-score defined as HCZ < -2 SD (WHO growth standards) (23)

[11]Low head circumference-for-gestational age z-score defined as HCGAZ < -2 SD (INTERGROWTH-21st Standards)(24)

[12]Preterm birth defined as birth at < 37 weeks gestation (59)

**Online supporting material**

Effects of prenatal small-quantity lipid-based nutrient supplements on pregnancy, birth and infant outcomes: a systematic review and meta-analysis of individual participant data from randomized controlled trials in low-income and middle-income countries

Dewey *et al.* (2024)

**Supplemental Table 4: Anthropometric outcomes at 6 mo among IFA/SOC groups, by trial and pooled estimates[1]**

| Country | First author, year | WAZ[2] | Underweight, WAZ < -2 SD (%)[3] | LAZ[2] | Stunted, LAZ < -2 SD (%)[4] | WLZ[2] | Wasted, WLZ < -2 SD (%)[5] |
|---|---|---|---|---|---|---|---|
| Bangladesh | Mridha, 2016 (citation) | -1.13 (1.07) | 19.8 | -1.32 (1.04) | 24.8 | -0.33 (1.00) | 4.7 |
| Ghana | Adu Afarwuah, 2015 (citation) | -0.55 (1.13) | 8.8 | -0.75 (0.99) | 10.2 | -0.05 (1.13) | 5.1 |
| Malawi | Ashorn, 2015 (citation) | -0.56 (1.14) | 9.1 | -1.22 (1.17) | 23.6 | 0.36 (1.17) | 1.6 |
| Guatemala | Hambidge, 2019 (citation) | -0.89 (1.04) | 15.0 | -1.44 (1.04) | 29.9 | 0.13 (0.96) | 1.9 |
| Pooled estimate | | -0.79 | 13.2 | -1.18 | 21.9 | 0.02 | 3.2 |

| Country | HCZ[2] | Low HCZ (%)[6] | MUACZ[2] | Low MUAC (%)[7] | Acute malnutrition (%)[8] |
|---|---|---|---|---|---|
| Bangladesh | -1.42 (0.91) | 25.2 | -0.16 (0.90) | 6.1 | 8.2 |
| Ghana | -0.78 (0.93) | 8.4 | -0.17 (1.00) | 8.4 | 8.5 |
| Malawi | -0.36 (1.05) | 5.2 | 0.09 (1.13) | 7.7 | 8.2 |
| Guatemala | -0.79 (1.03) | 10.9 | - | - | - |
| Pooled estimate | -0.84 | 12.5 | -0.08 | 6.4 | 8.2 |

HCAZ, head circumference-for-age z-score; IFA, iron and folic acid supplements; LAZ, length-for-age z-score; MUACZ, mid-upper arm circumference-for-age z-score; SOC, standard of care; WAZ, weight-for-age z-score; WLZ, weight-for-length z-score

[1]Values are mean (SD) or prevalence (%)

[2]WAZ, LAZ, WLZ, HCAZ and MUACZ are based on WHO Growth Standards (22,23).

[3]Underweight defined as WAZ < -2 SD (WHO Growth Standards (22)

[4]Stunting defined as LAZ < -2 SD (WHO Growth Standards) (22)

[5]Wasting defined as WLZ < - 2SD (WHO growth standards) (22)

[6]Low head circumference-for-age z-score defined as HCZ < -2 SD (WHO growth standards) (23)

[7]Low MUAC defined as MUACZ < -2 SD or MUAC < 12.5 cm (WHO growth standards) (23)

[8]Acute malnutrition defined as WLZ < -2 SD (WHO Growth Standards) (22) or MUAC < 125 mm

**Online supporting material**

Effects of prenatal small-quantity lipid-based nutrient supplements on pregnancy, birth and infant outcomes: a systematic review and meta-analysis of individual participant data from randomized controlled trials in low-income and middle-income countries

Dewey *et al.* (2024)

**Supplemental Table 5: Adverse outcomes among IFA/SOC groups, by trial and pooled estimates**

| Country | First author, year | Miscarriage (%)[1] | Stillbirth (%)[2] | Cesarean-Section (%) | Early neonatal mortality (≤ 7 d) (%) | Neonatal mortality (≤ 28 d) (%) |
|---|---|---|---|---|---|---|
| Bangladesh | Mridha, 2016 (citation) | 6.0 | 2.4 | 15.5 | 2.5 | 2.9 |
| Ghana | Adu Afarwuah, 2015 (citation) | 3.7 | 2.6 | 18.3 | 0.9 | 1.2 |
| Malawi | Ashorn, 2015 (citation) | 0.2 | 2.1 | 4.2 | 3.9 | 4.6 |
| Guatemala | Hambidge, 2019 (citation) | 14.8 | 0.4 | 27.0 | 1.3 | 1.8 |
| Pooled estimate | | 5.9 | 1.8 | 15.6 | 2.00 | 2.5 |

| **Country** | **Mortality 0-6 mo (%)** | **Stillbirth or early neonatal mortality (%)** | **Stillbirth or miscarriage (%)** | **Stillbirth or miscarriage and early neonatal mortality (%)** |
|---|---|---|---|---|
| Bangladesh | 3.8 | 4.6 | 8.4 | 10.6 |
| Ghana | 1.5 | 3.4 | 6.3 | 7.2 |
| Malawi | 5.0 | 5.8 | 2.3 | 6 |
| Guatemala | 2.7 | 1.5 | 15.2 | 16.3 |
| Pooled estimate | 3.2 | 3.7 | 7.7 | 9.7 |

IFA; iron and folic acid supplements; SOC, standard of care

[1]Miscarriage defined as embryo or fetal death < 28 weeks gestation (59)˙ In Bangladesh, Ghana and Malawi, mean gestational age at enrollment was ~13-17 weeks. In Guatemala, study participants were enrolled pre-conceptually. The higher prevalence of miscarriages in Guatemala reflect this difference in study design.

[2]Stillbirth defined as fetal death $\geq$ 28 weeks gestation (59)

Online Supplemental Material

Effects of prenatal small-quantity lipid-based nutrient supplements on pregnancy, birth and infant outcomes: a systematic review and meta-analysis of individual participant data from randomized controlled trials in low-income and middle-income countries

Dewey *et al.* (2024)

**Supplemental Table 6: Risk of bias assessment in each trial**

| **Country** | **Author** | **Random sequence generation** | **Allocation concealment** | **Blinding participants** | **Outcome assessment[1]** | **Incomplete outcome** | **Selective reporting** | **Other** |
|---|---|---|---|---|---|---|---|---|
| Bangladesh | Mridha 2016 (32) | low | low | high | low | low | low | low |
| Ghana | Adu Afarwuah 2015 (33) | low | low | high | low | low | low | low |
| Malawi | Ashorn 2015 (34) | low | low | high | low | low | low | low |
| Guatemala | Hambidge 2019 (35) | low | low | high | unclear | low | low | low |

[1]Due to the nature of the intervention, blinding of participants was not possible. We considered birth outcome assessment to be at low risk of bias only when it was clearly specified that data collectors who performed the anthropometric measurements were not aware of group allocation, and it would be unlikely that they could easily become aware of group allocation (i.e. observation of interventions materials in study communities, non-intervention passive control arms, etc.)

| Adu-Afarwuah 2016 (33) | | |
|---|---|---|
| **Bias** | **Authors' judgement** | **Support for judgement** |
| Random sequence generation (selection bias) | Low risk | **Quote**: "women were randomly allocated into one of 3 groups by using a computer-generated scheme (SAS version 9.3; SAS Institute) in blocks of 9…the study statistician at UC Davis, who designed the randomization scheme"<br>**Comment:** adequately done |
| Allocation concealment (selection bias) | Low risk | **Quote**: "Sheets bearing supplement allocations represented by 6 different color codes (3 for IFA and 3 for MMN) and an inscription "LNS" (for the LNS group) and numbered 1–1320 were placed in opaque envelopes and stacked in increasing order. At each enrollment, the study nurse shuffled the 9 topmost envelopes in the stack, and the woman picked one to reveal allocation."<br>**Comment:** adequately done |
| Blinding of participants and personnel (performance bias) | High risk | **Quote:** "Two individuals in Ghana who were independent of the research team color-coded the capsules by placing color stickers (which also included the letter P or L to indicate pregnancy or lactation) on the blister packs of IFA and MMN, so that no investigator, study worker, or participant knew the identities of the capsules except by the colors. Each fieldworker received from the field supervisor the coded allocations of only those women assigned to that fieldworker for follow-up home visits and was told not to reveal those codes to anyone. It was not possible to blind the fieldworkers and study participants to those consuming capsules vs. LNS (because of the starkly different characteristics)"<br>**Comment:** not done |
| Blinding of outcome assessment (detection bias) | Low risk | **Quote**: "…but none of the maternal or newborn anthropometrists was aware of the code allocations. Likewise, data analysts remained blinded until all preliminary analyses had been completed, and the allocation codes were broken."<br>**Comment:** adequately done |
| Incomplete outcome data (attrition bias) | Low risk | **Attrition:** Pregnancy outcomes: IFA group = 349/441; MMN group = 354/439; LNS group = 354/440; Birth anthropometric outcomes: IFA group = 305/441; MMN group = 318/439; LNS: 307/440<br>**Comment:** Reason for exclusion of participants provided, no attrition for pregnancy outcomes; reasons for attrition provided (additional details in Adu-Afarwuah *et al.* AJCN 2016) |
| Selective reporting (reporting bias) | Low risk | **Comment**: The trial was registered at clinicaltrials.gov (NCT00970866); SAP available online; outcomes described in the methods section reported in the results section |
| Other bias | Low risk | **Quote: "…**discovered that some mislabeling of IFA and MMN supplements had occurred due to miscommunication, and as a result, some women in the IFA and MMN groups had<br>received both IFA and MMN supplements during pregnancy. Throughout the study, we kept full records of when any batch of supplement arrived in Ghana, the date the |

| | | |
|---|---|---|
| | | distribution of each batch was started, and the batch number of each blister pack delivered to each woman at every home visit. Therefore, it was possible later to identify which subjects received both the IFA and MMN supplements during pregnancy…When we discovered the mislabeling (described above), 510 women had been enrolled. At the recommendation of the Data and Safety Monitoring Board and the funder, we calculated<br>the sample size by using the same parameters as before, but assuming 20% attrition based on data up to that point, and determined that if we enrolled an additional target number of 810 women (270/group), there would be sufficient statistical power to examine pregnancy outcomes even if we excluded women in the IFA and MMN groups who had the mixed "exposure" and those in the LNS groups who were pregnant during the same period".<br>**Comment**: adequately done |

**Funding**

Funded by a grant to the University of California, Davis, from the Bill & Melinda Gates Foundation.

| **Ashorn 2015 (34)** | | |
|---|---|---|
| **Bias** | **Authors' judgement** | **Support for judgement** |
| Random sequence generation (selection bias) | Low risk | **Quote:** "A study statistician not involved in data collection generated 4 randomization ode lists in blocks of 9 (one list for each of the 4 enrollment sites). In the randomization process, each participant number was allocated one of 9 possible letter codes (A, B, C, D, E, H, J, K, or M). Each letter code corresponded to one of the 3 interventions (i.e., each intervention matched with 3 separate letter codes). Another researcher not involved with the iLiNSDYAD trial then created individual randomization slips, each containing one unique identification number and the corresponding letter code.<br>**Comment**: adequately done. |
| Allocation concealment (selection bias) | Low risk | **Quote**: "The researcher sealed the slips into individual opaque randomization envelopes, marked each envelope with the trial name and an individual participant number, and sorted the envelopes in 4 stacks (one for each site), each ordered by the participant number shown on the envelope…For the actual enrollment and group allocation, a randomizer picked and shuffled the randomization envelopes for the 6 lowest participant numbers that had not yet been assigned to any participant. He or she then asked the potential participant to choose one, without showing her the envelope identifiers. The number on the envelope chosen by the woman became her participant number, and the contents of the envelope indicated her group allocation (in letter codes)".<br>**Comment:** adequately done |
| Blinding of participants and personnel (performance bias) | High risk | **Quote:** "The IFA and MMN interventions were provided by using double-masked procedures—that is, the capsules looked identical, and neither the participants nor the research team members were aware of the nutrient contents of the supplement capsules.<br>For the LNS group, we used single-masked procedures—that is, field workers who delivered the supplements knew which mothers were receiving LNS (but not a difference between IFA and MMN), and the participants were advised not to disclose information about their supplements to anyone other than an iLiNS team member."<br>**Comment:** not done |
| Blinding of outcome assessment (detection bias) | Low risk | **Quote:** "The data collectors who performed the anthropometric measurements or assessed other outcomes were not aware of group allocation. Researchers responsible for the data cleaning remained blind to the trial code until the database was fully cleaned.<br>**Comment**: adequately done |
| Incomplete outcome data (attrition bias) | Low risk | **Attrition:** Pregnancy outcomes: IFA group = 437/463; MMN group = 434/466; LNS group = 436/462; Birth weight: IFA group = 388/437; MMN group = 381/434; LNS: 394/436<br>**Quote:** "An analysis that included the twins and used the number of fetuses as a covariate also gave essentially similar results, and so did the Heckman's selection models that adjusted for the potential correlation between a tendency of missing data on outcome |

| | | |
|---|---|---|
| | | values and their actual values (details not shown)"; " Internal validity could have been compromised by a relatively large number of missing data, delay in anthropometric measurements of some participants, temporary discontinuation of the LNS distribution during the trial, and our inability to directly observe the consumption of the study supplements. Because the results were robust to several sensitivity analyses, we believe these factors did not significantly bias our conclusions. However, the smaller sample size than originally intended (due to budget reduction) limited the statistical power of the study. Therefore, although the results do not support the study hypothesis, they also do not rule out a modest intervention effect on birth size."<br>**Comment:** adequately done |
| Selective reporting (reporting bias) | Low risk | **Comment**: The trial was registered at clinicaltrials.gov (NCT01239693); SAP available online; outcomes described in the methods section reported in the results section |
| Other bias | Low risk | **Quote:** "During the trial implementation, international organizations involved in medium-quantity LNS distribution to children with acute malnutrition made a recommendation on a new quality assurance procedure for all such products. The recommendation involved the testing of LNS for the presence of *Cronobacter sakazakii* bacteria and in clinical practice withholding of the use of untested products or those that were found to contain any *C. sakazakii.* After consultation with members of the trial's data safety and monitoring board, the study team decided to withhold further distribution of LNS to the iLiNS-DYAD trial participants until the recommended testing had been completed. Because of this episode, a total of 160 pregnant women in the LNS group missed their study supplement for a period ranging from 1 to 20 d between 1 August 2012 and 21 August 2012. Of these women, 127 were provided with IFA capsules (1 capsule/d) while LNS was on hold; the other 33 were not located at their homes during the IFA distribution."<br>**Comment:** adequately done, disrupted exposure time < 3 weeks, any potential bias to the null hypothesis |
| **Funding**<br>Supported in part by a grant to the University of California, Davis, from the Bill & Melinda Gates Foundation, with additional funding from the Office of Health, Infectious Diseases, and Nutrition, Bureau for Global Health, US Agency for International Development (USAID) under terms of cooperative agreement AID-OAAA-1200005, through the Food and Nutrition Technical Assistance III Project (FANTA), managed by FHI 360. For data management and statistical analysis, the team received additional support from the Academy of Finland grant 252075 and the Medical Research Fund of Tampere University Hospital grant 9M004. YBC was supported by the Singapore Ministry of Health's National Medical Council under its Clinician Scientist Award. | | |

| **Mridha 2016 (32)** | | |
|---|---|---|
| **Bias** | **Authors' judgement** | **Support for judgement** |
| Random sequence generation (selection bias) | Low risk | **Quote:** "For the randomization, the study statistician at UC Davis first stratified the 64 clusters by subdistrict and union, and then assigned each cluster to 1 of 4 sets containing 16 clusters each. This procedure was then replicated several thousand times, and each randomization was tested for balance across groups with respect to mean cluster population, number of clinics and health workers per 1000 people, number of health-/nutrition-related nongovernmental organizations in the cluster, and the source of funding for the CHDP, as well as the SD of the cluster population size. The final randomization to the 4 arms was then chosen at random from the acceptable potential randomizations; and the letters A, B, C, and D were assigned to the 4 sets, randomly permuting them by sorting on a randomly generated, uniformly distributed number (with the use of SAS for Windows, release 9.2; SAS Institute) and assigning them respectively to control, child-only MNP, child-only LNS, and comprehensive LNS treatments".<br>**Comment:** adequately done |
| Allocation concealment (selection bias) | Low risk | **Quote:** " The final randomization to the 4 arms was then chosen at random from the acceptable potential randomizations; and the letters A, B, C, and D were assigned to the 4 sets, randomly permuting them by sorting on a randomly generated, uniformly distributed number (with the use of SAS for Windows, release 9.2; SAS Institute) and assigning them respectively to control, child-only MNP, child-only LNS, and comprehensive LNS treatments".<br>**Comment**: central randomization of a cluster-randomized trial |
| Blinding of participants and personnel (performance bias) | High risk | **Comment**: participant blinding not possible due to the nature of the intervention (LNS vs. IFA) |
| Blinding of outcome assessment (detection bias) | Low risk | **Quote:** "The trial was a researcher-blind, longitudinal, cluster randomized effectiveness trial"; " Data collection was performed by 2 separate teams: the "SDU visit team," which collected clinical and anthropometric data at the SDU, and the "home visit team," which enrolled mothers and collected baseline and follow-up data at participants' homes"… "To the extent possible, both study evaluation teams were kept blind to group assignment, although this was difficult for home visit team members because they might have seen supplements in the home"; "researchers responsible for the collection of outcome data were kept blind to study assignment."<br>**Comment:** adequately done |
| Incomplete outcome data (attrition bias) | Low risk | **Attrition**: LNS = 898/1047; IFA = 2551/2964<br>**Comment:** reasons given for loss to follow-up; missing outcome data balanced in numbers across intervention groups |
| Selective reporting (reporting bias) | Low risk | **Comment:** The trial was registered at ClinicalTrials.gov (NCT01715038); outcomes described in the methods section reported in the results section |

| | | |
|---|---|---|
| Other bias | Low risk | **Quote:** "LNS-PL distribution was interrupted from 8 August to 20 October 2012 to comply with a new quality-control criterion for ready-to-use supplementary foods implemented by the World Food Program, which required the absence of Cronobacter sakazakii (i.e., no samples testing positive at any amount) …During the interruption, women in all of the arms received IFA."; " A separate exploratory analysis was conducted to examine the effect of the intervention on children who were born before the interruption of LNS-PLs. To account for the interruption in LNS-PLs, we tested models that included the number of days the woman was participating in the study during the interruption period. However, we found that this did not improve the model fit compared with including the time interval variable and its interaction with intervention group in the model, and therefore we did not continue with this approach."<br>**Comment:** adequately done |

**Funding**
Supported by the Office of Health, Infectious Diseases, and Nutrition, Bureau for Global Health, US Agency for International Development (USAID) under the terms of cooperative agreement AID-OAA-A-12-00005, through the Food and Nutrition Technical Assistance III Project (FANTA), managed by FHI 360. Our research intervention was incorporated into the community heath and development program of LAMB, which was supported by Plan-Bangladesh in 6 of the 11 study unions. Nutriset S.A.S. prepared the lipid-based nutrient supplements for this trial, and Hudson Pharmaceuticals Ltd. prepared the iron and folic acid tablets.

| Hambidge 2019 (35) | | |
|---|---|---|
| **Bias** | **Authors' judgement** | **Support for judgement** |
| Random sequence generation (selection bias) | Low risk | **Quote:** **"**The DCC *(Data Coordinating Center at RTI International)* created the randomization scheme, centrally generating the allocation sequence for each site. To ensure geographic balance, a permuted block design stratified by GN clusters was used for assigning individual participants to a trial arm. The allocation ratio was 1:1:1 within blocks which randomly varied between sizes of 3, 6, or 9 for each site. Once the responsible home visitor research assistant identified an eligible participant, they received the random assignment generated by the site data manager from the centralized computerized data management system maintained by the DCC."<br>**Comment:** adequately done |
| Allocation concealment (selection bias) | Low risk | **Quote:** Once the responsible home visitor research assistant identified an eligible participant, they received the random assignment generated by the site data manager from the centralized computerized data management system maintained by the DCC."<br>**Comment**: adequately done |
| Blinding of participants and personnel (performance bias) | High risk | **Quote:** "This was an individually randomized, nonmasked, multisite controlled efficacy trial"<br>**Comment**: participant blinding not possible due to the nature of the intervention (LNS pre-conception vs. LNS at the end of the 1$^{st}$ trimester vs. standard of care) |
| Blinding of outcome assessment (detection bias) | Unclear risk | **Quote:** All anthropometry was performed by trained assessment teams who were not involved in the biweekly home visits.<br>**Comment:** unclear if the anthropometrists were blinded to intervention assignment |
| Incomplete outcome data (attrition bias) | Low risk | **Attrition**: data were not provided for Guatemala individually, but for all sites in this multi-site trial combined.<br>**Comment:** reasons given for loss to follow-up; missing outcome data balanced in numbers across intervention groups |
| Selective reporting (reporting bias) | Low risk | **Comment:** The trial was registered at ClinicalTrials.gov (NCT01883193); study protocol is available online; outcomes described in the methods section reported in the results section |
| Other bias | Low risk | **Comment:** no other potential sources of bias reported |
| **Funding**<br>Supported by Bill &Melinda Gates Foundation grant OPP1055867 and *Eunice Kennedy Shriver* National Institute of Child Health and Human Development and Office of Dietary Supplements, NIH grant U10 HD<br>076474. | | |

#### Supplemental Table 7A: Sensitivity analyses for main effects of SQ-LNS vs IFA/SOC on birth outcomes[1]

| Birth outcomes | MD or RR (95% CI)[2]<br>N (trials); p-value<br>Excluding gestational age not measured by ultrasound | MD or RR (95% CI)[2]<br>N (trials); p-value<br>Anthropometry within 72 h of birth | MD or RR (95% CI)[2]<br>N (trials); p-value<br>Excluding Guatemala | MD or RR (95% CI)[2]<br>N (trials); p-value<br>Including pre-conception |
|---|---|---|---|---|
| Birth weight (g) | N/A | 49.3 (26.2, 72.3)<br>4468 (4); p<0.001 | 47.6 (22.1, 73.0)<br>4834 (3); p<0.001 | 42.8 (20.2, 65.5)<br>5461 (4); p<0.001 |
| Weight-for-age z score (WAZ) | N/A | 0.12 (0.07, 0.18)<br>4468 (4); p<0.001 | 0.12 (0.05, 0.18)<br>4834 (3); p<0.001 | 0.10 (0.05, 0.16)<br>5461 (4); p<0.001 |
| Weight-for-gestational age z-score (WGAZ) | 0.13 (0.05, 0.21)<br>1717 (3); p=0.001 | 0.15 (0.07, 0.24)<br>1129 (3); p=0.001 | 0.09 (-0.02, 0.19)<br>1370 (2); p=0.099 | 0.11 (0.04, 0.19)<br>1871 (3); p=0.003 |
| Low birth weight (LBW) | N/A | 0.91 (0.82, 1.01)<br>4468 (4); p=0.075 | 0.90 (0.80, 1.00)<br>4834 (3); p=0.054 | 0.90 (0.82, 1.00)<br>5461 (4); p=0.048 |
| Birth weight < 2 kg | N/A | 0.75 (0.57, 1.00)<br>4035 (3); p=0.050 | 0.78 (0.60, 1.02)<br>4834 (3); p=0.069 | 0.81 (0.63, 1.05)<br>5461 (4); p=0.108 |
| Small-for-gestational age (SGA) | 0.86 (0.73, 1.01)<br>1717 (3); p=0.070 | 0.97 (0.92, 1.02)<br>4378 (4); p=0.243 | 0.97 (0.92, 1.02)<br>4834 (3); p=0.199 | 0.96 (0.92, 1.01)<br>5335 (4); p=0.133 |
| Large-for-gestational age (LGA) | 0.95 (0.53, 1.72)<br>1370 (2); p=0.878 | 1.64 (0.81, 3.32)<br>3786 (2); p=0.168 | 1.00 (0.61, 1.65)<br>4834 (3); p=0.984 | 1.00 (0.61, 1.65)<br>4834 (3); p=0.984 |
| Birth length (cm) | N/A | 0.19 (0.08, 0.31)<br>4217 (3); p=0.001 | 0.19 (0.07, 0.31)<br>4575 (3); p=0.002 | 0.15 (0.04, 0.25)<br>5202 (4); p=0.005 |
| Length-for-age z score (LAZ) | N/A | 0.11 (0.05, 0.17)<br>4217 (3); p<0.001 | 0.11 (0.04, 0.17)<br>4575 (3); p=0.001 | 0.09 (0.03, 0.14)<br>5202 (4); p=0.002 |
| Length-for-gestational age z-score (LGAZ) | 0.13 (0.05, 0.21)<br>1460 (3); p=0.002 | 0.14 (0.05, 0.23)<br>880 (2); p=0.002 | 0.10 (-0.03, 0.22)<br>1113 (2); p=0.123 | 0.09 (0.03, 0.15)<br>1614 (3); p=0.004 |
| Newborn stunting | N/A | 0.83 (0.75, 0.93)<br>4217 (3); p=0.001 | 0.84 (0.74, 0.95)<br>4575 (3); p=0.005 | 0.87 (0.78, 0.97)<br>5202 (4); p=0.014 |
| Low LGAZ | 0.95 (0.71, 1.26)<br>1460 (3); p=0.713 | 0.90 (0.79, 1.03)<br>4127 (3); p=0.128 | 0.90 (0.80, 1.02)<br>4575 (3); p=0.099 | 0.91 (0.81, 1.03)<br>5076 (4); p=0.123 |
| BMI-for-age z-score (BMIZ) | N/A | 0.11 (0.04, 0.17)<br>4217 (3); p=0.001 | 0.10 (0.03, 0.17)<br>4563 (3); p=0.004 | 0.09 (0.03, 0.16)<br>5190 (4); p=0.004 |

| | | | | |
|---|---|---|---|---|
| Low BMIZ | N/A | 0.89 (0.81, 0.98)<br>4217 (3); p=0.016 | 0.90 (0.81, 0.99)<br>4563 (3); p=0.029 | 0.90 (0.82, 0.99)<br>5190 (4); p=0.035 |
| Head circumference (cm) | N/A | 0.10 (0.03, 0.18)<br>4218 (3); p=0.005 | 0.12 (0.04, 0.20)<br>4577 (3); p=0.003 | 0.10 (0.03, 0.17)<br>5204 (4); p=0.006 |
| Head circumference-for-age z score (HCZ) | N/A | 0.10 (0.04, 0.15)<br>4218 (3); p=0.001 | 0.10 (0.04, 0.17)<br>4577 (3); p=0.002 | 0.09 (0.03, 0.14)<br>5204 (4); p=0.003 |
| Head circumference-for-gestational age z score (HCGAZ) | 0.11 (0.02, 0.20)<br>1461 (3); p=0.019 | 0.13 (0.03, 0.24)<br>880 (2); p=0.014 | 0.13 (0.00, 0.25)<br>1114 (2); p=0.048 | 0.09 (-0.01, 0.19)<br>1615 (3); p=0.067 |
| Low HCZ | N/A | 0.87 (0.76, 0.99)<br>4218 (3); p=0.030 | 0.84 (0.75, 0.96)<br>4577 (3); p=0.008 | 0.86 (0.76, 0.97)<br>5204 (4); p=0.013 |
| Low HCGAZ | 0.74 (0.37, 1.51)<br>1461 (3); p=0.414 | 0.88 (0.76, 1.03)<br>4128 (3); p=0.105 | 0.88 (0.76, 1.02)<br>4577 (3); p=0.081 | 0.88 (0.77, 1.02)<br>5078 (4); p=0.092 |
| Mid-upper arm circumference (MUAC) (cm) | N/A | 0.08 (0.01, 0.14)<br>3786 (2); p=0.027 | 0.08 (0.02, 0.14)<br>4581 (3); p=0.009 | 0.08 (0.02, 0.14)<br>4581 (3); p=0.009 |
| Duration of gestation (wk) | 0.15 (-0.03, 0.33)<br>1880 (3); p=0.107 | N/A | 0.14 (0.01, 0.27)<br>4981 (3); p=0.031 | 0.10 (-0.02, 0.22)<br>5509 (4); p=0.105 |
| Preterm birth | 0.90 (0.68, 1.18)<br>1880 (3); p=0.446 | N/A | 0.94 (0.80, 1.10)<br>4981 (3); p=0.432 | 0.96 (0.82, 1.12)<br>5509 (4); p=0.612 |

[1]BMIZ, BMI-for-age z-score; HCGAZ, head circumference-for-gestational age z-score; HCZ, head circumference-for-age z-score; IFA, iron and folic acid supplement; LAZ, length-for-age z-score; LBW, low birth weight; LGA, large-for-gestational age; LGAZ, length-for-gestational age z-score; MD, mean difference; MUAC, mid-upper arum circumference; RR, relative risk; SGA, small-for-gestational age; SOC, standard of care; SQ-LNS, small-quantity lipid-based nutrient supplement; WAZ, weight-for-age z-score; WGAZ, weight-for-gestational age z-score

[2]For continuous outcomes, values are MDs: LNS – IFA/SOC (95% CIs). For binary outcomes, values are RRs: LNS compared with IFA/SOC (95% CIs).

#### Supplemental Table 7B: Sensitivity analyses for main effects of SQ-LNS vs IFA/SOC on infant anthropometric outcomes at 6 mo of age[1]

| Infant anthropometric outcomes at 6 mo | MD or PR (95% CI)[2] N (trials); p-value Excluding Women First and DYAD-M Simple Follow-up | MD or PR (95% CI)[2] N (trials); p-value Excluding Guatemala | MD or PR (95% CI)[2] N (trials); p-value Including pre-conception |
|---|---|---|---|
| Weight-for-age z-score (WAZ) | 0.02 (-0.06, 0.11)<br>4322 (3); p=0.589 | 0.02 (-0.06, 0.11)<br>4596 (3); p=0.602 | 0.05 (-0.02, 0.12)<br>5210 (4); p=0.176 |
| Underweight | 0.96 (0.80, 1.17)<br>4322 (3); p=0.714 | 0.96 (0.80, 1.16)<br>4596 (3); p=0.684 | 0.90 (0.78, 1.03)<br>5210 (4); p=0.123 |
| Length-for-age z-score (LAZ) | 0.04 (-0.03, 0.12)<br>4325 (3); p=0.267 | 0.04 (-0.04, 0.11)<br>4599 (3); p=0.311 | 0.06 (0.00, 0.13)<br>5213 (4); p=0.063 |
| Stunted | 0.95 (0.82, 1.10)<br>4325 (3); p=0.480 | 0.93 (0.81, 1.07)<br>4599 (3); p=0.325 | 0.88 (0.79, 0.99)<br>5213 (4); p=0.037 |
| Weight-for-length z-score (WLZ) | -0.01 (-0.10, 0.07)<br>4321 (3); p=0.769 | -0.01 (-0.09, 0.07)<br>4595 (3); p=0.824 | 0.00 (-0.07, 0.07)<br>5209 (4); p=0.960 |
| Wasted | 0.90 (0.65, 1.24)<br>4321 (3); p=0.507 | 0.95 (0.69, 1.31)<br>4595 (3); p=0.749 | 0.94 (0.69, 1.27)<br>5209 (4); p=0.675 |
| Head circumference-for-age z-score (HCZ) | 0.01 (-0.06, 0.07)<br>4324 (3); p=0.838 | 0.01 (-0.06, 0.07)<br>4598 (3); p=0.836 | 0.00 (-0.05, 0.06)<br>5209 (4); p=0.886 |
| Low HCZ | 0.91 (0.80, 1.04)<br>4324 (3); p=0.154 | 0.91 (0.81, 1.03)<br>4598 (3); p=0.149 | 0.93 (0.83, 1.05)<br>5209 (4); p=0.232 |
| MUAC-for-age z-score (MUACZ) | -0.01 (-0.09, 0.08)<br>4326 (3); p=0.896 | 0.00 (-0.07, 0.08)<br>4600 (3); p=0.949 | 0.00 (-0.07, 0.08)<br>4600 (3); p=0.949 |
| Low MUAC [MUACZ < -2 SD or MUAC < 125 mm] | 1.01 (0.78, 1.32)<br>4326 (3); p=0.930 | 1.00 (0.78, 1.29)<br>4600 (3); p=0.987 | 1.00 (0.78, 1.29)<br>4600 (3); p=0.987 |
| Acute malnutrition [WLZ < -2 SD or MUAC < 125 mm] | 0.99 (0.78, 1.25)<br>4321 (3); p=0.918 | 1.00 (0.79, 1.25)<br>4595 (3); p=0.970 | 1.00 (0.79, 1.25)<br>4595 (3); p=0.970 |

[1]HCZ, head circumference-for-age z-score; IFA, iron and folic acid supplement; LAZ, length-for-age z-score; MD, mean difference; MUAC, mid-upper arum circumference; MUACZ, mid-upper arm circumference-for-age z-score; PR, prevalence ratio; SOC, standard of care; SQ-LNS, small-quantity lipid-based nutrient supplement; WAZ, weight-for-age z-score; WLZ, weight-for-length z-score

[2]For continuous outcomes, values are MDs: LNS – IFA/SOC (95% CIs). For binary outcomes, values are PRs: LNS compared with IFA/SOC (95% CIs).

#### Supplemental Table 7C: Sensitivity analyses for main effects of SQ-LNS vs IFA/SOC on adverse outcomes[1]

| Adverse outcomes | RR (95% CI) N (trials); p-value Excluding Women First and DYAD-M Simple Follow-up | RR (95% CI) N (trials); p-value Excluding Guatemala | RR (95% CI) N (trials); p-value Including pre-conception |
|---|---|---|---|
| Cesarian-Section | 1.11 (0.90, 1.38) 4916 (3); p=0.339 | 1.12 (0.91, 1.38) 5228 (3); p=0.281 | 1.03 (0.90, 1.19) 5891 (4); p=0.642 |
| Miscarriage | 0.88 (0.69, 1.13) 5240 (3); p=0.319 | 0.89 (0.69, 1.13) 5565 (3); p=0.337 | 0.86 (0.70, 1.06) 6327 (4); p=0.150 |
| Stillbirth | 1.16 (0.78, 1.72) 5240 (3); p=0.457 | 1.24 (0.85, 1.81) 5565 (3); p=0.270 | 1.32 (0.91, 1.92) 6327 (4); p=0.146 |
| Miscarriage or stillbirth | 0.97 (0.77, 1.21) 5240 (3); p=0.785 | 1.00 (0.80, 1.25) 5565 (3); p=0.999 | 0.98 (0.82, 1.17) 6327 (4); p=0.793 |
| Early neonatal mortality | 0.78 (0.47, 1.32) 4746 (3); p=0.356 | 0.76 (0.47, 1.24) 5053 (3); p=0.277 | 0.76 (0.48, 1.20) 5698 (4); p=0.233 |
| Miscarriage or stillbirth or early neonatal mortality | 0.92 (0.74, 1.14) 5240 (3); p=0.445 | 0.94 (0.76, 1.16) 5565 (3); p=0.569 | 0.94 (0.78, 1.11) 6327 (4); p=0.454 |
| Neonatal mortality | 0.90 (0.57, 1.40) 4766 (3); p=0.635 | 0.84 (0.55, 1.28) 5076 (3); p=0.414 | 0.88 (0.60, 1.30) 5720 (4); p=0.521 |
| Mortality 0-6 mo | 0.99 (0.70, 1.40) 4809 (3); p=0.955 | 0.97 (0.69, 1.35) 5121 (3); p=0.843 | 0.98 (0.72, 1.33) 5754 (4); p=0.893 |

[1]GRADE, Grading of Recommendations Assessment, Development and Evaluation; IFA, iron and folic acid supplement; RR, relative risk; SOC, standard of care; SQ-LNS, small-quantity lipid-based nutrient supplement

[2]Values are RRs: LNS compared with IFA/SOC (95% CIs).

**Online supplmental material**

Effects of prenatal small-quantity lipid-based nutrient supplements on pregnancy, birth and infant outcomes: a systematic review and meta-analysis of individual participant data from randomized controlled trials in low-income and middle-income countries

Dewey *et al.* (2024)

**Supplemental Table 8: Sensitivity analyses for main effects of SQ-LNS vs IFA/SOC on birth outcomes among those with ultrasound data and among those with anthropometry within 72 h of birth1**

| | **Sex** | | | |
|---|---|---|---|---|
| | **N (trials)[2]** | **P-Int[3]** | **Male** | **Female** |
| **Gestational age measured by ultrasound** | | | | |
| Small-for-gestational age (SGA)[4] | 1717 (3) | 0.512 | 0.88 (0.71, 1.10) | 0.79 (0.62, 1.02) |
| Low LGAZ[5] | 1460 (3) | 0.518 | 0.88 (0.59, 1.31) | 1.08 (0.72, 1.61) |
| Duration of gestation (wk) | 1847 (3) | 0.047 | 0.01 (-0.23, 0.25) | 0.30 (0.09, 0.51) |
| Preterm birth[6] | 1847 (3) | 0.915 | 0.91 (0.59, 1.41) | 0.89 (0.59, 1.35) |
| **Anthropometry within 72 h of birth** | | | | |
| Birth weight (g) | 4468 (4) | 0.005 | 22.3 (-14.3, 58.9) | 86.0 (58.3, 113.6) |
| Weight-for-age z score (WAZ)[7] | 4468 (4) | 0.004 | 0.05 (-0.03, 0.14) | 0.21 (0.14, 0.28) |
| Weight-for-gestational age z-score (WGAZ) | 1129 (3) | 0.296 | 0.19 (0.07, 0.31) | 0.11 (0.00, 0.22) |
| Low birth weight[8] | 4468 (4) | 0.007 | 1.04 (0.89, 1.22) | 0.80 (0.71, 0.90) |
| Small-for-gestational age (SGA) | 4378 (4) | 0.327 | 0.99 (0.91, 1.07) | 0.94 (0.87, 1.01) |
| Birth length (cm) | 4217 (3) | 0.035 | 0.06 (-0.13, 0.25) | 0.35 (0.20, 0.50) |
| Length-for-age z score (LAZ) | 4217 (3) | 0.036 | 0.04 (-0.06, 0.14) | 0.19 (0.11, 0.27) |
| Length-for-gestational age z-score (LGAZ) | 880 (2) | 0.512 | 0.16 (0.01, 0.32) | 0.09 (-0.04, 0.22) |
| Newborn stunting | 4217 (3) | 0.173 | 0.91 (0.76, 1.10) | 0.74 (0.63, 0.87) |
| Low LGAZ | 4127 (3) | 0.919 | 0.93 (0.76, 1.13) | 0.92 (0.73, 1.16) |
| BMI-for-age z-score (BMIZ) | 4217 (3) | 0.024 | 0.03 (-0.07, 0.13) | 0.19 (0.10, 0.27) |
| Low BMIZ | 4217 (3) | 0.011 | 1.02 (0.89, 1.17) | 0.76 (0.65, 0.90) |
| Head circumference (cm) | 4218 (3) | 0.051 | 0.04 (-0.08, 0.16) | 0.19 (0.09, 0.29) |
| Head circumference-for-age z score (HCZ) | 4218 (3) | 0.036 | 0.03 (-0.06, 0.13) | 0.17 (0.08, 0.25) |
| Head circumference-for-gestational age z score (HCGAZ) | 880 (2) | 0.392 | 0.08 (-0.05, 0.22) | 0.17 (0.02, 0.32) |
| Low HCZ | 4218 (3) | 0.784 | 0.88 (0.69, 1.12) | 0.84 (0.73, 0.98) |
| Mid-upper arm circumference (MUAC) (cm) | 3786 (2) | 0.071 | 0.02 (-0.08, 0.12) | 0.13 (0.05, 0.21) |

| | Maternal BMI N (trials)[2] | P-Int | ≥ 20 kg/m² | < 20 kg/m² |
|---|---|---|---|---|
| **Gestational age measured by ultrasound** | | | | |
| Small-for-gestational age (SGA)[4] | 1700 (3) | 0.722 | 0.85 (0.68, 1.07) | 0.88 (0.67, 1.16) |
| Low LGAZ[5] | 1444 (3) | 0.861 | 0.99 (0.67, 1.47) | 0.94 (0.60, 1.48) |
| Duration of gestation (wk) | 1863 (3) | 0.596 | 0.15 (-0.06, 0.35) | 0.06 (-0.33, 0.46) |
| Preterm birth[6] | 1497 (2) | 0.761 | 0.88 (0.61, 1.28) | 0.93 (0.55, 1.57) |
| **Anthropometry within 72 h of birth** | | | | |
| Birth weight (g) | 4368 (4) | 0.614 | 53.4 (24.6, 82.3) | 49.5 (17.4, 81.6) |
| Weight-for-age z score (WAZ)[7] | 4368 (4) | 0.712 | 0.13 (0.06, 0.21) | 0.12 (0.04, 0.20) |
| Weight-for-gestational age z-score (WGAZ) | 1116 (3) | 0.238 | 0.19 (0.09, 0.28) | -0.01 (-0.23, 0.21) |
| Low birth weight[8] | 4368 (4) | 0.615 | 0.91 (0.77, 1.06) | 0.90 (0.80, 1.01) |
| Small-for-gestational age (SGA) | 4278 (4) | 0.828 | 0.95 (0.87, 1.03) | 0.97 (0.90, 1.05) |
| Birth length (cm) | 4118 (3) | 0.373 | 0.19 (0.04, 0.34) | 0.24 (0.06, 0.42) |
| Length-for-age z score (LAZ) | 4118 (3) | 0.523 | 0.12 (0.03, 0.20) | 0.12 (0.03, 0.22) |
| Length-for-gestational age z-score (LGAZ) | 868 (2) | 0.516 | 0.15 (0.06, 0.25) | 0.02 (-0.29, 0.34) |
| Newborn stunting | 4118 (3) | 0.346 | 0.89 (0.74, 1.08) | 0.80 (0.70, 0.92) |
| Low LGAZ | 4028 (3) | 0.590 | 0.96 (0.78, 1.18) | 0.86 (0.71, 1.05) |
| BMI-for-age z-score (BMIZ) | 4118 (3) | 0.672 | 0.11 (0.03, 0.19) | 0.12 (0.02, 0.21) |
| Low BMIZ | 4118 (3) | 0.894 | 0.84 (0.71, 1.01) | 0.90 (0.79, 1.02) |
| Head circumference (cm) | 4119 (3) | 0.147 | 0.08 (-0.01, 0.17) | 0.16 (0.05, 0.28) |
| Head circumference-for-age z score (HCZ) | 4119 (3) | 0.210 | 0.09 (0.01, 0.16) | 0.13 (0.04, 0.23) |
| Head circumference-for-gestational age z score (HCGAZ) | 868 (2) | 0.615 | 0.14 (0.03, 0.25) | 0.10 (-0.23, 0.44) |
| Low HCZ | 4119 (3) | 0.094 | 0.97 (0.79, 1.18) | 0.79 (0.67, 0.93) |
| Mid-upper arm circumference (MUAC) (cm) | 3688 (2) | 0.518 | 0.06 (-0.02, 0.14) | 0.07 (-0.02, 0.16) |

| | Maternal age | | | |
|---|---|---|---|---|
| | N (trials) | P-Int | ≥ 25 y | < 25 y |
| **Gestational age measured by ultrasound** | | | | |
| Small-for-gestational age (SGA)[4] | 1717 (3) | 0.716 | 0.85 (0.65, 1.10) | 0.88 (0.70, 1.10) |
| Low LGAZ[5] | 1460 (3) | 0.410 | 0.85 (0.53, 1.34) | 1.05 (0.73, 1.50) |
| Duration of gestation (wk) | 1880 (3) | 0.015 | -0.04 (-0.28, 0.19) | 0.32 (0.07, 0.58) |
| Preterm birth[6] | 1880 (3) | 0.975 | 0.93 (0.63, 1.37) | 0.90 (0.61, 1.32) |
| **Anthropometry within 72 h of birth** | | | | |
| Birth weight (g) | 4468 (4) | 0.114 | 80.9 (44.0, 117.9) | 36.7 (5.8, 67.5) |
| Weight-for-age z score (WAZ)[7] | 4468 (4) | 0.115 | 0.19 (0.11, 0.28) | 0.09 (0.02, 0.17) |
| Weight-for-gestational age z-score (WGAZ) | 1129 (3) | 0.220 | 0.26 (0.14, 0.39) | 0.09 (-0.05, 0.23) |
| Low birth weight[8] | 4468 (4) | 0.327 | 0.85 (0.69, 1.04) | 0.94 (0.84, 1.05) |
| Small-for-gestational age (SGA) | 4378 (4) | 0.026 | 0.85 (0.76, 0.95) | 1.01 (0.95, 1.09) |
| Birth length (cm) | 4217 (3) | 0.313 | 0.30 (0.10, 0.50) | 0.13 (-0.03, 0.28) |
| Length-for-age z score (LAZ) | 4217 (3) | 0.354 | 0.16 (0.05, 0.27) | 0.08 (-0.01, 0.16) |
| Length-for-gestational age z-score (LGAZ) | 880 (2) | 0.102 | 0.23 (0.06, 0.40) | 0.00 (-0.16, 0.17) |
| Newborn stunting | 4217 (3) | 0.329 | 0.93 (0.71, 1.21) | 0.80 (0.70, 0.93) |
| Low LGAZ | 4127 (3) | 0.722 | 0.97 (0.76, 1.24) | 0.89 (0.75, 1.06) |
| BMI-for-age z-score (BMIZ) | 4217 (3) | 0.191 | 0.17 (0.07, 0.27) | 0.08 (0.00, 0.16) |
| Low BMIZ | 4217 (3) | 0.380 | 0.81 (0.64, 1.02) | 0.93 (0.84, 1.04) |
| Head circumference (cm) | 4218 (3) | 0.283 | 0.20 (0.07, 0.34) | 0.05 (-0.05, 0.14) |
| Head circumference-for-age z score (HCZ) | 4218 (3) | 0.272 | 0.17 (0.07, 0.27) | 0.05 (-0.03, 0.13) |
| Head circumference-for-gestational age z score (HCGAZ) | 880 (2) | 0.903 | 0.19 (0.02, 0.37) | 0.11 (-0.06, 0.28) |
| Low HCZ | 4218 (3) | 0.628 | 0.81 (0.64, 1.01) | 0.89 (0.74, 1.08) |
| Mid-upper arm circumference (MUAC) (cm) | 3786 (2) | 0.452 | 0.11 (0.01, 0.20) | 0.06 (-0.03, 0.14) |

| | Maternal inflammation | | | |
|---|---|---|---|---|
| | N (trials) | P-Int | Not Inflamed | Inflamed |
| **Gestational age measured by ultrasound** | | | | |
| Small-for-gestational age (SGA)[4] | 1342 (2) | 0.488 | 0.93 (0.71, 1.22) | 0.83 (0.62, 1.10) |
| Low LGAZ[5] | 1092 (2) | 0.548 | 0.91 (0.56, 1.48) | 1.04 (0.65, 1.66) |
| Duration of gestation (wk) | 1481 (2) | 0.252 | 0.12 (-0.17, 0.40) | 0.39 (0.03, 0.75) |
| Preterm birth[6] | 1481 (2) | 0.374 | 0.96 (0.62, 1.47) | 0.71 (0.45, 1.13) |
| **Anthropometry within 72 h of birth** | | | | |
| Birth weight (g) | 1689 (3) | 0.047 | 33.5 (-9.6, 76.6) | 118.4 (43.7, 193.0) |
| Weight-for-age z score (WAZ)[7] | 1689 (3) | 0.046 | 0.08 (-0.02, 0.18) | 0.27 (0.10, 0.45) |
| Weight-for-gestational age z-score (WGAZ) | 767 (2) | 0.076 | 0.02 (-0.15, 0.19) | 0.24 (0.03, 0.45) |
| Low birth weight[8] | 1689 (3) | 0.086 | 0.91 (0.77, 1.06) | 0.64 (0.44, 0.93) |
| Small-for-gestational age (SGA) | 1689 (3) | 0.975 | 0.95 (0.84, 1.07) | 0.93 (0.73, 1.18) |
| Birth length (cm) | 1445 (2) | 0.120 | 0.05 (-0.19, 0.28) | 0.49 (0.06, 0.91) |
| Length-for-age z score (LAZ) | 1445 (2) | 0.127 | 0.02 (-0.10, 0.14) | 0.24 (0.02, 0.46) |
| Length-for-gestational age z-score (LGAZ) | - | - | - | - |
| Newborn stunting | 1445 (2) | 0.124 | 0.92 (0.73, 1.15) | 0.64 (0.40, 1.00) |
| Low LGAZ | 1445 (2) | 0.558 | 0.90 (0.69, 1.18) | 0.80 (0.42, 1.54) |
| BMI-for-age z-score (BMIZ) | 1445 (2) | 0.052 | 0.11 (0.00, 0.23) | 0.36 (0.14, 0.59) |
| Low BMIZ | 1445 (2) | 0.084 | 0.82 (0.69, 0.97) | 0.52 (0.34, 0.81) |
| Head circumference (cm) | 1444 (2) | 0.719 | 0.14 (-0.02, 0.30) | 0.32 (0.02, 0.61) |
| Head circumference-for-age z score (HCZ) | 1444 (2) | 0.547 | 0.11 (-0.02, 0.23) | 0.26 (0.02, 0.49) |
| Head circumference-for-gestational age z score (HCGAZ) | - | - | - | - |
| Low HCZ | 1444 (2) | 0.143 | 0.90 (0.73, 1.10) | 0.63 (0.41, 0.95) |
| Mid-upper arm circumference (MUAC) (cm) | 1445 (2) | 0.123 | 0.06 (-0.04, 0.16) | 0.24 (0.05, 0.43) |

| | Food insecurity | | | |
|---|---|---|---|---|
| | N (trials) | P-Int | None/mild | Moderate/severe |
| **Gestational age measured by ultrasound** | | | | |
| Small-for-gestational age (SGA)[4] | 1360 (2) | 0.705 | 0.90 (0.68, 1.19) | 0.89 (0.68, 1.18) |
| Low LGAZ[5] | 1106 (2) | 0.948 | 1.03 (0.62, 1.71) | 0.99 (0.64, 1.55) |
| Duration of gestation (wk) | 1499 (2) | 0.804 | 0.24 (-0.05, 0.52) | 0.17 (-0.19, 0.53) |
| Preterm birth[6] | 1499 (2) | 0.999 | 0.90 (0.57, 1.41) | 0.87 (0.57, 1.32) |
| **Anthropometry within 72 h of birth** | | | | |
| Birth weight (g) | 4030 (3) | 0.128 | 31.0 (-0.9, 63.0) | 65.3 (24.3, 106.4) |
| Weight-for-age z score (WAZ)[7] | 4030 (3) | 0.136 | 0.08 (0.00, 0.16) | 0.16 (0.06, 0.26) |
| Weight-for-gestational age z-score (WGAZ) | 781 (2) | 0.496 | 0.18 (0.00, 0.36) | 0.05 (-0.15, 0.25) |
| Low birth weight[8] | 4030 (3) | 0.413 | 0.96 (0.84, 1.10) | 0.88 (0.74, 1.05) |
| Small-for-gestational age (SGA) | 4030 (3) | 0.806 | 0.98 (0.92, 1.05) | 0.96 (0.87, 1.07) |
| Birth length (cm) | 3782 (2) | 0.027 | 0.09 (-0.06, 0.23) | 0.30 (0.11, 0.49) |
| Length-for-age z score (LAZ) | 3782 (2) | 0.049 | 0.05 (-0.03, 0.13) | 0.16 (0.06, 0.26) |
| Length-for-gestational age z-score (LGAZ) | - | - | - | - |
| Newborn stunting | 3782 (2) | 0.041 | 0.95 (0.82, 1.10) | 0.72 (0.59, 0.89) |
| Low LGAZ | 3782 (2) | 0.431 | 0.87 (0.72, 1.05) | 0.97 (0.78, 1.21) |
| BMI-for-age z-score (BMIZ) | 3782 (2) | 0.484 | 0.09 (0.00, 0.17) | 0.13 (0.01, 0.26) |
| Low BMIZ | 3782 (2) | 0.676 | 0.91 (0.80, 1.04) | 0.89 (0.77, 1.01) |
| Head circumference (cm) | 3783 (2) | 0.057 | 0.03 (-0.06, 0.13) | 0.20 (0.06, 0.35) |
| Head circumference-for-age z score (HCZ) | 3783 (2) | 0.131 | 0.04 (-0.04, 0.12) | 0.16 (0.04, 0.28) |
| Head circumference-for-gestational age z score (HCGAZ) | - | - | - | - |
| Low HCZ | 3783 (2) | 0.999 | 0.87 (0.72, 1.06) | 0.87 (0.71, 1.05) |
| Mid-upper arm circumference (MUAC) (cm) | 3784 (2) | 0.330 | 0.06 (-0.03, 0.14) | 0.09 (0.00, 0.18) |

| | Compliance | | | |
|---|---|---|---|---|
| | N (trials) | P-Int | ≥ 4 d/wk | < 4 d/wk |
| **Gestational age measured by ultrasound** | | | | |
| Small-for-gestational age (SGA)[4] | 1364 (2) | 0.926 | 0.89 (0.72, 1.09) | 0.86 (0.45, 1.65) |
| Low LGAZ[5] | 1112 (2) | 0.988 | 0.97 (0.68, 1.38) | 0.99 (0.30, 3.25) |
| Duration of gestation (wk) | 1505 (2) | 0.909 | 0.24 (-0.01, 0.48) | 0.16 (-0.46, 0.78) |
| Preterm birth[6] | 1505 (2) | 0.426 | 0.83 (0.59, 1.15) | 1.20 (0.53, 2.73) |
| **Anthropometry within 72 h of birth** | | | | |
| Birth weight (g) | 4004 (3) | 0.270 | 65.0 (32.5, 97.4) | 20.1 (-56.8, 96.9) |
| Weight-for-age z score (WAZ)[7] | 4004 (3) | 0.245 | 0.16 (0.08, 0.24) | 0.05 (-0.14, 0.23) |
| Weight-for-gestational age z-score (WGAZ) | 781 (2) | 0.402 | 0.11 (-0.04, 0.25) | 0.30 (-0.08, 0.69) |
| Low birth weight[8] | 3760 (2) | 0.602 | 0.85 (0.74, 0.99) | 0.93 (0.72, 1.19) |
| Small-for-gestational age (SGA) | 4004 (3) | 0.453 | 0.94 (0.86, 1.01) | 1.01 (0.86, 1.20) |
| Birth length (cm) | 3758 (2) | 0.196 | 0.26 (0.10, 0.42) | -0.01 (-0.38, 0.36) |
| Length-for-age z score (LAZ) | 3758 (2) | 0.151 | 0.14 (0.06, 0.23) | -0.02 (-0.21, 0.18) |
| Length-for-gestational age z-score (LGAZ) | - | - | - | - |
| Newborn stunting | 3758 (2) | 0.072 | 0.76 (0.64, 0.90) | 1.08 (0.78, 1.50) |
| Low LGAZ | 3758 (2) | 0.596 | 0.82 (0.68, 0.98) | 0.91 (0.63, 1.31) |
| BMI-for-age z-score (BMIZ) | 3758 (2) | 0.778 | 0.14 (0.05, 0.23) | 0.10 (-0.09, 0.29) |
| Low BMIZ | 3758 (2) | 0.449 | 0.83 (0.74, 0.94) | 0.92 (0.72, 1.17) |
| Head circumference (cm) | 3759 (2) | 0.857 | 0.17 (0.05, 0.29) | 0.09 (-0.16, 0.35) |
| Head circumference-for-age z score (HCZ) | 3759 (2) | 0.701 | 0.14 (0.05, 0.24) | 0.07 (-0.12, 0.26) |
| Head circumference-for-gestational age z score (HCGAZ) | - | - | - | - |
| Low HCZ | 3759 (2) | 0.869 | 0.83 (0.69, 1.00) | 0.85 (0.65, 1.13) |
| Mid-upper arm circumference (MUAC) (cm) | 3760 (2) | 0.754 | 0.11 (0.02, 0.19) | 0.07 (-0.08, 0.22) |

| | Maternal malaria | | | |
|---|---|---|---|---|
| | N (trials) | P-Int | No malaria | Malaria |
| **Gestational age measured by ultrasound** | | | | |
| Small-for-gestational age (SGA)[4] | 1368 (2) | 0.016 | 0.99 (0.79, 1.24) | 0.60 (0.40, 0.89) |
| Low LGAZ[5] | 1112 (2) | 0.769 | 0.96 (0.64, 1.44) | 0.99 (0.55, 1.77) |
| Duration of gestation (wk) | 1511 (2) | 0.982 | 0.21 (-0.03, 0.45) | 0.15 (-0.47, 0.78) |
| Preterm birth[6] | 1511 (2) | 0.218 | 0.97 (0.69, 1.36) | 0.58 (0.28, 1.19) |
| **Anthropometry within 72 h of birth** | | | | |
| Birth weight (g) | 797 (2) | 0.283 | 69.8 (4.6, 135.1) | 148.0 (17.6, 278.3) |
| Weight-for-age z score (WAZ)[7] | 797 (2) | 0.295 | 0.16 (0.01, 0.30) | 0.33 (0.02, 0.64) |
| Weight-for-gestational age z-score (WGAZ) | 786 (2) | 0.122 | 0.09 (-0.05, 0.24) | 0.37 (0.06, 0.67) |
| Low birth weight[8] | 797 (2) | 0.287 | 0.84 (0.55, 1.28) | 0.56 (0.23, 1.35) |
| Small-for-gestational age (SGA) | 797 (2) | 0.034 | 0.94 (0.71, 1.26) | 0.50 (0.28, 0.88) |
| Birth length (cm) | - | - | - | - |
| Length-for-age z score (LAZ) | - | - | - | - |
| Length-for-gestational age z-score (LGAZ) | - | - | - | - |
| Newborn stunting | - | - | - | - |
| Low LGAZ | - | - | - | - |
| BMI-for-age z-score (BMIZ) | - | - | - | - |
| Low BMIZ | - | - | - | - |
| Head circumference (cm) | - | - | - | - |
| Head circumference-for-age z score (HCZ) | - | - | - | - |
| Head circumference-for-gestational age z score (HCGAZ) | - | - | - | - |
| Low HCZ | - | - | - | - |
| Mid-upper arm circumference (MUAC) (cm) | - | - | - | - |

BMIZ, BMI-for-age z-score; HCGAZ, head circumference-for-gestational age z-score; HCZ, head circumference-for-age z-score; IFA, iron and folic acid supplements; LGAZ, length-for-gestational age z-score; MUAC, mid-upper arm circumference; SGA, small-for-gestational age; SOC, standard of care; SQ-LNS, small-quantity lipid-based nutrient supplement; WAZ, weight-for-age z-score; WGAZ, weight-for-gestational age z-socre

[1]Values within indicated strata are pooled MD (95% CI) for continuous outcomes and pooled RR (95%CI) for dichotomous outcomes.

[2]N (trials) is the number of participants included in analysis with the number of trials included within the parentheses

[3]P-int is the P-for-interaction from the pooled interaction term between intervention and indicated effect modifier

[4]Small-for-gestational age defined as < 10th percentile weight-for-gestational age (INTERGROWTH-21st Standards) (24)

[5]WGAZ, LGAZ, HCGAZ are based on INTERGROWTH-21st Standards (24). Low length-for-gestational age z-score defined as LGAZ < -2 SD (INTERGROWTH-21st Standards)(24). Low head circumference-for-gestational age z-score defined as HCGAZ < -2 SD (INTERGROWTH-21st Standards)(24)

[6]Preterm birth defined as birth at < 37 weeks gestation (59)

[7]WAZ, LAZ, BMIZ, HCAZ are based on WHO Growth Standards (22,23). BMIZ is used as a proxy for weight-for-length z-score because the latter is not calculated for children with lengths < 45 cm (22). Newborn stunting defined as LAZ < -2 SD (WHO Growth Standards) (22). Low BMI z-score defined as BMIZ < - 2SD (WHO growth standards) (22). Low head circumference-for-age z-score defined as HCZ < -2 SD (WHO growth standards) (23)

[8]Low birth weight defined as weight < 2.5 kg at birth (59)

1

**Online Supplemental Material**

Effects of prenatal small-quantity lipid-based nutrient supplements on pregnancy, birth and infant outcomes: a systematic review and meta-analysis of individual participant data from randomized controlled trials in low- and middle-income countries
Dewey *et al.* (2024)

**Supplemental Figure 1. Summary risk of bias as a percentage of all included studies for the effects of prenatal SQ-LNS on pregnancy, birth and infant outcomes**

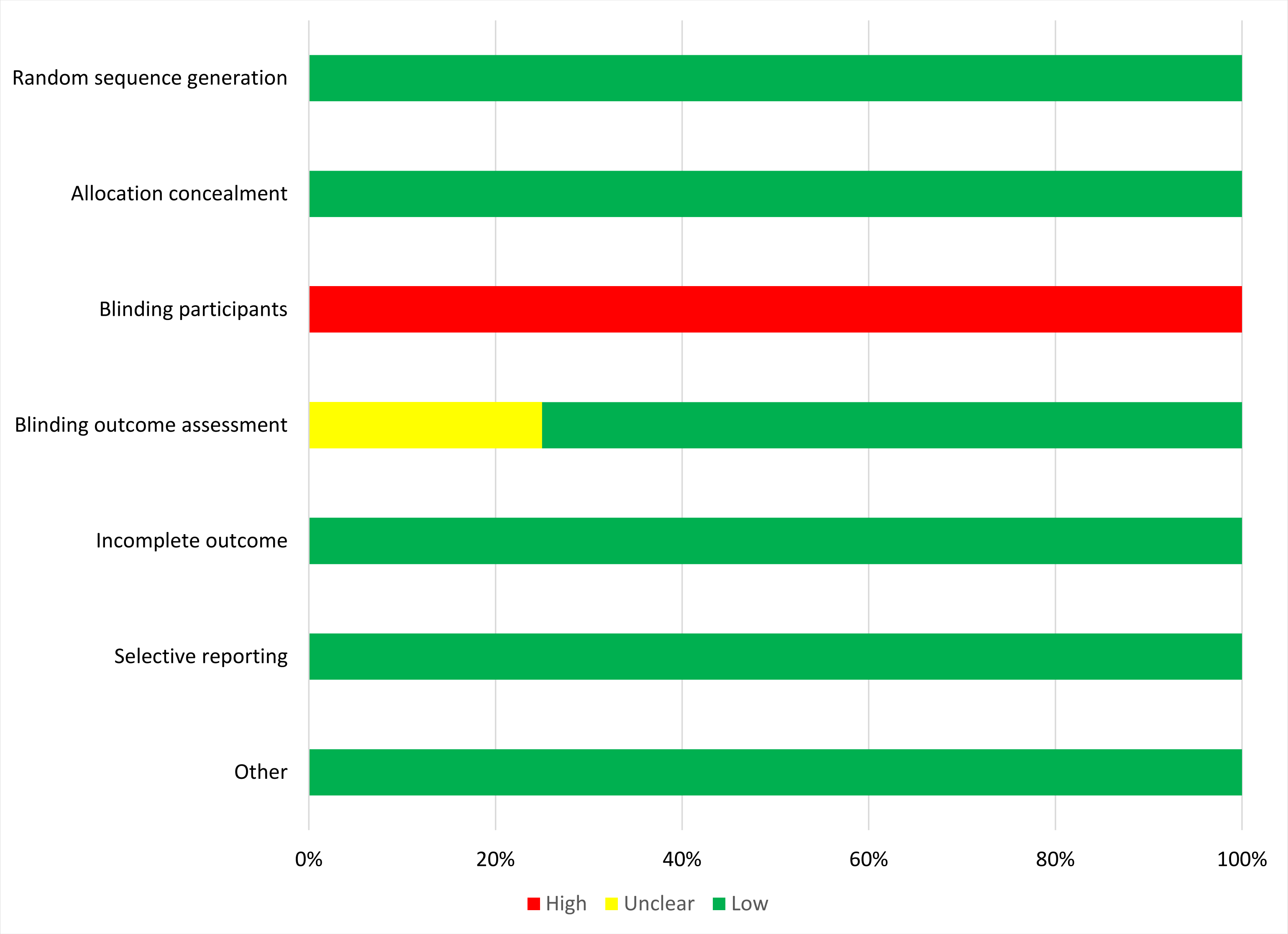

### Supplemental figure 2: Forest plots of main effects for all outcomes, SQ-LNS vs IFA/SOC

#### Contents

**Supplemental figure 2A: Mean difference in birth weight** 5

**Supplemental figure 2B: Mean difference in birth weight-for-age z score** 6

**Supplemental figure 2C: Mean difference in weight-for-gestational age z-score** 7

**Supplemental figure 2D: Low birth weight relative risk** 8

**Supplemental figure 2E: Low birth weight risk difference** 9

**Supplemental figure 2F: Birth weight < 2 kg relative risk** 10

**Supplemental figure 2G: Birth weight < 2 kg risk difference** 11

**Supplemental figure 2H: Small-for-gestational age relative risk** 12

**Supplemental figure 2I: Small-for-gestational age risk difference** 13

**Supplemental figure 2J: Large-for-gestational age relative risk** 14

**Supplemental figure 2K: Large-for-gestational age risk difference** 15

**Supplemental figure 2L: Mean difference in birth length** 16

**Supplemental figure 2M: Mean difference in birth length-for-age z score** 17

**Supplemental figure 2N: Mean difference in length-for-gestational age z-score** 18

**Supplemental figure 2O: Newborn stunting relative risk** **19**

**Supplemental figure 2P: Newborn stunting risk difference** **20**

**Supplemental figure 2Q: Low LGAZ relative risk** **21**

**Supplemental figure 2R: Low LGAZ risk difference** **22**

**Supplemental figure 2S: Mean difference in birth BMI-for-age z-score** **23**

**Supplemental figure 2T: Low BMIZ relative risk** **24**

**Supplemental figure 2U: Low BMIZ risk difference** **25**

**Supplemental figure 2V: Mean difference in birth head circumference** **26**

**Supplemental figure 2W: Mean difference in birth head circumference-for-age z score** **27**

**Supplemental figure 2X: Mean difference in birth head circumference-for-gestational age z score** **28**

**Supplemental figure 2Y: Low HCZ relative risk** **29**

**Supplemental figure 2Z: Low HCZ risk difference** **30**

**Supplemental figure 2AA: Low HCGAZ relative risk** **31**

**Supplemental figure 2AB: Low HCGAZ risk difference** **32**

**Supplemental figure 2AC: Mean difference in birth mid-upper arm circumference** **33**

**Supplemental figure 2AD: Mean difference in duration of gestation** **34**

**Supplemental figure 2AE: Preterm birth relative risk** **35**

**Supplemental figure 2AF: Preterm birth risk difference** **36**

**Supplemental figure 2AG: Mean difference in 6 mo weight-for-age z-score** **37**

**Supplemental figure 2AH: 6 mo underweight prevalence ratio** **38**

**Supplemental figure 2AI: 6 mo underweight prevalence difference** **39**

**Supplemental figure 2AJ: Mean difference in 6 mo length-for-age z-score** **40**

**Supplemental figure 2AK: 6 mo stunting prevalence ratio** **41**

**Supplemental figure 2AL: 6 mo stunting prevalence difference** **42**

**Supplemental figure 2AM: Mean difference in 6 mo weight-for-length z-score** **43**

**Supplemental figure 2AN: 6 mo wasting prevalence ratio** **44**

**Supplemental figure 2AO: 6 mo wasting prevalence difference** **45**

**Supplemental figure 2AP: Mean difference in 6 mo head circumference-for-age z-score** **46**

**Supplemental figure 2AQ: 6 mo low HCZ prevalence ratio** **47**

**Supplemental figure 2AR: 6 mo low HCZ prevalence difference** **48**

**Supplemental figure 2AS: Mean difference in 6 mo MUAC-for-age z-score** **49**

**Supplemental figure 2AT: 6 mo low MUAC prevalence ratio** **50**

**Supplemental figure 2AU: 6 mo low MUAC prevalence difference** **51**

**Supplemental figure 2AV: 6 mo acute malnutrition prevalence ratio** **52**

**Supplemental figure 2AW: 6 mo acute malnutrition prevalence difference** **53**

**Supplemental figure 2AX: Cesarian-section relative risk** **54**

**Supplemental figure 2AY: Cesarian-section risk difference** **55**

**Supplemental figure 2AZ: Miscarriage relative risk** **56**

**Supplemental figure 2BA: Miscarriage risk difference** **57**

**Supplemental figure 2BB: Stillbirth relative risk** **58**

**Supplemental figure 2BC: Stillbirth risk difference** **59**

**Supplemental figure 2BD: Miscarriage or stillbirth relative risk** **60**

**Supplemental figure 2BE: Miscarriage or stillbirth risk difference** **61**

**Supplemental figure 2BF: Early neonatal mortality relative risk** **62**

**Supplemental figure 2BG: Early neonatal mortality risk difference** **63**

**Supplemental figure 2BH: Miscarriage or stillbirth or early neonatal mortality relative risk** **64**

**Supplemental figure 2BI: Miscarriage or stillbirth or early neonatal mortality risk difference** **65**

**Supplemental figure 2BJ: Neonatal mortality relative risk** **66**

**Supplemental figure 2BK: Neonatal mortality risk difference** **67**

**Supplemental figure 2BL: Mortality 0-6 mo relative risk** **68**

**Supplemental figure 2BM: Mortality 0-6 mo risk difference** **69**

These figures are forest plots showing the study-level estimates of intervention effect with the pooled estimate in the bottom summary rows. Individual study estimates were generated from log binomial regression for dichotomous outcomes and simple linear regression for continuous outcomes with clustered observations using robust standard errors for cluster-randomized trials. Pooled estimates were generated using inverse variance weighting in both fixed and random effects models. For continuous outcomes the intervention effect is measured by the difference in mean of the SQ-LNS group minus IFA/SOC. For dichotomous outcomes analyzed via prevalence/risk ratios the effect estimate is the prevalence/risk in the SQ-LNS group divided by the prevalence/risk in the IFA/SOC group. For dichotomous outcomes analyzed via prevalence/risk differences the effect estimate is the prevalence/risk in the SQ-LNS group minus the prevalence/risk in the IFA/SOC group. The labels on the left y-axis correspond to trial level information. The values on the right indicate the study level effect estimate, confidence interval, and weighting for deriving the pooled estimate.

LAZ, length-for-age z-score; WLZ, weight-for-length z-score; WAZ, weight for-age z-score; MUACZ, mid-upper arm circumference z-score; BMI, body mass index; HCZ, head circumference-for-age z-score; LGAZ, length-for-gestational-age z-score; HCGAZ, head circumference-for-gestational-age z-score; BMIZ, body mass index-for-age z-score; IFA/SOC, Iron and folic acid or standard of care; MD, mean difference; MMS, multiple micronutrient supplement; MUAC, mid-upper arm circumference; PR, prevalence ratio; PD, prevalence difference; RD, risk difference; RR, relative risk; SOC, standard of care; SQ-LNS, small-quantity lipid-based nutrient supplements; WGAZ, weight-for-gestational age z-score.

### Supplemental figure 2A: Mean difference in birth weight

| Country | Trial | SQ−LNS N | IFA/SOC N | MD (95% CI) | Fixed W | Random W |
|---|---|---|---|---|---|---|
| Ghana | DYAD−G | 307 | 305 | 80 (12, 148) | 0.11 | 0.11 |
| Malawi | DYAD−M | 387 | 386 | 48 (−16, 112) | 0.12 | 0.12 |
| Bangladesh | RDNS | 898 | 2551 | 41 (11, 71) | 0.55 | 0.55 |
| Guatemala | WOMEN FIRST | 223 | 216 | 53 (4, 101) | 0.21 | 0.21 |
| **I² = 0.00, Tau² = 0.00** | | **1815** | **3458** | | | |
| **Fixed** | | | | **49 (26, 71)** | | |
| **Random** | | | | **49 (26, 71)** | | |

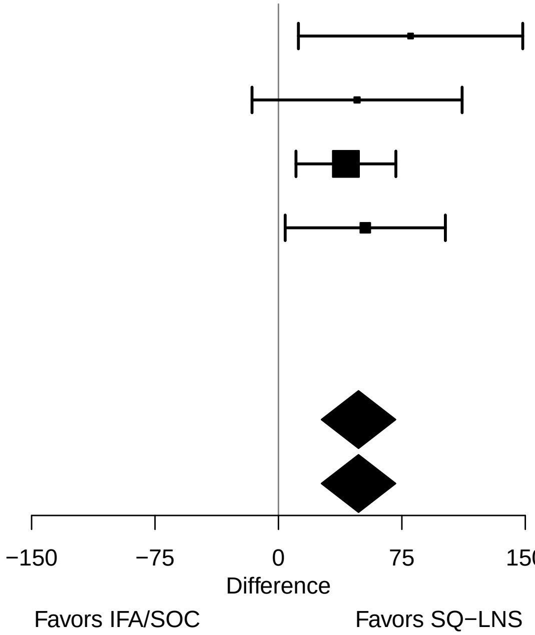

### Supplemental figure 2B: Mean difference in birth weight-for-age z score

| Country | Trial | SQ−LNS N | IFA/SOC N | MD (95% CI) | Fixed W | Random W |
|---|---|---|---|---|---|---|
| Ghana | DYAD−G | 307 | 305 | 0.18 (0.03, 0.33) | 0.13 | 0.13 |
| Malawi | DYAD−M | 387 | 386 | 0.11 (−0.04, 0.26) | 0.14 | 0.14 |
| Bangladesh | RDNS | 898 | 2551 | 0.10 (0.03, 0.18) | 0.54 | 0.54 |
| Guatemala | WOMEN FIRST | 223 | 216 | 0.13 (0.01, 0.26) | 0.20 | 0.20 |
| **I² = 0.00, Tau² = 0.00** | | **1815** | **3458** | | | |
| **Fixed** | | | | **0.12 (0.06, 0.17)** | | |
| **Random** | | | | **0.12 (0.06, 0.17)** | | |

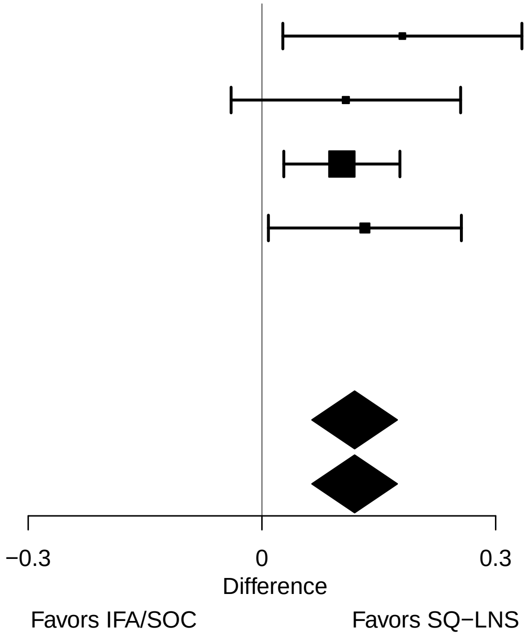

#### Supplemental figure 2C: Mean difference in weight-for-gestational age z-score

| Country | Trial | SQ−LNS N | IFA/SOC N | MD (95% CI) | Fixed W | Random W |
|---|---|---|---|---|---|---|
| Ghana | DYAD−G | 298 | 299 | 0.16 (0.01, 0.32) | 0.24 | 0.27 |
| Malawi | DYAD−M | 387 | 386 | 0.03 (−0.11, 0.17) | 0.30 | 0.31 |
| Bangladesh | RDNS | | | | | |
| Guatemala | WOMEN FIRST | 181 | 166 | 0.18 (0.07, 0.29) | 0.46 | 0.42 |
| **$I^2 = 0.31$, $Tau^2 = 0.00$** | | **866** | **851** | | | |
| **Fixed** | | | | **0.13 (0.05, 0.21)** | | |
| **Random** | | | | **0.13 (0.03, 0.22)** | | |

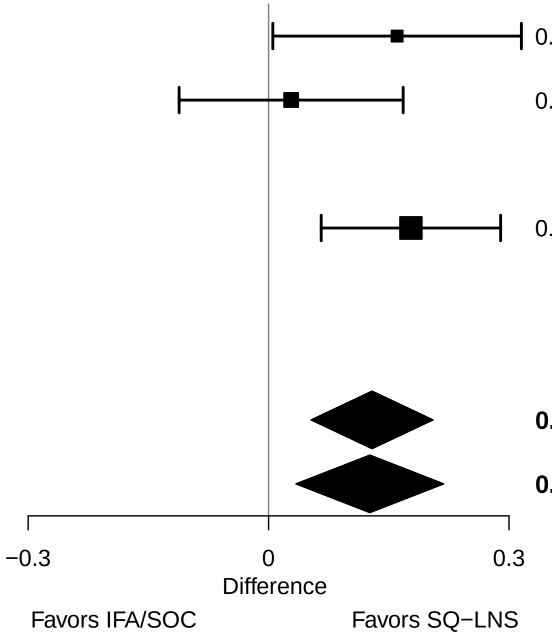

### Supplemental figure 2D: Low birth weight relative risk

| Country | Trial | SQ−LNS N | IFA/SOC N | RR (95% CI) | Fixed W | Random W |
|---|---|---|---|---|---|---|
| Ghana | DYAD−G | 307 | 305 | 0.62 (0.39, 0.98) | 0.05 | 0.05 |
| Malawi | DYAD−M | 387 | 386 | 0.92 (0.64, 1.32) | 0.08 | 0.08 |
| Bangladesh | RDNS | 898 | 2551 | 0.92 (0.82, 1.03) | 0.78 | 0.78 |
| Guatemala | WOMEN FIRST | 223 | 216 | 0.84 (0.60, 1.19) | 0.09 | 0.09 |
| **$I^2$ = 0.00, $Tau^2$ = 0.00** | | **1815** | **3458** | | | |
| **Fixed** | | | | **0.89 (0.80, 0.99)** | | |
| **Random** | | | | **0.89 (0.80, 0.99)** | | |

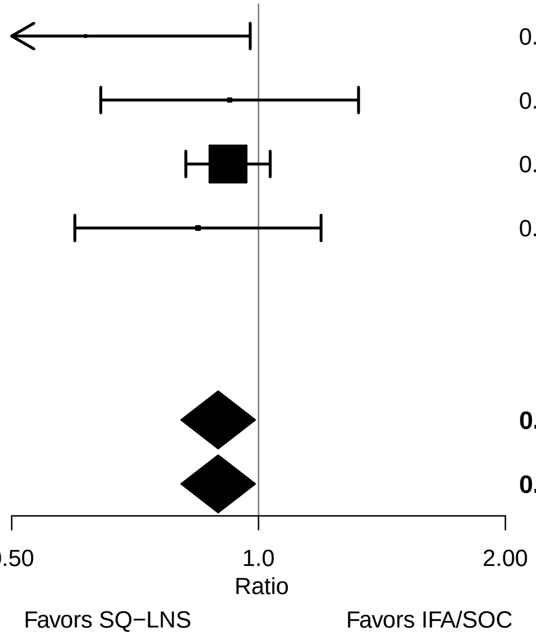

#### Supplemental figure 2E: Low birth weight risk difference

| Country | Trial | SQ−LNS N | IFA/SOC N | RD (95% CI) | Fixed W | Random W |
|---|---|---|---|---|---|---|
| Ghana | DYAD−G | 307 | 305 | −0.05 (−0.10, 0.00) | 0.21 | 0.21 |
| Malawi | DYAD−M | 387 | 386 | −0.01 (−0.06, 0.04) | 0.22 | 0.22 |
| Bangladesh | RDNS | 898 | 2551 | −0.03 (−0.07, 0.01) | 0.31 | 0.31 |
| Guatemala | WOMEN FIRST | 223 | 216 | −0.02 (−0.07, 0.02) | 0.26 | 0.26 |
| **I² = 0.00, Tau² = 0.00** | | **1815** | **3458** | | | |
| **Fixed** | | | | **−0.03 (−0.05, −0.01)** | | |
| **Random** | | | | **−0.03 (−0.05, −0.01)** | | |

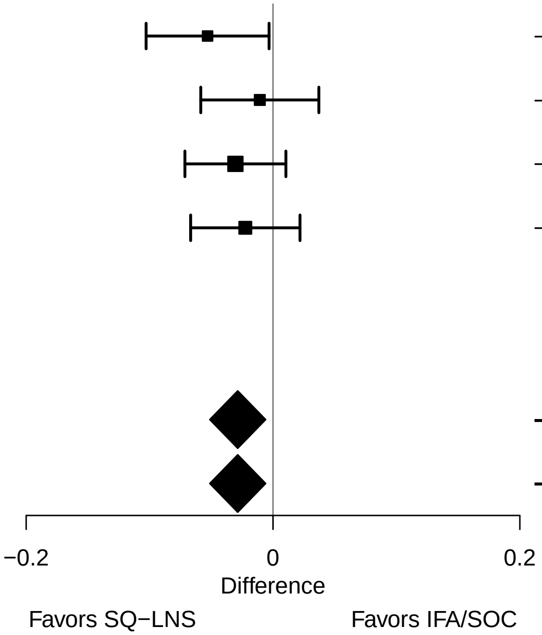

#### Supplemental figure 2F: Birth weight < 2 kg relative risk

| Country | Trial | SQ−LNS N | IFA/SOC N | RR (95% CI) | Fixed W | Random W |
|---|---|---|---|---|---|---|
| Ghana | DYAD−G | 307 | 305 | 0.43 (0.11, 1.63) | 0.04 | 0.04 |
| Malawi | DYAD−M | 387 | 386 | 1.00 (0.45, 2.19) | 0.11 | 0.11 |
| Bangladesh | RDNS | 898 | 2551 | 0.78 (0.59, 1.04) | 0.84 | 0.84 |
| Guatemala | WOMEN FIRST | 223 | 216 | 0.48 (0.04, 5.69) | 0.01 | 0.01 |
| **I² = 0.00, Tau² = 0.00** | | **1815** | **3458** | | | |
| **Fixed** | | | | **0.78 (0.60, 1.01)** | | |
| **Random** | | | | **0.78 (0.60, 1.01)** | | |

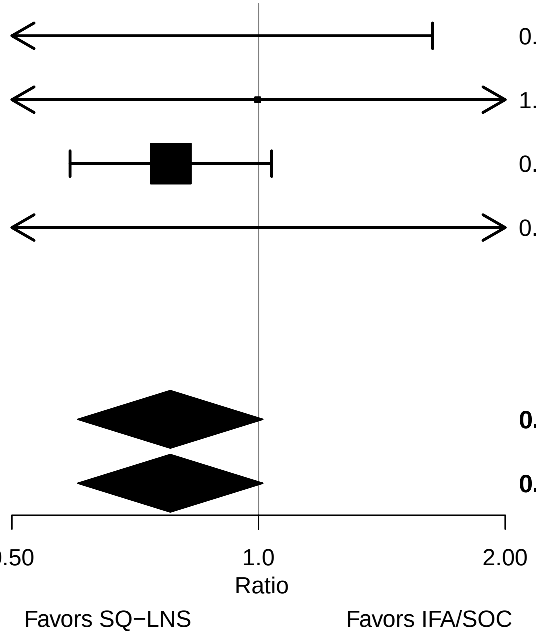

#### Supplemental figure 2G: Birth weight < 2 kg risk difference

| Country | Trial | SQ−LNS N | IFA/SOC N | RD (95% CI) | Fixed W | Random W |
|---|---|---|---|---|---|---|
| Ghana | DYAD−G | 307 | 305 | −0.01 (−0.03, 0.01) | 0.21 | 0.21 |
| Malawi | DYAD−M | 387 | 386 | 0.00 (−0.02, 0.02) | 0.14 | 0.14 |
| Bangladesh | RDNS | 898 | 2551 | −0.02 (−0.03, 0.00) | 0.29 | 0.29 |
| Guatemala | WOMEN FIRST | 223 | 216 | 0.00 (−0.02, 0.01) | 0.36 | 0.36 |
| **I² = 0.00, Tau² = 0.00** | | **1815** | **3458** | | | |
| **Fixed** | | | | **−0.01 (−0.02, 0.00)** | | |
| **Random** | | | | **−0.01 (−0.02, 0.00)** | | |

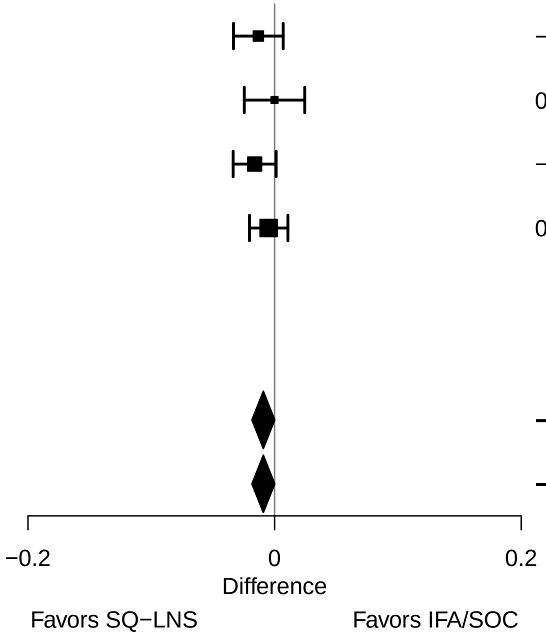

### Supplemental figure 2H: Small-for-gestational age relative risk

| Country | Trial | SQ−LNS N | IFA/SOC N | RR (95% CI) | Fixed W | Random W |
|---|---|---|---|---|---|---|
| Ghana | DYAD−G | 307 | 305 | 0.82 (0.60, 1.12) | 0.02 | 0.02 |
| Malawi | DYAD−M | 387 | 386 | 0.96 (0.75, 1.22) | 0.04 | 0.04 |
| Bangladesh | RDNS | 898 | 2551 | 0.97 (0.92, 1.02) | 0.91 | 0.91 |
| Guatemala | WOMEN FIRST | 181 | 166 | 0.77 (0.56, 1.07) | 0.02 | 0.02 |
| **I² = 0.00, Tau² = 0.00** | | **1773** | **3408** | | | |
| **Fixed** | | | | **0.96 (0.92, 1.01)** | | |
| **Random** | | | | **0.96 (0.92, 1.01)** | | |

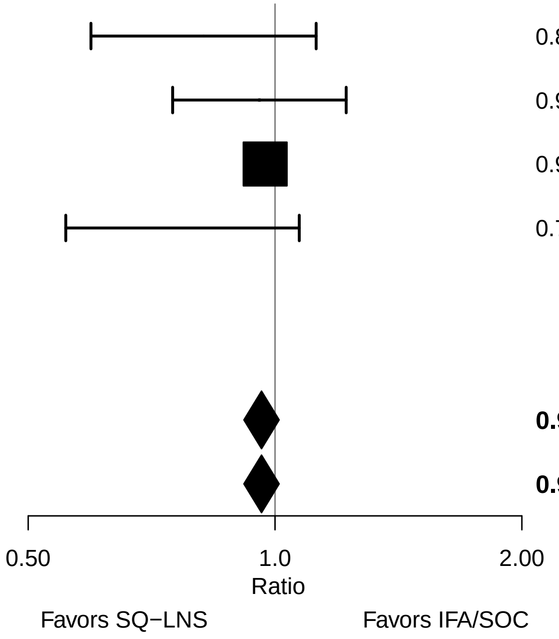

#### Supplemental figure 2I: Small-for-gestational age risk difference

| Country | Trial | SQ−LNS N | IFA/SOC N | RD (95% CI) | Fixed W | Random W |
|---|---|---|---|---|---|---|
| Ghana | DYAD−G | 307 | 305 | −0.04 (−0.10, 0.02) | 0.14 | 0.14 |
| Malawi | DYAD−M | 387 | 386 | −0.01 (−0.07, 0.05) | 0.15 | 0.15 |
| Bangladesh | RDNS | 898 | 2551 | −0.02 (−0.05, 0.01) | 0.62 | 0.62 |
| Guatemala | WOMEN FIRST | 181 | 166 | −0.06 (−0.14, 0.02) | 0.10 | 0.10 |
| **I² = 0.00, Tau² = 0.00** | | **1773** | **3408** | | | |
| **Fixed** | | | | **−0.02 (−0.05, 0.00)** | | |
| **Random** | | | | **−0.02 (−0.05, 0.00)** | | |

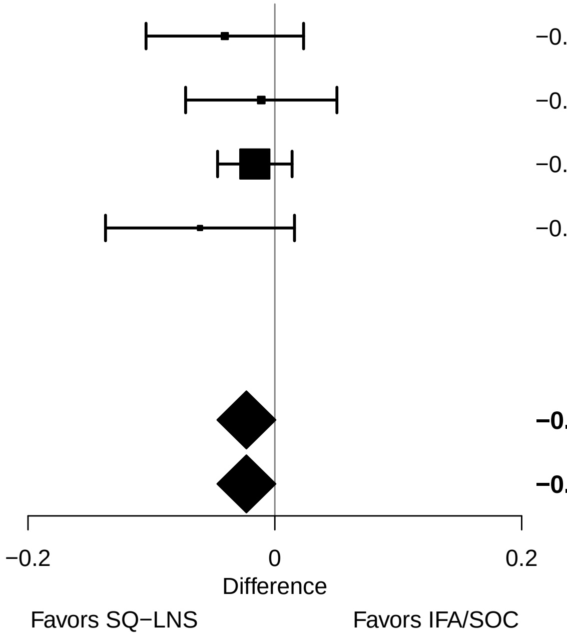

#### Supplemental figure 2J: Large-for-gestational age relative risk

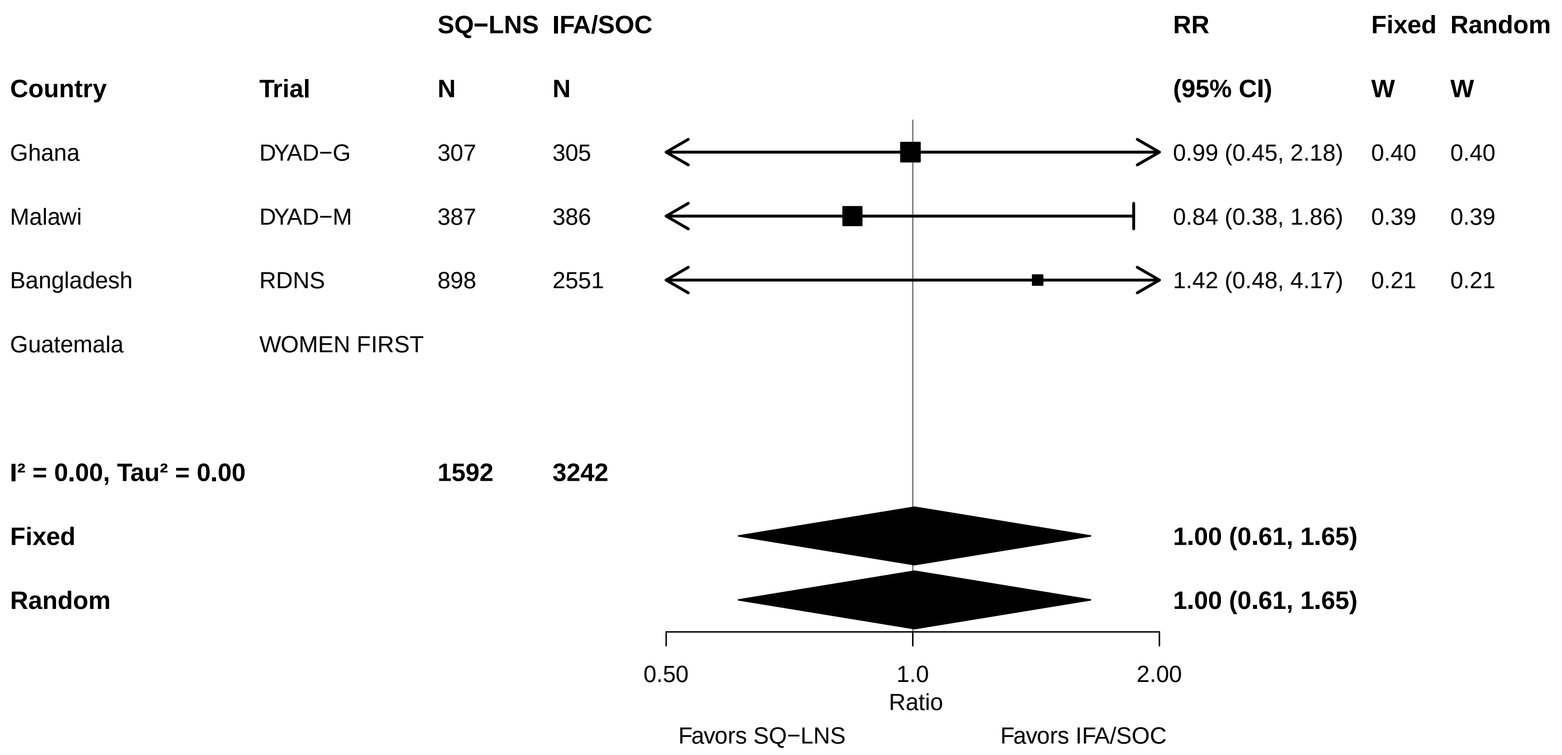

### Supplemental figure 2K: Large-for-gestational age risk difference

| | | SQ−LNS | IFA/SOC | RD | Fixed | Random |
|---|---|---|---|---|---|---|
| **Country** | **Trial** | **N** | **N** | **(95% CI)** | **W** | **W** |
| Ghana | DYAD−G | 307 | 305 | 0.00 (−0.03, 0.03) | 0.02 | 0.02 |
| Malawi | DYAD−M | 387 | 386 | −0.01 (−0.03, 0.02) | 0.03 | 0.03 |
| Bangladesh | RDNS | 898 | 2551 | 0.00 (0.00, 0.01) | 0.95 | 0.95 |
| Guatemala | WOMEN FIRST | | | | | |
| **I² = 0.00, Tau² = 0.00** | | **1592** | **3242** | | | |
| **Fixed** | | | | **0.00 (0.00, 0.01)** | | |
| **Random** | | | | **0.00 (0.00, 0.01)** | | |

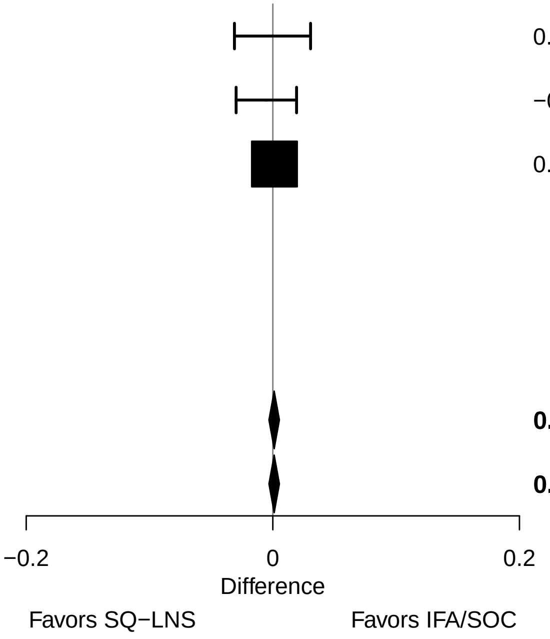

#### Supplemental figure 2L: Mean difference in birth length

| Country | Trial | SQ−LNS N | IFA/SOC N | MD (95% CI) | Fixed W | Random W |
|---|---|---|---|---|---|---|
| Ghana | DYAD−G | 307 | 305 | 0.25 (−0.05, 0.55) | 0.12 | 0.12 |
| Malawi | DYAD−M | 253 | 263 | 0.30 (−0.07, 0.66) | 0.08 | 0.08 |
| Bangladesh | RDNS | 897 | 2550 | 0.17 (0.03, 0.31) | 0.57 | 0.57 |
| Guatemala | WOMEN FIRST | 223 | 216 | 0.22 (−0.01, 0.44) | 0.23 | 0.23 |
| **I² = 0.00, Tau² = 0.00** | | **1680** | **3334** | | | |
| **Fixed** | | | | **0.20 (0.09, 0.31)** | | |
| **Random** | | | | **0.20 (0.09, 0.31)** | | |

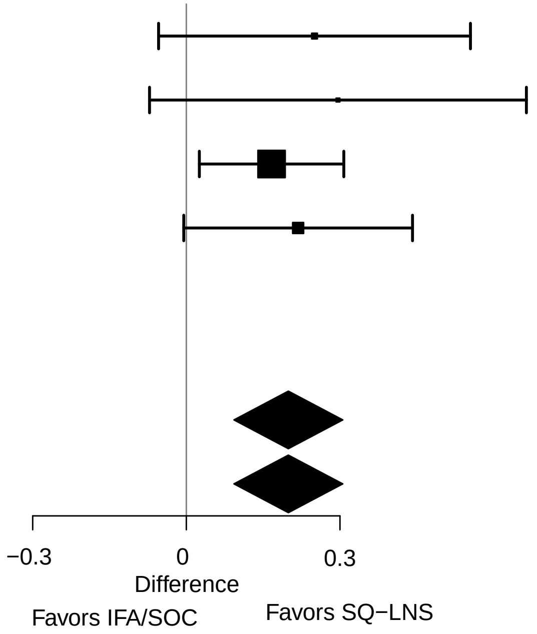

#### Supplemental figure 2M: Mean difference in birth length-for-age z score

| Country | Trial | SQ−LNS N | IFA/SOC N | MD (95% CI) | Fixed W | Random W |
|---|---|---|---|---|---|---|
| Ghana | DYAD−G | 307 | 305 | 0.12 (−0.04, 0.28) | 0.13 | 0.13 |
| Malawi | DYAD−M | 253 | 263 | 0.17 (−0.02, 0.37) | 0.09 | 0.09 |
| Bangladesh | RDNS | 897 | 2550 | 0.09 (0.02, 0.17) | 0.60 | 0.60 |
| Guatemala | WOMEN FIRST | 223 | 216 | 0.14 (0.01, 0.27) | 0.19 | 0.19 |
| **I² = 0.00, Tau² = 0.00** | | **1680** | **3334** | | | |
| **Fixed** | | | | **0.11 (0.06, 0.17)** | | |
| **Random** | | | | **0.11 (0.06, 0.17)** | | |

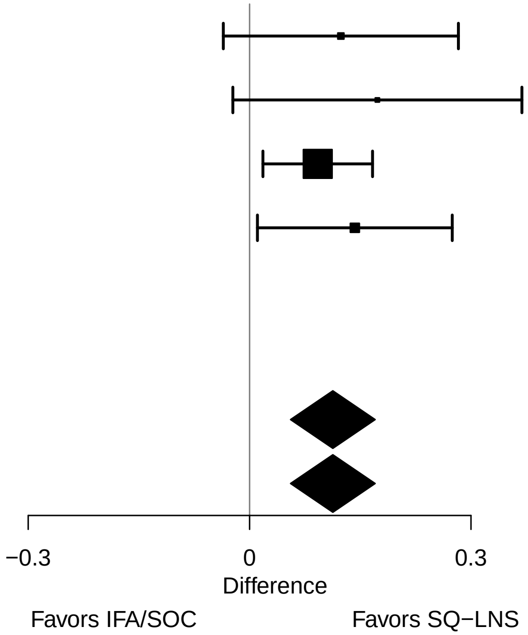

#### Supplemental figure 2N: Mean difference in length-for-gestational age z-score

| Country | Trial | SQ−LNS N | IFA/SOC N | MD (95% CI) | Fixed W | Random W |
|---|---|---|---|---|---|---|
| Ghana | DYAD−G | 298 | 299 | 0.10 (−0.06, 0.26) | 0.26 | 0.26 |
| Malawi | DYAD−M | 253 | 263 | 0.09 (−0.11, 0.28) | 0.17 | 0.17 |
| Bangladesh | RDNS | | | | | |
| Guatemala | WOMEN FIRST | 181 | 166 | 0.15 (0.04, 0.26) | 0.57 | 0.57 |
| **I² = 0.00, Tau² = 0.00** | | **732** | **728** | | | |
| **Fixed** | | | | **0.13 (0.05, 0.21)** | | |
| **Random** | | | | **0.13 (0.05, 0.21)** | | |

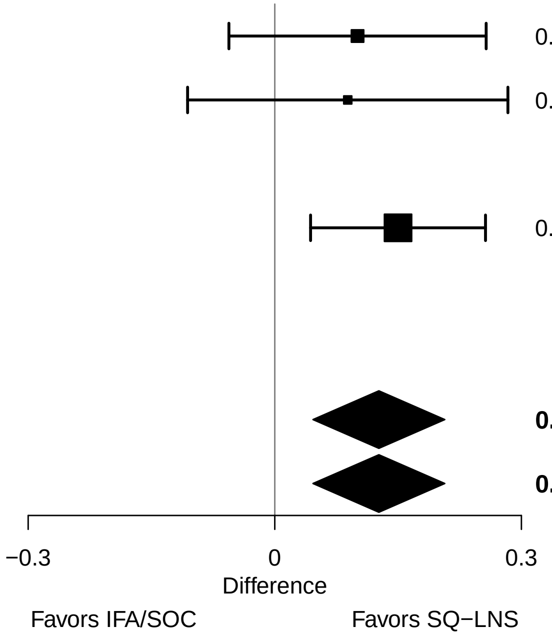

#### Supplemental figure 2O: Newborn stunting relative risk

| Country | Trial | SQ−LNS N | IFA/SOC N | RR (95% CI) | Fixed W | Random W |
|---|---|---|---|---|---|---|
| Ghana | DYAD−G | 307 | 305 | 0.99 (0.61, 1.61) | 0.06 | 0.06 |
| Malawi | DYAD−M | 253 | 263 | 0.85 (0.57, 1.28) | 0.08 | 0.08 |
| Bangladesh | RDNS | 897 | 2550 | 0.83 (0.73, 0.95) | 0.74 | 0.74 |
| Guatemala | WOMEN FIRST | 223 | 216 | 0.73 (0.53, 1.02) | 0.12 | 0.12 |
| **I² = 0.00, Tau² = 0.00** | | **1680** | **3334** | | | |
| **Fixed** | | | | **0.83 (0.74, 0.93)** | | |
| **Random** | | | | **0.83 (0.74, 0.93)** | | |

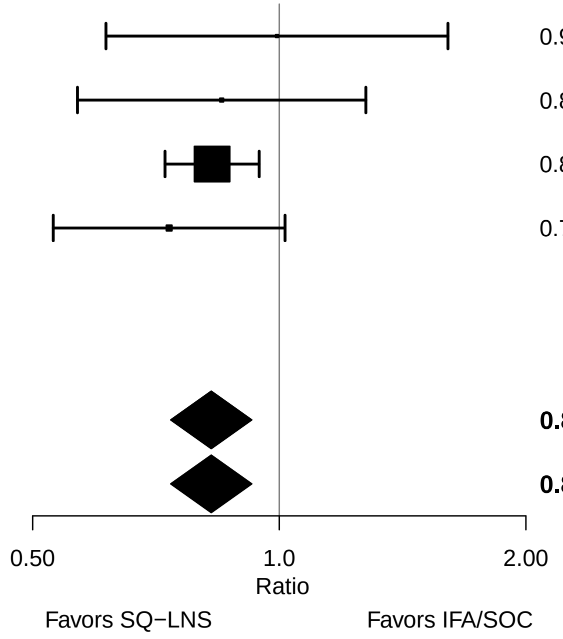

#### Supplemental figure 2P: Newborn stunting risk difference

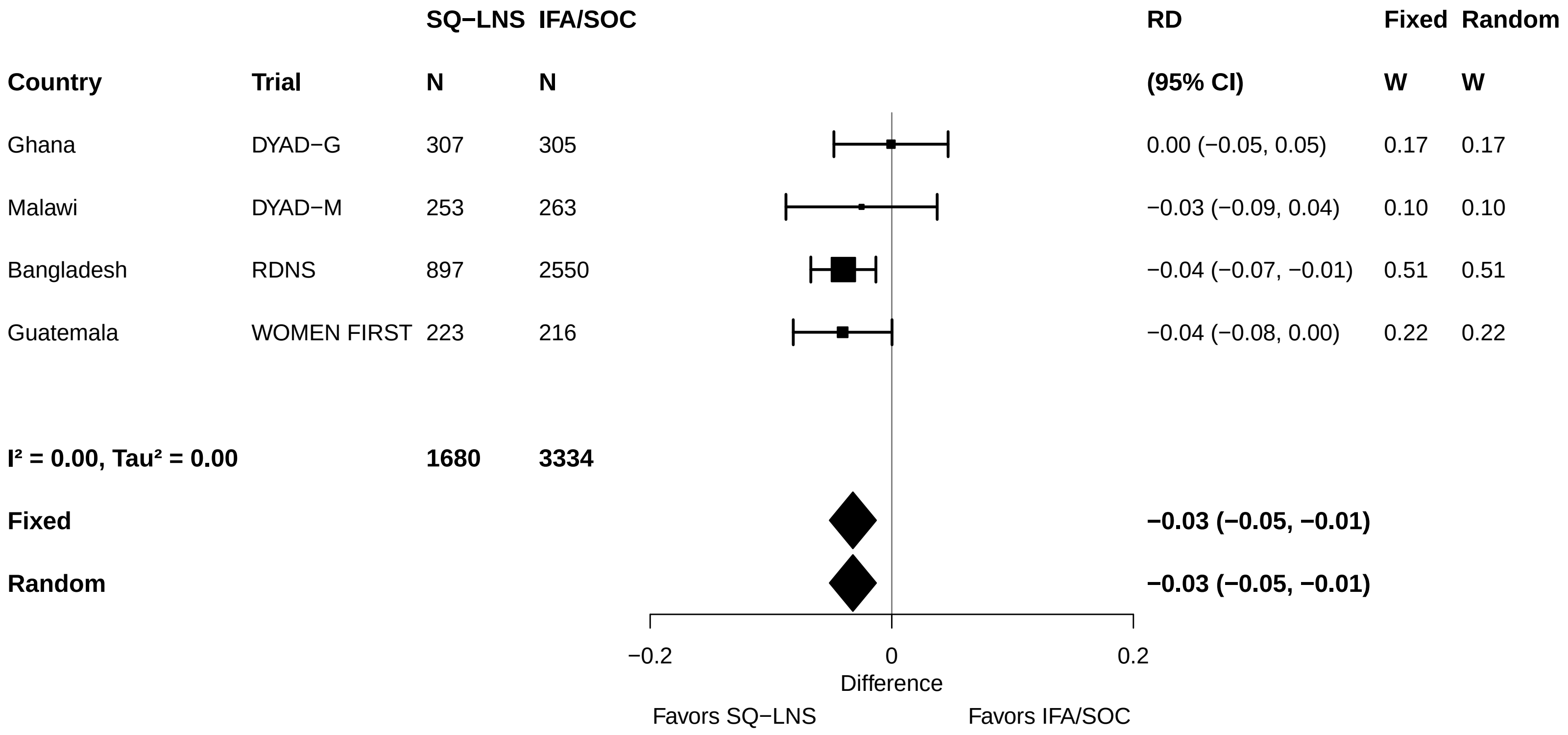

#### Supplemental figure 2Q: Low LGAZ relative risk

| Country | Trial | SQ−LNS N | IFA/SOC N | RR (95% CI) | Fixed W | Random W |
|---|---|---|---|---|---|---|
| Ghana | DYAD−G | 307 | 305 | 1.19 (0.61, 2.32) | 0.03 | 0.03 |
| Malawi | DYAD−M | 253 | 263 | 0.93 (0.63, 1.36) | 0.09 | 0.09 |
| Bangladesh | RDNS | 897 | 2550 | 0.89 (0.78, 1.01) | 0.83 | 0.83 |
| Guatemala | WOMEN FIRST | 181 | 166 | 0.85 (0.49, 1.46) | 0.05 | 0.05 |
| **I² = 0.00, Tau² = 0.00** | | **1638** | **3284** | | | |
| **Fixed** | | | | **0.90 (0.80, 1.01)** | | |
| **Random** | | | | **0.90 (0.80, 1.01)** | | |

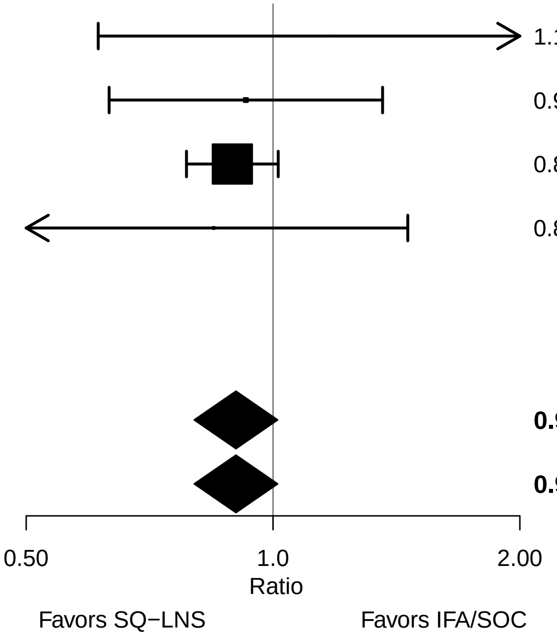

#### Supplemental figure 2R: Low LGAZ risk difference

| Country | Trial | SQ−LNS N | IFA/SOC N | RD (95% CI) | Fixed W | Random W |
|---|---|---|---|---|---|---|
| Ghana | DYAD−G | 307 | 305 | 0.01 (−0.03, 0.05) | 0.24 | 0.24 |
| Malawi | DYAD−M | 253 | 263 | −0.01 (−0.08, 0.05) | 0.07 | 0.07 |
| Bangladesh | RDNS | 897 | 2550 | −0.02 (−0.05, 0.00) | 0.49 | 0.49 |
| Guatemala | WOMEN FIRST | 181 | 166 | −0.01 (−0.05, 0.03) | 0.19 | 0.19 |
| **I² = 0.00, Tau² = 0.00** | | **1638** | **3284** | | | |
| **Fixed** | | | | **−0.01 (−0.03, 0.01)** | | |
| **Random** | | | | **−0.01 (−0.03, 0.01)** | | |

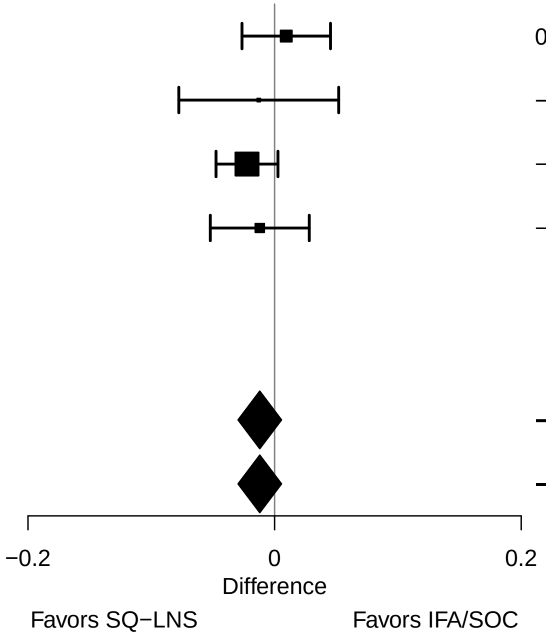

#### Supplemental figure 2S: Mean difference in birth BMI-for-age z-score

| Country | Trial | SQ−LNS N | IFA/SOC N | MD (95% CI) | Fixed W | Random W |
|---|---|---|---|---|---|---|
| Ghana | DYAD−G | 307 | 305 | 0.19 (0.03, 0.36) | 0.14 | 0.14 |
| Malawi | DYAD−M | 249 | 255 | 0.01 (−0.19, 0.21) | 0.10 | 0.10 |
| Bangladesh | RDNS | 897 | 2550 | 0.09 (0.01, 0.18) | 0.55 | 0.55 |
| Guatemala | WOMEN FIRST | 223 | 216 | 0.09 (−0.04, 0.22) | 0.22 | 0.22 |
| **I² = 0.00, Tau² = 0.00** | | **1676** | **3326** | | | |
| **Fixed** | | | | **0.10 (0.04, 0.16)** | | |
| **Random** | | | | **0.10 (0.04, 0.16)** | | |

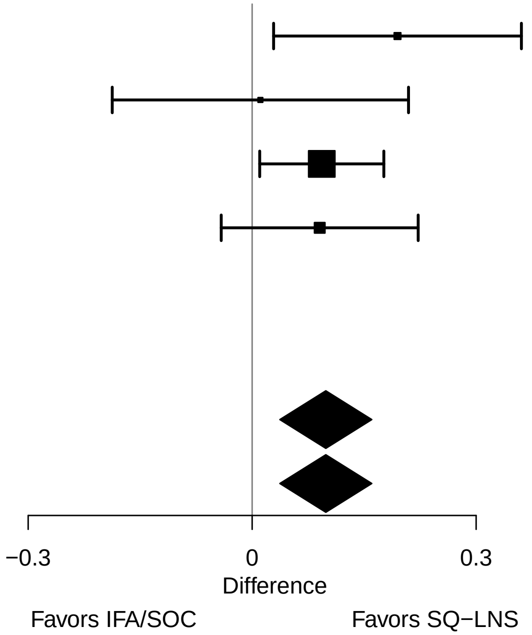

#### Supplemental figure 2T: Low BMIZ relative risk

| Country | Trial | SQ−LNS N | IFA/SOC N | RR (95% CI) | Fixed W | Random W |
|---|---|---|---|---|---|---|
| Ghana | DYAD−G | 307 | 305 | 0.54 (0.31, 0.94) | 0.03 | 0.15 |
| Malawi | DYAD−M | 249 | 255 | 1.14 (0.62, 2.10) | 0.02 | 0.12 |
| Bangladesh | RDNS | 897 | 2550 | 0.91 (0.82, 1.00) | 0.92 | 0.62 |
| Guatemala | WOMEN FIRST | 223 | 216 | 0.76 (0.40, 1.46) | 0.02 | 0.11 |
| **I² = 0.26, Tau² = 0.02** | | **1676** | **3326** | | | |
| **Fixed** | | | | **0.89 (0.81, 0.98)** | | |
| **Random** | | | | **0.85 (0.67, 1.08)** | | |

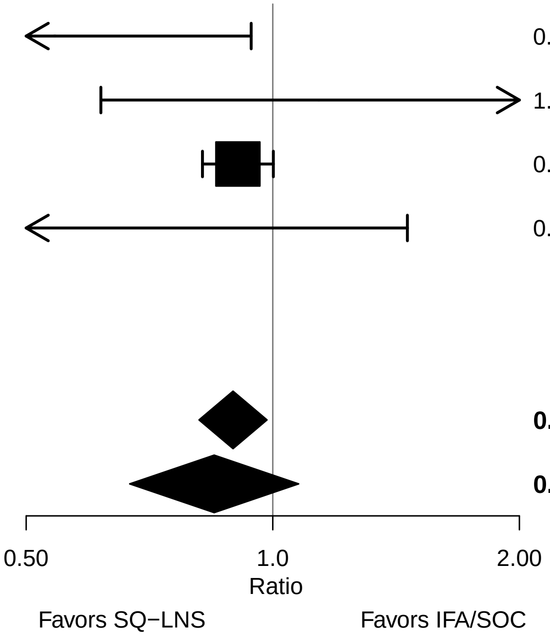

#### Supplemental figure 2U: Low BMIZ risk difference

| Country | Trial | SQ−LNS N | IFA/SOC N | RD (95% CI) | Fixed W | Random W |
|---|---|---|---|---|---|---|
| Ghana | DYAD−G | 307 | 305 | −0.05 (−0.09, −0.01) | 0.19 | 0.21 |
| Malawi | DYAD−M | 249 | 255 | 0.01 (−0.04, 0.06) | 0.17 | 0.19 |
| Bangladesh | RDNS | 897 | 2550 | −0.03 (−0.06, 0.00) | 0.37 | 0.34 |
| Guatemala | WOMEN FIRST | 223 | 216 | −0.02 (−0.05, 0.02) | 0.27 | 0.27 |
| **$I^2$ = 0.20, $Tau^2$ = 0.00** | | **1676** | **3326** | | | |
| **Fixed** | | | | **−0.02 (−0.04, 0.00)** | | |
| **Random** | | | | **−0.02 (−0.05, 0.00)** | | |

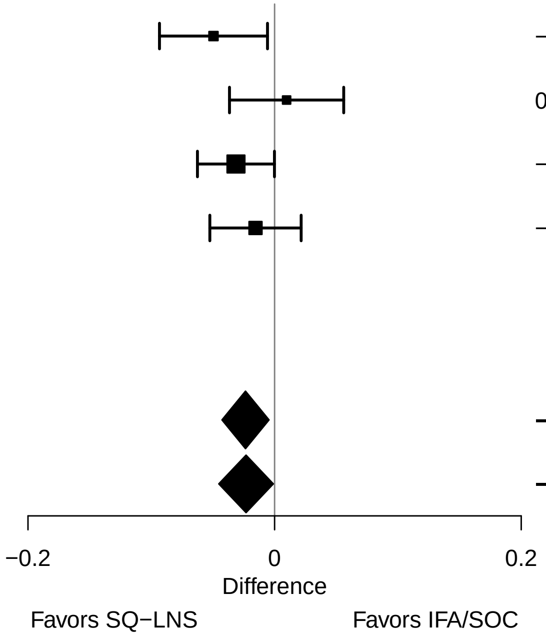

### Supplemental figure 2V: Mean difference in birth head circumference

| **Country** | **Trial** | **SQ−LNS N** | **IFA/SOC N** | **MD (95% CI)** | **Fixed W** | **Random W** |
|---|---|---|---|---|---|---|
| Ghana | DYAD−G | 307 | 305 | 0.29 (0.05, 0.53) | 0.09 | 0.09 |
| Malawi | DYAD−M | 253 | 264 | 0.13 (−0.11, 0.36) | 0.09 | 0.09 |
| Bangladesh | RDNS | 897 | 2551 | 0.10 (0.01, 0.18) | 0.63 | 0.63 |
| Guatemala | WOMEN FIRST | 223 | 216 | 0.07 (−0.09, 0.24) | 0.19 | 0.19 |
| **$I^2$ = 0.00, $Tau^2$ = 0.00** | | **1680** | **3336** | | | |
| **Fixed** | | | | **0.11 (0.04, 0.18)** | | |
| **Random** | | | | **0.11 (0.04, 0.18)** | | |

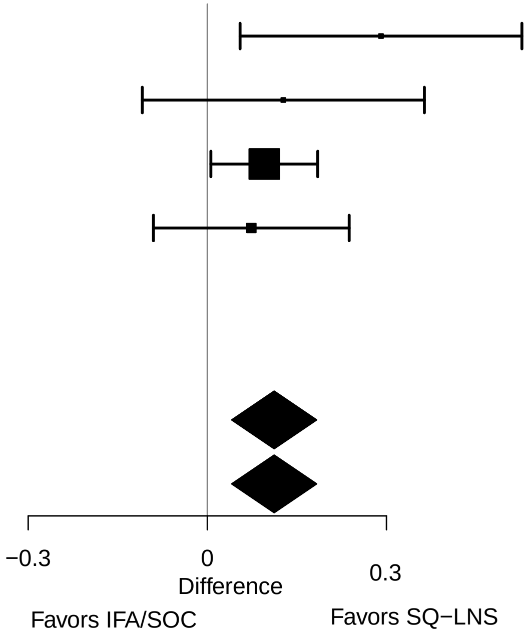

#### Supplemental figure 2W: Mean difference in birth head circumference-for-age z score

| Country | Trial | SQ−LNS N | IFA/SOC N | MD (95% CI) | Fixed W | Random W |
|---|---|---|---|---|---|---|
| Ghana | DYAD−G | 307 | 305 | 0.21 (0.03, 0.39) | 0.10 | 0.10 |
| Malawi | DYAD−M | 253 | 264 | 0.12 (−0.07, 0.31) | 0.09 | 0.09 |
| Bangladesh | RDNS | 897 | 2551 | 0.08 (0.01, 0.16) | 0.62 | 0.62 |
| Guatemala | WOMEN FIRST | 223 | 216 | 0.09 (−0.04, 0.23) | 0.19 | 0.19 |
| **I² = 0.00, Tau² = 0.00** | | **1680** | **3336** | | | |
| **Fixed** | | | | **0.10 (0.04, 0.16)** | | |
| **Random** | | | | **0.10 (0.04, 0.16)** | | |

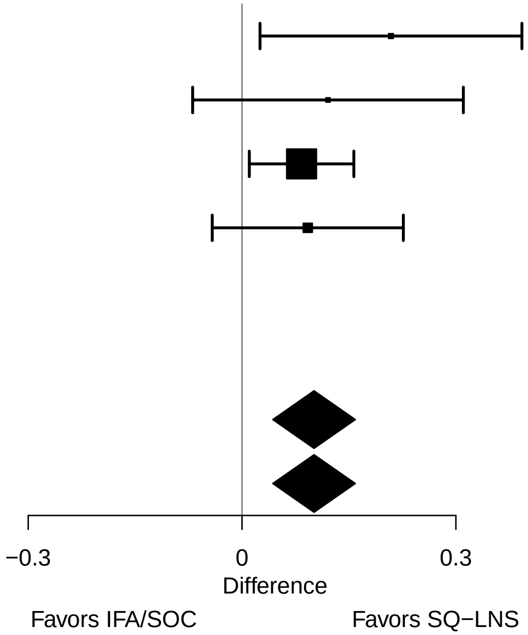

#### Supplemental figure 2X: Mean difference in birth head circumference-for-gestational age z score

| Country | Trial | SQ−LNS N | IFA/SOC N | MD (95% CI) | Fixed W | Random W |
|---|---|---|---|---|---|---|
| Ghana | DYAD−G | 298 | 299 | 0.19 (0.02, 0.37) | 0.27 | 0.27 |
| Malawi | DYAD−M | 253 | 264 | 0.06 (−0.12, 0.24) | 0.27 | 0.27 |
| Bangladesh | RDNS | | | | | |
| Guatemala | WOMEN FIRST | 181 | 166 | 0.09 (−0.04, 0.23) | 0.46 | 0.46 |
| **I² = 0.00, Tau² = 0.00** | | **732** | **729** | | | |
| **Fixed** | | | | **0.11 (0.02, 0.20)** | | |
| **Random** | | | | **0.11 (0.02, 0.20)** | | |

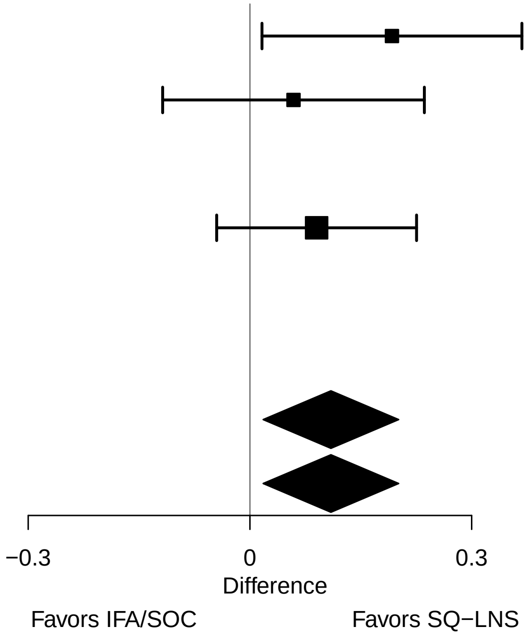

#### Supplemental figure 2Y: Low HCZ relative risk

| Country | Trial | SQ−LNS N | IFA/SOC N | RR (95% CI) | Fixed W | Random W |
|---|---|---|---|---|---|---|
| Ghana | DYAD−G | 307 | 305 | 0.83 (0.47, 1.47) | 0.05 | 0.05 |
| Malawi | DYAD−M | 253 | 264 | 0.78 (0.38, 1.62) | 0.03 | 0.03 |
| Bangladesh | RDNS | 897 | 2551 | 0.85 (0.75, 0.96) | 0.89 | 0.89 |
| Guatemala | WOMEN FIRST | 223 | 216 | 0.97 (0.53, 1.77) | 0.04 | 0.04 |
| **I² = 0.00, Tau² = 0.00** | | **1680** | **3336** | | | |
| **Fixed** | | | | **0.85 (0.75, 0.96)** | | |
| **Random** | | | | **0.85 (0.75, 0.96)** | | |

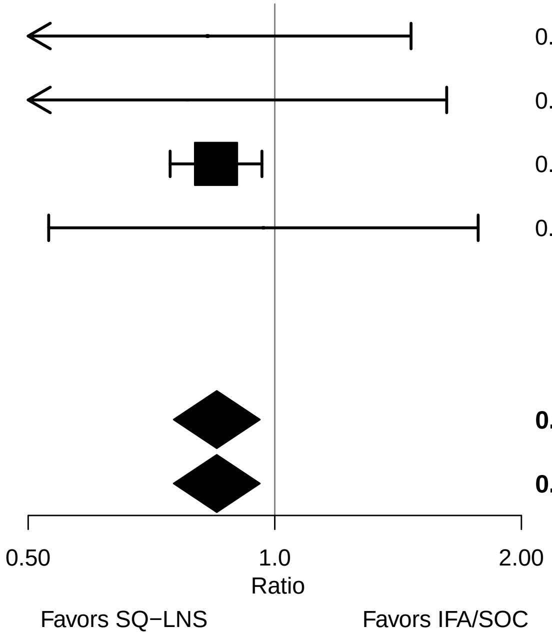

### Supplemental figure 2Z: Low HCZ risk difference

| Country | Trial | SQ−LNS N | IFA/SOC N | RD (95% CI) | Fixed W | Random W |
|---|---|---|---|---|---|---|
| Ghana | DYAD−G | 307 | 305 | −0.01 (−0.05, 0.03) | 0.20 | 0.20 |
| Malawi | DYAD−M | 253 | 264 | −0.01 (−0.05, 0.03) | 0.22 | 0.22 |
| Bangladesh | RDNS | 897 | 2551 | −0.04 (−0.07, −0.01) | 0.42 | 0.42 |
| Guatemala | WOMEN FIRST | 223 | 216 | 0.00 (−0.05, 0.04) | 0.15 | 0.15 |
| **$I^2$ = 0.00, $Tau^2$ = 0.00** | | **1680** | **3336** | | | |
| **Fixed** | | | | **−0.02 (−0.04, 0.00)** | | |
| **Random** | | | | **−0.02 (−0.04, 0.00)** | | |

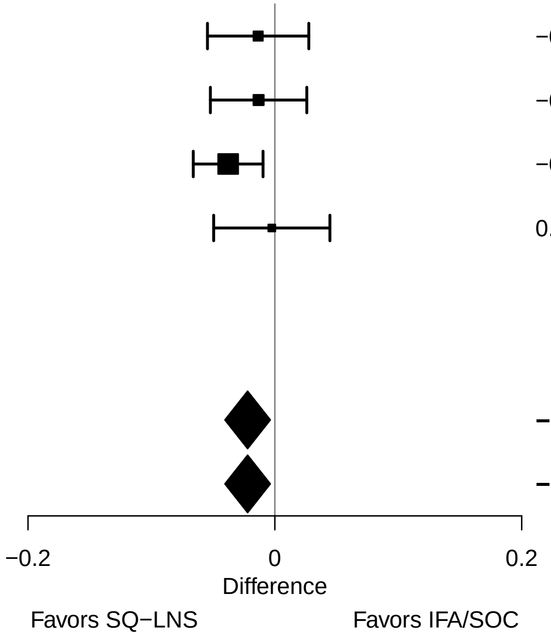

#### Supplemental figure 2AA: Low HCGAZ relative risk

| Country | Trial | SQ−LNS N | IFA/SOC N | RR (95% CI) | Fixed W | Random W |
|---|---|---|---|---|---|---|
| Ghana | DYAD−G | 307 | 305 | 0.50 (0.17, 1.44) | 0.02 | 0.02 |
| Malawi | DYAD−M | 253 | 264 | 2.09 (0.39, 11.29) | 0.01 | 0.01 |
| Bangladesh | RDNS | 897 | 2551 | 0.88 (0.76, 1.02) | 0.96 | 0.96 |
| Guatemala | WOMEN FIRST | 181 | 166 | 0.73 (0.23, 2.32) | 0.02 | 0.02 |
| **I² = 0.00, Tau² = 0.00** | | **1638** | **3286** | | | |
| **Fixed** | | | | **0.88 (0.76, 1.01)** | | |
| **Random** | | | | **0.88 (0.76, 1.01)** | | |

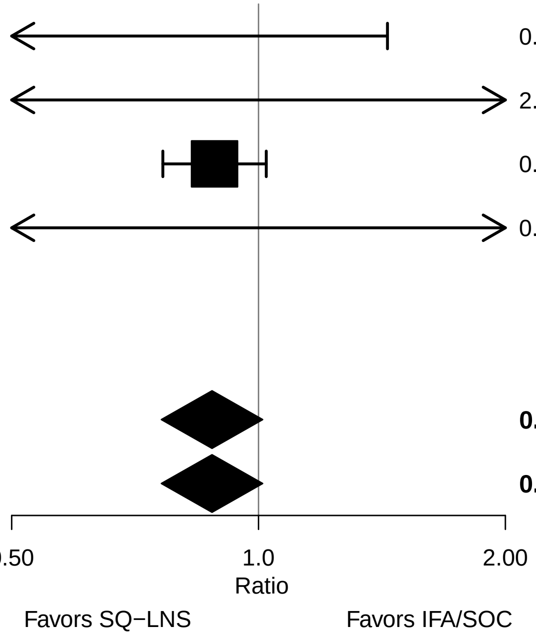

### Supplemental figure 2AB: Low HCGAZ risk difference

| Country | Trial | SQ−LNS N | IFA/SOC N | RD (95% CI) | Fixed W | Random W |
|---|---|---|---|---|---|---|
| Ghana | DYAD−G | 307 | 305 | −0.02 (−0.04, 0.01) | 0.24 | 0.25 |
| Malawi | DYAD−M | 253 | 264 | 0.01 (−0.01, 0.03) | 0.41 | 0.34 |
| Bangladesh | RDNS | 897 | 2551 | −0.02 (−0.05, 0.00) | 0.23 | 0.24 |
| Guatemala | WOMEN FIRST | 181 | 166 | −0.01 (−0.04, 0.03) | 0.13 | 0.17 |
| **I² = 0.32, Tau² = 0.00** | | **1638** | **3286** | | | |
| **Fixed** | | | | **−0.01 (−0.02, 0.01)** | | |
| **Random** | | | | **−0.01 (−0.02, 0.01)** | | |

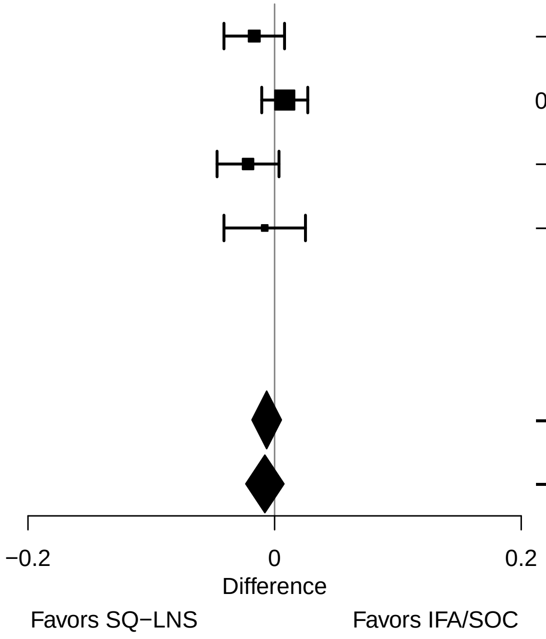

#### Supplemental figure 2AC: Mean difference in birth mid-upper arm circumference

| Country | Trial | SQ−LNS N | IFA/SOC N | MD (95% CI) | Fixed W | Random W |
|---|---|---|---|---|---|---|
| Ghana | DYAD−G | 307 | 305 | 0.16 (0.01, 0.31) | 0.16 | 0.16 |
| Malawi | DYAD−M | 256 | 264 | 0.11 (−0.05, 0.27) | 0.14 | 0.14 |
| Bangladesh | RDNS | 898 | 2551 | 0.06 (−0.02, 0.13) | 0.69 | 0.69 |
| Guatemala | WOMEN FIRST | | | | | |
| **I² = 0.00, Tau² = 0.00** | | **1461** | **3120** | | | |
| **Fixed** | | | | **0.08 (0.02, 0.14)** | | |
| **Random** | | | | **0.08 (0.02, 0.14)** | | |

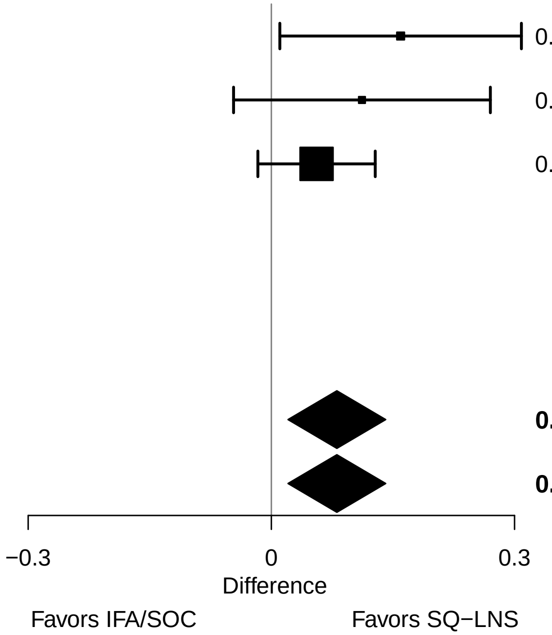

### Supplemental figure 2AD: Mean difference in duration of gestation

| Country | Trial | SQ−LNS N | IFA/SOC N | MD (95% CI) | Fixed W | Random W |
|---|---|---|---|---|---|---|
| Ghana | DYAD−G | 331 | 328 | 0.14 (−0.16, 0.44) | 0.15 | 0.15 |
| Malawi | DYAD−M | 436 | 437 | 0.19 (−0.21, 0.59) | 0.09 | 0.09 |
| Bangladesh | RDNS | 898 | 2551 | 0.13 (−0.02, 0.29) | 0.60 | 0.60 |
| Guatemala | WOMEN FIRST | 190 | 177 | 0.03 (−0.27, 0.33) | 0.16 | 0.16 |
| **I² = 0.00, Tau² = 0.00** | | **1855** | **3493** | | | |
| **Fixed** | | | | **0.12 (0.01, 0.24)** | | |
| **Random** | | | | **0.12 (0.01, 0.24)** | | |

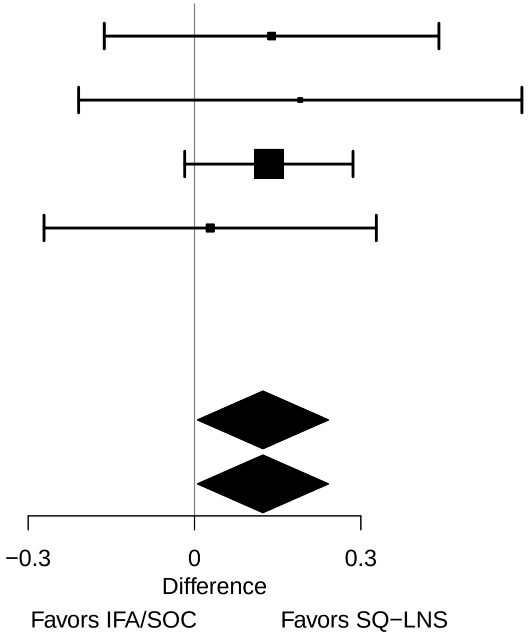

### Supplemental figure 2AE: Preterm birth relative risk

| Country | Trial | SQ−LNS N | IFA/SOC N | RR (95% CI) | Fixed W | Random W |
|---|---|---|---|---|---|---|
| Ghana | DYAD−G | 331 | 328 | 0.99 (0.62, 1.58) | 0.12 | 0.12 |
| Malawi | DYAD−M | 436 | 437 | 0.85 (0.58, 1.24) | 0.18 | 0.18 |
| Bangladesh | RDNS | 898 | 2551 | 0.95 (0.78, 1.16) | 0.65 | 0.65 |
| Guatemala | WOMEN FIRST | 190 | 177 | 1.00 (0.51, 1.94) | 0.06 | 0.06 |
| **I² = 0.00, Tau² = 0.00** | | **1855** | **3493** | | | |
| **Fixed** | | | | **0.94 (0.80, 1.10)** | | |
| **Random** | | | | **0.94 (0.80, 1.10)** | | |

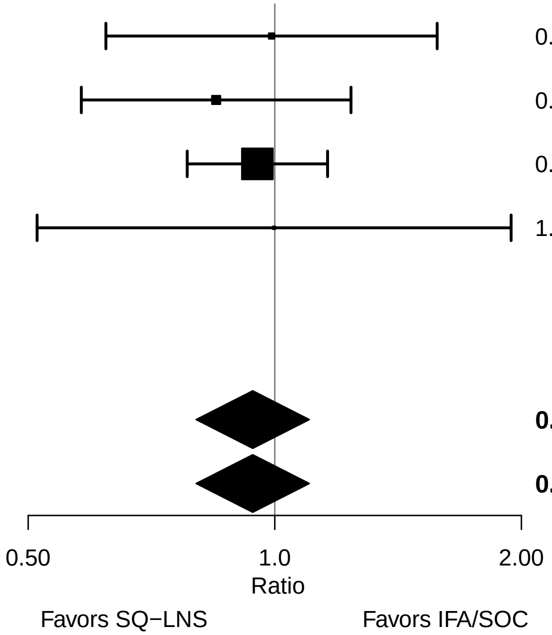

#### Supplemental figure 2AF: Preterm birth risk difference

| Country | Trial | SQ−LNS N | IFA/SOC N | RD (95% CI) | Fixed W | Random W |
|---|---|---|---|---|---|---|
| Ghana | DYAD−G | 331 | 328 | 0.00 (−0.05, 0.04) | 0.17 | 0.17 |
| Malawi | DYAD−M | 436 | 437 | −0.02 (−0.06, 0.02) | 0.20 | 0.20 |
| Bangladesh | RDNS | 898 | 2551 | −0.01 (−0.03, 0.02) | 0.50 | 0.50 |
| Guatemala | WOMEN FIRST | 190 | 177 | 0.00 (−0.05, 0.05) | 0.13 | 0.13 |
| **I² = 0.00, Tau² = 0.00** | | **1855** | **3493** | | | |
| **Fixed** | | | | **−0.01 (−0.03, 0.01)** | | |
| **Random** | | | | **−0.01 (−0.03, 0.01)** | | |

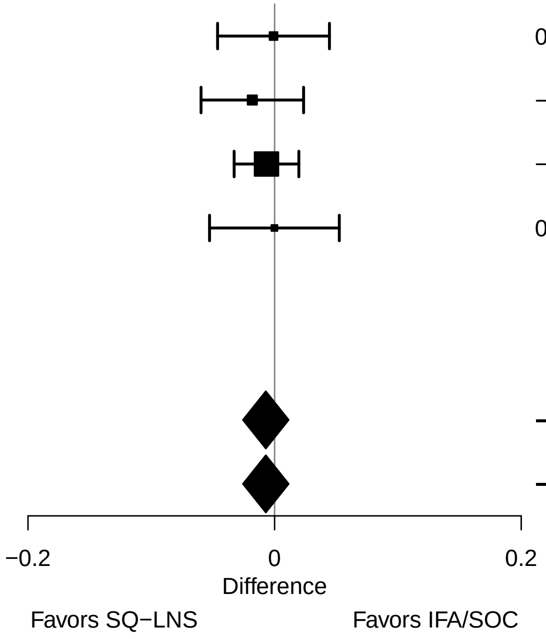

#### Supplemental figure 2AG: Mean difference in 6 mo weight-for-age z-score

| Country | Trial | SQ−LNS N | IFA/SOC N | MD (95% CI) | Fixed W | Random W |
|---|---|---|---|---|---|---|
| Ghana | DYAD−G | 281 | 272 | 0.07 (−0.11, 0.26) | 0.16 | 0.16 |
| Malawi | DYAD−M | 350 | 364 | 0.00 (−0.16, 0.17) | 0.21 | 0.21 |
| Bangladesh | RDNS | 866 | 2463 | 0.01 (−0.10, 0.13) | 0.43 | 0.43 |
| Guatemala | WOMEN FIRST | 219 | 214 | 0.17 (0.00, 0.33) | 0.21 | 0.21 |
| **I² = 0.00, Tau² = 0.00** | | **1716** | **3313** | | | |
| **Fixed** | | | | **0.05 (−0.02, 0.13)** | | |
| **Random** | | | | **0.05 (−0.02, 0.13)** | | |

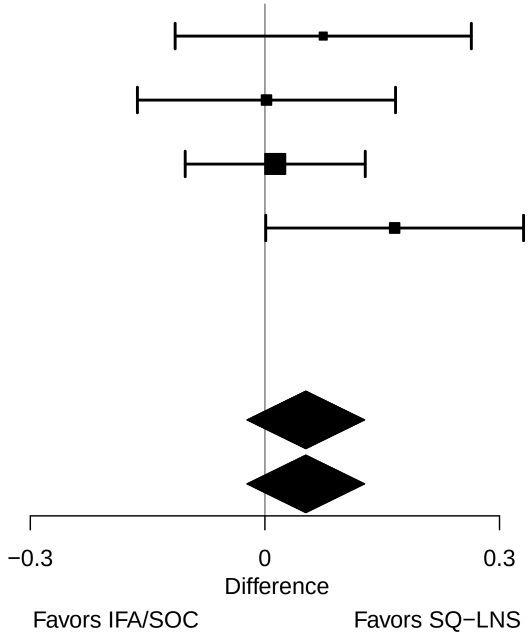

### Supplemental figure 2AH: 6 mo underweight prevalence ratio

| Country | Trial | SQ−LNS N | IFA/SOC N | PR (95% CI) | Fixed W | Random W |
|---|---|---|---|---|---|---|
| Ghana | DYAD−G | 281 | 272 | 0.85 (0.48, 1.48) | 0.07 | 0.11 |
| Malawi | DYAD−M | 350 | 364 | 1.04 (0.66, 1.65) | 0.11 | 0.15 |
| Bangladesh | RDNS | 866 | 2463 | 0.96 (0.77, 1.20) | 0.48 | 0.41 |
| Guatemala | WOMEN FIRST | 219 | 214 | 0.67 (0.52, 0.87) | 0.33 | 0.33 |
| **I² = 0.41, Tau² = 0.01** | | **1716** | **3313** | | | |
| **Fixed** | | | | **0.85 (0.73, 0.99)** | | |
| **Random** | | | | **0.85 (0.70, 1.04)** | | |

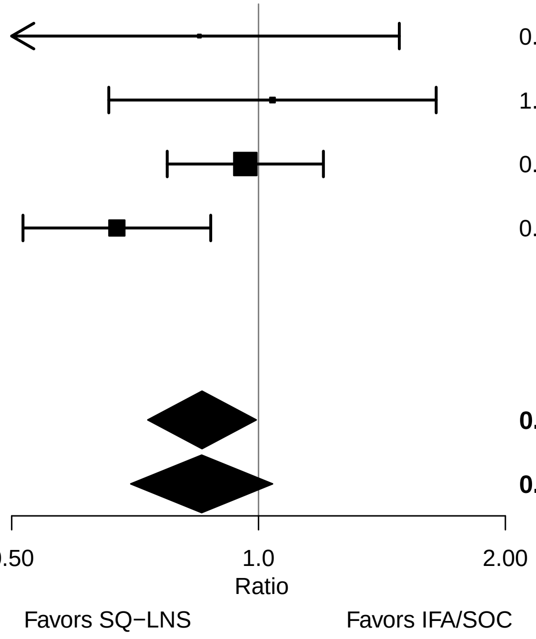

#### Supplemental figure 2AI: 6 mo underweight prevalence difference

| Country | Trial | SQ−LNS N | IFA/SOC N | RD (95% CI) | Fixed W | Random W |
|---|---|---|---|---|---|---|
| Ghana | DYAD−G | 281 | 272 | −0.01 (−0.06, 0.03) | 0.18 | 0.20 |
| Malawi | DYAD−M | 350 | 364 | 0.00 (−0.04, 0.05) | 0.21 | 0.23 |
| Bangladesh | RDNS | 866 | 2463 | −0.01 (−0.05, 0.03) | 0.22 | 0.23 |
| Guatemala | WOMEN FIRST | 219 | 214 | −0.05 (−0.08, −0.02) | 0.38 | 0.34 |
| **I² = 0.37, Tau² = 0.00** | | **1716** | **3313** | | | |
| **Fixed** | | | | **−0.02 (−0.04, 0.00)** | | |
| **Random** | | | | **−0.02 (−0.04, 0.00)** | | |

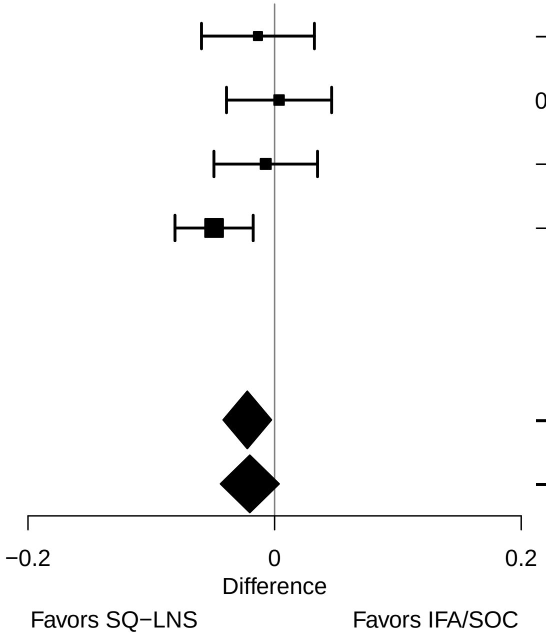

#### Supplemental figure 2AJ: Mean difference in 6 mo length-for-age z-score

| Country | Trial | SQ−LNS N | IFA/SOC N | MD (95% CI) | Fixed W | Random W |
|---|---|---|---|---|---|---|
| Ghana | DYAD−G | 282 | 274 | 0.08 (−0.09, 0.25) | 0.16 | 0.17 |
| Malawi | DYAD−M | 350 | 365 | 0.02 (−0.15, 0.18) | 0.17 | 0.18 |
| Bangladesh | RDNS | 866 | 2462 | 0.03 (−0.06, 0.13) | 0.53 | 0.52 |
| Guatemala | WOMEN FIRST | 219 | 214 | 0.20 (0.02, 0.39) | 0.14 | 0.14 |
| **I² = 0.02, Tau² = 0.00** | | **1717** | **3315** | | | |
| **Fixed** | | | | **0.06 (−0.01, 0.13)** | | |
| **Random** | | | | **0.06 (−0.01, 0.13)** | | |

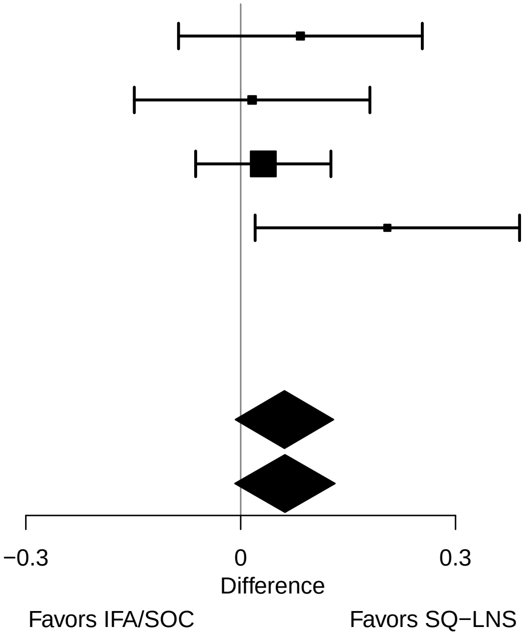

### Supplemental figure 2AK: 6 mo stunting prevalence ratio

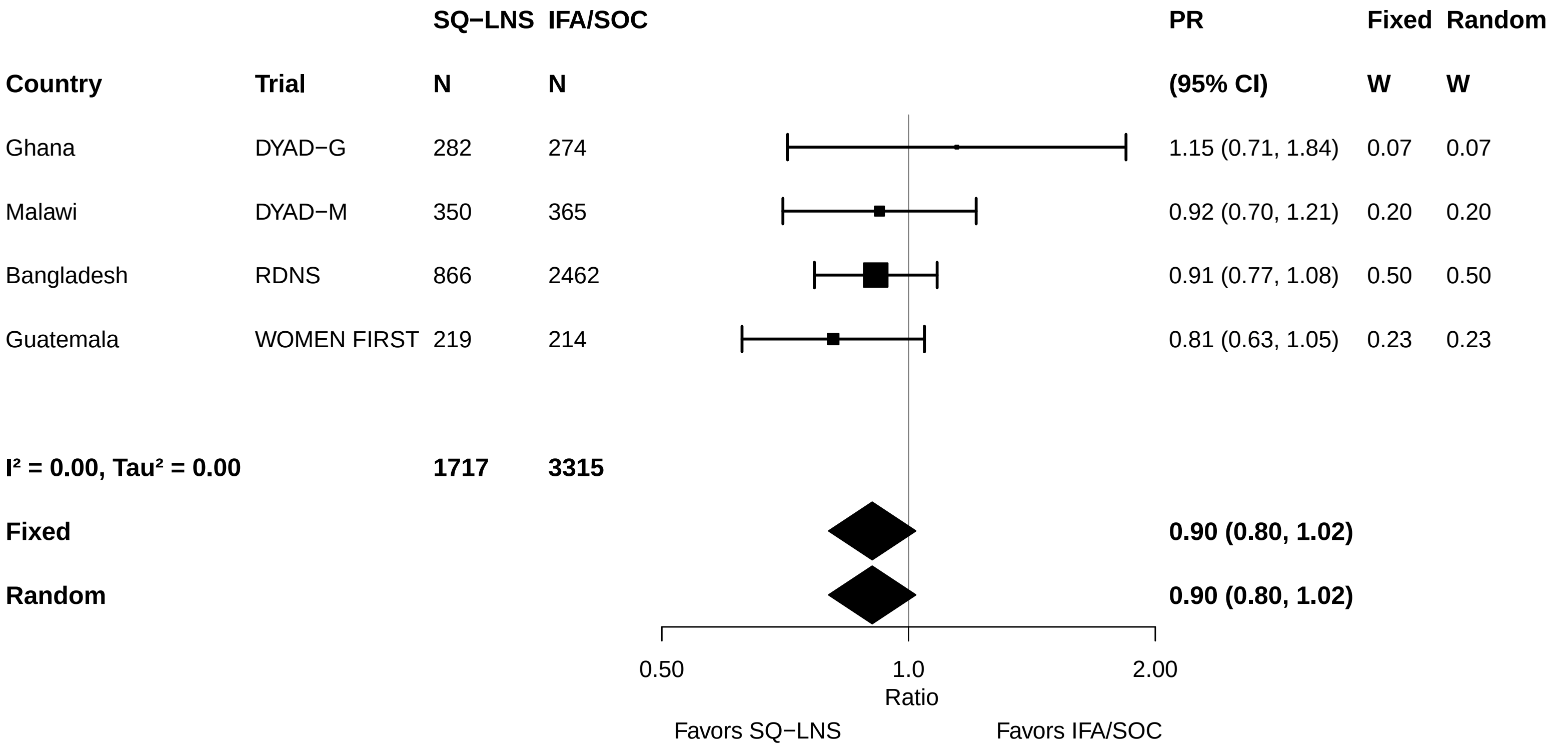

#### Supplemental figure 2AL: 6 mo stunting prevalence difference

| Country | Trial | SQ−LNS N | IFA/SOC N | RD (95% CI) | Fixed W | Random W |
|---|---|---|---|---|---|---|
| Ghana | DYAD−G | 282 | 274 | 0.01 (−0.04, 0.07) | 0.25 | 0.25 |
| Malawi | DYAD−M | 350 | 365 | −0.02 (−0.08, 0.04) | 0.18 | 0.18 |
| Bangladesh | RDNS | 866 | 2462 | −0.02 (−0.06, 0.02) | 0.43 | 0.43 |
| Guatemala | WOMEN FIRST | 219 | 214 | −0.06 (−0.12, 0.01) | 0.15 | 0.15 |
| **$I^2$ = 0.00, $Tau^2$ = 0.00** | | **1717** | **3315** | | | |
| **Fixed** | | | | **−0.02 (−0.04, 0.01)** | | |
| **Random** | | | | **−0.02 (−0.04, 0.01)** | | |

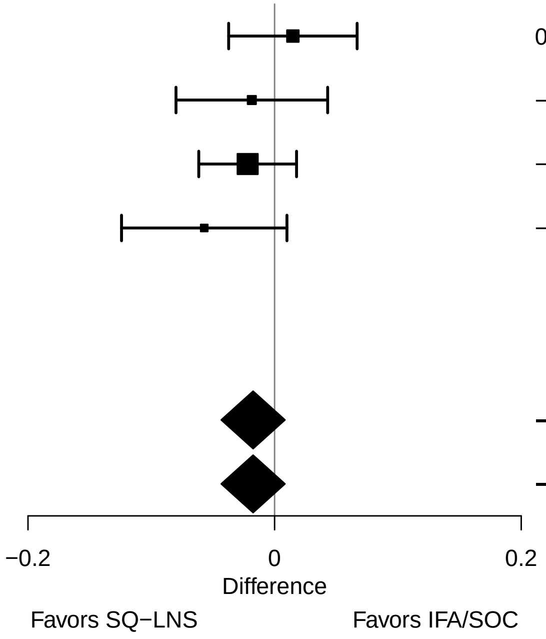

#### Supplemental figure 2AM: Mean difference in 6 mo weight-for-length z-score

| Country | Trial | SQ−LNS N | IFA/SOC N | MD (95% CI) | Fixed W | Random W |
|---|---|---|---|---|---|---|
| Ghana | DYAD−G | 281 | 272 | 0.03 (−0.16, 0.22) | 0.15 | 0.15 |
| Malawi | DYAD−M | 350 | 364 | −0.03 (−0.20, 0.14) | 0.18 | 0.18 |
| Bangladesh | RDNS | 866 | 2462 | −0.02 (−0.12, 0.09) | 0.48 | 0.48 |
| Guatemala | WOMEN FIRST | 219 | 214 | 0.05 (−0.12, 0.22) | 0.18 | 0.18 |
| **I² = 0.00, Tau² = 0.00** | | **1716** | **3312** | | | |
| **Fixed** | | | | **0.00 (−0.07, 0.07)** | | |
| **Random** | | | | **0.00 (−0.07, 0.07)** | | |

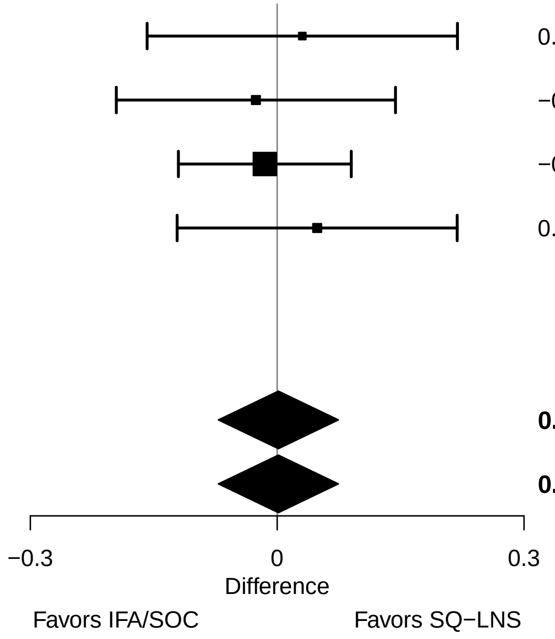

#### Supplemental figure 2AN: 6 mo wasting prevalence ratio

| Country | Trial | SQ−LNS N | IFA/SOC N | PR (95% CI) | Fixed W | Random W |
|---|---|---|---|---|---|---|
| Ghana | DYAD−G | 281 | 272 | 0.48 (0.20, 1.18) | 0.12 | 0.25 |
| Malawi | DYAD−M | 350 | 364 | 1.39 (0.49, 3.96) | 0.09 | 0.20 |
| Bangladesh | RDNS | 866 | 2462 | 1.01 (0.71, 1.46) | 0.75 | 0.46 |
| Guatemala | WOMEN FIRST | 219 | 214 | 0.24 (0.04, 1.47) | 0.03 | 0.09 |
| **$I^2$ = 0.39, $Tau^2$ = 0.17** | | **1716** | **3312** | | | |
| **Fixed** | | | | **0.91 (0.66, 1.25)** | | |
| **Random** | | | | **0.79 (0.43, 1.44)** | | |

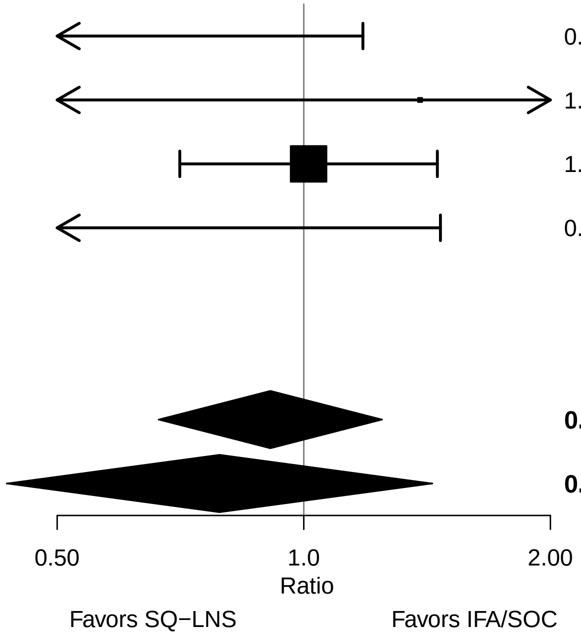

### Supplemental figure 2AO: 6 mo wasting prevalence difference

| Country | Trial | SQ−LNS N | IFA/SOC N | RD (95% CI) | Fixed W | Random W |
|---|---|---|---|---|---|---|
| Ghana | DYAD−G | 281 | 272 | −0.03 (−0.06, 0.01) | 0.09 | 0.13 |
| Malawi | DYAD−M | 350 | 364 | 0.01 (−0.01, 0.03) | 0.21 | 0.24 |
| Bangladesh | RDNS | 866 | 2462 | 0.00 (−0.02, 0.02) | 0.31 | 0.30 |
| Guatemala | WOMEN FIRST | 219 | 214 | −0.01 (−0.03, 0.00) | 0.39 | 0.34 |
| **I² = 0.36, Tau² = 0.00** | | **1716** | **3312** | | | |
| **Fixed** | | | | **−0.01 (−0.02, 0.00)** | | |
| **Random** | | | | **−0.01 (−0.02, 0.01)** | | |

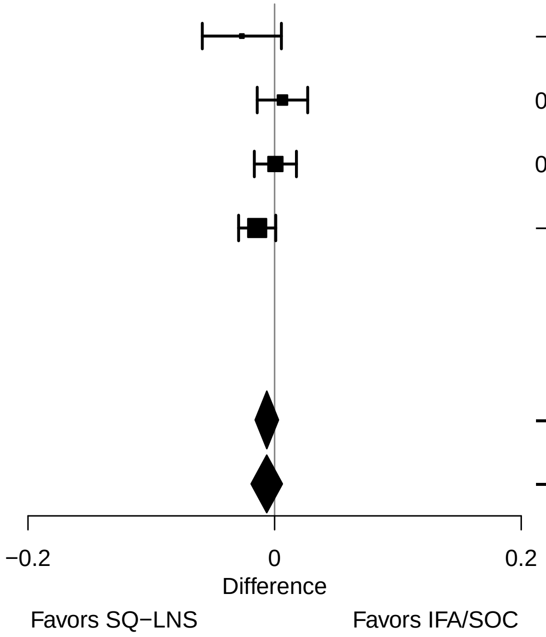

#### Supplemental figure 2AP: Mean difference in 6 mo head circumference-for-age z-score

| Country | Trial | SQ−LNS N | IFA/SOC N | MD (95% CI) | Fixed W | Random W |
|---|---|---|---|---|---|---|
| Ghana | DYAD−G | 282 | 274 | 0.07 (−0.09, 0.22) | 0.16 | 0.16 |
| Malawi | DYAD−M | 350 | 363 | −0.05 (−0.20, 0.10) | 0.16 | 0.16 |
| Bangladesh | RDNS | 866 | 2463 | 0.01 (−0.07, 0.09) | 0.57 | 0.57 |
| Guatemala | WOMEN FIRST | 219 | 211 | 0.06 (−0.12, 0.24) | 0.11 | 0.11 |
| **I² = 0.00, Tau² = 0.00** | | **1717** | **3311** | | | |
| **Fixed** | | | | **0.01 (−0.05, 0.07)** | | |
| **Random** | | | | **0.01 (−0.05, 0.07)** | | |

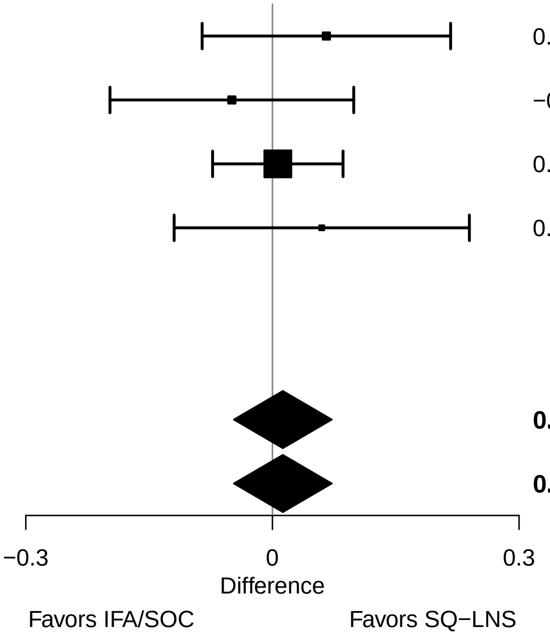

#### Supplemental figure 2AQ: 6 mo low HCZ prevalence ratio

| Country | Trial | SQ−LNS N | IFA/SOC N | PR (95% CI) | Fixed W | Random W |
|---|---|---|---|---|---|---|
| Ghana | DYAD−G | 282 | 274 | 0.80 (0.45, 1.44) | 0.04 | 0.04 |
| Malawi | DYAD−M | 350 | 363 | 1.15 (0.63, 2.09) | 0.04 | 0.04 |
| Bangladesh | RDNS | 866 | 2463 | 0.91 (0.80, 1.04) | 0.86 | 0.86 |
| Guatemala | WOMEN FIRST | 219 | 211 | 0.75 (0.45, 1.26) | 0.06 | 0.06 |
| **I² = 0.00, Tau² = 0.00** | | **1717** | **3311** | | | |
| **Fixed** | | | | **0.90 (0.80, 1.02)** | | |
| **Random** | | | | **0.90 (0.80, 1.02)** | | |

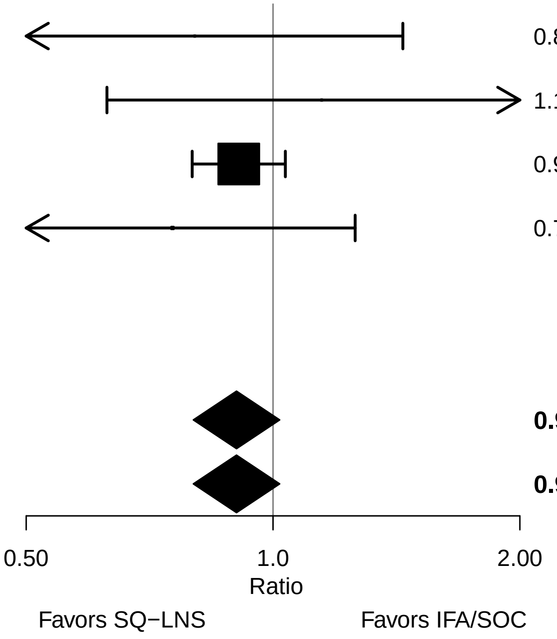

#### Supplemental figure 2AR: 6 mo low HCZ prevalence difference

| Country | Trial | SQ−LNS N | IFA/SOC N | RD (95% CI) | Fixed W | Random W |
|---|---|---|---|---|---|---|
| Ghana | DYAD−G | 282 | 274 | −0.02 (−0.06, 0.03) | 0.18 | 0.18 |
| Malawi | DYAD−M | 350 | 363 | 0.01 (−0.03, 0.04) | 0.30 | 0.30 |
| Bangladesh | RDNS | 866 | 2463 | −0.02 (−0.05, 0.01) | 0.36 | 0.36 |
| Guatemala | WOMEN FIRST | 219 | 211 | −0.03 (−0.07, 0.02) | 0.16 | 0.16 |
| **I² = 0.00, Tau² = 0.00** | | **1717** | **3311** | | | |
| **Fixed** | | | | **−0.01 (−0.03, 0.01)** | | |
| **Random** | | | | **−0.01 (−0.03, 0.01)** | | |

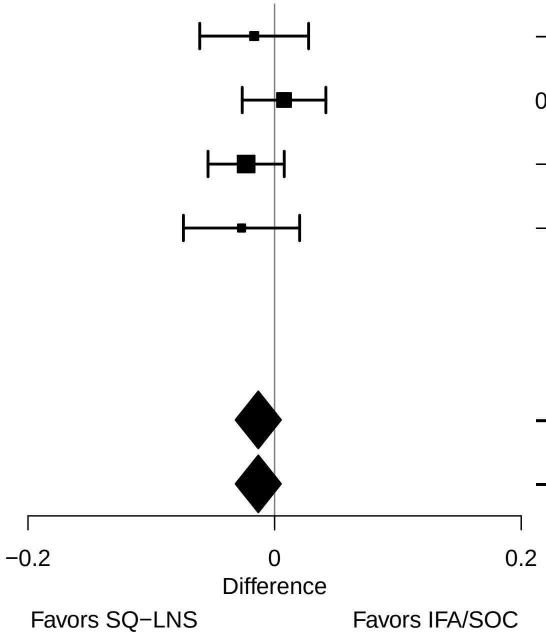

#### Supplemental figure 2AS: Mean difference in 6 mo MUAC-for-age z-score

| Country | Trial | SQ−LNS N | IFA/SOC N | MD (95% CI) | Fixed W | Random W |
|---|---|---|---|---|---|---|
| Ghana | DYAD−G | 282 | 274 | −0.01 (−0.18, 0.15) | 0.21 | 0.21 |
| Malawi | DYAD−M | 350 | 365 | 0.07 (−0.09, 0.23) | 0.23 | 0.23 |
| Bangladesh | RDNS | 866 | 2463 | −0.02 (−0.12, 0.08) | 0.55 | 0.55 |
| Guatemala | WOMEN FIRST | | | | | |
| **I² = 0.00, Tau² = 0.00** | | **1498** | **3102** | | | |
| **Fixed** | | | | **0.00 (−0.07, 0.08)** | | |
| **Random** | | | | **0.00 (−0.07, 0.08)** | | |

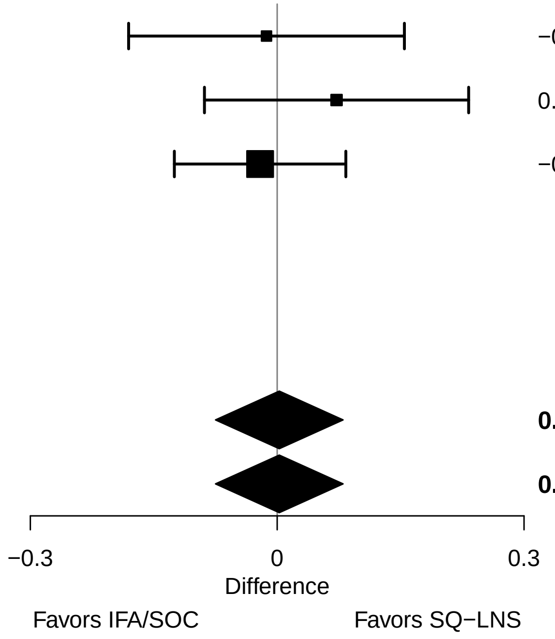

### Supplemental figure 2AT: 6 mo low MUAC prevalence ratio

| Country | Trial | SQ−LNS N | IFA/SOC N | PR (95% CI) | Fixed W | Random W |
|---|---|---|---|---|---|---|
| Ghana | DYAD−G | 282 | 274 | 0.97 (0.56, 1.69) | 0.21 | 0.21 |
| Malawi | DYAD−M | 350 | 365 | 0.74 (0.43, 1.30) | 0.21 | 0.21 |
| Bangladesh | RDNS | 866 | 2463 | 1.13 (0.81, 1.58) | 0.58 | 0.58 |
| Guatemala | WOMEN FIRST | | | | | |
| **I² = 0.00, Tau² = 0.00** | | **1498** | **3102** | | | |
| **Fixed** | | | | **1.00 (0.78, 1.29)** | | |
| **Random** | | | | **1.00 (0.78, 1.29)** | | |

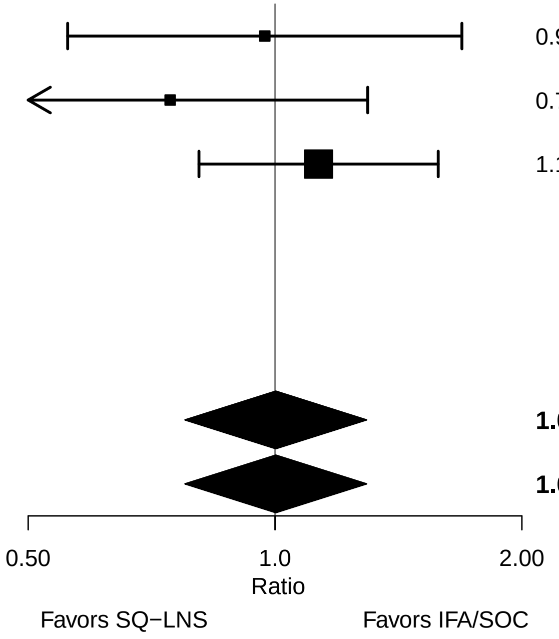

#### Supplemental figure 2AU: 6 mo low MUAC prevalence difference

| Country | Trial | SQ−LNS N | IFA/SOC N | RD (95% CI) | Fixed W | Random W |
|---|---|---|---|---|---|---|
| Ghana | DYAD−G | 282 | 274 | 0.00 (−0.05, 0.04) | 0.15 | 0.15 |
| Malawi | DYAD−M | 350 | 365 | −0.02 (−0.06, 0.02) | 0.24 | 0.24 |
| Bangladesh | RDNS | 866 | 2463 | 0.01 (−0.01, 0.03) | 0.61 | 0.61 |
| Guatemala | WOMEN FIRST | | | | | |
| **I² = 0.00, Tau² = 0.00** | | **1498** | **3102** | | | |
| **Fixed** | | | | **0.00 (−0.02, 0.02)** | | |
| **Random** | | | | **0.00 (−0.02, 0.02)** | | |

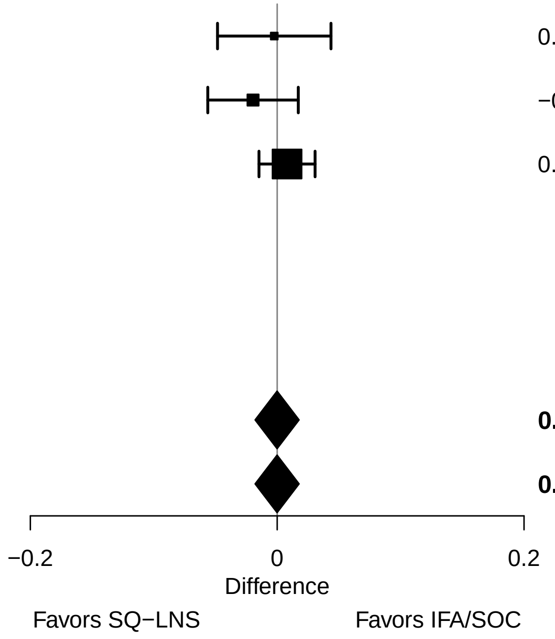

### Supplemental figure 2AV: 6 mo acute malnutrition prevalence ratio

| Country | Trial | SQ−LNS N | IFA/SOC N | PR (95% CI) | Fixed W | Random W |
|---|---|---|---|---|---|---|
| Ghana | DYAD−G | 281 | 272 | 1.01 (0.58, 1.75) | 0.17 | 0.17 |
| Malawi | DYAD−M | 350 | 364 | 0.83 (0.50, 1.39) | 0.19 | 0.19 |
| Bangladesh | RDNS | 866 | 2462 | 1.05 (0.79, 1.39) | 0.64 | 0.64 |
| Guatemala | WOMEN FIRST | | | | | |
| **I² = 0.00, Tau² = 0.00** | | **1497** | **3098** | | | |
| **Fixed** | | | | **1.00 (0.79, 1.25)** | | |
| **Random** | | | | **1.00 (0.79, 1.25)** | | |

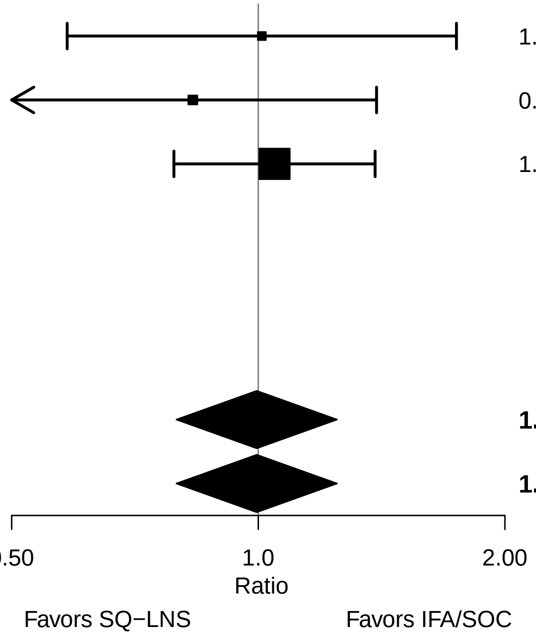

#### Supplemental figure 2AW: 6 mo acute malnutrition prevalence difference

| Country | Trial | SQ−LNS N | IFA/SOC N | RD (95% CI) | Fixed W | Random W |
|---|---|---|---|---|---|---|
| Ghana | DYAD−G | 281 | 272 | 0.00 (−0.05, 0.05) | 0.16 | 0.16 |
| Malawi | DYAD−M | 350 | 364 | −0.01 (−0.05, 0.02) | 0.23 | 0.23 |
| Bangladesh | RDNS | 866 | 2462 | 0.00 (−0.02, 0.03) | 0.61 | 0.61 |
| Guatemala | WOMEN FIRST | | | | | |
| **I² = 0.00, Tau² = 0.00** | | **1497** | **3098** | | | |
| **Fixed** | | | | **0.00 (−0.02, 0.02)** | | |
| **Random** | | | | **0.00 (−0.02, 0.02)** | | |

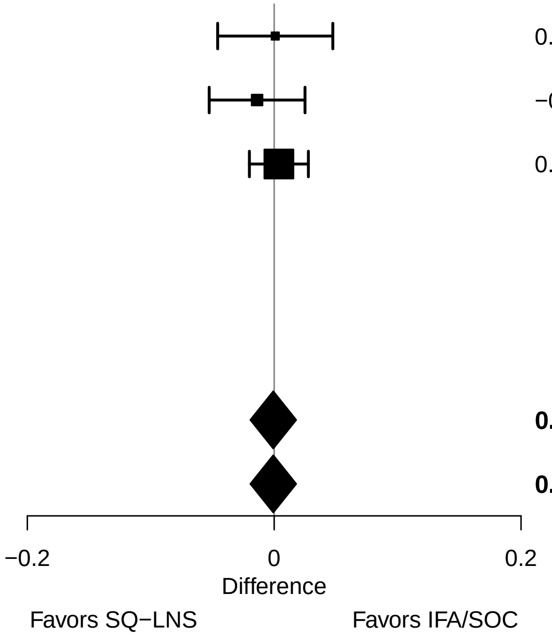

#### Supplemental figure 2AX: Cesarian-section relative risk

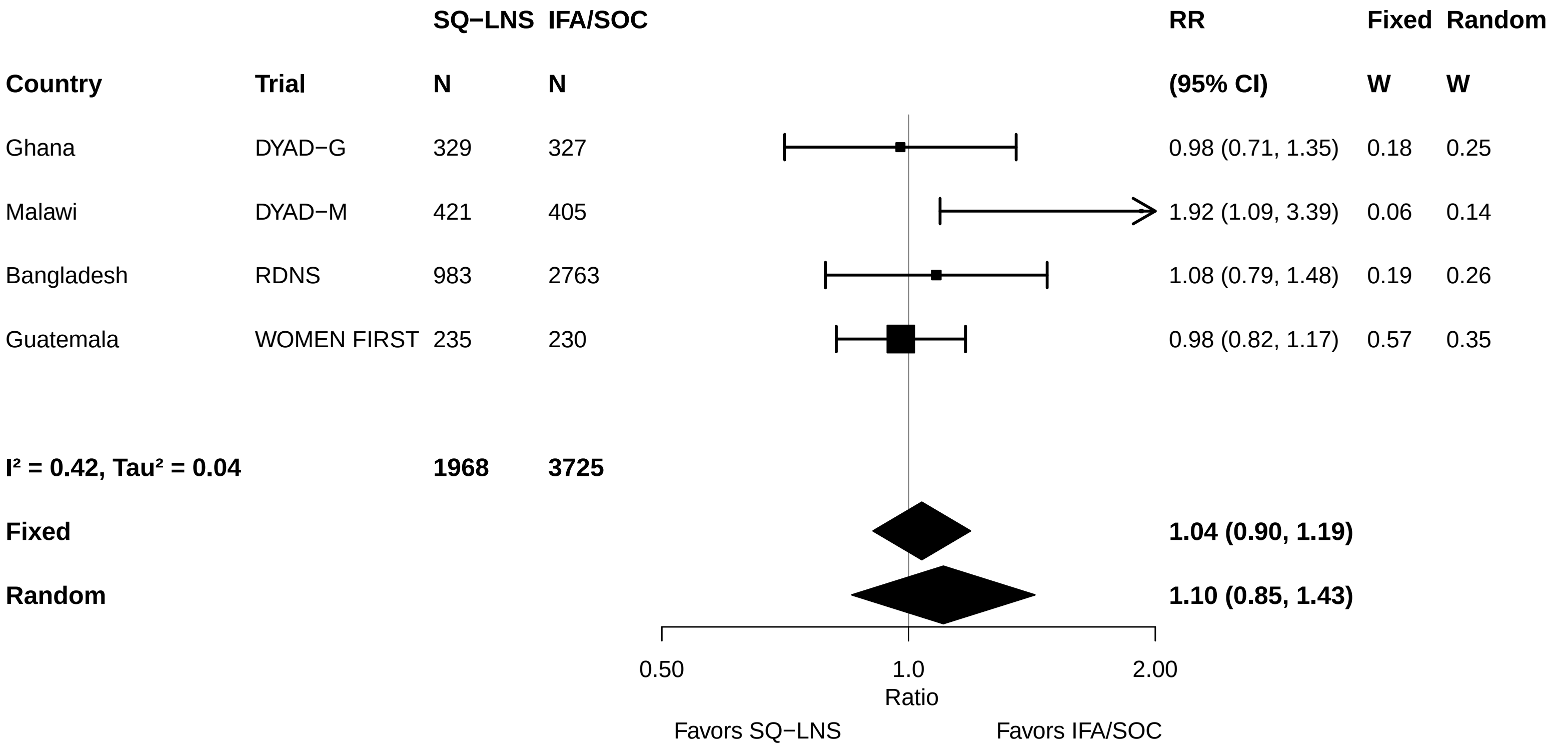

### Supplemental figure 2AY: Cesarian-section risk difference

| Country | Trial | SQ−LNS N | IFA/SOC N | RD (95% CI) | Fixed W | Random W |
|---|---|---|---|---|---|---|
| Ghana | DYAD−G | 329 | 327 | 0.00 (−0.06, 0.06) | 0.14 | 0.14 |
| Malawi | DYAD−M | 421 | 405 | 0.04 (0.01, 0.07) | 0.47 | 0.47 |
| Bangladesh | RDNS | 983 | 2763 | 0.01 (−0.04, 0.06) | 0.19 | 0.19 |
| Guatemala | WOMEN FIRST | 235 | 230 | −0.01 (−0.05, 0.04) | 0.21 | 0.21 |
| **I² = 0.01, Tau² = 0.00** | | **1968** | **3725** | | | |
| **Fixed** | | | | **0.02 (0.00, 0.04)** | | |
| **Random** | | | | **0.02 (0.00, 0.04)** | | |

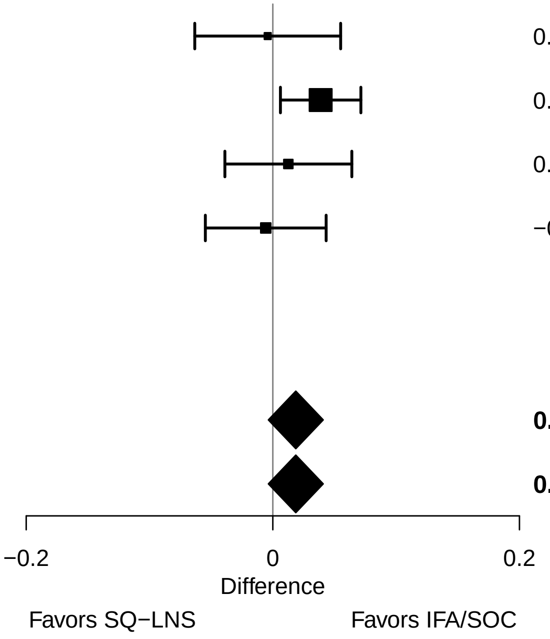

#### Supplemental figure 2AZ: Miscarriage relative risk

| Country | Trial | SQ−LNS N | IFA/SOC N | RR (95% CI) | Fixed W | Random W |
|---|---|---|---|---|---|---|
| Ghana | DYAD−G | 354 | 349 | 0.68 (0.30, 1.58) | 0.07 | 0.07 |
| Malawi | DYAD−M | 431 | 433 | 4.02 (0.45, 35.81) | 0.01 | 0.01 |
| Bangladesh | RDNS | 1044 | 2954 | 0.89 (0.69, 1.15) | 0.72 | 0.72 |
| Guatemala | WOMEN FIRST | 267 | 270 | 0.81 (0.50, 1.31) | 0.20 | 0.20 |
| **$I^2 = 0.00$, $Tau^2 = 0.00$** | | **2096** | **4006** | | | |
| **Fixed** | | | | **0.87 (0.70, 1.08)** | | |
| **Random** | | | | **0.87 (0.70, 1.08)** | | |

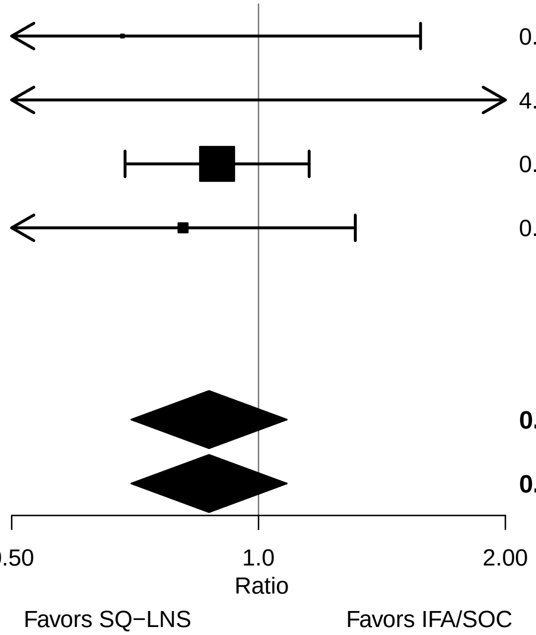

#### Supplemental figure 2BA: Miscarriage risk difference

| Country | Trial | SQ−LNS N | IFA/SOC N | RD (95% CI) | Fixed W | Random W |
|---|---|---|---|---|---|---|
| Ghana | DYAD−G | 354 | 349 | −0.01 (−0.04, 0.01) | 0.09 | 0.09 |
| Malawi | DYAD−M | 431 | 433 | 0.01 (0.00, 0.02) | 0.60 | 0.60 |
| Bangladesh | RDNS | 1044 | 2954 | −0.01 (−0.02, 0.01) | 0.29 | 0.29 |
| Guatemala | WOMEN FIRST | 267 | 270 | −0.03 (−0.09, 0.04) | 0.01 | 0.01 |
| **I² = 0.27, Tau² = 0.00** | | **2096** | **4006** | | | |
| **Fixed** | | | | **0.00 (−0.01, 0.01)** | | |
| **Random** | | | | **0.00 (−0.01, 0.01)** | | |

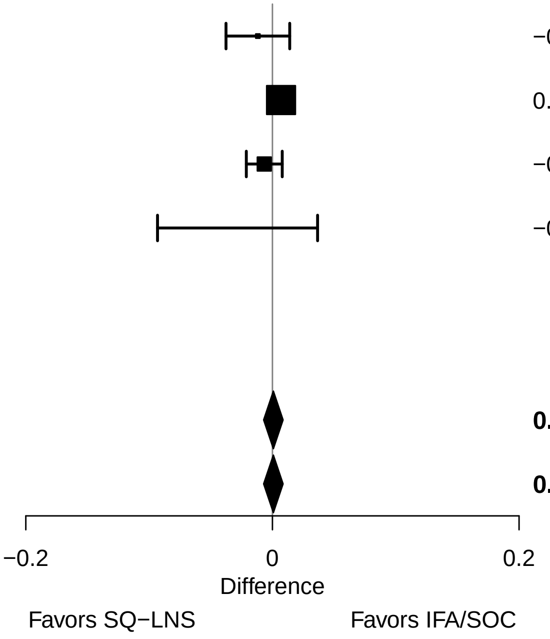

### Supplemental figure 2BB: Stillbirth relative risk

| Country | Trial | SQ−LNS N | IFA/SOC N | RR (95% CI) | Fixed W | Random W |
|---|---|---|---|---|---|---|
| Ghana | DYAD−G | 354 | 349 | 0.44 (0.14, 1.41) | 0.10 | 0.23 |
| Malawi | DYAD−M | 431 | 433 | 1.56 (0.68, 3.57) | 0.20 | 0.29 |
| Bangladesh | RDNS | 1044 | 2954 | 1.35 (0.85, 2.15) | 0.66 | 0.35 |
| Guatemala | WOMEN FIRST | 267 | 270 | 6.07 (0.86, 42.74) | 0.04 | 0.13 |
| **I² = 0.49, Tau² = 0.51** | | **2096** | **4006** | | | |
| **Fixed** | | | | **1.31 (0.90, 1.91)** | | |
| **Random** | | | | **[illegible] ([illegible], 3.18)** | | |

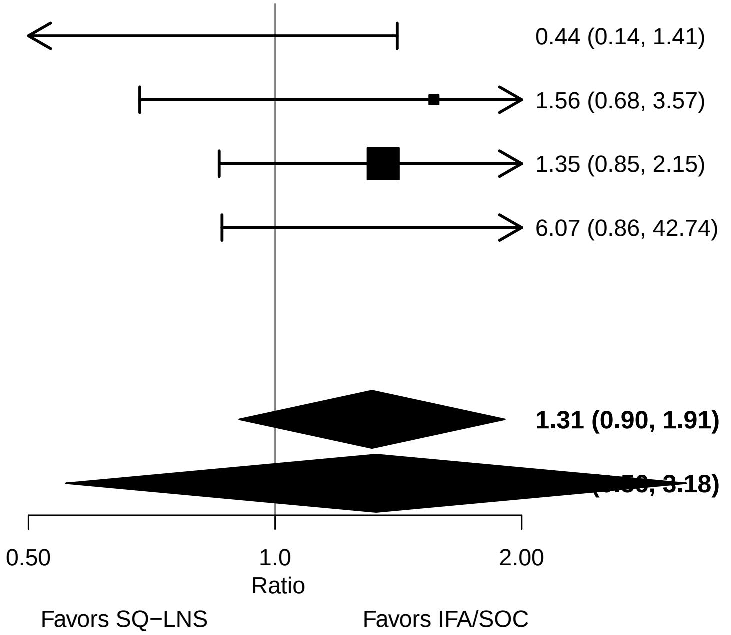

### Supplemental figure 2BC: Stillbirth risk difference

| Country | Trial | SQ−LNS N | IFA/SOC N | RD (95% CI) | Fixed W | Random W |
|---|---|---|---|---|---|---|
| Ghana | DYAD−G | 354 | 349 | −0.01 (−0.03, 0.01) | 0.19 | 0.23 |
| Malawi | DYAD−M | 431 | 433 | 0.01 (−0.01, 0.03) | 0.17 | 0.21 |
| Bangladesh | RDNS | 1044 | 2954 | 0.01 (−0.01, 0.02) | 0.39 | 0.31 |
| Guatemala | WOMEN FIRST | 267 | 270 | 0.02 (0.00, 0.04) | 0.24 | 0.25 |
| **I² = 0.53, Tau² = 0.00** | | **2096** | **4006** | | | |
| **Fixed** | | | | **0.01 (0.00, 0.02)** | | |
| **Random** | | | | **0.01 (−0.01, 0.02)** | | |

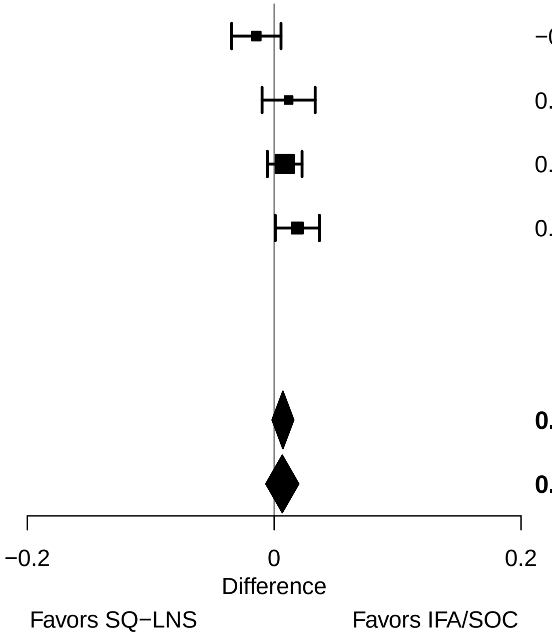

### Supplemental figure 2BD: Miscarriage or stillbirth relative risk

| Country | Trial | SQ−LNS N | IFA/SOC N | RR (95% CI) | Fixed W | Random W |
|---|---|---|---|---|---|---|
| Ghana | DYAD−G | 354 | 349 | 0.58 (0.30, 1.14) | 0.08 | 0.18 |
| Malawi | DYAD−M | 431 | 433 | 1.81 (0.84, 3.87) | 0.06 | 0.15 |
| Bangladesh | RDNS | 1044 | 2954 | 1.01 (0.79, 1.29) | 0.61 | 0.37 |
| Guatemala | WOMEN FIRST | 267 | 270 | 0.94 (0.64, 1.38) | 0.25 | 0.30 |
| **$I^2 = 0.39$, $Tau^2 = 0.08$** | | **2096** | **4006** | | | |
| **Fixed** | | | | **0.98 (0.81, 1.19)** | | |
| **Random** | | | | **0.98 (0.68, 1.42)** | | |

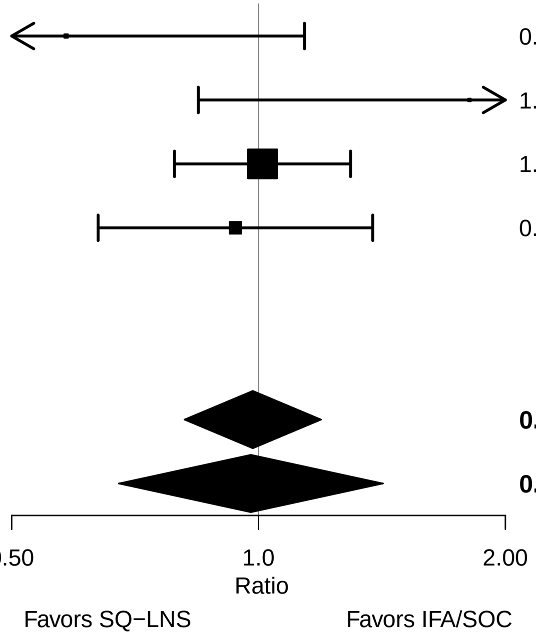

### Supplemental figure 2BE: Miscarriage or stillbirth risk difference

| Country | Trial | SQ−LNS N | IFA/SOC N | RD (95% CI) | Fixed W | Random W |
|---|---|---|---|---|---|---|
| Ghana | DYAD−G | 354 | 349 | −0.03 (−0.06, 0.01) | 0.18 | 0.22 |
| Malawi | DYAD−M | 431 | 433 | 0.02 (0.00, 0.04) | 0.34 | 0.32 |
| Bangladesh | RDNS | 1044 | 2954 | 0.00 (−0.02, 0.02) | 0.43 | 0.36 |
| Guatemala | WOMEN FIRST | 267 | 270 | −0.01 (−0.07, 0.05) | 0.06 | 0.09 |
| **I² = 0.40, Tau² = 0.00** | | **2096** | **4006** | | | |
| **Fixed** | | | | **0.00 (−0.01, 0.02)** | | |
| **Random** | | | | **0.00 (−0.02, 0.02)** | | |

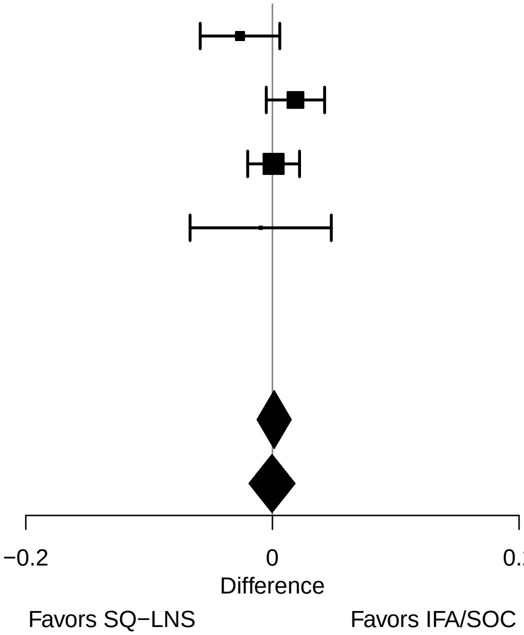

#### Supplemental figure 2BF: Early neonatal mortality relative risk

| Country | Trial | SQ−LNS N | IFA/SOC N | RR (95% CI) | Fixed W | Random W |
|---|---|---|---|---|---|---|
| Ghana | DYAD−G | 332 | 324 | 1.95 (0.49, 7.74) | 0.12 | 0.13 |
| Malawi | DYAD−M | 399 | 412 | 0.52 (0.22, 1.19) | 0.32 | 0.33 |
| Bangladesh | RDNS | 935 | 2651 | 0.79 (0.40, 1.52) | 0.51 | 0.50 |
| Guatemala | WOMEN FIRST | 228 | 225 | 0.33 (0.03, 3.31) | 0.04 | 0.05 |
| **I² = 0.04, Tau² = 0.02** | | **1894** | **3612** | | | |
| **Fixed** | | | | **0.74 (0.46, 1.19)** | | |
| **Random** | | | | **0.74 (0.45, 1.22)** | | |

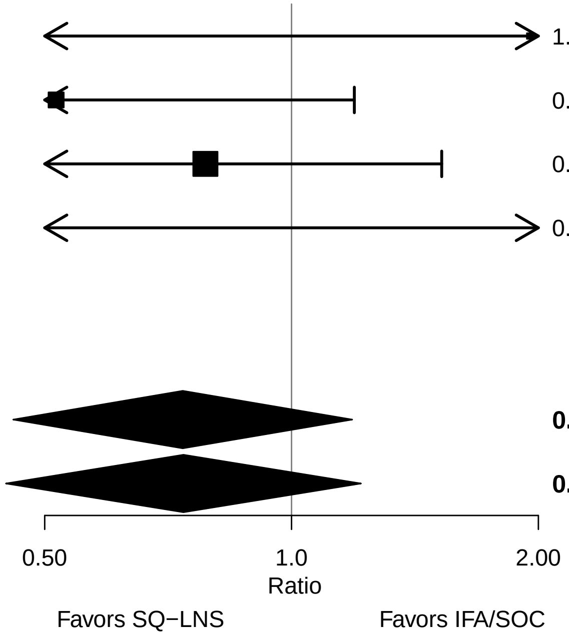

### Supplemental figure 2BG: Early neonatal mortality risk difference

| Country | Trial | SQ−LNS N | IFA/SOC N | RD (95% CI) | Fixed W | Random W |
|---|---|---|---|---|---|---|
| Ghana | DYAD−G | 332 | 324 | 0.01 (−0.01, 0.03) | 0.23 | 0.25 |
| Malawi | DYAD−M | 399 | 412 | −0.02 (−0.04, 0.00) | 0.13 | 0.18 |
| Bangladesh | RDNS | 935 | 2651 | −0.01 (−0.02, 0.01) | 0.40 | 0.33 |
| Guatemala | WOMEN FIRST | 228 | 225 | −0.01 (−0.03, 0.01) | 0.24 | 0.25 |
| **I² = 0.22, Tau² = 0.00** | | **1894** | **3612** | | | |
| **Fixed** | | | | **0.00 (−0.01, 0.00)** | | |
| **Random** | | | | **−0.01 (−0.02, 0.01)** | | |

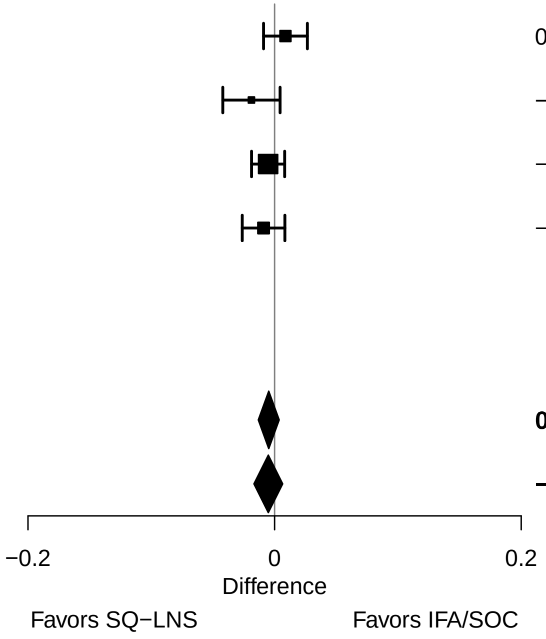

#### Supplemental figure 2BH: Miscarriage or stillbirth or early neonatal mortality relative risk

| Country | Trial | SQ−LNS N | IFA/SOC N | RR (95% CI) | Fixed W | Random W |
|---|---|---|---|---|---|---|
| Ghana | DYAD−G | 354 | 349 | 0.75 (0.42, 1.34) | 0.10 | 0.10 |
| Malawi | DYAD−M | 431 | 433 | 1.00 (0.59, 1.70) | 0.13 | 0.13 |
| Bangladesh | RDNS | 1044 | 2954 | 0.97 (0.75, 1.24) | 0.56 | 0.56 |
| Guatemala | WOMEN FIRST | 267 | 270 | 0.90 (0.60, 1.34) | 0.22 | 0.22 |
| **I² = 0.00, Tau² = 0.00** | | **2096** | **4006** | | | |
| **Fixed** | | | | **0.93 (0.77, 1.12)** | | |
| **Random** | | | | **0.93 (0.77, 1.12)** | | |

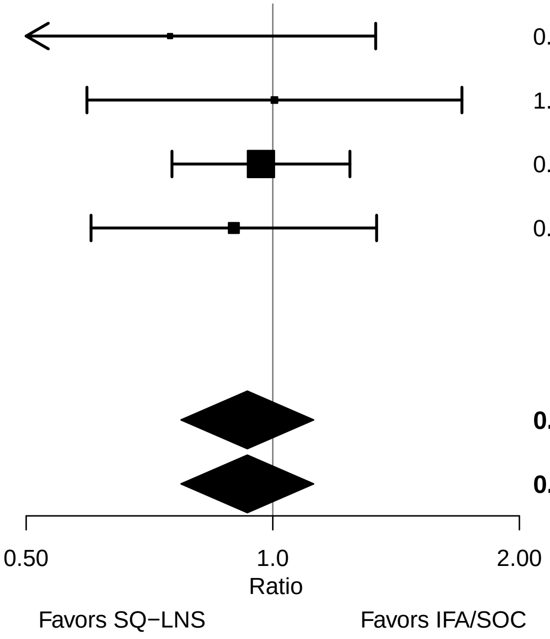

#### Supplemental figure 2BI: Miscarriage or stillbirth or early neonatal mortality risk difference

| Country | Trial | SQ−LNS N | IFA/SOC N | RD (95% CI) | Fixed W | Random W |
|---|---|---|---|---|---|---|
| Ghana | DYAD−G | 354 | 349 | −0.02 (−0.05, 0.02) | 0.22 | 0.22 |
| Malawi | DYAD−M | 431 | 433 | 0.00 (−0.03, 0.03) | 0.28 | 0.28 |
| Bangladesh | RDNS | 1044 | 2954 | 0.00 (−0.03, 0.02) | 0.43 | 0.43 |
| Guatemala | WOMEN FIRST | 267 | 270 | −0.02 (−0.08, 0.05) | 0.07 | 0.07 |
| **I² = 0.00, Tau² = 0.00** | | **2096** | **4006** | | | |
| **Fixed** | | | | **−0.01 (−0.02, 0.01)** | | |
| **Random** | | | | **−0.01 (−0.02, 0.01)** | | |

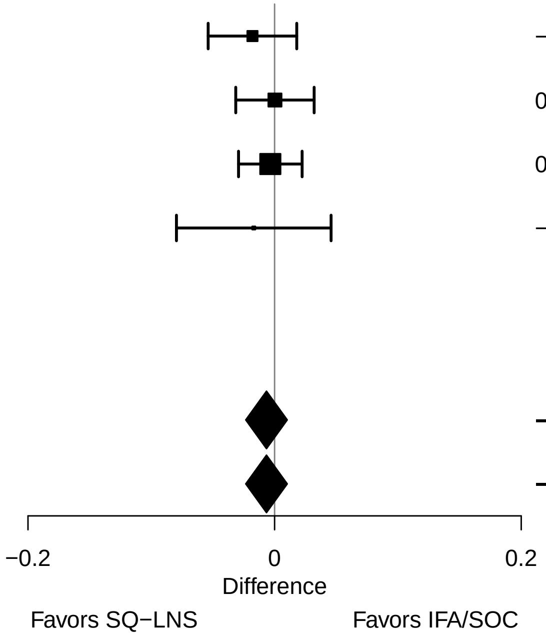

#### Supplemental figure 2BJ: Neonatal mortality relative risk

| Country | Trial | SQ−LNS N | IFA/SOC N | RR (95% CI) | Fixed W | Random W |
|---|---|---|---|---|---|---|
| Ghana | DYAD−G | 332 | 324 | 1.71 (0.50, 5.78) | 0.11 | 0.11 |
| Malawi | DYAD−M | 401 | 414 | 0.54 (0.26, 1.15) | 0.28 | 0.28 |
| Bangladesh | RDNS | 942 | 2663 | 0.92 (0.53, 1.59) | 0.52 | 0.52 |
| Guatemala | WOMEN FIRST | 227 | 225 | 0.74 (0.21, 2.69) | 0.10 | 0.10 |
| **$I^2$ = 0.00, $Tau^2$ = 0.00** | | **1902** | **3626** | | | |
| **Fixed** | | | | **0.83 (0.56, 1.24)** | | |
| **Random** | | | | **0.83 (0.56, 1.24)** | | |

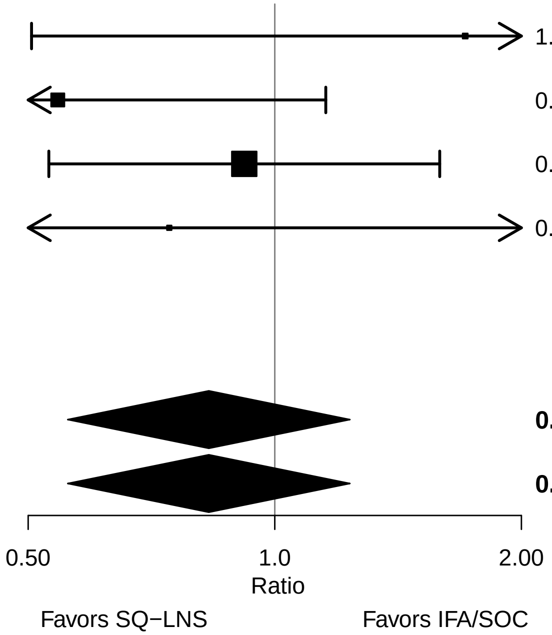

### Supplemental figure 2BK: Neonatal mortality risk difference

| Country | Trial | SQ−LNS N | IFA/SOC N | RD (95% CI) | Fixed W | Random W |
|---|---|---|---|---|---|---|
| Ghana | DYAD−G | 332 | 324 | 0.01 (−0.01, 0.03) | 0.23 | 0.23 |
| Malawi | DYAD−M | 401 | 414 | −0.02 (−0.05, 0.00) | 0.14 | 0.14 |
| Bangladesh | RDNS | 942 | 2663 | 0.00 (−0.02, 0.01) | 0.41 | 0.41 |
| Guatemala | WOMEN FIRST | 227 | 225 | 0.00 (−0.02, 0.02) | 0.22 | 0.22 |
| **$I^2$ = 0.10, $Tau^2$ = 0.00** | | **1902** | **3626** | | | |
| **Fixed** | | | | **0.00 (−0.01, 0.01)** | | |
| **Random** | | | | **0.00 (−0.01, 0.01)** | | |

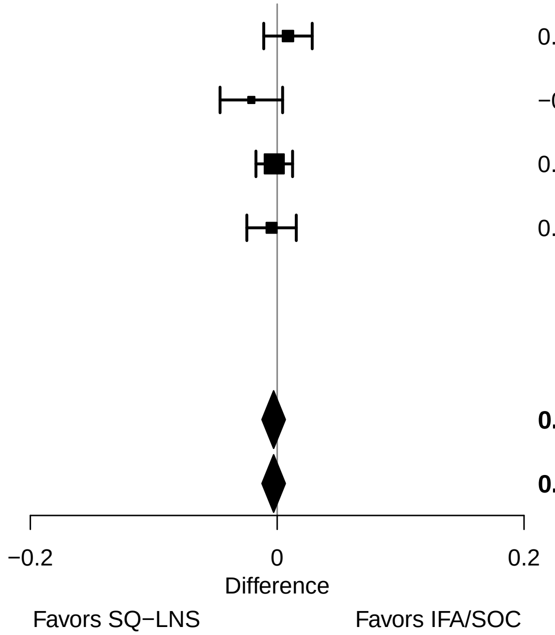

#### Supplemental figure 2BL: Mortality 0-6 mo relative risk

| Country | Trial | SQ−LNS N | IFA/SOC N | RR (95% CI) | Fixed W | Random W |
|---|---|---|---|---|---|---|
| Ghana | DYAD−G | 332 | 324 | 1.56 (0.52, 4.72) | 0.08 | 0.08 |
| Malawi | DYAD−M | 406 | 416 | 0.73 (0.38, 1.40) | 0.23 | 0.23 |
| Bangladesh | RDNS | 954 | 2689 | 1.01 (0.67, 1.53) | 0.58 | 0.58 |
| Guatemala | WOMEN FIRST | 225 | 220 | 0.81 (0.32, 2.05) | 0.11 | 0.11 |
| **$I^2$ = 0.00, $Tau^2$ = 0.00** | | **1917** | **3649** | | | |
| **Fixed** | | | | **0.95 (0.69, 1.30)** | | |
| **Random** | | | | **0.95 (0.69, 1.30)** | | |

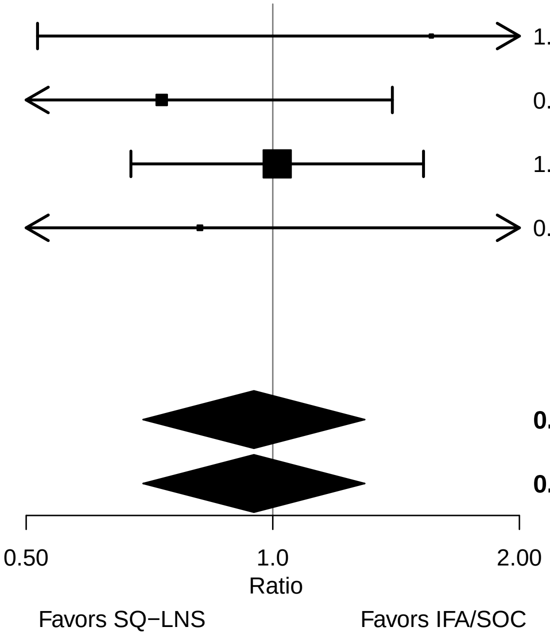

#### Supplemental figure 2BM: Mortality 0-6 mo risk difference

| Country | Trial | SQ−LNS N | IFA/SOC N | RD (95% CI) | Fixed W | Random W |
|---|---|---|---|---|---|---|
| Ghana | DYAD−G | 332 | 324 | 0.01 (−0.01, 0.03) | 0.24 | 0.24 |
| Malawi | DYAD−M | 406 | 416 | −0.01 (−0.04, 0.01) | 0.14 | 0.14 |
| Bangladesh | RDNS | 954 | 2689 | 0.00 (−0.02, 0.02) | 0.43 | 0.43 |
| Guatemala | WOMEN FIRST | 225 | 220 | −0.01 (−0.03, 0.02) | 0.19 | 0.19 |
| **I² = 0.00, Tau² = 0.00** | | **1917** | **3649** | | | |
| **Fixed** | | | | **0.00 (−0.01, 0.01)** | | |
| **Random** | | | | **0.00 (−0.01, 0.01)** | | |

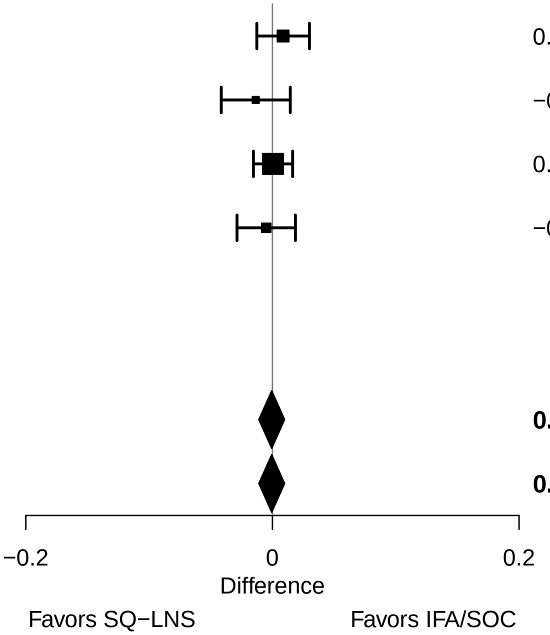

### Supplemental figure 3: Pooled plots for infant outcomes at birth and at 6 mo, stratified by potential effect modifiers, SQ-LNS vs IFA/SOC

These figures show pooled effects of SQ-LNS within study-level and individual-level characteristic subgroups along with the p-for-interaction. For definitions of effect modifiers, see Box 1 in the main paper. Individual study estimates were generated from log-binomial regression for dichotomous outcomes and simple linear regression for continuous outcomes with clustered observations using robust standard errors for cluster-randomized trials. Pooled sub-group estimates and statistical testing of the pooled interaction term were generated using inverse-variance weighting. For continuous outcomes the intervention effect is measured by the difference in mean of the SQ-LNS group minus IFA/SOC. For dichotomous outcomes analyzed via prevalence/risk ratios, the effect estimate is the prevalence/risk in the SQ-LNS group divided by the prevalence/risk in the IFA/SOC group. For dichotomous outcomes analyzed via prevalence/risk differences, the effect estimate is the prevalence/risk in the SQ-LNS group minus the prevalence/risk in the IFA/SOC group. The labels on the left y-axis correspond to the characteristic subgroups and their sample sizes. The values on the right indicate the pooled prevalence ratio and confidence interval within that subgroup.

LAZ, length-for-age z-score; WLZ, weight-for-length z-score; WAZ, weight for-age z-score; MUACZ, mid-upper arm circumference z-score; BMI, body mass index; HCZ, head circumference-for-age z-score; LGAZ, length-for-gestational-age z-score; HCGAZ, head circumference-for-gestational-age z-score; BMIZ, body mass index-for-age z-score; IFA/SOC, Iron and folic acid or standard of care; MD, mean difference; MMS, multiple micronutrient supplement; MUAC, mid-upper arm circumference; PR, prevalence ratio; PD, prevalence difference; RD, risk difference; RR, relative risk; SOC, standard of care; SQ-LNS, small-quantity lipid-based nutrient supplements; WGAZ, weight-for-gestational age z-score.

### Supplemental figure 3A: Sex

#### 3A1: Mean differences for birth outcomes

| Outcome (p−for−interaction) | SQ−LNS N | IFA/SOC N | Mean difference (95% CI) |
|---|---|---|---|
| Weight (kg) (p=0.017) | | | |
| Male | 901 | 1736 | 0.03 (−0.01, 0.06) |
| Female | 914 | 1722 | 0.08 (0.05, 0.11) |
| Weight−for−age z score (p=0.014) | | | |
| Male | 901 | 1736 | 0.06 (−0.02, 0.14) |
| Female | 914 | 1722 | 0.19 (0.13, 0.26) |
| Weight−for−gestational age z−score (p=0.765) | | | |
| Male | 425 | 423 | 0.13 (0.02, 0.25) |
| Female | 441 | 428 | 0.11 (0.01, 0.21) |
| Birth length (cm) (p=0.133) | | | |
| Male | 831 | 1677 | 0.12 (−0.05, 0.29) |
| Female | 849 | 1657 | 0.30 (0.16, 0.45) |
| Length−for−age z−score (p=0.139) | | | |
| Male | 831 | 1677 | 0.07 (−0.02, 0.16) |
| Female | 849 | 1657 | 0.16 (0.09, 0.24) |
| Length−for−gestational age z−score (p=0.683) | | | |
| Male | 355 | 365 | 0.14 (0.00, 0.27) |
| Female | 377 | 363 | 0.09 (−0.02, 0.21) |
| BMI z−score (p=0.025) | | | |
| Male | 829 | 1670 | 0.03 (−0.06, 0.12) |
| Female | 847 | 1656 | 0.17 (0.09, 0.25) |
| Head circumference (cm) (p=0.101) | | | |
| Male | 832 | 1678 | 0.06 (−0.05, 0.18) |
| Female | 848 | 1658 | 0.18 (0.09, 0.28) |
| Head circumference−for−age z−score (p=0.073) | | | |
| Male | 832 | 1678 | 0.05 (−0.04, 0.14) |
| Female | 848 | 1658 | 0.16 (0.08, 0.24) |
| Head circumference−for−gestational age z score (p=0.391) | | | |
| Male | 356 | 365 | 0.07 (−0.05, 0.20) |
| Female | 376 | 364 | 0.14 (0.02, 0.26) |
| Mid−upper arm circumference (cm) (p=0.193) | | | |
| Male | 730 | 1562 | 0.04 (−0.04, 0.13) |
| Female | 731 | 1558 | 0.12 (0.04, 0.19) |
| Duration (wk) (p=0.003) | | | |
| Male | 918 | 1748 | −0.06 (−0.24, 0.13) |
| Female | 922 | 1726 | 0.30 (0.15, 0.45) |

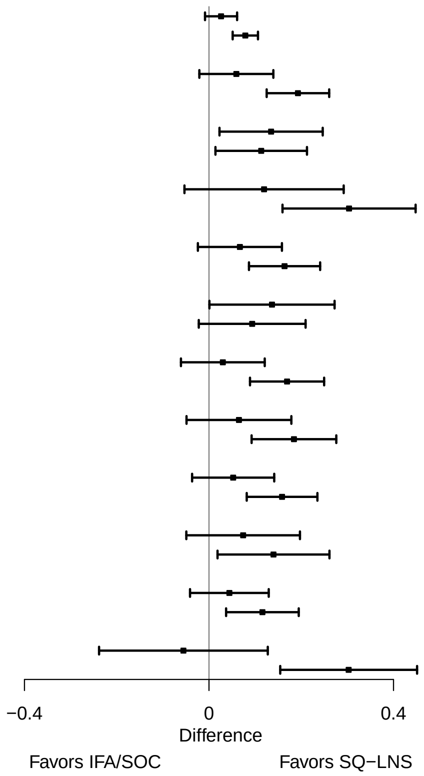

**Supplemental figure 3A: Sex**

#### 3A2: Relative risks for birth outcomes

| Outcome (p−for−interaction) | SQ−LNS N | IFA/SOC N | Relative risk (95% CI) |
|---|---|---|---|
| Low birth weight (p=0.016) | | | |
| Male | 901 | 1736 | 1.01 (0.86, 1.18) |
| Female | 914 | 1722 | 0.80 (0.71, 0.90) |
| Birth weight < 2 kg (p=0.018) | | | |
| Male | 797 | 1620 | 1.14 (0.79, 1.66) |
| Female | 795 | 1622 | 0.50 (0.30, 0.84) |
| Small−for−gestational age (p=0.283) | | | |
| Male | 881 | 1707 | 0.99 (0.91, 1.06) |
| Female | 892 | 1701 | 0.93 (0.87, 1.00) |
| Large−for−gestational age (p=0.589) | | | |
| Male | 344 | 339 | 1.06 (0.49, 2.30) |
| Female | 350 | 352 | 0.78 (0.35, 1.75) |
| Newborn stunting (p=0.332) | | | |
| Male | 831 | 1677 | 0.88 (0.74, 1.06) |
| Female | 849 | 1657 | 0.76 (0.64, 0.90) |
| Low LGAZ (p=0.722) | | | |
| Male | 811 | 1648 | 0.94 (0.79, 1.13) |
| Female | 827 | 1636 | 0.89 (0.72, 1.09) |
| Low BMIZ (p=0.017) | | | |
| Male | 829 | 1670 | 1.02 (0.89, 1.16) |
| Female | 847 | 1656 | 0.78 (0.66, 0.92) |
| Low HCZ (p=0.848) | | | |
| Male | 832 | 1678 | 0.86 (0.68, 1.08) |
| Female | 848 | 1658 | 0.83 (0.71, 0.96) |
| Low HCGAZ (p=0.859) | | | |
| Male | 696 | 1519 | 0.89 (0.70, 1.14) |
| Female | 689 | 1503 | 0.85 (0.69, 1.05) |
| Preterm birth (p=0.080) | | | |
| Male | 918 | 1748 | 1.08 (0.86, 1.36) |
| Female | 922 | 1726 | 0.82 (0.65, 1.03) |

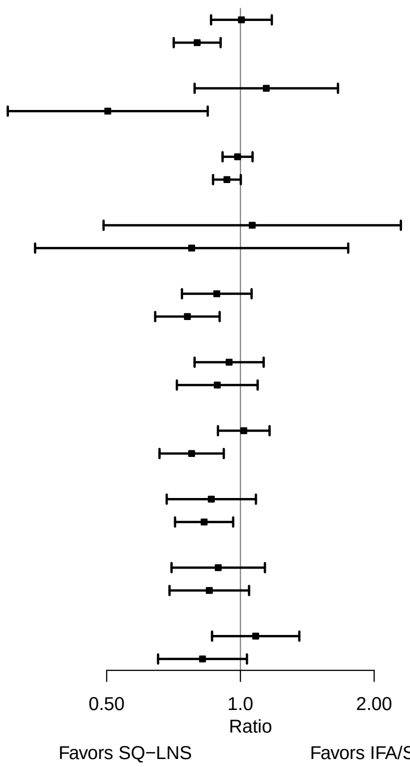

**Supplemental figure 3A: Sex**

#### 3A3: Mean differences for 6 mo outcomes

| Outcome (p−for−interaction) | SQ−LNS N | IFA/SOC N | Mean difference (95% CI) |
|---|---|---|---|
| Weight−for−age z score (p=0.005) | | | |
| Male | 849 | 1661 | −0.05 (−0.15, 0.06) |
| Female | 867 | 1652 | 0.12 (0.03, 0.20) |
| Length−for−age z−score (p=0.008) | | | |
| Male | 849 | 1660 | −0.02 (−0.11, 0.08) |
| Female | 868 | 1655 | 0.12 (0.04, 0.20) |
| Weight−for−length z−score (p=0.075) | | | |
| Male | 849 | 1660 | −0.06 (−0.15, 0.04) |
| Female | 867 | 1652 | 0.05 (−0.04, 0.14) |
| Head circumference−for−age z−score (p=0.035) | | | |
| Male | 849 | 1659 | −0.05 (−0.13, 0.02) |
| Female | 868 | 1652 | 0.06 (−0.02, 0.14) |
| MUAC−for−age z−score (p=0.025) | | | |
| Male | 747 | 1550 | −0.07 (−0.16, 0.03) |
| Female | 751 | 1552 | 0.07 (−0.03, 0.16) |

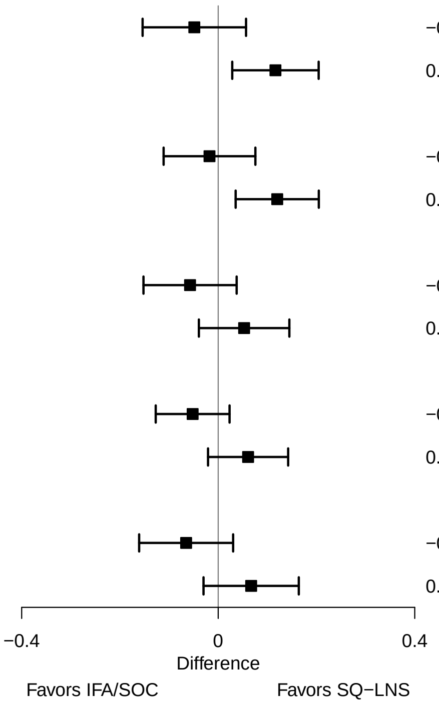

**Supplemental figure 3A: Sex**

#### 3A4: Prevalence ratios for 6 mo outcomes

| Outcome (p−for−interaction) | SQ−LNS N | IFA/SOC N | Prevalence ratio (95% CI) |
|---|---|---|---|
| Underweight (p=0.001) | | | |
| Male | 849 | 1661 | 1.08 (0.88, 1.32) |
| Female | 867 | 1652 | 0.74 (0.59, 0.93) |
| Stunted (p=0.530) | | | |
| Male | 849 | 1660 | 0.93 (0.79, 1.09) |
| Female | 868 | 1655 | 0.87 (0.73, 1.04) |
| Wasted (p=0.503) | | | |
| Male | 747 | 1549 | 0.88 (0.55, 1.40) |
| Female | 750 | 1549 | 1.08 (0.64, 1.82) |
| Low HCAZ (p=0.001) | | | |
| Male | 849 | 1659 | 1.09 (0.94, 1.26) |
| Female | 868 | 1652 | 0.70 (0.56, 0.87) |
| Low MUAC (p=0.972) | | | |
| Male | 747 | 1550 | 1.19 (0.85, 1.65) |
| Female | 751 | 1552 | 1.02 (0.71, 1.45) |
| Acute malnutrition (p=0.365) | | | |
| Male | 747 | 1549 | 0.91 (0.66, 1.25) |
| Female | 750 | 1549 | 1.09 (0.79, 1.49) |

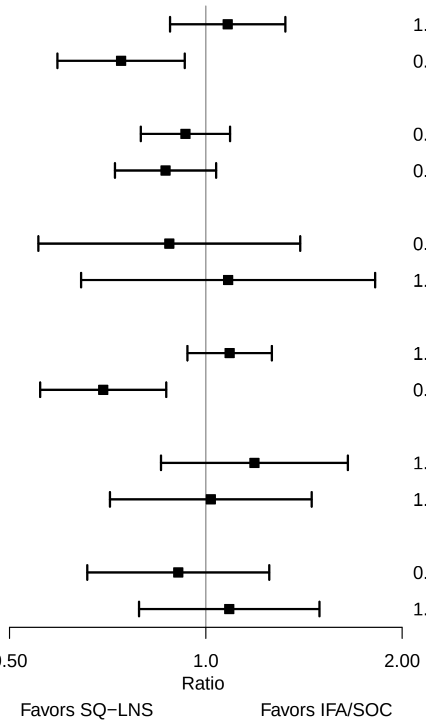

### Supplemental figure 3B: Birth order

#### 3B1: Mean differences for birth outcomes

| Outcome (p−for−interaction) | SQ−LNS N | IFA/SOC N | Mean difference (95% CI) |
|---|---|---|---|
| Weight (kg) (p=0.983) | | | |
| Later born | 1250 | 2281 | 0.05 (0.03, 0.08) |
| Firstborn | 564 | 1171 | 0.05 (0.01, 0.10) |
| Weight−for−age z score (p=0.890) | | | |
| Later born | 1250 | 2281 | 0.13 (0.07, 0.19) |
| Firstborn | 564 | 1171 | 0.14 (0.03, 0.24) |
| Weight−for−gestational age z−score (p=0.129) | | | |
| Later born | 677 | 663 | 0.10 (0.02, 0.19) |
| Firstborn | 189 | 187 | 0.24 (0.06, 0.43) |
| Birth length (cm) (p=0.861) | | | |
| Later born | 1150 | 2180 | 0.21 (0.08, 0.34) |
| Firstborn | 529 | 1148 | 0.21 (−0.02, 0.43) |
| Length−for−age z−score (p=0.997) | | | |
| Later born | 1150 | 2180 | 0.12 (0.06, 0.19) |
| Firstborn | 529 | 1148 | 0.10 (−0.01, 0.22) |
| Length−for−gestational age z−score (p=0.156) | | | |
| Later born | 578 | 562 | 0.10 (0.00, 0.19) |
| Firstborn | 154 | 165 | 0.22 (0.01, 0.42) |
| BMI z−score (p=0.961) | | | |
| Later born | 1148 | 2175 | 0.10 (0.03, 0.17) |
| Firstborn | 527 | 1145 | 0.12 (0.01, 0.23) |
| Head circumference (cm) (p=0.582) | | | |
| Later born | 1150 | 2182 | 0.13 (0.05, 0.22) |
| Firstborn | 529 | 1148 | 0.08 (−0.06, 0.23) |
| Head circumference−for−age z−score (p=0.430) | | | |
| Later born | 1150 | 2182 | 0.12 (0.05, 0.19) |
| Firstborn | 529 | 1148 | 0.06 (−0.05, 0.17) |
| Head circumference−for−gestational age z score (p=0.986) | | | |
| Later born | 578 | 564 | 0.08 (−0.02, 0.18) |
| Firstborn | 154 | 164 | 0.08 (−0.12, 0.29) |
| Mid−upper arm circumference (cm) (p=0.760) | | | |
| Later born | 945 | 1975 | 0.07 (0.01, 0.13) |
| Firstborn | 515 | 1139 | 0.11 (0.00, 0.21) |
| Duration (wk) (p=0.406) | | | |
| Later born | 1261 | 2288 | 0.15 (0.00, 0.29) |
| Firstborn | 593 | 1199 | 0.06 (−0.15, 0.27) |

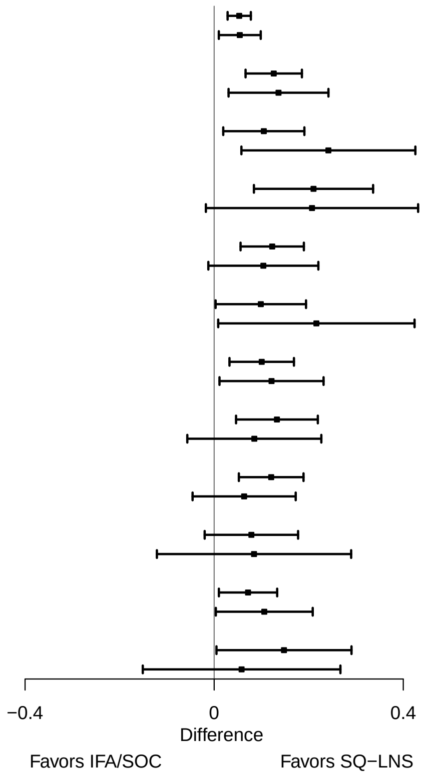

**Supplemental figure 3B: Birth order**

#### 3B2: Relative risks for birth outcomes

| Outcome (p−for−interaction) | SQ−LNS N | IFA/SOC N | Relative risk (95% CI) |
|---|---|---|---|
| Low birth weight (p=0.972) | | | |
| Later born | 1250 | 2281 | 0.88 (0.77, 0.99) |
| Firstborn | 564 | 1171 | 0.89 (0.76, 1.05) |
| Birth weight < 2 kg (p=0.602) | | | |
| Later born | 1041 | 2074 | 0.84 (0.58, 1.23) |
| Firstborn | 550 | 1162 | 0.72 (0.50, 1.04) |
| Small−for−gestational age (p=0.633) | | | |
| Later born | 1210 | 2232 | 0.94 (0.87, 1.00) |
| Firstborn | 562 | 1170 | 0.98 (0.90, 1.06) |
| Large−for−gestational age (p=0.316) | | | |
| Later born | 1041 | 2074 | 0.88 (0.51, 1.53) |
| Firstborn | 550 | 1162 | 1.85 (0.52, 6.58) |
| Newborn stunting (p=0.195) | | | |
| Later born | 1150 | 2180 | 0.86 (0.74, 1.01) |
| Firstborn | 529 | 1148 | 0.76 (0.64, 0.91) |
| Low LGAZ (p=0.809) | | | |
| Later born | 1110 | 2131 | 0.90 (0.76, 1.06) |
| Firstborn | 527 | 1147 | 0.88 (0.72, 1.09) |
| Low BMIZ (p=0.766) | | | |
| Later born | 939 | 1968 | 0.91 (0.78, 1.06) |
| Firstborn | 513 | 1136 | 0.88 (0.78, 1.00) |
| Low HCZ (p=0.069) | | | |
| Later born | 1150 | 2182 | 0.92 (0.81, 1.05) |
| Firstborn | 529 | 1148 | 0.74 (0.61, 0.91) |
| Low HCGAZ (p=0.142) | | | |
| Later born | 729 | 1757 | 0.98 (0.79, 1.21) |
| Firstborn | 474 | 1094 | 0.76 (0.61, 0.95) |
| Preterm birth (p=0.517) | | | |
| Later born | 1261 | 2288 | 0.90 (0.73, 1.11) |
| Firstborn | 593 | 1199 | 1.00 (0.80, 1.26) |

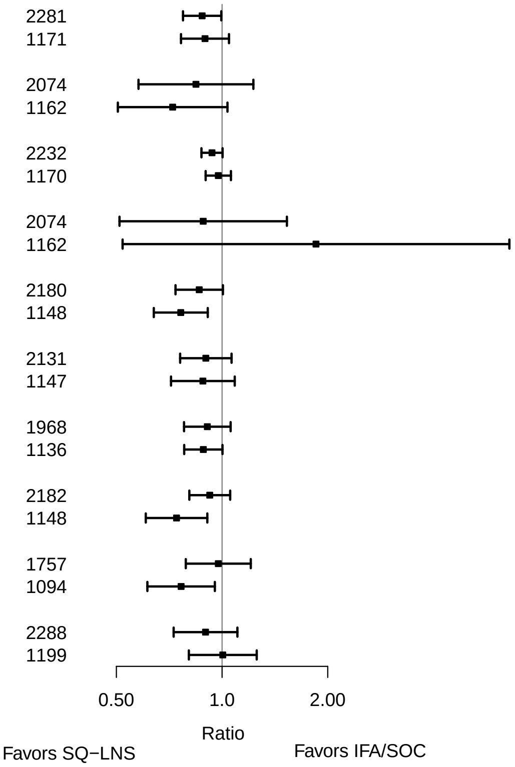

**Supplemental figure 3B: Birth order**

#### 3B3: Mean differences for 6 mo outcomes

| Outcome (p−for−interaction) | SQ−LNS N | IFA/SOC N | Mean difference (95% CI) |
|---|---|---|---|
| Weight−for−age z score (p=0.143) | | | |
| Later born | 1176 | 2196 | 0.06 (−0.02, 0.14) |
| Firstborn | 540 | 1113 | 0.00 (−0.13, 0.12) |
| Length−for−age z−score (p=0.134) | | | |
| Later born | 1176 | 2199 | 0.08 (0.00, 0.15) |
| Firstborn | 541 | 1112 | −0.04 (−0.15, 0.06) |
| Weight−for−length z−score (p=0.594) | | | |
| Later born | 1176 | 2196 | 0.00 (−0.08, 0.08) |
| Firstborn | 540 | 1112 | −0.02 (−0.13, 0.08) |
| Head circumference−for−age z−score (p=0.386) | | | |
| Later born | 1176 | 2194 | 0.03 (−0.04, 0.09) |
| Firstborn | 541 | 1113 | −0.01 (−0.11, 0.09) |
| MUAC−for−age z−score (p=0.879) | | | |
| Later born | 971 | 1995 | −0.01 (−0.09, 0.08) |
| Firstborn | 527 | 1103 | 0.01 (−0.12, 0.13) |

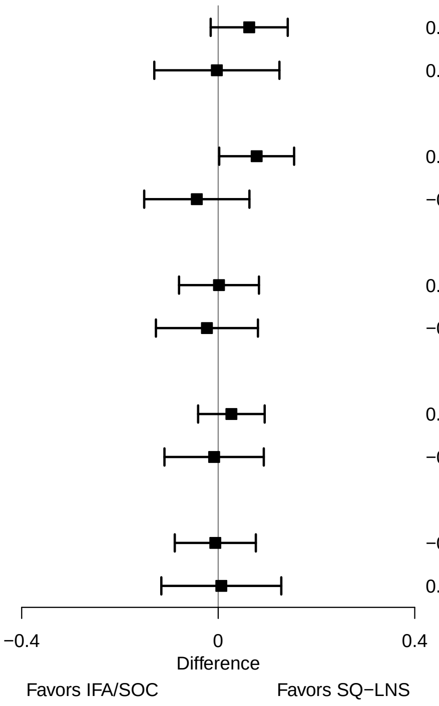

**Supplemental figure 3B: Birth order**

#### 3B4: Prevalence ratios for 6 mo outcomes

| Outcome (p−for−interaction) | SQ−LNS N | IFA/SOC N | Prevalence ratio (95% CI) |
|---|---|---|---|
| Underweight (p=0.026) | | | |
| Later born | 1176 | 2196 | 0.79 (0.66, 0.93) |
| Firstborn | 540 | 1113 | 1.11 (0.87, 1.42) |
| Stunted (p=0.074) | | | |
| Later born | 1176 | 2199 | 0.84 (0.72, 0.97) |
| Firstborn | 541 | 1112 | 1.04 (0.85, 1.28) |
| Wasted (p=0.766) | | | |
| Later born | 691 | 1695 | 0.92 (0.61, 1.39) |
| Firstborn | 456 | 1035 | 0.98 (0.56, 1.70) |
| Low HCAZ (p=0.202) | | | |
| Later born | 1176 | 2194 | 0.84 (0.73, 0.97) |
| Firstborn | 541 | 1113 | 0.99 (0.81, 1.21) |
| Low MUAC (p=0.117) | | | |
| Later born | 971 | 1995 | 0.83 (0.60, 1.16) |
| Firstborn | 527 | 1103 | 1.38 (0.92, 2.06) |
| Acute malnutrition (p=0.171) | | | |
| Later born | 971 | 1992 | 0.89 (0.68, 1.17) |
| Firstborn | 526 | 1102 | 1.27 (0.87, 1.85) |

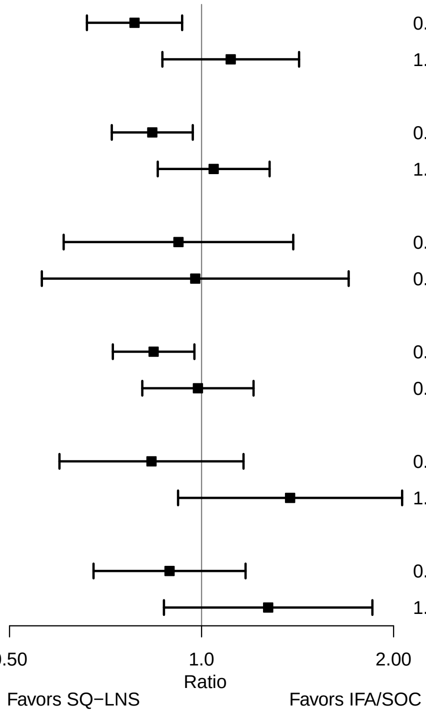

### Supplemental figure 3C: Maternal height

#### 3C1: Mean differences for birth outcomes

| Outcome (p−for−interaction) | SQ−LNS N | IFA/SOC N | Mean difference (95% CI) |
|---|---|---|---|
| Weight (kg) (p=0.656) | | | |
| At least 150.1 | 1154 | 1960 | 0.04 (0.01, 0.08) |
| Less than 150.1 | 624 | 1415 | 0.04 (0.01, 0.08) |
| Weight−for−age z score (p=0.682) | | | |
| At least 150.1 | 1154 | 1960 | 0.11 (0.03, 0.18) |
| Less than 150.1 | 624 | 1415 | 0.11 (0.03, 0.20) |
| Weight−for−gestational age z−score (p=0.922) | | | |
| At least 150.1 | 662 | 625 | 0.09 (−0.02, 0.19) |
| Less than 150.1 | 198 | 218 | 0.15 (0.04, 0.27) |
| Birth length (cm) (p=0.767) | | | |
| At least 150.1 | 1038 | 1856 | 0.17 (0.02, 0.33) |
| Less than 150.1 | 605 | 1395 | 0.15 (−0.02, 0.32) |
| Length−for−age z−score (p=0.795) | | | |
| At least 150.1 | 1038 | 1856 | 0.10 (0.02, 0.18) |
| Less than 150.1 | 605 | 1395 | 0.10 (0.00, 0.19) |
| Length−for−gestational age z−score (p=0.999) | | | |
| At least 150.1 | 546 | 521 | 0.09 (−0.03, 0.21) |
| Less than 150.1 | 180 | 199 | 0.10 (−0.04, 0.23) |
| BMI z−score (p=0.591) | | | |
| At least 150.1 | 1035 | 1851 | 0.09 (0.01, 0.17) |
| Less than 150.1 | 604 | 1392 | 0.10 (0.01, 0.19) |
| Head circumference (cm) (p=0.804) | | | |
| At least 150.1 | 1037 | 1857 | 0.12 (0.02, 0.22) |
| Less than 150.1 | 606 | 1396 | 0.06 (−0.07, 0.19) |
| Head circumference−for−age z−score (p=0.800) | | | |
| At least 150.1 | 1037 | 1857 | 0.10 (0.02, 0.18) |
| Less than 150.1 | 606 | 1396 | 0.07 (−0.04, 0.17) |
| Head circumference−for−gestational age z score (p=0.741) | | | |
| At least 150.1 | 545 | 522 | 0.12 (0.00, 0.25) |
| Less than 150.1 | 181 | 199 | 0.04 (−0.12, 0.20) |
| Mid−upper arm circumference (cm) (p=0.263) | | | |
| At least 150.1 | 993 | 1831 | 0.05 (−0.02, 0.13) |
| Less than 150.1 | 432 | 1206 | 0.08 (−0.02, 0.18) |
| Duration (wk) (p=0.269) | | | |
| At least 150.1 | 1209 | 2017 | 0.06 (−0.10, 0.22) |
| Less than 150.1 | 609 | 1393 | 0.18 (−0.03, 0.39) |

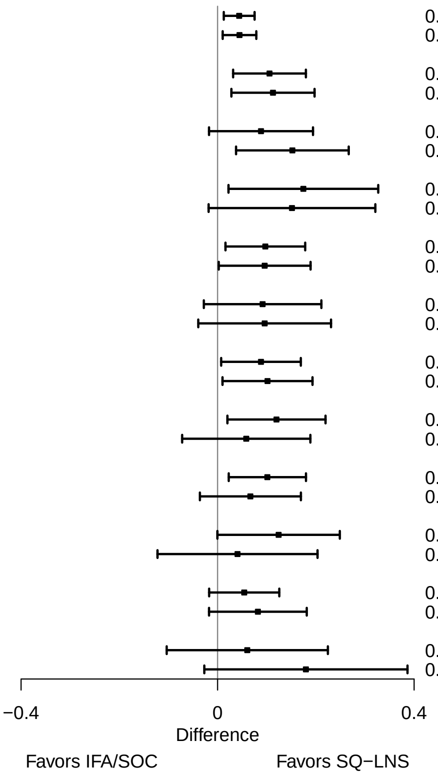

**Supplemental figure 3C: Maternal height**

#### 3C2: Relative risks for birth outcomes

| Outcome (p−for−interaction) | SQ−LNS N | IFA/SOC N | Relative risk (95% CI) |
|---|---|---|---|
| Low birth weight (p=0.365) | | | |
| At least 150.1 | 863 | 1679 | 0.89 (0.77, 1.04) |
| Less than 150.1 | 613 | 1398 | 0.94 (0.82, 1.08) |
| Birth weight < 2 kg (p=0.653) | | | |
| At least 150.1 | 816 | 1653 | 0.92 (0.60, 1.42) |
| Less than 150.1 | 438 | 1208 | 0.76 (0.50, 1.15) |
| Small−for−gestational age (p=0.829) | | | |
| At least 150.1 | 1145 | 1953 | 0.95 (0.87, 1.04) |
| Less than 150.1 | 591 | 1372 | 0.97 (0.91, 1.03) |
| Newborn stunting (p=0.755) | | | |
| At least 150.1 | 747 | 1575 | 0.83 (0.64, 1.08) |
| Less than 150.1 | 594 | 1378 | 0.85 (0.72, 1.00) |
| Low LGAZ (p=0.071) | | | |
| At least 150.1 | 700 | 1549 | 0.79 (0.65, 0.96) |
| Less than 150.1 | 419 | 1188 | 1.03 (0.85, 1.24) |
| Low BMIZ (p=0.935) | | | |
| At least 150.1 | 697 | 1544 | 0.91 (0.80, 1.04) |
| Less than 150.1 | 418 | 1185 | 0.92 (0.79, 1.07) |
| Low HCZ (p=0.605) | | | |
| At least 150.1 | 699 | 1550 | 0.89 (0.76, 1.04) |
| Less than 150.1 | 420 | 1189 | 0.83 (0.67, 1.03) |
| Preterm birth (p=0.252) | | | |
| At least 150.1 | 1209 | 2017 | 1.03 (0.83, 1.27) |
| Less than 150.1 | 609 | 1393 | 0.85 (0.65, 1.12) |

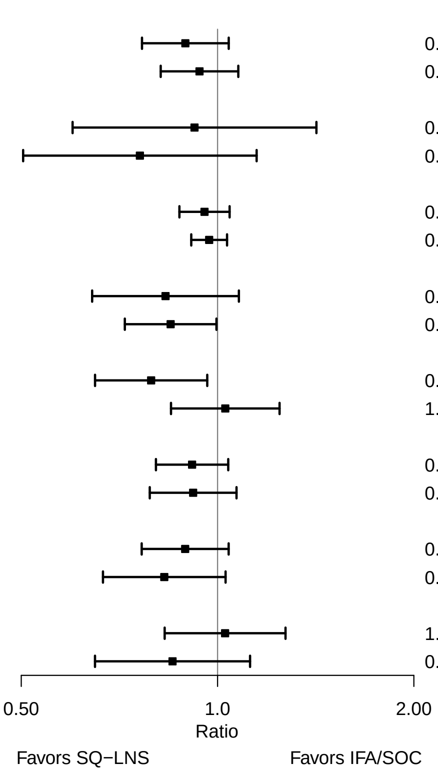

**Supplemental figure 3C: Maternal height**

#### 3C3: Mean differences for 6 mo outcomes

| Outcome (p−for−interaction) | SQ−LNS N | IFA/SOC N | Mean difference (95% CI) |
|---|---|---|---|
| Weight−for−age z score (p=0.513) | | | |
| At least 150.1 | 1085 | 1882 | 0.05 (−0.05, 0.14) |
| Less than 150.1 | 598 | 1359 | 0.01 (−0.10, 0.12) |
| Length−for−age z−score (p=0.490) | | | |
| At least 150.1 | 1086 | 1884 | 0.03 (−0.06, 0.12) |
| Less than 150.1 | 598 | 1359 | 0.08 (−0.01, 0.17) |
| Weight−for−length z−score (p=0.073) | | | |
| At least 150.1 | 1085 | 1881 | 0.04 (−0.05, 0.13) |
| Less than 150.1 | 598 | 1359 | −0.06 (−0.15, 0.04) |
| Head circumference−for−age z−score (p=0.861) | | | |
| At least 150.1 | 1086 | 1884 | 0.01 (−0.07, 0.08) |
| Less than 150.1 | 598 | 1355 | 0.00 (−0.08, 0.09) |
| MUAC−for−age z−score (p=0.664) | | | |
| At least 150.1 | 1039 | 1858 | 0.02 (−0.06, 0.10) |
| Less than 150.1 | 427 | 1172 | −0.05 (−0.17, 0.06) |

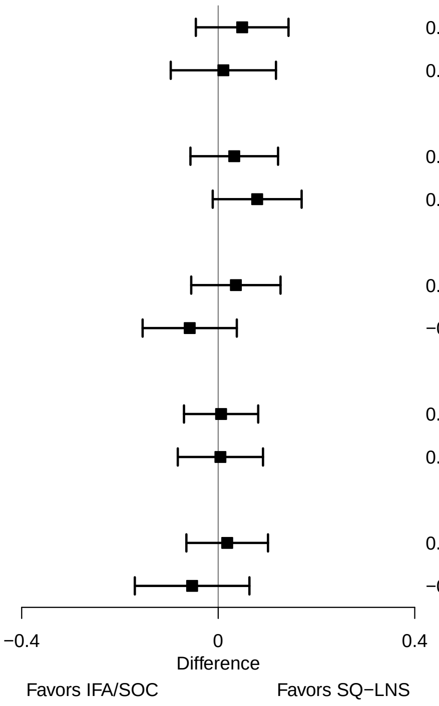

**Supplemental figure 3C: Maternal height**

#### 3C4: Prevalence ratios for 6 mo outcomes

| Outcome (p−for−interaction) | SQ−LNS N | IFA/SOC N | Prevalence ratio (95% CI) |
|---|---|---|---|
| Underweight (p=0.478) | | | |
| At least 150.1 | 820 | 1633 | 0.91 (0.69, 1.21) |
| Less than 150.1 | 587 | 1343 | 0.93 (0.77, 1.13) |
| Stunted (p=0.797) | | | |
| At least 150.1 | 1086 | 1884 | 0.95 (0.79, 1.16) |
| Less than 150.1 | 598 | 1359 | 0.89 (0.77, 1.01) |
| Low HCAZ (p=0.646) | | | |
| At least 150.1 | 820 | 1633 | 0.95 (0.79, 1.14) |
| Less than 150.1 | 587 | 1339 | 0.89 (0.76, 1.03) |
| Low MUAC (p=0.037) | | | |
| At least 150.1 | 1039 | 1858 | 1.14 (0.85, 1.53) |
| Less than 150.1 | 427 | 1172 | 0.88 (0.58, 1.33) |
| Acute malnutrition (p=0.079) | | | |
| At least 150.1 | 1038 | 1854 | 1.09 (0.83, 1.44) |
| Less than 150.1 | 427 | 1172 | 0.88 (0.64, 1.21) |

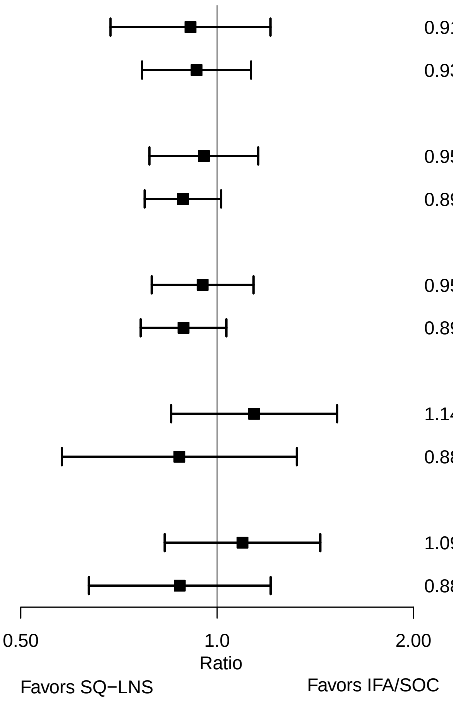

### Supplemental figure 3D: Maternal BMI

#### 3D1: Mean differences for birth outcomes

| Outcome (p−for−interaction) | SQ−LNS N | IFA/SOC N | Mean difference (95% CI) |
|---|---|---|---|
| Weight (kg) (p=0.233) | | | |
| At least 20 | 1041 | 1806 | 0.04 (0.02, 0.07) |
| Less than 20 | 736 | 1567 | 0.06 (0.03, 0.09) |
| Weight−for−age z score (p=0.276) | | | |
| At least 20 | 1041 | 1806 | 0.11 (0.04, 0.18) |
| Less than 20 | 736 | 1567 | 0.14 (0.06, 0.22) |
| Weight−for−gestational age z−score (p=0.995) | | | |
| At least 20 | 613 | 588 | 0.15 (0.07, 0.23) |
| Less than 20 | 246 | 253 | 0.06 (−0.10, 0.23) |
| Birth length (cm) (p=0.186) | | | |
| At least 20 | 975 | 1749 | 0.16 (0.02, 0.30) |
| Less than 20 | 667 | 1501 | 0.27 (0.10, 0.45) |
| Length−for−age z−score (p=0.303) | | | |
| At least 20 | 975 | 1749 | 0.10 (0.02, 0.18) |
| Less than 20 | 667 | 1501 | 0.14 (0.05, 0.23) |
| Length−for−gestational age z−score (p=0.791) | | | |
| At least 20 | 548 | 532 | 0.13 (0.04, 0.22) |
| Less than 20 | 177 | 187 | 0.12 (−0.09, 0.33) |
| BMI z−score (p=0.239) | | | |
| At least 20 | 974 | 1748 | 0.09 (0.01, 0.16) |
| Less than 20 | 664 | 1494 | 0.13 (0.04, 0.22) |
| Head circumference (cm) (p=0.111) | | | |
| At least 20 | 973 | 1751 | 0.07 (−0.01, 0.16) |
| Less than 20 | 669 | 1501 | 0.17 (0.05, 0.28) |
| Head circumference−for−age z−score (p=0.185) | | | |
| At least 20 | 973 | 1751 | 0.08 (0.01, 0.15) |
| Less than 20 | 669 | 1501 | 0.13 (0.04, 0.22) |
| Head circumference−for−gestational age z score (p=0.884) | | | |
| At least 20 | 546 | 533 | 0.11 (0.01, 0.21) |
| Less than 20 | 179 | 187 | 0.10 (−0.11, 0.31) |
| Mid−upper arm circumference (cm) (p=0.081) | | | |
| At least 20 | 773 | 1548 | 0.03 (−0.04, 0.10) |
| Less than 20 | 651 | 1488 | 0.11 (0.03, 0.20) |
| Duration (wk) (p=0.840) | | | |
| At least 20 | 1062 | 1820 | 0.11 (−0.05, 0.27) |
| Less than 20 | 755 | 1588 | 0.12 (−0.08, 0.31) |

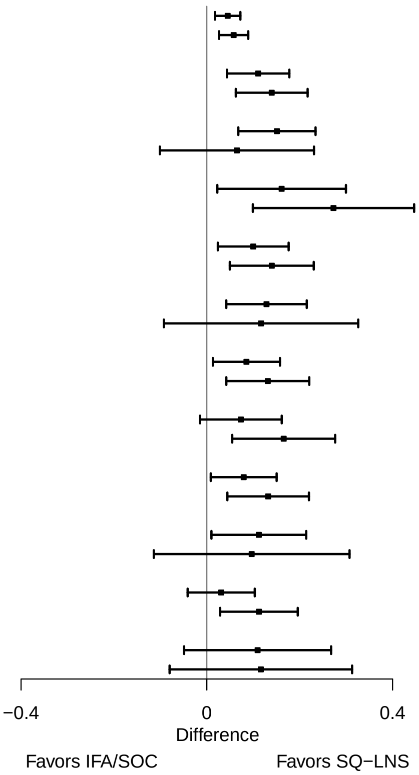

**Supplemental figure 3D: Maternal BMI**

#### 3D2: Relative risks for birth outcomes

| Outcome (p−for−interaction) | SQ−LNS N | IFA/SOC N | Relative risk (95% CI) |
|---|---|---|---|
| Low birth weight (p=0.248) | | | |
| At least 20 | 1041 | 1806 | 0.93 (0.80, 1.08) |
| Less than 20 | 736 | 1567 | 0.87 (0.76, 0.98) |
| Birth weight < 2 kg (p=0.220) | | | |
| At least 20 | 582 | 1357 | 1.08 (0.66, 1.78) |
| Less than 20 | 671 | 1502 | 0.69 (0.46, 1.02) |
| Small−for−gestational age (p=0.923) | | | |
| At least 20 | 1003 | 1759 | 0.95 (0.87, 1.03) |
| Less than 20 | 732 | 1564 | 0.96 (0.90, 1.03) |
| Large−for−gestational age (p=0.488) | | | |
| At least 20 | 582 | 1357 | 0.77 (0.31, 1.88) |
| Less than 20 | 671 | 1502 | 1.46 (0.52, 4.12) |
| Newborn stunting (p=0.342) | | | |
| At least 20 | 975 | 1749 | 0.89 (0.75, 1.06) |
| Less than 20 | 667 | 1501 | 0.80 (0.69, 0.93) |
| Low LGAZ (p=0.640) | | | |
| At least 20 | 937 | 1702 | 0.95 (0.78, 1.15) |
| Less than 20 | 663 | 1498 | 0.88 (0.75, 1.04) |
| Low BMIZ (p=0.683) | | | |
| At least 20 | 974 | 1748 | 0.89 (0.75, 1.05) |
| Less than 20 | 664 | 1494 | 0.89 (0.79, 1.00) |
| Low HCZ (p=0.049) | | | |
| At least 20 | 973 | 1751 | 0.97 (0.81, 1.17) |
| Less than 20 | 669 | 1501 | 0.77 (0.66, 0.90) |
| Low HCGAZ (p=0.157) | | | |
| At least 20 | 637 | 1411 | 0.96 (0.75, 1.24) |
| Less than 20 | 531 | 1363 | 0.81 (0.69, 0.95) |
| Preterm birth (p=0.687) | | | |
| At least 20 | 887 | 1653 | 0.92 (0.71, 1.19) |
| Less than 20 | 741 | 1578 | 0.98 (0.78, 1.24) |

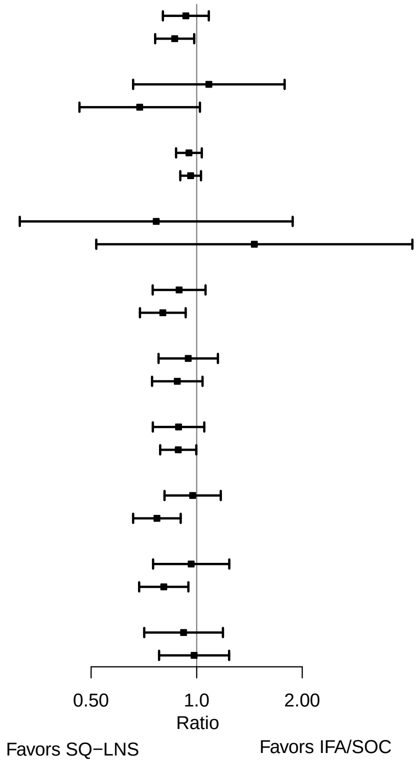

**Supplemental figure 3D: Maternal BMI**

#### 3D3: Mean differences for 6 mo outcomes

| Outcome (p−for−interaction) | SQ−LNS N | IFA/SOC N | Mean difference (95% CI) |
|---|---|---|---|
| Weight−for−age z score (p=0.402) | | | |
| At least 20 | 987 | 1730 | 0.05 (−0.04, 0.14) |
| Less than 20 | 695 | 1510 | 0.05 (−0.06, 0.16) |
| Length−for−age z−score (p=0.922) | | | |
| At least 20 | 987 | 1732 | 0.09 (0.00, 0.18) |
| Less than 20 | 696 | 1510 | 0.05 (−0.05, 0.14) |
| Weight−for−length z−score (p=0.115) | | | |
| At least 20 | 987 | 1730 | −0.02 (−0.10, 0.06) |
| Less than 20 | 695 | 1509 | 0.03 (−0.07, 0.14) |
| Head circumference−for−age z−score (p=0.773) | | | |
| At least 20 | 987 | 1728 | 0.02 (−0.05, 0.09) |
| Less than 20 | 696 | 1510 | 0.02 (−0.07, 0.11) |
| MUAC−for−age z−score (p=0.529) | | | |
| At least 20 | 787 | 1530 | −0.01 (−0.10, 0.08) |
| Less than 20 | 678 | 1499 | 0.02 (−0.08, 0.12) |

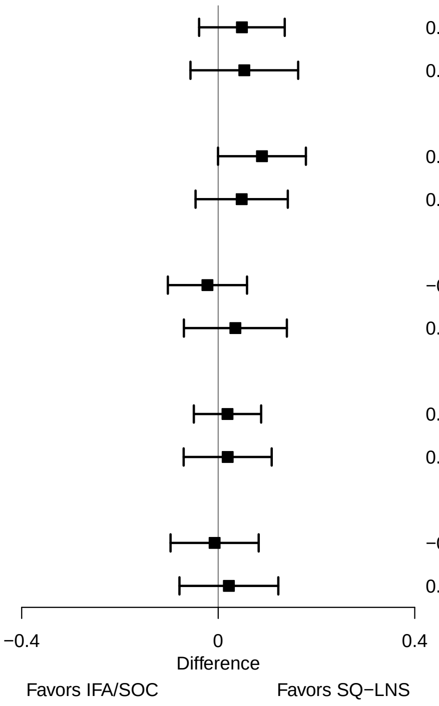

**Supplemental figure 3D: Maternal BMI**

#### 3D4: Prevalence ratios for 6 mo outcomes

| Outcome (p−for−interaction) | SQ−LNS N | IFA/SOC N | Prevalence ratio (95% CI) |
|---|---|---|---|
| Underweight (p=0.838) | | | |
| At least 20 | 987 | 1730 | 0.83 (0.67, 1.04) |
| Less than 20 | 695 | 1510 | 0.95 (0.79, 1.14) |
| Stunted (p=0.802) | | | |
| At least 20 | 987 | 1732 | 0.88 (0.75, 1.03) |
| Less than 20 | 696 | 1510 | 0.91 (0.78, 1.06) |
| Wasted (p=0.916) | | | |
| At least 20 | 787 | 1528 | 0.85 (0.48, 1.52) |
| Less than 20 | 677 | 1497 | 1.01 (0.71, 1.44) |
| Low HCAZ (p=0.595) | | | |
| At least 20 | 787 | 1529 | 0.93 (0.77, 1.13) |
| Less than 20 | 678 | 1498 | 0.88 (0.73, 1.06) |
| Low MUAC (p=0.665) | | | |
| At least 20 | 787 | 1530 | 0.93 (0.61, 1.43) |
| Less than 20 | 678 | 1499 | 1.06 (0.78, 1.45) |
| Acute malnutrition (p=0.605) | | | |
| At least 20 | 787 | 1528 | 0.94 (0.65, 1.38) |
| Less than 20 | 677 | 1497 | 1.06 (0.80, 1.39) |

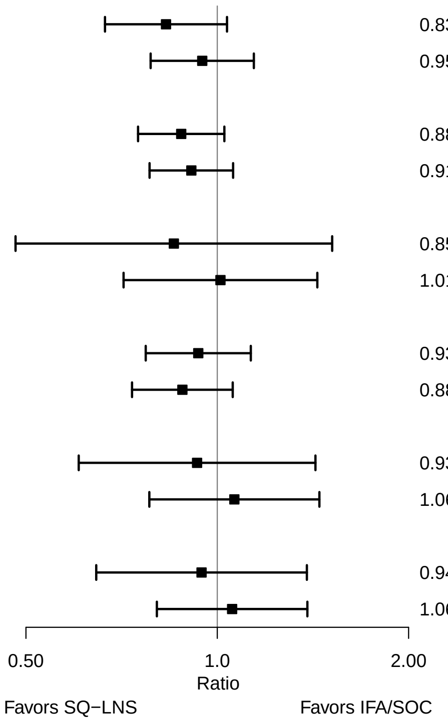

### Supplemental figure 3E: Maternal age

#### 3E1: Mean differences for birth outcomes

| Outcome (p−for−interaction) | SQ−LNS N | IFA/SOC N | Mean difference (95% CI) |
|---|---|---|---|
| Weight (kg) (p=0.257) | | | |
| At least 25 | 723 | 1173 | 0.07 (0.03, 0.11) |
| Less than 25 | 1092 | 2285 | 0.04 (0.01, 0.07) |
| Weight−for−age z score (p=0.395) | | | |
| At least 25 | 723 | 1173 | 0.15 (0.07, 0.24) |
| Less than 25 | 1092 | 2285 | 0.10 (0.03, 0.17) |
| Weight−for−gestational age z−score (p=0.256) | | | |
| At least 25 | 451 | 455 | 0.22 (0.12, 0.33) |
| Less than 25 | 415 | 396 | 0.07 (−0.05, 0.19) |
| Birth length (cm) (p=0.522) | | | |
| At least 25 | 668 | 1113 | 0.26 (0.08, 0.45) |
| Less than 25 | 1012 | 2221 | 0.14 (0.00, 0.29) |
| Length−for−age z−score (p=0.699) | | | |
| At least 25 | 668 | 1113 | 0.13 (0.04, 0.23) |
| Less than 25 | 1012 | 2221 | 0.09 (0.01, 0.17) |
| Length−for−gestational age z−score (p=0.192) | | | |
| At least 25 | 397 | 395 | 0.18 (0.05, 0.32) |
| Less than 25 | 335 | 333 | 0.02 (−0.12, 0.17) |
| BMI z−score (p=0.245) | | | |
| At least 25 | 667 | 1111 | 0.14 (0.05, 0.24) |
| Less than 25 | 1009 | 2215 | 0.08 (0.00, 0.15) |
| Head circumference (cm) (p=0.290) | | | |
| At least 25 | 668 | 1115 | 0.19 (0.07, 0.32) |
| Less than 25 | 1012 | 2221 | 0.06 (−0.03, 0.16) |
| Head circumference−for−age z−score (p=0.395) | | | |
| At least 25 | 668 | 1115 | 0.15 (0.06, 0.25) |
| Less than 25 | 1012 | 2221 | 0.07 (−0.01, 0.15) |
| Head circumference−for−gestational age z score (p=0.862) | | | |
| At least 25 | 397 | 397 | 0.14 (0.00, 0.27) |
| Less than 25 | 335 | 332 | 0.10 (−0.05, 0.24) |
| Mid−upper arm circumference (cm) (p=0.621) | | | |
| At least 25 | 582 | 1012 | 0.10 (0.01, 0.18) |
| Less than 25 | 879 | 2108 | 0.07 (−0.01, 0.14) |
| Duration (wk) (p=0.020) | | | |
| At least 25 | 744 | 1189 | −0.06 (−0.27, 0.15) |
| Less than 25 | 1111 | 2304 | 0.23 (0.06, 0.39) |

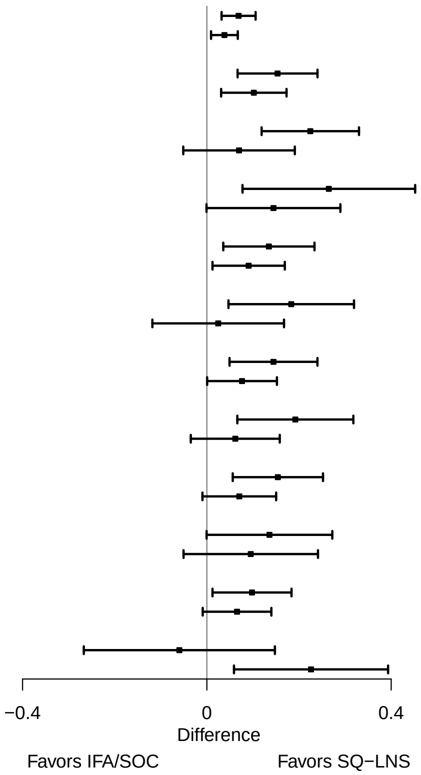

**Supplemental figure 3E: Maternal age**

#### 3E2: Relative risks for birth outcomes

| Outcome (p−for−interaction) | SQ−LNS N | IFA/SOC N | Relative risk (95% CI) |
|---|---|---|---|
| Low birth weight (p=0.481) | | | |
| At least 25 | 723 | 1173 | 0.85 (0.70, 1.03) |
| Less than 25 | 1092 | 2285 | 0.92 (0.82, 1.03) |
| Birth weight < 2 kg (p=0.022) | | | |
| At least 25 | 633 | 1070 | 1.26 (0.77, 2.08) |
| Less than 25 | 959 | 2172 | 0.64 (0.47, 0.86) |
| Small−for−gestational age (p=0.052) | | | |
| At least 25 | 705 | 1151 | 0.87 (0.77, 0.97) |
| Less than 25 | 1068 | 2257 | 1.00 (0.94, 1.06) |
| Large−for−gestational age (p=0.279) | | | |
| At least 25 | 633 | 1070 | 0.83 (0.45, 1.54) |
| Less than 25 | 959 | 2172 | 1.43 (0.59, 3.47) |
| Newborn stunting (p=0.343) | | | |
| At least 25 | 668 | 1113 | 0.91 (0.72, 1.16) |
| Less than 25 | 1012 | 2221 | 0.80 (0.70, 0.92) |
| Low LGAZ (p=0.956) | | | |
| At least 25 | 650 | 1091 | 0.94 (0.74, 1.18) |
| Less than 25 | 988 | 2193 | 0.91 (0.77, 1.06) |
| Low BMIZ (p=0.462) | | | |
| At least 25 | 667 | 1111 | 0.83 (0.65, 1.05) |
| Less than 25 | 1009 | 2215 | 0.93 (0.84, 1.03) |
| Low HCZ (p=0.920) | | | |
| At least 25 | 668 | 1115 | 0.84 (0.66, 1.06) |
| Less than 25 | 1012 | 2221 | 0.85 (0.71, 1.01) |
| Low HCGAZ (p=0.395) | | | |
| At least 25 | 510 | 956 | 0.97 (0.71, 1.32) |
| Less than 25 | 875 | 2066 | 0.84 (0.71, 1.00) |
| Preterm birth (p=0.236) | | | |
| At least 25 | 744 | 1189 | 1.07 (0.82, 1.39) |
| Less than 25 | 1111 | 2304 | 0.87 (0.71, 1.06) |

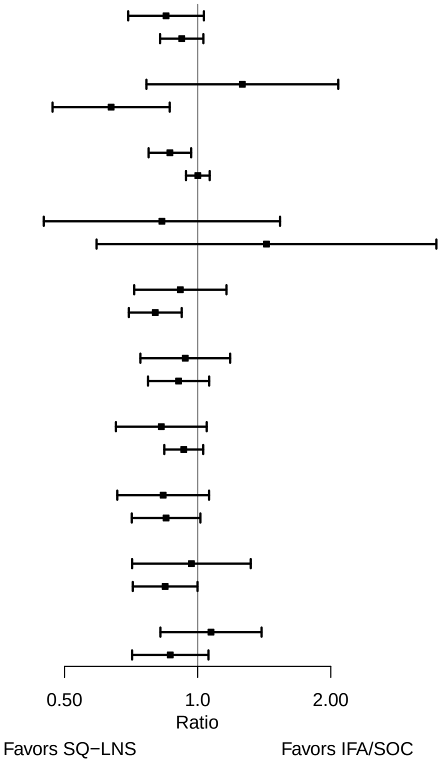

**Supplemental figure 3E: Maternal age**

#### 3E3: Mean differences for 6 mo outcomes

| Outcome (p−for−interaction) | SQ−LNS N | IFA/SOC N | Mean difference (95% CI) |
|---|---|---|---|
| Weight−for−age z score (p=0.924) | | | |
| At least 25 | 671 | 1128 | 0.04 (−0.08, 0.15) |
| Less than 25 | 1045 | 2185 | 0.04 (−0.05, 0.13) |
| Length−for−age z−score (p=0.267) | | | |
| At least 25 | 672 | 1129 | 0.11 (0.00, 0.21) |
| Less than 25 | 1045 | 2186 | 0.03 (−0.06, 0.11) |
| Weight−for−length z−score (p=0.111) | | | |
| At least 25 | 671 | 1128 | −0.04 (−0.15, 0.07) |
| Less than 25 | 1045 | 2184 | 0.02 (−0.05, 0.10) |
| Head circumference−for−age z−score (p=0.698) | | | |
| At least 25 | 672 | 1127 | 0.03 (−0.07, 0.12) |
| Less than 25 | 1045 | 2184 | 0.00 (−0.07, 0.08) |
| MUAC−for−age z−score (p=0.109) | | | |
| At least 25 | 582 | 1025 | −0.07 (−0.18, 0.04) |
| Less than 25 | 916 | 2077 | 0.03 (−0.05, 0.12) |

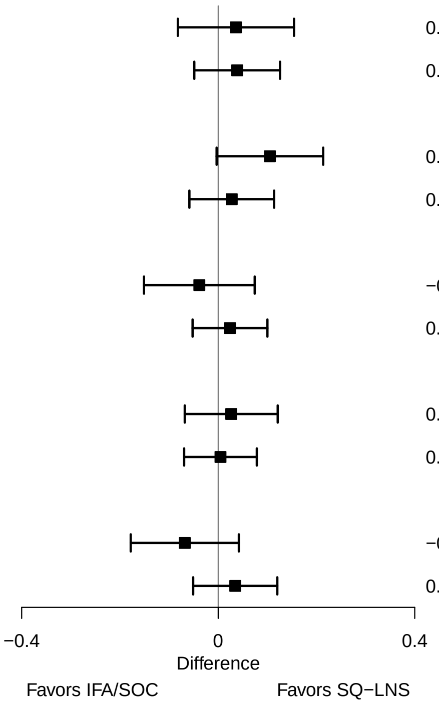

**Supplemental figure 3E: Maternal age**

#### 3E4: Prevalence ratios for 6 mo outcomes

| Outcome (p−for−interaction) | SQ−LNS N | IFA/SOC N | Prevalence ratio (95% CI) |
|---|---|---|---|
| Underweight (p=0.830) | | | |
| At least 25 | 671 | 1128 | 0.85 (0.66, 1.10) |
| Less than 25 | 1045 | 2185 | 0.96 (0.81, 1.14) |
| Stunted (p=0.953) | | | |
| At least 25 | 672 | 1129 | 0.92 (0.75, 1.14) |
| Less than 25 | 1045 | 2186 | 0.90 (0.77, 1.05) |
| Wasted (p=0.519) | | | |
| At least 25 | 581 | 1024 | 0.81 (0.50, 1.32) |
| Less than 25 | 916 | 2074 | 1.09 (0.74, 1.62) |
| Low HCAZ (p=0.341) | | | |
| At least 25 | 672 | 1127 | 0.81 (0.66, 1.00) |
| Less than 25 | 1045 | 2184 | 0.95 (0.80, 1.11) |
| Low MUAC (p=0.289) | | | |
| At least 25 | 582 | 1025 | 0.88 (0.59, 1.32) |
| Less than 25 | 916 | 2077 | 1.14 (0.87, 1.51) |
| Acute malnutrition (p=0.384) | | | |
| At least 25 | 581 | 1024 | 0.92 (0.66, 1.28) |
| Less than 25 | 916 | 2074 | 1.08 (0.84, 1.37) |

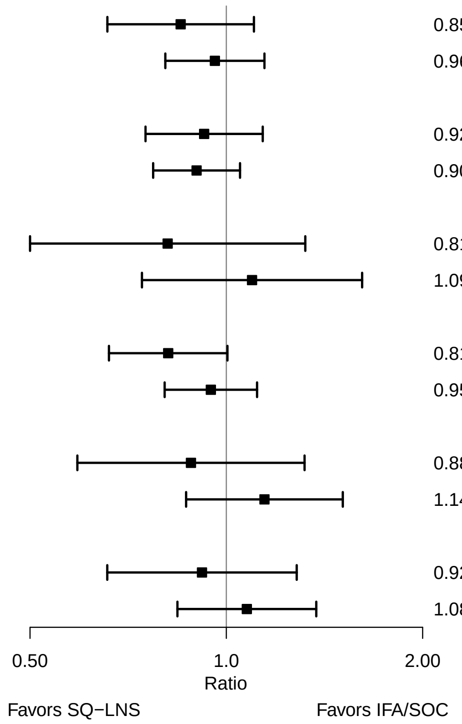

### Supplemental figure 3F: Maternal education

#### 3F1: Mean differences for birth outcomes

| Outcome (p−for−interaction) | SQ−LNS N | IFA/SOC N | Mean difference (95% CI) |
|---|---|---|---|
| Weight (kg) (p=0.929) | | | |
| Primary or greater | 1212 | 2328 | 0.05 (0.02, 0.07) |
| Incomplete or no formal | 600 | 1126 | 0.05 (0.00, 0.10) |
| Weight−for−age z score (p=0.981) | | | |
| Primary or greater | 1212 | 2328 | 0.12 (0.05, 0.18) |
| Incomplete or no formal | 600 | 1126 | 0.11 (0.00, 0.22) |
| Weight−for−gestational age z−score (p=0.773) | | | |
| Primary or greater | 468 | 446 | 0.17 (0.07, 0.27) |
| Incomplete or no formal | 395 | 401 | 0.04 (−0.09, 0.17) |
| Birth length (cm) (p=0.690) | | | |
| Primary or greater | 1194 | 2310 | 0.19 (0.07, 0.32) |
| Incomplete or no formal | 484 | 1022 | 0.17 (−0.08, 0.42) |
| Length−for−age z−score (p=0.665) | | | |
| Primary or greater | 1194 | 2310 | 0.11 (0.04, 0.18) |
| Incomplete or no formal | 484 | 1022 | 0.10 (−0.03, 0.23) |
| Length−for−gestational age z−score (p=0.369) | | | |
| Primary or greater | 450 | 429 | 0.15 (0.06, 0.24) |
| Incomplete or no formal | 280 | 297 | 0.04 (−0.13, 0.20) |
| BMI z−score (p=0.638) | | | |
| Primary or greater | 1193 | 2310 | 0.11 (0.04, 0.18) |
| Incomplete or no formal | 481 | 1014 | 0.05 (−0.08, 0.18) |
| Head circumference (cm) (p=0.797) | | | |
| Primary or greater | 1194 | 2311 | 0.09 (0.00, 0.17) |
| Incomplete or no formal | 484 | 1023 | 0.12 (−0.04, 0.28) |
| Head circumference−for−age z−score (p=0.678) | | | |
| Primary or greater | 1194 | 2311 | 0.08 (0.01, 0.15) |
| Incomplete or no formal | 484 | 1023 | 0.12 (−0.01, 0.24) |
| Head circumference−for−gestational age z score (p=0.726) | | | |
| Primary or greater | 450 | 429 | 0.11 (0.00, 0.23) |
| Incomplete or no formal | 280 | 298 | 0.10 (−0.06, 0.25) |
| Mid−upper arm circumference (cm) (p=0.967) | | | |
| Primary or greater | 985 | 2117 | 0.07 (0.00, 0.14) |
| Incomplete or no formal | 474 | 1001 | 0.08 (−0.02, 0.18) |
| Duration (wk) (p=0.741) | | | |
| Primary or greater | 1205 | 2320 | 0.08 (−0.06, 0.22) |
| Incomplete or no formal | 645 | 1168 | 0.16 (−0.06, 0.39) |

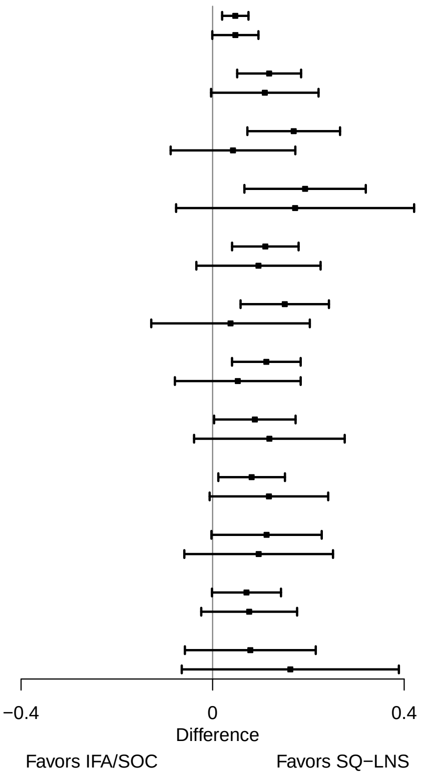

**Supplemental figure 3F: Maternal education**

#### 3F2: Relative risks for birth outcomes

| Outcome (p−for−interaction) | SQ−LNS N | IFA/SOC N | Relative risk (95% CI) |
|---|---|---|---|
| Low birth weight (p=0.216) | | | |
| Primary or greater | 1212 | 2328 | 0.86 (0.76, 0.98) |
| Incomplete or no formal | 600 | 1126 | 1.01 (0.85, 1.20) |
| Birth weight < 2 kg (p=0.607) | | | |
| Primary or greater | 758 | 1892 | 0.78 (0.55, 1.10) |
| Incomplete or no formal | 524 | 1041 | 0.91 (0.61, 1.36) |
| Small−for−gestational age (p=0.466) | | | |
| Primary or greater | 1172 | 2286 | 0.97 (0.91, 1.03) |
| Incomplete or no formal | 598 | 1118 | 0.94 (0.85, 1.03) |
| Large−for−gestational age (p=0.539) | | | |
| Primary or greater | 1003 | 2134 | 0.92 (0.45, 1.88) |
| Incomplete or no formal | 586 | 1104 | 1.06 (0.51, 2.17) |
| Newborn stunting (p=0.285) | | | |
| Primary or greater | 1194 | 2310 | 0.80 (0.68, 0.93) |
| Incomplete or no formal | 484 | 1022 | 0.94 (0.78, 1.13) |
| Low LGAZ (p=0.627) | | | |
| Primary or greater | 985 | 2116 | 0.88 (0.76, 1.03) |
| Incomplete or no formal | 470 | 1000 | 0.95 (0.76, 1.19) |
| Low BMIZ (p=0.089) | | | |
| Primary or greater | 1193 | 2310 | 0.85 (0.76, 0.95) |
| Incomplete or no formal | 481 | 1014 | 1.06 (0.88, 1.29) |
| Low HCZ (p=0.935) | | | |
| Primary or greater | 1194 | 2311 | 0.86 (0.73, 1.01) |
| Incomplete or no formal | 484 | 1023 | 0.85 (0.68, 1.07) |
| Low HCGAZ (p=0.372) | | | |
| Primary or greater | 943 | 2079 | 0.84 (0.68, 1.03) |
| Incomplete or no formal | 261 | 777 | 0.99 (0.75, 1.32) |
| Preterm birth (p=0.671) | | | |
| Primary or greater | 1027 | 2158 | 1.00 (0.81, 1.23) |
| Incomplete or no formal | 633 | 1153 | 0.90 (0.72, 1.12) |

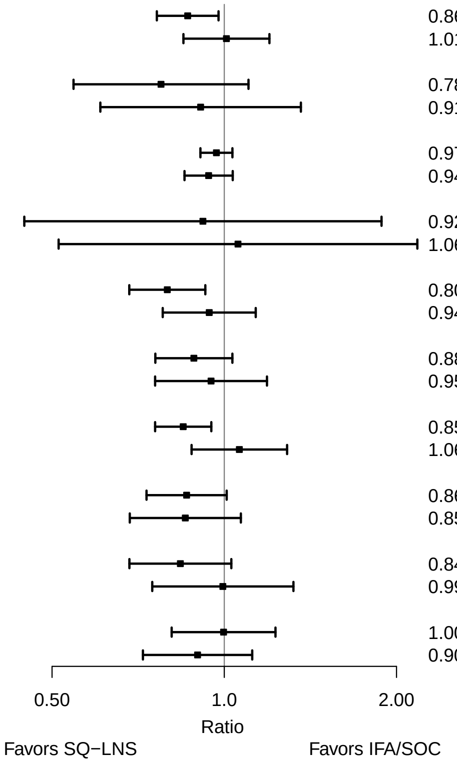

**Supplemental figure 3F: Maternal education**

#### 3F3: Mean differences for 6 mo outcomes

| Outcome (p−for−interaction) | SQ−LNS N | IFA/SOC N | Mean difference (95% CI) |
|---|---|---|---|
| Weight−for−age z score (p=0.559) | | | |
| Primary or greater | 1162 | 2235 | 0.06 (−0.02, 0.14) |
| Incomplete or no formal | 553 | 1077 | −0.01 (−0.14, 0.12) |
| Length−for−age z−score (p=0.631) | | | |
| Primary or greater | 1163 | 2236 | 0.05 (−0.03, 0.13) |
| Incomplete or no formal | 553 | 1078 | 0.05 (−0.07, 0.17) |
| Weight−for−length z−score (p=0.085) | | | |
| Primary or greater | 1162 | 2234 | 0.03 (−0.05, 0.10) |
| Incomplete or no formal | 553 | 1077 | −0.08 (−0.21, 0.05) |
| Head circumference−for−age z−score (p=0.637) | | | |
| Primary or greater | 1163 | 2234 | 0.03 (−0.04, 0.09) |
| Incomplete or no formal | 553 | 1076 | −0.03 (−0.14, 0.08) |
| MUAC−for−age z−score (p=0.572) | | | |
| Primary or greater | 957 | 2042 | −0.01 (−0.10, 0.07) |
| Incomplete or no formal | 540 | 1059 | 0.00 (−0.12, 0.12) |

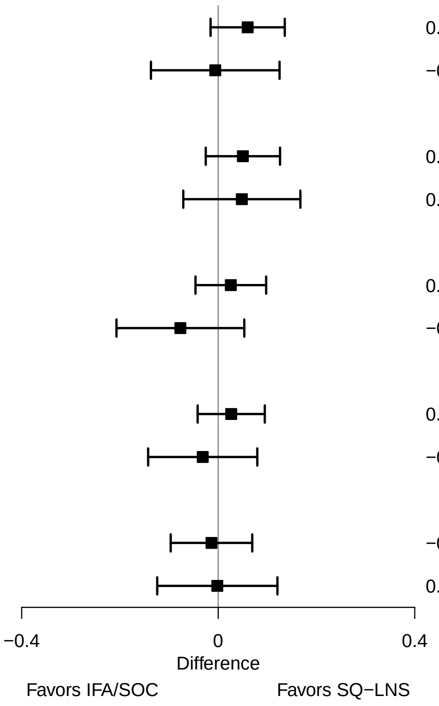

**Supplemental figure 3F: Maternal education**

#### 3F4: Prevalence ratios for 6 mo outcomes

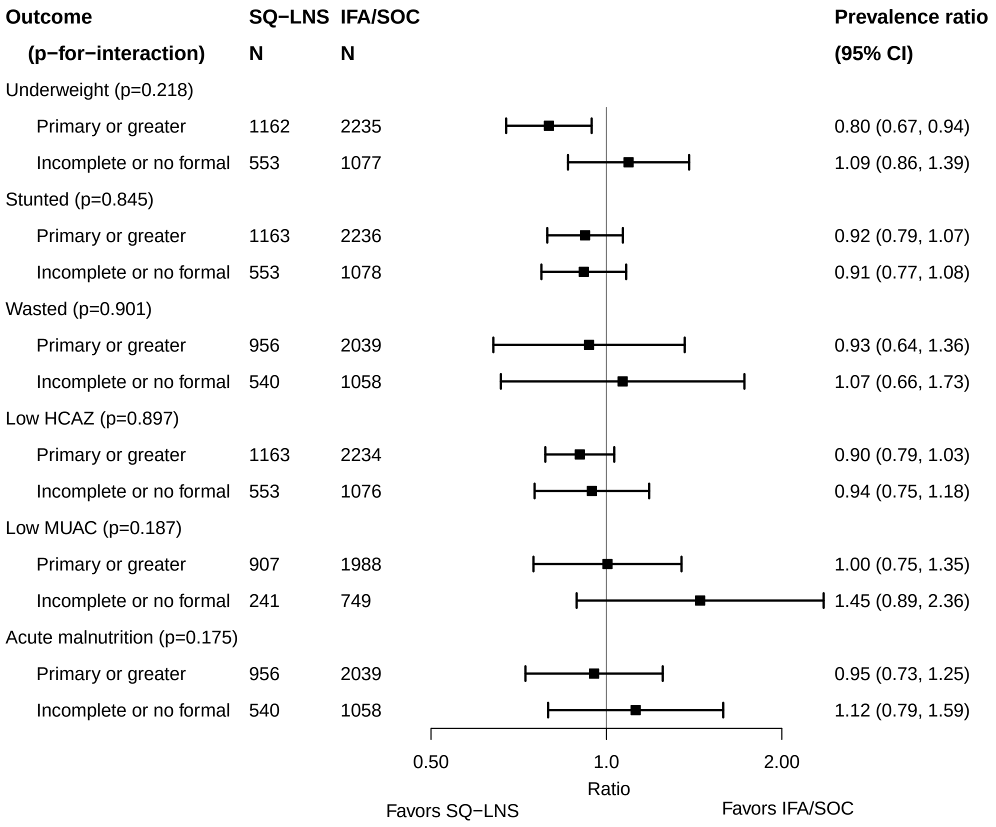

### Supplemental figure 3G: Baseline anemia status

#### 3G1: Mean differences for birth outcomes

| Outcome (p−for−interaction) | SQ−LNS N | IFA/SOC N | Mean difference (95% CI) |
|---|---|---|---|
| Weight (kg) (p=0.633) | | | |
| Anemia | 435 | 429 | 0.04 (−0.02, 0.10) |
| No anemia | 796 | 704 | 0.05 (0.01, 0.09) |
| Weight−for−age z score (p=0.654) | | | |
| Anemia | 435 | 429 | 0.10 (−0.04, 0.23) |
| No anemia | 796 | 704 | 0.13 (0.03, 0.22) |
| Weight−for−gestational age z−score (p=0.653) | | | |
| Anemia | 277 | 287 | 0.11 (−0.05, 0.27) |
| No anemia | 408 | 398 | 0.07 (−0.07, 0.20) |
| Birth length (cm) (p=0.936) | | | |
| Anemia | 374 | 362 | 0.20 (−0.11, 0.51) |
| No anemia | 723 | 648 | 0.15 (−0.06, 0.37) |
| Length−for−age z−score (p=0.814) | | | |
| Anemia | 374 | 362 | 0.12 (−0.05, 0.28) |
| No anemia | 723 | 648 | 0.08 (−0.03, 0.19) |
| Length−for−gestational age z−score (p=0.606) | | | |
| Anemia | 216 | 220 | 0.14 (−0.06, 0.34) |
| No anemia | 335 | 342 | 0.07 (−0.08, 0.23) |
| BMI z−score (p=0.975) | | | |
| Anemia | 371 | 358 | 0.14 (−0.01, 0.28) |
| No anemia | 722 | 644 | 0.12 (0.01, 0.23) |
| Head circumference (cm) (p=0.877) | | | |
| Anemia | 374 | 364 | 0.17 (−0.04, 0.38) |
| No anemia | 722 | 647 | 0.09 (−0.05, 0.23) |
| Head circumference−for−age z−score (p=0.749) | | | |
| Anemia | 374 | 364 | 0.15 (−0.01, 0.32) |
| No anemia | 722 | 647 | 0.07 (−0.04, 0.18) |
| Head circumference−for−gestational age z score (p=0.605) | | | |
| Anemia | 216 | 222 | 0.09 (−0.11, 0.29) |
| No anemia | 335 | 341 | 0.15 (−0.01, 0.31) |
| Mid−upper arm circumference (cm) (p=0.881) | | | |
| Anemia | 375 | 364 | 0.11 (−0.02, 0.25) |
| No anemia | 725 | 647 | 0.07 (−0.01, 0.16) |
| Duration (wk) (p=0.173) | | | |
| Anemia | 471 | 466 | −0.07 (−0.38, 0.24) |
| No anemia | 833 | 741 | 0.19 (−0.03, 0.42) |

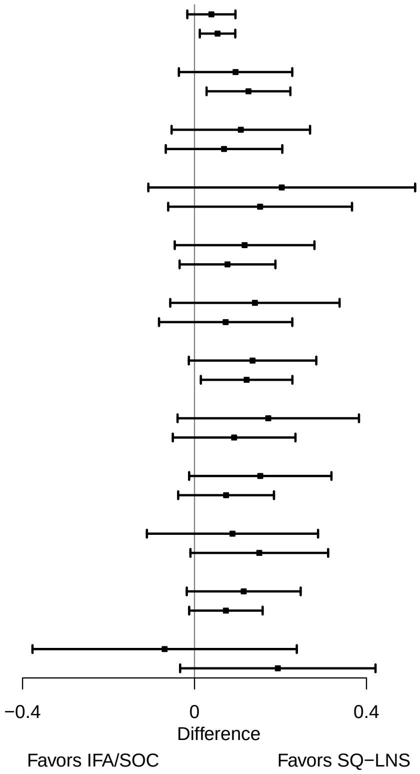

**Supplemental figure 3G: Baseline anemia status**

#### 3G2: Relative risks for birth outcomes

| **Outcome (p−for−interaction)** | **SQ−LNS N** | **IFA/SOC N** | **Relative risk (95% CI)** |
|---|---|---|---|
| Low birth weight (p=0.785) | | | |
| Anemia | 435 | 429 | 0.89 (0.71, 1.11) |
| No anemia | 796 | 704 | 0.84 (0.70, 1.01) |
| Birth weight < 2 kg (p=0.744) | | | |
| Anemia | 435 | 429 | 1.06 (0.55, 2.05) |
| No anemia | 796 | 704 | 0.84 (0.50, 1.42) |
| Small−for−gestational age (p=0.359) | | | |
| Anemia | 435 | 429 | 0.88 (0.73, 1.07) |
| No anemia | 796 | 704 | 0.98 (0.87, 1.12) |
| Large−for−gestational age (p=0.750) | | | |
| Anemia | 282 | 289 | 0.82 (0.33, 2.06) |
| No anemia | 412 | 402 | 0.98 (0.48, 1.98) |
| Newborn stunting (p=0.406) | | | |
| Anemia | 374 | 362 | 0.96 (0.72, 1.29) |
| No anemia | 723 | 648 | 0.83 (0.64, 1.07) |
| Low LGAZ (p=0.773) | | | |
| Anemia | 374 | 362 | 0.94 (0.66, 1.36) |
| No anemia | 723 | 648 | 0.87 (0.67, 1.13) |
| Low BMIZ (p=0.602) | | | |
| Anemia | 371 | 358 | 0.85 (0.63, 1.15) |
| No anemia | 722 | 644 | 0.77 (0.64, 0.92) |
| Low HCZ (p=0.436) | | | |
| Anemia | 374 | 364 | 0.76 (0.56, 1.05) |
| No anemia | 722 | 647 | 0.92 (0.72, 1.17) |
| Preterm birth (p=0.141) | | | |
| Anemia | 471 | 466 | 1.20 (0.86, 1.66) |
| No anemia | 833 | 741 | 0.90 (0.67, 1.21) |

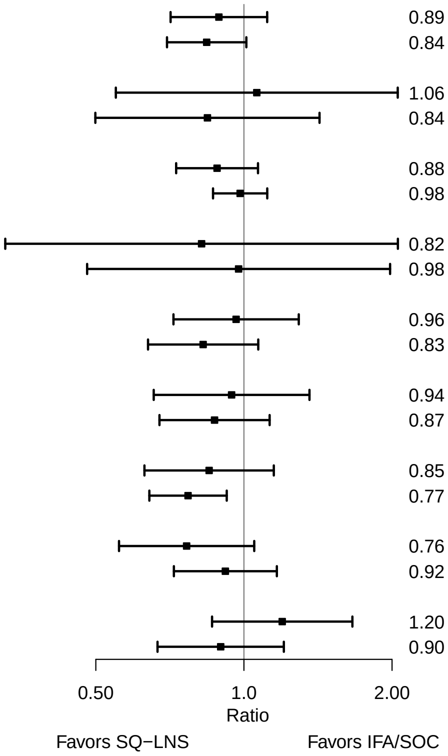

**Supplemental figure 3G: Baseline anemia status**

#### 3G3: Mean differences for 6 mo outcomes

| Outcome (p−for−interaction) | SQ−LNS N | IFA/SOC N | Mean difference (95% CI) |
|---|---|---|---|
| Weight−for−age z score (p=0.763) | | | |
| Anemia | 405 | 413 | 0.04 (−0.11, 0.19) |
| No anemia | 749 | 654 | 0.06 (−0.07, 0.19) |
| Length−for−age z−score (p=0.960) | | | |
| Anemia | 406 | 413 | 0.05 (−0.10, 0.19) |
| No anemia | 749 | 657 | 0.05 (−0.07, 0.18) |
| Weight−for−length z−score (p=0.850) | | | |
| Anemia | 405 | 413 | 0.02 (−0.12, 0.16) |
| No anemia | 749 | 654 | 0.03 (−0.08, 0.15) |
| Head circumference−for−age z−score (p=0.340) | | | |
| Anemia | 406 | 412 | −0.04 (−0.16, 0.09) |
| No anemia | 749 | 656 | 0.06 (−0.04, 0.15) |
| MUAC−for−age z−score (p=0.939) | | | |
| Anemia | 406 | 413 | 0.04 (−0.11, 0.18) |
| No anemia | 749 | 657 | 0.04 (−0.06, 0.15) |

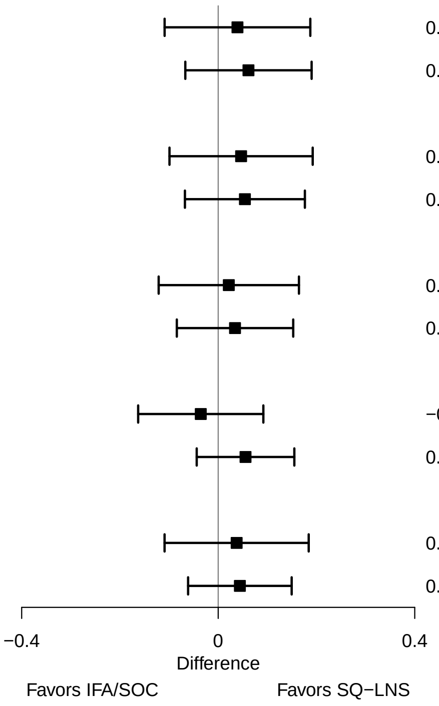

**Supplemental figure 3G: Baseline anemia status**

#### 3G4: Prevalence ratios for 6 mo outcomes

| Outcome (p−for−interaction) | SQ−LNS N | IFA/SOC N | Prevalence ratio (95% CI) |
|---|---|---|---|
| Underweight (p=0.292) | | | |
| Anemia | 405 | 413 | 1.03 (0.69, 1.53) |
| No anemia | 749 | 654 | 0.79 (0.59, 1.07) |
| Stunted (p=0.477) | | | |
| Anemia | 406 | 413 | 1.00 (0.76, 1.31) |
| No anemia | 749 | 657 | 0.86 (0.69, 1.08) |
| Wasted (p=0.215) | | | |
| Anemia | 405 | 413 | 1.18 (0.52, 2.72) |
| No anemia | 749 | 654 | 0.62 (0.36, 1.06) |
| Low HCAZ (p=0.454) | | | |
| Anemia | 406 | 412 | 1.01 (0.71, 1.45) |
| No anemia | 749 | 656 | 0.85 (0.67, 1.08) |
| Low MUAC (p=0.196) | | | |
| Anemia | 406 | 413 | 1.15 (0.69, 1.91) |
| No anemia | 749 | 657 | 0.76 (0.51, 1.12) |
| Acute malnutrition (p=0.060) | | | |
| Anemia | 405 | 413 | 1.30 (0.80, 2.13) |
| No anemia | 749 | 654 | 0.72 (0.51, 1.02) |

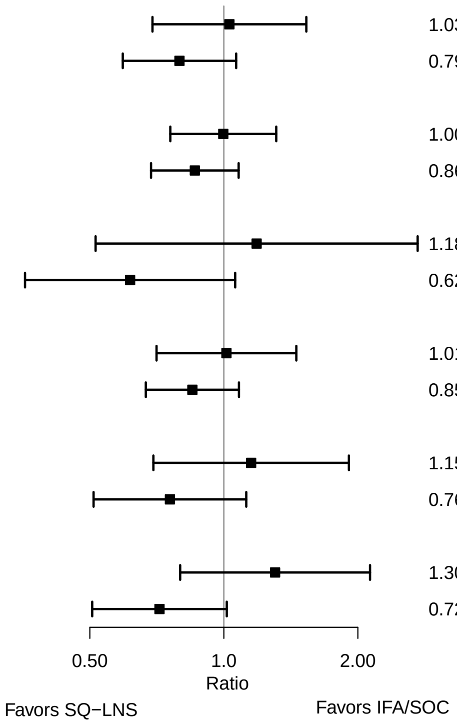

### Supplemental figure 3H: Baseline inflammation status

#### 3H1: Mean differences for birth outcomes

| Outcome (p−for−interaction) | SQ−LNS N | IFA/SOC N | Mean difference (95% CI) |
|---|---|---|---|
| Weight (kg) (p=0.118) | | | |
| Not | 816 | 772 | 0.03 (0.00, 0.07) |
| Inflamed | 401 | 343 | 0.09 (0.03, 0.16) |
| Weight−for−age z score (p=0.091) | | | |
| Not | 816 | 772 | 0.08 (−0.01, 0.17) |
| Inflamed | 401 | 343 | 0.23 (0.08, 0.37) |
| Weight−for−gestational age z−score (p=0.327) | | | |
| Not | 377 | 387 | 0.04 (−0.09, 0.18) |
| Inflamed | 294 | 284 | 0.14 (−0.03, 0.30) |
| Birth length (cm) (p=0.308) | | | |
| Not | 749 | 704 | 0.12 (−0.08, 0.31) |
| Inflamed | 337 | 292 | 0.39 (0.06, 0.72) |
| Length−for−age z−score (p=0.280) | | | |
| Not | 749 | 704 | 0.06 (−0.04, 0.16) |
| Inflamed | 337 | 292 | 0.21 (0.03, 0.38) |
| Length−for−gestational age z−score (p=0.717) | | | |
| Not | 310 | 319 | 0.07 (−0.09, 0.23) |
| Inflamed | 230 | 233 | 0.12 (−0.07, 0.31) |
| BMI z−score (p=0.242) | | | |
| Not | 746 | 702 | 0.09 (−0.01, 0.20) |
| Inflamed | 336 | 286 | 0.20 (0.02, 0.37) |
| Head circumference (cm) (p=0.778) | | | |
| Not | 747 | 705 | 0.13 (0.00, 0.27) |
| Inflamed | 338 | 292 | 0.22 (0.00, 0.45) |
| Head circumference−for−age z−score (p=0.607) | | | |
| Not | 747 | 705 | 0.11 (0.00, 0.21) |
| Inflamed | 338 | 292 | 0.20 (0.01, 0.38) |
| Head circumference−for−gestational age z score (p=0.844) | | | |
| Not | 309 | 320 | 0.15 (−0.01, 0.32) |
| Inflamed | 231 | 233 | 0.12 (−0.07, 0.32) |
| Mid−upper arm circumference (cm) (p=0.189) | | | |
| Not | 749 | 705 | 0.06 (−0.03, 0.14) |
| Inflamed | 339 | 292 | 0.19 (0.04, 0.33) |
| Duration (wk) (p=0.090) | | | |
| Not | 858 | 805 | 0.02 (−0.18, 0.22) |
| Inflamed | 430 | 382 | 0.39 (0.06, 0.72) |

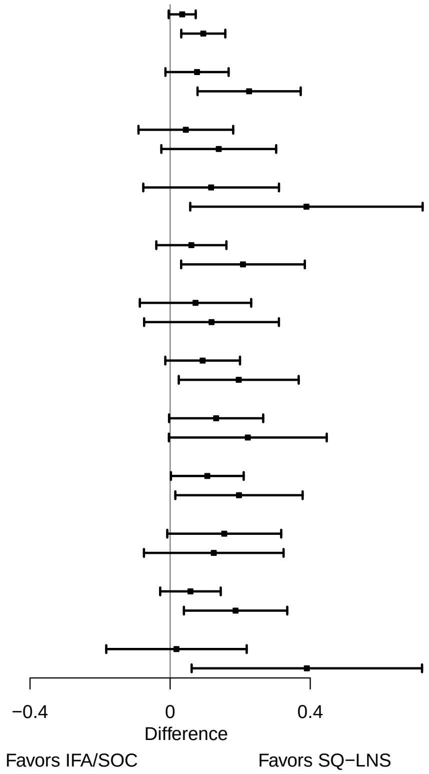

**Supplemental figure 3H: Baseline inflammation status**

#### 3H2: Relative risks for birth outcomes

| Outcome (p−for−interaction) | SQ−LNS N | IFA/SOC N | Relative risk (95% CI) |
|---|---|---|---|
| Low birth weight (p=0.140) | | | |
| Not | 816 | 772 | 0.89 (0.76, 1.05) |
| Inflamed | 401 | 343 | 0.67 (0.49, 0.92) |
| Birth weight < 2 kg (p=0.808) | | | |
| Not | 816 | 772 | 0.91 (0.55, 1.50) |
| Inflamed | 401 | 343 | 0.85 (0.36, 2.01) |
| Small−for−gestational age (p=0.963) | | | |
| Not | 816 | 772 | 0.95 (0.85, 1.07) |
| Inflamed | 401 | 343 | 0.94 (0.77, 1.15) |
| Large−for−gestational age (p=0.176) | | | |
| Not | 384 | 390 | 0.65 (0.30, 1.41) |
| Inflamed | 296 | 286 | 1.43 (0.62, 3.27) |
| Newborn stunting (p=0.189) | | | |
| Not | 749 | 704 | 0.91 (0.73, 1.12) |
| Inflamed | 337 | 292 | 0.69 (0.49, 0.97) |
| Low LGAZ (p=0.808) | | | |
| Not | 749 | 704 | 0.90 (0.71, 1.14) |
| Inflamed | 337 | 292 | 0.87 (0.59, 1.29) |
| Low BMIZ (p=0.324) | | | |
| Not | 746 | 702 | 0.82 (0.69, 0.97) |
| Inflamed | 336 | 286 | 0.67 (0.46, 0.98) |
| Low HCZ (p=0.095) | | | |
| Not | 747 | 705 | 0.91 (0.74, 1.11) |
| Inflamed | 338 | 292 | 0.60 (0.41, 0.89) |
| Low HCGAZ (p=0.700) | | | |
| Not | 747 | 705 | 0.79 (0.59, 1.07) |
| Inflamed | 338 | 292 | 0.91 (0.52, 1.59) |
| Preterm birth (p=0.284) | | | |
| Not | 858 | 805 | 1.09 (0.82, 1.46) |
| Inflamed | 430 | 382 | 0.80 (0.55, 1.15) |

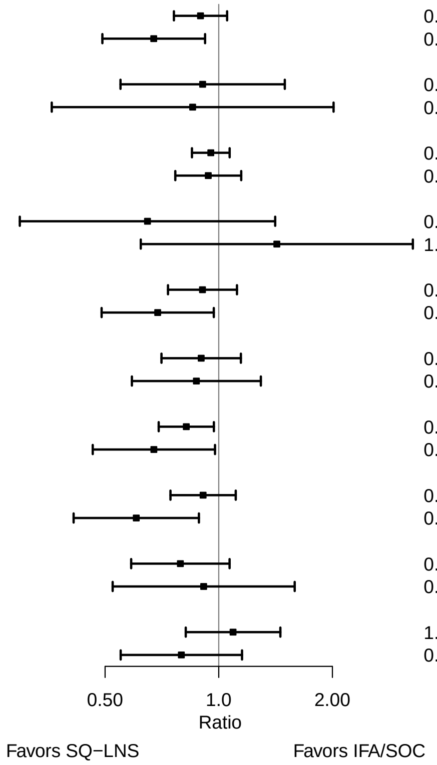

**Supplemental figure 3H: Baseline inflammation status**

#### 3H3: Mean differences for 6 mo outcomes

| Outcome (p−for−interaction) | SQ−LNS N | IFA/SOC N | Mean difference (95% CI) |
|---|---|---|---|
| Weight−for−age z score (p=0.341) | | | |
| Not | 769 | 733 | 0.01 (−0.11, 0.14) |
| Inflamed | 373 | 318 | 0.12 (−0.04, 0.29) |
| Length−for−age z−score (p=0.372) | | | |
| Not | 770 | 735 | 0.03 (−0.08, 0.15) |
| Inflamed | 373 | 319 | 0.13 (−0.03, 0.29) |
| Weight−for−length z−score (p=0.453) | | | |
| Not | 769 | 733 | 0.00 (−0.12, 0.12) |
| Inflamed | 373 | 318 | 0.07 (−0.10, 0.23) |
| Head circumference−for−age z−score (p=0.060) | | | |
| Not | 770 | 734 | −0.02 (−0.11, 0.08) |
| Inflamed | 373 | 318 | 0.13 (−0.02, 0.28) |
| MUAC−for−age z−score (p=0.536) | | | |
| Not | 770 | 735 | 0.01 (−0.10, 0.13) |
| Inflamed | 373 | 319 | 0.08 (−0.07, 0.23) |

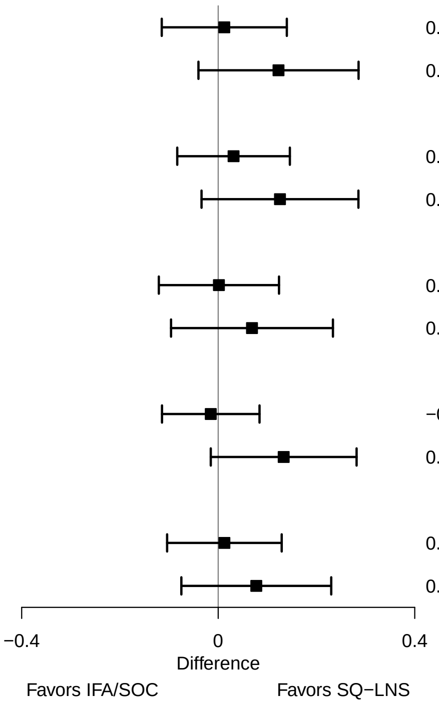

**Supplemental figure 3H: Baseline inflammation status**

#### 3H4: Prevalence ratios for 6 mo outcomes

| Outcome (p−for−interaction) | SQ−LNS N | IFA/SOC N | Prevalence ratio (95% CI) |
|---|---|---|---|
| Underweight (p=0.708) | | | |
| Not | 769 | 733 | 0.85 (0.62, 1.15) |
| Inflamed | 373 | 318 | 0.79 (0.49, 1.27) |
| Stunted (p=0.245) | | | |
| Not | 770 | 735 | 0.93 (0.75, 1.15) |
| Inflamed | 373 | 319 | 0.78 (0.58, 1.05) |
| Wasted (p=0.569) | | | |
| Not | 769 | 733 | 0.79 (0.44, 1.42) |
| Inflamed | 373 | 318 | 0.70 (0.29, 1.69) |
| Low HCAZ (p=0.184) | | | |
| Not | 770 | 734 | 0.94 (0.73, 1.21) |
| Inflamed | 373 | 318 | 0.72 (0.50, 1.02) |
| Low MUAC (p=0.753) | | | |
| Not | 770 | 735 | 0.89 (0.61, 1.30) |
| Inflamed | 373 | 319 | 0.93 (0.48, 1.79) |
| Acute malnutrition (p=0.851) | | | |
| Not | 769 | 733 | 0.90 (0.63, 1.29) |
| Inflamed | 373 | 318 | 0.86 (0.48, 1.52) |

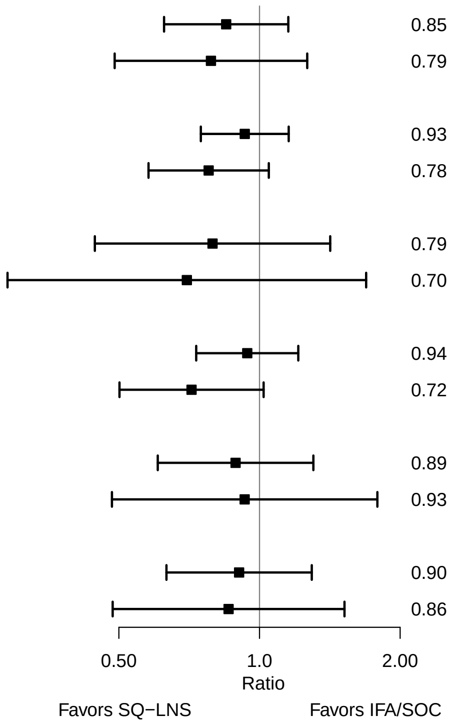

### Supplemental figure 3I: Baseline malaria status

#### 3I1: Mean differences for birth outcomes

| Outcome (p−for−interaction) | SQ−LNS N | IFA/SOC N | Mean difference (95% CI) |
|---|---|---|---|
| Weight (kg) (p=0.053) | | | |
| No malaria | 574 | 582 | 0.05 (−0.01, 0.10) |
| Malaria | 118 | 109 | 0.16 (0.06, 0.27) |
| Weight−for−age z score (p=0.024) | | | |
| No malaria | 574 | 582 | 0.09 (−0.02, 0.21) |
| Malaria | 118 | 109 | 0.41 (0.16, 0.66) |
| Weight−for−gestational age z−score (p=0.268) | | | |
| No malaria | 567 | 576 | 0.07 (−0.04, 0.19) |
| Malaria | 116 | 109 | 0.22 (−0.01, 0.45) |
| Birth length (cm) (p=0.989) | | | |
| No malaria | 470 | 485 | 0.28 (0.03, 0.53) |
| Malaria | 89 | 83 | 0.27 (−0.35, 0.90) |
| Length−for−age z−score (p=0.794) | | | |
| No malaria | 470 | 485 | 0.14 (0.01, 0.27) |
| Malaria | 89 | 83 | 0.19 (−0.15, 0.52) |
| Length−for−gestational age z−score (p=0.715) | | | |
| No malaria | 463 | 479 | 0.11 (−0.02, 0.24) |
| Malaria | 87 | 83 | 0.08 (−0.22, 0.38) |
| BMI z−score (p=0.235) | | | |
| No malaria | 467 | 481 | 0.10 (−0.04, 0.24) |
| Malaria | 88 | 79 | 0.32 (0.02, 0.62) |
| Head circumference (cm) (p=0.977) | | | |
| No malaria | 471 | 486 | 0.21 (0.03, 0.39) |
| Malaria | 88 | 83 | 0.17 (−0.26, 0.59) |
| Head circumference−for−age z−score (p=0.703) | | | |
| No malaria | 471 | 486 | 0.16 (0.02, 0.30) |
| Malaria | 88 | 83 | 0.19 (−0.16, 0.55) |
| Head circumference−for−gestational age z score (p=0.969) | | | |
| No malaria | 464 | 480 | 0.13 (0.00, 0.27) |
| Malaria | 86 | 83 | 0.10 (−0.23, 0.42) |
| Mid−upper arm circumference (cm) (p=0.269) | | | |
| No malaria | 473 | 486 | 0.11 (0.00, 0.23) |
| Malaria | 89 | 83 | 0.27 (0.00, 0.54) |
| Duration (wk) (p=0.761) | | | |
| No malaria | 629 | 638 | 0.14 (−0.12, 0.40) |
| Malaria | 136 | 127 | 0.18 (−0.45, 0.82) |

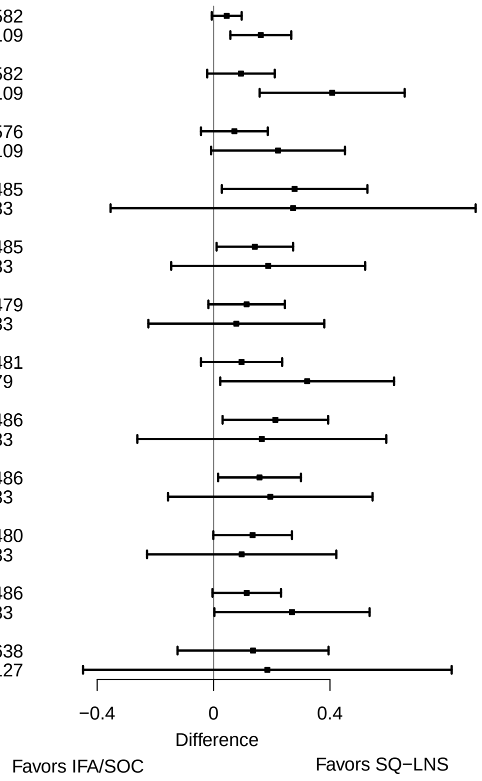

**Supplemental figure 3I: Baseline malaria status**

#### 3I2: Relative risks for birth outcomes

| Outcome (p−for−interaction) | SQ−LNS N | IFA/SOC N | Relative risk (95% CI) |
|---|---|---|---|
| Low birth weight (p=0.097) | | | |
| No malaria | 574 | 582 | 0.88 (0.64, 1.21) |
| Malaria | 118 | 109 | 0.53 (0.28, 0.98) |
| Small−for−gestational age (p=0.016) | | | |
| No malaria | 574 | 582 | 1.01 (0.81, 1.26) |
| Malaria | 118 | 109 | 0.60 (0.41, 0.89) |
| Newborn stunting (p=0.673) | | | |
| No malaria | 470 | 485 | 0.88 (0.61, 1.28) |
| Malaria | 89 | 83 | 0.96 (0.56, 1.64) |
| Low LGAZ (p=0.780) | | | |
| No malaria | 470 | 485 | 0.96 (0.64, 1.44) |
| Malaria | 89 | 83 | 0.98 (0.55, 1.76) |
| Low HCZ (p=0.254) | | | |
| No malaria | 471 | 486 | 0.93 (0.57, 1.52) |
| Malaria | 88 | 83 | 0.40 (0.10, 1.61) |
| Preterm birth (p=0.271) | | | |
| No malaria | 629 | 638 | 0.98 (0.71, 1.36) |
| Malaria | 136 | 127 | 0.61 (0.30, 1.25) |

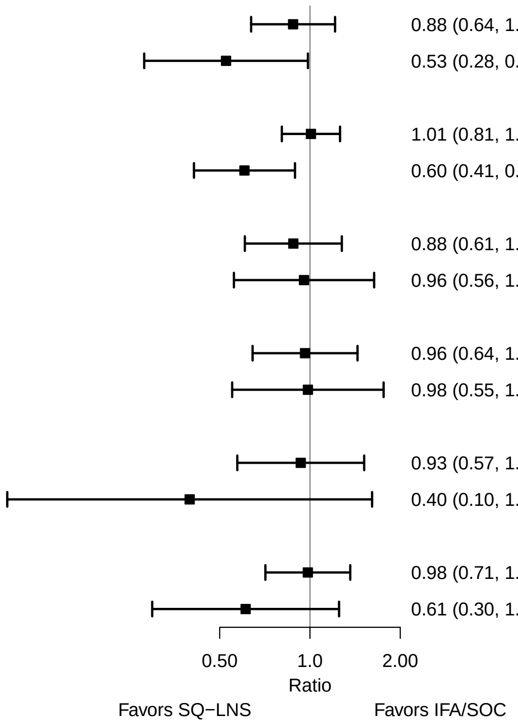

**Supplemental figure 3I: Baseline malaria status**

#### 3I3: Mean differences for 6 mo outcomes

| Outcome (p−for−interaction) | SQ−LNS N | IFA/SOC N | Mean difference (95% CI) |
|---|---|---|---|
| Weight−for−age z score (p=0.092) | | | |
| No malaria | 523 | 534 | −0.01 (−0.14, 0.13) |
| Malaria | 106 | 102 | 0.27 (−0.02, 0.57) |
| Length−for−age z−score (p=0.133) | | | |
| No malaria | 524 | 537 | 0.02 (−0.11, 0.15) |
| Malaria | 106 | 102 | 0.24 (−0.04, 0.53) |
| Weight−for−length z−score (p=0.379) | | | |
| No malaria | 523 | 534 | −0.02 (−0.16, 0.12) |
| Malaria | 106 | 102 | 0.16 (−0.15, 0.47) |
| Head circumference−for−age z−score (p=0.210) | | | |
| No malaria | 524 | 535 | −0.01 (−0.13, 0.10) |
| Malaria | 106 | 102 | 0.14 (−0.13, 0.40) |
| MUAC−for−age z−score (p=0.540) | | | |
| No malaria | 524 | 537 | 0.02 (−0.11, 0.15) |
| Malaria | 106 | 102 | 0.13 (−0.13, 0.40) |

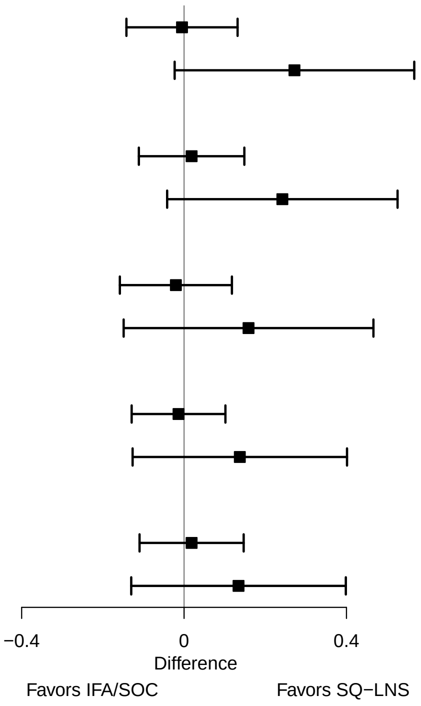

**Supplemental figure 3I: Baseline malaria status**

#### 3I4: Prevalence ratios for 6 mo outcomes

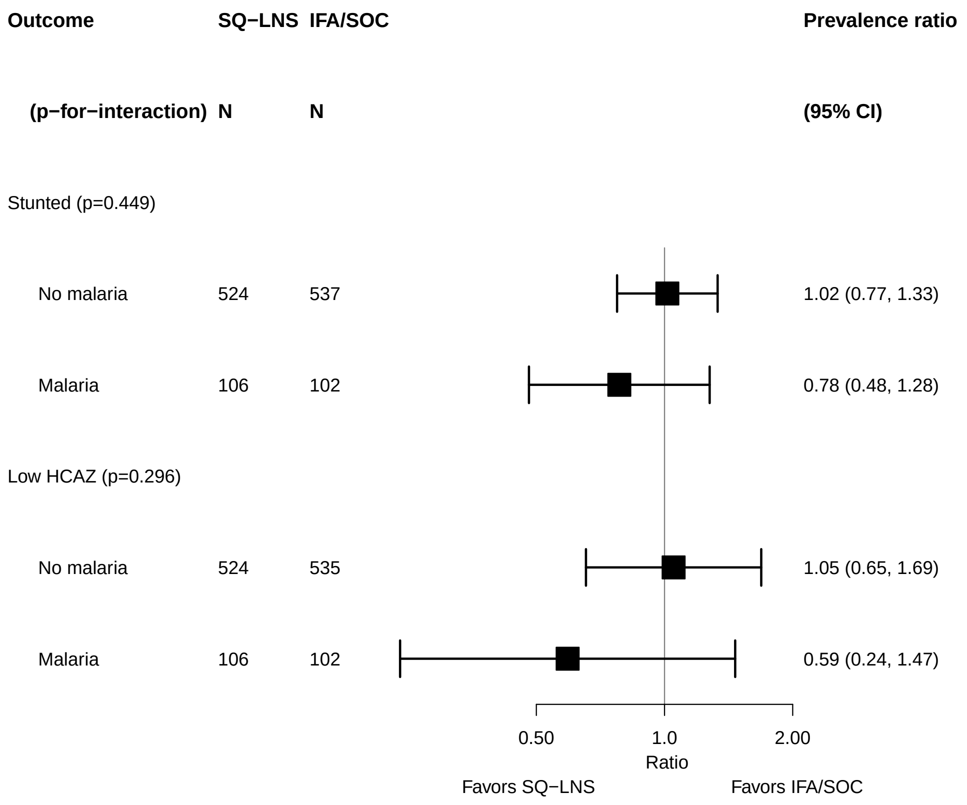

### Supplemental figure 3J: Gestational age at supplementation

#### 3J1: Mean differences for birth outcomes

| Outcome (p−for−interaction) | SQ−LNS N | IFA/SOC N | Mean difference (95% CI) |
|---|---|---|---|
| Weight (kg) (p=0.283) | | | |
| < 14 wks | 851 | 1847 | 0.04 (0.01, 0.07) |
| >= 14 wks | 949 | 1586 | 0.06 (0.03, 0.10) |
| Weight−for−age z score (p=0.316) | | | |
| < 14 wks | 851 | 1847 | 0.10 (0.03, 0.17) |
| >= 14 wks | 949 | 1586 | 0.15 (0.07, 0.23) |
| Weight−for−gestational age z−score (p=0.900) | | | |
| < 14 wks | 115 | 117 | 0.13 (−0.12, 0.37) |
| >= 14 wks | 570 | 568 | 0.08 (−0.03, 0.20) |
| Birth length (cm) (p=0.226) | | | |
| < 14 wks | 836 | 1840 | 0.13 (0.00, 0.27) |
| >= 14 wks | 829 | 1469 | 0.30 (0.11, 0.49) |
| Length−for−age z−score (p=0.182) | | | |
| < 14 wks | 836 | 1840 | 0.08 (0.00, 0.15) |
| >= 14 wks | 829 | 1469 | 0.16 (0.06, 0.26) |
| Length−for−gestational age z−score (p=0.787) | | | |
| < 14 wks | 100 | 111 | 0.13 (−0.15, 0.41) |
| >= 14 wks | 451 | 451 | 0.09 (−0.05, 0.22) |
| BMI z−score (p=0.620) | | | |
| < 14 wks | 836 | 1838 | 0.09 (0.01, 0.18) |
| >= 14 wks | 825 | 1463 | 0.12 (0.03, 0.21) |
| Head circumference (cm) (p=0.213) | | | |
| < 14 wks | 835 | 1841 | 0.06 (−0.04, 0.15) |
| >= 14 wks | 830 | 1470 | 0.20 (0.07, 0.33) |
| Head circumference−for−age z−score (p=0.192) | | | |
| < 14 wks | 835 | 1841 | 0.06 (−0.02, 0.13) |
| >= 14 wks | 830 | 1470 | 0.17 (0.06, 0.27) |
| Head circumference−for−gestational age z score (p=0.773) | | | |
| < 14 wks | 99 | 111 | 0.09 (−0.20, 0.39) |
| >= 14 wks | 452 | 452 | 0.13 (−0.01, 0.27) |
| Mid−upper arm circumference (cm) (p=0.262) | | | |
| < 14 wks | 655 | 1673 | 0.04 (−0.04, 0.11) |
| >= 14 wks | 806 | 1447 | 0.13 (0.04, 0.21) |
| Duration (wk) (p=0.308) | | | |
| < 14 wks | 685 | 1689 | 0.07 (−0.10, 0.24) |
| >= 14 wks | 980 | 1627 | 0.21 (0.01, 0.41) |

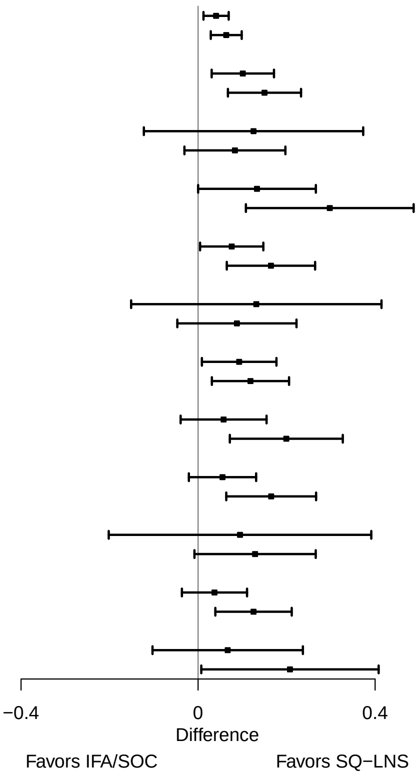

**Supplemental figure 3J: Gestational age at supplementation**

#### 3J2: Relative risks for birth outcomes

| Outcome (p−for−interaction) | SQ−LNS N | IFA/SOC N | Relative risk (95% CI) |
|---|---|---|---|
| Low birth weight (p=0.802) | | | |
| < 14 wks | 851 | 1847 | 0.90 (0.80, 1.00) |
| >= 14 wks | 949 | 1586 | 0.89 (0.76, 1.05) |
| Birth weight < 2 kg (p=0.860) | | | |
| < 14 wks | 602 | 1606 | 0.77 (0.55, 1.09) |
| >= 14 wks | 683 | 1331 | 0.86 (0.55, 1.36) |
| Small−for−gestational age (p=0.993) | | | |
| < 14 wks | 670 | 1679 | 0.97 (0.90, 1.05) |
| >= 14 wks | 922 | 1563 | 0.96 (0.88, 1.04) |
| Large−for−gestational age (p=0.060) | | | |
| < 14 wks | 618 | 1632 | 0.57 (0.17, 1.93) |
| >= 14 wks | 587 | 1224 | 1.60 (0.70, 3.69) |
| Newborn stunting (p=0.485) | | | |
| < 14 wks | 836 | 1840 | 0.86 (0.75, 0.97) |
| >= 14 wks | 829 | 1469 | 0.81 (0.65, 1.01) |
| Low LGAZ (p=0.811) | | | |
| < 14 wks | 655 | 1672 | 0.90 (0.78, 1.04) |
| >= 14 wks | 802 | 1446 | 0.90 (0.74, 1.10) |
| Low BMIZ (p=0.806) | | | |
| < 14 wks | 836 | 1838 | 0.89 (0.78, 1.02) |
| >= 14 wks | 825 | 1463 | 0.91 (0.80, 1.03) |
| Low HCZ (p=0.923) | | | |
| < 14 wks | 835 | 1841 | 0.86 (0.72, 1.04) |
| >= 14 wks | 830 | 1470 | 0.83 (0.67, 1.05) |
| Low HCGAZ (p=0.505) | | | |
| < 14 wks | 618 | 1632 | 0.83 (0.65, 1.07) |
| >= 14 wks | 586 | 1224 | 0.94 (0.73, 1.20) |
| Preterm birth (p=0.442) | | | |
| < 14 wks | 685 | 1689 | 0.88 (0.70, 1.11) |
| >= 14 wks | 980 | 1627 | 0.97 (0.77, 1.23) |

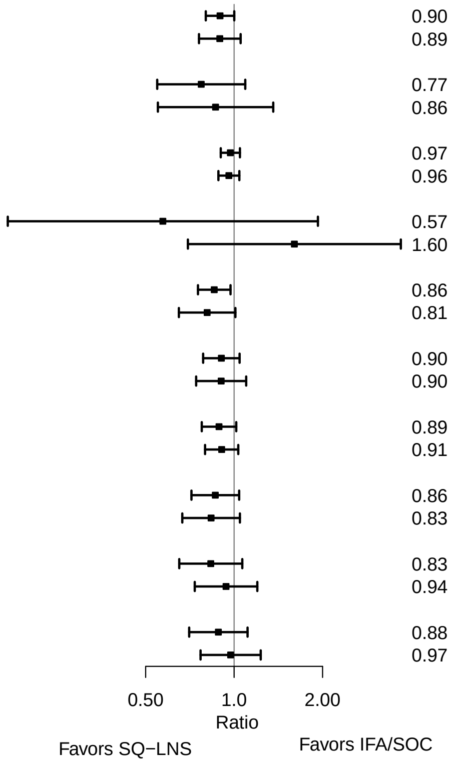

**Supplemental figure 3J: Gestational age at supplementation**

#### 3J3: Mean differences for 6 mo outcomes

| Outcome (p−for−interaction) | SQ−LNS N | IFA/SOC N | Mean difference (95% CI) |
|---|---|---|---|
| Weight−for−age z score (p=0.756) | | | |
| < 14 wks | 830 | 1790 | 0.04 (−0.04, 0.13) |
| >= 14 wks | 867 | 1492 | 0.03 (−0.08, 0.14) |
| Length−for−age z−score (p=0.780) | | | |
| < 14 wks | 830 | 1790 | 0.07 (−0.02, 0.16) |
| >= 14 wks | 868 | 1494 | 0.05 (−0.05, 0.15) |
| Weight−for−length z−score (p=0.712) | | | |
| < 14 wks | 830 | 1790 | −0.01 (−0.10, 0.07) |
| >= 14 wks | 867 | 1491 | 0.00 (−0.11, 0.11) |
| Head circumference−for−age z−score (p=0.513) | | | |
| < 14 wks | 830 | 1787 | −0.01 (−0.09, 0.07) |
| >= 14 wks | 868 | 1493 | 0.02 (−0.07, 0.11) |
| MUAC−for−age z−score (p=0.939) | | | |
| < 14 wks | 653 | 1625 | −0.02 (−0.11, 0.08) |
| >= 14 wks | 840 | 1471 | 0.01 (−0.09, 0.11) |

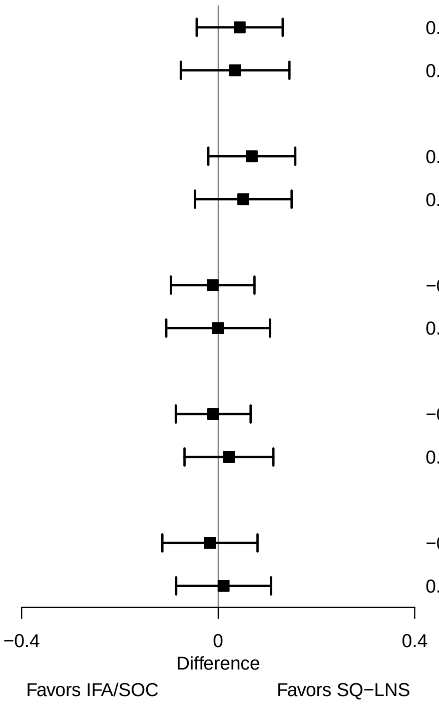

**Supplemental figure 3J: Gestational age at supplementation**

#### 3J4: Prevalence ratios for 6 mo outcomes

| Outcome (p−for−interaction) | SQ−LNS N | IFA/SOC N | Prevalence ratio (95% CI) |
|---|---|---|---|
| Underweight (p=0.983) | | | |
| < 14 wks | 830 | 1790 | 0.90 (0.76, 1.07) |
| >= 14 wks | 867 | 1492 | 0.96 (0.72, 1.26) |
| Stunted (p=0.131) | | | |
| < 14 wks | 830 | 1790 | 0.83 (0.72, 0.96) |
| >= 14 wks | 868 | 1494 | 1.02 (0.84, 1.25) |
| Wasted (p=0.731) | | | |
| < 14 wks | 605 | 1580 | 1.03 (0.68, 1.55) |
| >= 14 wks | 537 | 1148 | 0.75 (0.44, 1.29) |
| Low HCAZ (p=0.911) | | | |
| < 14 wks | 782 | 1743 | 0.89 (0.75, 1.05) |
| >= 14 wks | 566 | 1174 | 0.90 (0.73, 1.12) |
| Low MUAC (p=0.010) | | | |
| < 14 wks | 653 | 1625 | 1.36 (0.97, 1.91) |
| >= 14 wks | 840 | 1471 | 0.76 (0.55, 1.07) |
| Acute malnutrition (p=0.094) | | | |
| < 14 wks | 653 | 1625 | 1.20 (0.88, 1.63) |
| >= 14 wks | 839 | 1467 | 0.83 (0.61, 1.13) |

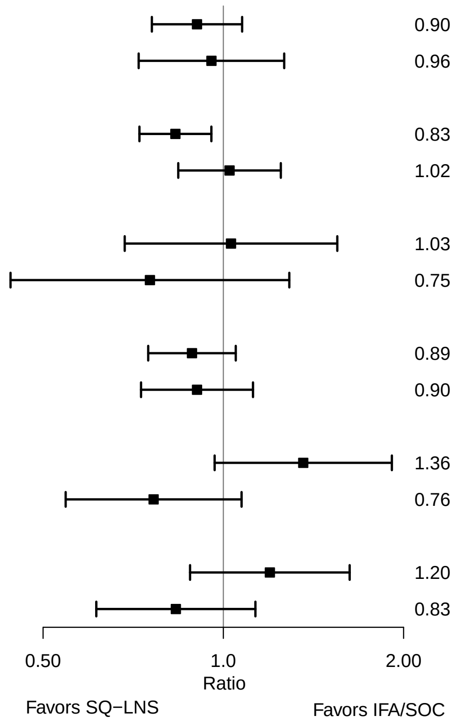

### Supplemental figure 3K: Compliance with supplementation

#### 3K1: Mean differences for birth outcomes

| Outcome (p−for−interaction) | SQ−LNS N | IFA/SOC N | Mean difference (95% CI) |
|---|---|---|---|
| Weight (kg) (p=0.105) | | | |
| >= 4/7 | 1184 | 2961 | 0.07 (0.04, 0.10) |
| Less than 4/7 | 400 | 249 | 0.00 (−0.07, 0.07) |
| Weight−for−age z score (p=0.123) | | | |
| >= 4/7 | 1184 | 2961 | 0.16 (0.08, 0.23) |
| Less than 4/7 | 400 | 249 | 0.01 (−0.17, 0.19) |
| Weight−for−gestational age z−score (p=0.949) | | | |
| >= 4/7 | 602 | 626 | 0.09 (−0.02, 0.20) |
| Less than 4/7 | 80 | 56 | 0.13 (−0.23, 0.48) |
| Birth length (cm) (p=0.113) | | | |
| >= 4/7 | 1063 | 2847 | 0.27 (0.13, 0.42) |
| Less than 4/7 | 389 | 241 | −0.04 (−0.38, 0.30) |
| Length−for−age z−score (p=0.115) | | | |
| >= 4/7 | 1063 | 2847 | 0.15 (0.07, 0.23) |
| Less than 4/7 | 389 | 241 | −0.02 (−0.20, 0.16) |
| Length−for−gestational age z−score (p=0.296) | | | |
| >= 4/7 | 482 | 513 | 0.12 (−0.01, 0.25) |
| Less than 4/7 | 69 | 48 | −0.10 (−0.47, 0.28) |
| BMI z−score (p=0.590) | | | |
| >= 4/7 | 1059 | 2840 | 0.12 (0.04, 0.20) |
| Less than 4/7 | 389 | 240 | 0.07 (−0.11, 0.25) |
| Head circumference (cm) (p=0.534) | | | |
| >= 4/7 | 1063 | 2849 | 0.17 (0.07, 0.28) |
| Less than 4/7 | 389 | 241 | 0.06 (−0.17, 0.30) |
| Head circumference−for−age z−score (p=0.466) | | | |
| >= 4/7 | 1063 | 2849 | 0.15 (0.06, 0.23) |
| Less than 4/7 | 389 | 241 | 0.05 (−0.13, 0.23) |
| Head circumference−for−gestational age z score (p=0.860) | | | |
| >= 4/7 | 482 | 514 | 0.13 (0.00, 0.26) |
| Less than 4/7 | 69 | 48 | 0.17 (−0.28, 0.62) |
| Mid−upper arm circumference (cm) (p=0.508) | | | |
| >= 4/7 | 1067 | 2849 | 0.11 (0.03, 0.18) |
| Less than 4/7 | 389 | 241 | 0.04 (−0.10, 0.19) |
| Duration (wk) (p=0.903) | | | |
| >= 4/7 | 1235 | 3019 | 0.12 (−0.05, 0.30) |
| Less than 4/7 | 421 | 264 | 0.12 (−0.22, 0.46) |

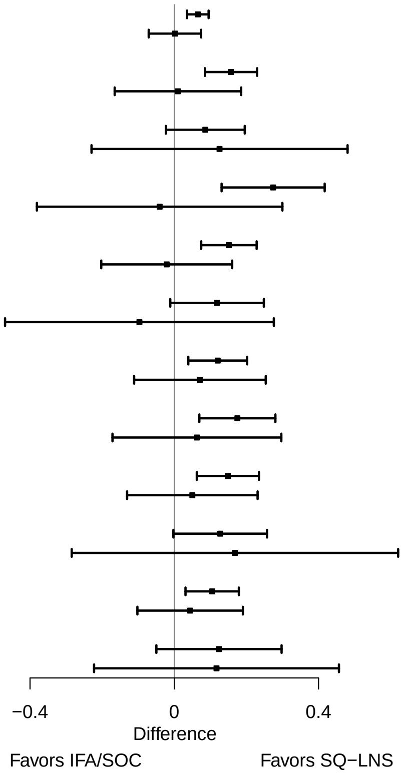

**Supplemental figure 3K: Compliance with supplementation**

#### 3K2: Relative risks for birth outcomes

| Outcome (p−for−interaction) | SQ−LNS N | IFA/SOC N | Relative risk (95% CI) |
|---|---|---|---|
| Low birth weight (p=0.364) | | | |
| >= 4/7 | 1184 | 2961 | 0.84 (0.74, 0.96) |
| Less than 4/7 | 400 | 249 | 0.96 (0.75, 1.23) |
| Small−for−gestational age (p=0.540) | | | |
| >= 4/7 | 1184 | 2961 | 0.94 (0.87, 1.01) |
| Less than 4/7 | 400 | 249 | 1.00 (0.85, 1.18) |
| Newborn stunting (p=0.081) | | | |
| >= 4/7 | 1063 | 2847 | 0.78 (0.66, 0.92) |
| Less than 4/7 | 389 | 241 | 1.08 (0.79, 1.48) |
| Low LGAZ (p=0.743) | | | |
| >= 4/7 | 1063 | 2847 | 0.85 (0.72, 1.01) |
| Less than 4/7 | 389 | 241 | 0.90 (0.63, 1.27) |
| Low BMIZ (p=0.565) | | | |
| >= 4/7 | 1059 | 2840 | 0.86 (0.76, 0.97) |
| Less than 4/7 | 389 | 240 | 0.91 (0.73, 1.15) |
| Low HCZ (p=0.909) | | | |
| >= 4/7 | 1063 | 2849 | 0.82 (0.69, 0.99) |
| Less than 4/7 | 389 | 241 | 0.84 (0.65, 1.11) |
| Low HCGAZ (p=0.613) | | | |
| >= 4/7 | 821 | 2601 | 0.84 (0.67, 1.07) |
| Less than 4/7 | 378 | 226 | 0.76 (0.54, 1.06) |
| Preterm birth (p=0.845) | | | |
| >= 4/7 | 1235 | 3019 | 0.93 (0.75, 1.15) |
| Less than 4/7 | 421 | 264 | 0.98 (0.67, 1.43) |

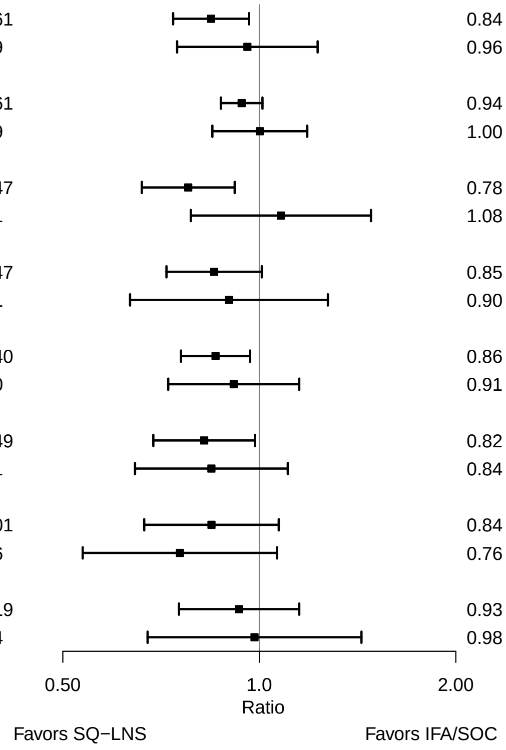

**Supplemental figure 3K: Compliance with supplementation**

#### 3K3: Mean differences for 6 mo outcomes

| Outcome (p−for−interaction) | SQ−LNS N | IFA/SOC N | Mean difference (95% CI) |
|---|---|---|---|
| Weight−for−age z score (p=0.937) | | | |
| >= 4/7 | 1111 | 2830 | 0.01 (−0.07, 0.09) |
| Less than 4/7 | 378 | 244 | 0.01 (−0.19, 0.21) |
| Length−for−age z−score (p=0.147) | | | |
| >= 4/7 | 1112 | 2833 | 0.07 (−0.01, 0.15) |
| Less than 4/7 | 378 | 243 | −0.05 (−0.22, 0.11) |
| Weight−for−length z−score (p=0.180) | | | |
| >= 4/7 | 1111 | 2830 | −0.06 (−0.13, 0.02) |
| Less than 4/7 | 378 | 243 | 0.04 (−0.14, 0.23) |
| Head circumference−for−age z−score (p=0.291) | | | |
| >= 4/7 | 1112 | 2831 | 0.03 (−0.04, 0.10) |
| Less than 4/7 | 378 | 244 | −0.05 (−0.19, 0.10) |
| MUAC−for−age z−score (p=0.916) | | | |
| >= 4/7 | 1112 | 2833 | 0.00 (−0.08, 0.07) |
| Less than 4/7 | 378 | 244 | −0.02 (−0.20, 0.16) |

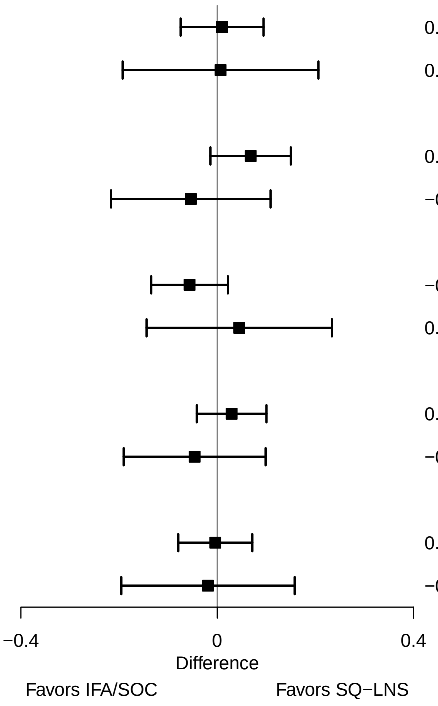

**Supplemental figure 3K: Compliance with supplementation**

#### 3K4: Prevalence ratios for 6 mo outcomes

| Outcome (p−for−interaction) | SQ−LNS N | IFA/SOC N | Prevalence ratio (95% CI) |
|---|---|---|---|
| Underweight (p=0.635) | | | |
| >= 4/7 | 1111 | 2830 | 0.99 (0.83, 1.19) |
| Less than 4/7 | 378 | 244 | 0.89 (0.57, 1.40) |
| Stunted (p=0.190) | | | |
| >= 4/7 | 1112 | 2833 | 0.90 (0.76, 1.06) |
| Less than 4/7 | 378 | 243 | 1.10 (0.80, 1.51) |
| Low HCAZ (p=0.370) | | | |
| >= 4/7 | 1112 | 2831 | 0.86 (0.74, 0.99) |
| Less than 4/7 | 378 | 244 | 0.99 (0.73, 1.34) |
| Low MUAC (p=0.512) | | | |
| >= 4/7 | 1112 | 2833 | 1.04 (0.80, 1.36) |
| Less than 4/7 | 378 | 244 | 0.92 (0.49, 1.76) |
| Acute malnutrition (p=0.166) | | | |
| >= 4/7 | 1111 | 2830 | 1.07 (0.85, 1.33) |
| Less than 4/7 | 378 | 243 | 0.73 (0.41, 1.32) |

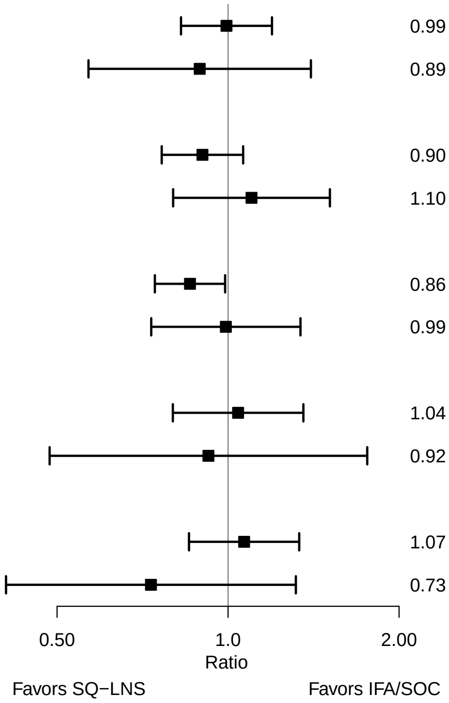

### Supplemental figure 3L: Household socio-economic status

#### 3L1: Mean differences for birth outcomes

| Outcome (p−for−interaction) | SQ−LNS N | IFA/SOC N | Mean difference (95% CI) |
|---|---|---|---|
| Weight (kg) (p=0.909) | | | |
| At least median | 874 | 1681 | 0.05 (0.02, 0.08) |
| Less than median | 914 | 1739 | 0.05 (0.01, 0.09) |
| Weight−for−age z score (p=0.933) | | | |
| At least median | 874 | 1681 | 0.12 (0.04, 0.20) |
| Less than median | 914 | 1739 | 0.12 (0.03, 0.21) |
| Weight−for−gestational age z−score (p=0.280) | | | |
| At least median | 383 | 389 | 0.18 (0.05, 0.30) |
| Less than median | 463 | 431 | 0.05 (−0.08, 0.17) |
| Birth length (cm) (p=0.829) | | | |
| At least median | 816 | 1626 | 0.19 (0.03, 0.36) |
| Less than median | 838 | 1673 | 0.19 (0.00, 0.38) |
| Length−for−age z−score (p=0.951) | | | |
| At least median | 816 | 1626 | 0.11 (0.02, 0.19) |
| Less than median | 838 | 1673 | 0.12 (0.01, 0.22) |
| Length−for−gestational age z−score (p=0.347) | | | |
| At least median | 326 | 335 | 0.18 (0.04, 0.31) |
| Less than median | 387 | 365 | 0.06 (−0.07, 0.19) |
| BMI z−score (p=0.952) | | | |
| At least median | 814 | 1625 | 0.09 (0.00, 0.17) |
| Less than median | 836 | 1666 | 0.10 (0.00, 0.19) |
| Head circumference (cm) (p=0.565) | | | |
| At least median | 814 | 1628 | 0.12 (0.02, 0.22) |
| Less than median | 840 | 1673 | 0.11 (−0.03, 0.24) |
| Head circumference−for−age z−score (p=0.706) | | | |
| At least median | 814 | 1628 | 0.12 (0.04, 0.19) |
| Less than median | 840 | 1673 | 0.11 (0.00, 0.21) |
| Head circumference−for−gestational age z score (p=0.143) | | | |
| At least median | 324 | 336 | 0.21 (0.08, 0.35) |
| Less than median | 389 | 365 | 0.01 (−0.13, 0.15) |
| Mid−upper arm circumference (cm) (p=0.560) | | | |
| At least median | 717 | 1540 | 0.06 (−0.02, 0.14) |
| Less than median | 742 | 1576 | 0.10 (0.01, 0.18) |
| Duration (wk) (p=0.345) | | | |
| At least median | 885 | 1708 | 0.16 (0.01, 0.31) |
| Less than median | 940 | 1746 | 0.07 (−0.10, 0.24) |

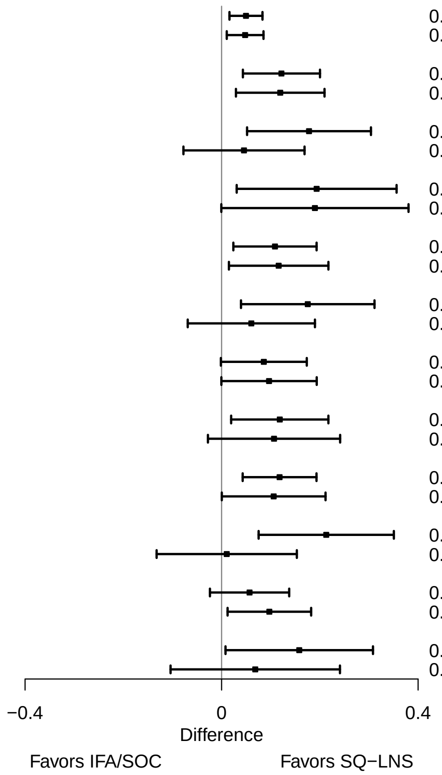

**Supplemental figure 3L: Household socio-economic status**

#### 3L2: Relative risks for birth outcomes

| Outcome (p−for−interaction) | SQ−LNS N | IFA/SOC N | Relative risk (95% CI) |
|---|---|---|---|
| Low birth weight (p=0.895) | | | |
| At least median | 874 | 1681 | 0.92 (0.79, 1.07) |
| Less than median | 914 | 1739 | 0.90 (0.78, 1.03) |
| Birth weight < 2 kg (p=0.161) | | | |
| At least median | 774 | 1593 | 0.98 (0.68, 1.42) |
| Less than median | 815 | 1642 | 0.64 (0.42, 0.98) |
| Small−for−gestational age (p=0.012) | | | |
| At least median | 854 | 1666 | 1.04 (0.96, 1.12) |
| Less than median | 899 | 1711 | 0.90 (0.83, 0.97) |
| Large−for−gestational age (p=0.669) | | | |
| At least median | 774 | 1593 | 0.89 (0.44, 1.78) |
| Less than median | 815 | 1642 | 1.10 (0.53, 2.29) |
| Newborn stunting (p=0.789) | | | |
| At least median | 816 | 1626 | 0.85 (0.70, 1.02) |
| Less than median | 838 | 1673 | 0.85 (0.71, 1.03) |
| Low LGAZ (p=0.763) | | | |
| At least median | 796 | 1611 | 0.88 (0.73, 1.06) |
| Less than median | 823 | 1645 | 0.93 (0.77, 1.12) |
| Low BMIZ (p=0.882) | | | |
| At least median | 814 | 1625 | 0.91 (0.78, 1.06) |
| Less than median | 836 | 1666 | 0.90 (0.77, 1.04) |
| Low HCZ (p=0.641) | | | |
| At least median | 814 | 1628 | 0.89 (0.76, 1.04) |
| Less than median | 840 | 1673 | 0.83 (0.69, 1.01) |
| Low HCGAZ (p=0.018) | | | |
| At least median | 548 | 1348 | 1.08 (0.89, 1.32) |
| Less than median | 513 | 1345 | 0.74 (0.59, 0.93) |
| Preterm birth (p=0.759) | | | |
| At least median | 885 | 1708 | 0.96 (0.76, 1.23) |
| Less than median | 940 | 1746 | 0.92 (0.74, 1.14) |

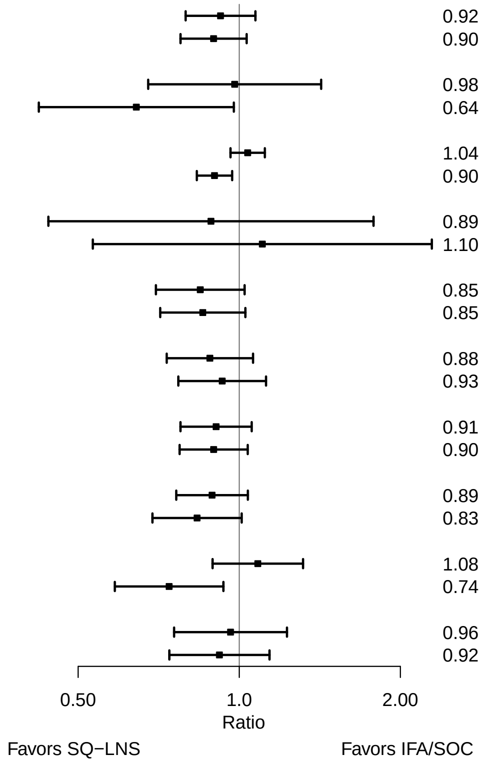

Supplemental figure 3L: Household socio-economic status

#### 3L3: Mean differences for 6 mo outcomes

| Outcome (p−for−interaction) | SQ−LNS N | IFA/SOC N | Mean difference (95% CI) |
|---|---|---|---|
| Weight−for−age z score (p=0.706) | | | |
| At least median | 821 | 1616 | 0.03 (−0.07, 0.13) |
| Less than median | 874 | 1671 | 0.04 (−0.06, 0.14) |
| Length−for−age z−score (p=0.332) | | | |
| At least median | 822 | 1616 | 0.01 (−0.08, 0.10) |
| Less than median | 874 | 1673 | 0.07 (−0.01, 0.16) |
| Weight−for−length z−score (p=0.497) | | | |
| At least median | 821 | 1615 | 0.04 (−0.04, 0.12) |
| Less than median | 874 | 1671 | −0.02 (−0.11, 0.08) |
| Head circumference−for−age z−score (p=0.637) | | | |
| At least median | 822 | 1614 | 0.02 (−0.06, 0.09) |
| Less than median | 874 | 1671 | −0.02 (−0.10, 0.07) |
| MUAC−for−age z−score (p=0.318) | | | |
| At least median | 720 | 1524 | −0.04 (−0.13, 0.05) |
| Less than median | 775 | 1575 | 0.02 (−0.07, 0.11) |

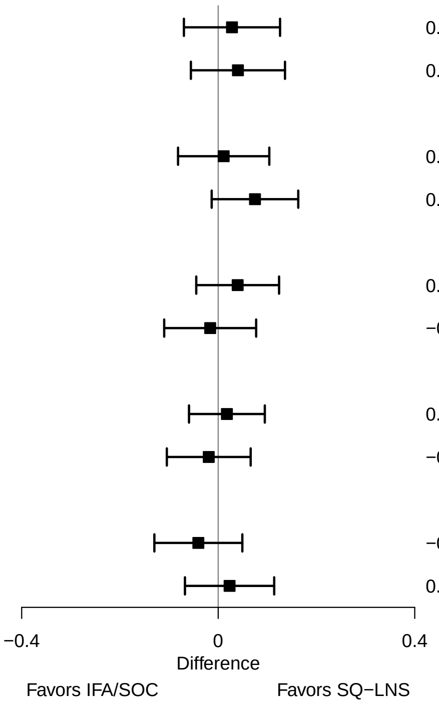

**Supplemental figure 3L: Household socio-economic status**

#### 3L4: Prevalence ratios for 6 mo outcomes

| Outcome (p−for−interaction) | SQ−LNS N | IFA/SOC N | Prevalence ratio (95% CI) |
|---|---|---|---|
| Underweight (p=0.498) | | | |
| At least median | 821 | 1616 | 0.96 (0.73, 1.26) |
| Less than median | 874 | 1671 | 0.89 (0.73, 1.09) |
| Stunted (p=0.664) | | | |
| At least median | 822 | 1616 | 0.94 (0.79, 1.13) |
| Less than median | 874 | 1673 | 0.90 (0.77, 1.04) |
| Wasted (p=0.992) | | | |
| At least median | 719 | 1522 | 0.91 (0.56, 1.49) |
| Less than median | 775 | 1573 | 0.98 (0.67, 1.44) |
| Low HCAZ (p=0.757) | | | |
| At least median | 822 | 1614 | 0.88 (0.74, 1.06) |
| Less than median | 874 | 1671 | 0.92 (0.76, 1.10) |
| Low MUAC (p=0.725) | | | |
| At least median | 720 | 1524 | 1.07 (0.74, 1.54) |
| Less than median | 775 | 1575 | 1.00 (0.75, 1.35) |
| Acute malnutrition (p=0.811) | | | |
| At least median | 719 | 1522 | 0.97 (0.69, 1.36) |
| Less than median | 775 | 1573 | 1.03 (0.80, 1.34) |

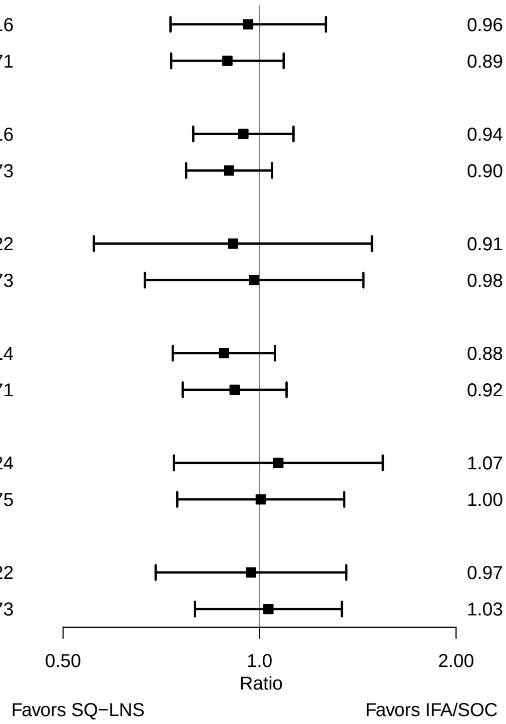

### Supplemental figure 3M: Household food security

#### 3M1: Mean differences for birth outcomes

| Outcome (p−for−interaction) | SQ−LNS N | IFA/SOC N | Mean difference (95% CI) |
|---|---|---|---|
| Weight (kg) (p=0.103) | | | |
| Mild to secure | 945 | 1884 | 0.03 (0.00, 0.06) |
| Moderate to severe | 643 | 1352 | 0.07 (0.03, 0.11) |
| Weight−for−age z score (p=0.135) | | | |
| Mild to secure | 945 | 1884 | 0.08 (0.01, 0.15) |
| Moderate to severe | 643 | 1352 | 0.16 (0.06, 0.25) |
| Weight−for−gestational age z−score (p=0.526) | | | |
| Mild to secure | 347 | 326 | 0.14 (−0.01, 0.29) |
| Moderate to severe | 334 | 353 | 0.03 (−0.12, 0.18) |
| Birth length (cm) (p=0.024) | | | |
| Mild to secure | 897 | 1844 | 0.09 (−0.05, 0.23) |
| Moderate to severe | 556 | 1271 | 0.34 (0.15, 0.52) |
| Length−for−age z−score (p=0.047) | | | |
| Mild to secure | 897 | 1844 | 0.06 (−0.02, 0.13) |
| Moderate to severe | 556 | 1271 | 0.17 (0.08, 0.27) |
| Length−for−gestational age z−score (p=0.700) | | | |
| Mild to secure | 300 | 287 | 0.12 (−0.05, 0.28) |
| Moderate to severe | 247 | 272 | 0.07 (−0.12, 0.25) |
| BMI z−score (p=0.249) | | | |
| Mild to secure | 897 | 1841 | 0.07 (−0.01, 0.15) |
| Moderate to severe | 552 | 1266 | 0.13 (0.02, 0.24) |
| Head circumference (cm) (p=0.053) | | | |
| Mild to secure | 898 | 1844 | 0.04 (−0.05, 0.13) |
| Moderate to severe | 555 | 1273 | 0.21 (0.07, 0.35) |
| Head circumference−for−age z−score (p=0.144) | | | |
| Mild to secure | 898 | 1844 | 0.05 (−0.03, 0.12) |
| Moderate to severe | 555 | 1273 | 0.16 (0.05, 0.28) |
| Head circumference−for−gestational age z score (p=0.556) | | | |
| Mild to secure | 301 | 286 | 0.18 (0.00, 0.35) |
| Moderate to severe | 246 | 274 | 0.06 (−0.12, 0.24) |
| Mid−upper arm circumference (cm) (p=0.110) | | | |
| Mild to secure | 900 | 1844 | 0.04 (−0.04, 0.12) |
| Moderate to severe | 557 | 1273 | 0.12 (0.04, 0.20) |
| Duration (wk) (p=0.351) | | | |
| Mild to secure | 981 | 1919 | 0.06 (−0.08, 0.21) |
| Moderate to severe | 678 | 1389 | 0.21 (−0.03, 0.46) |

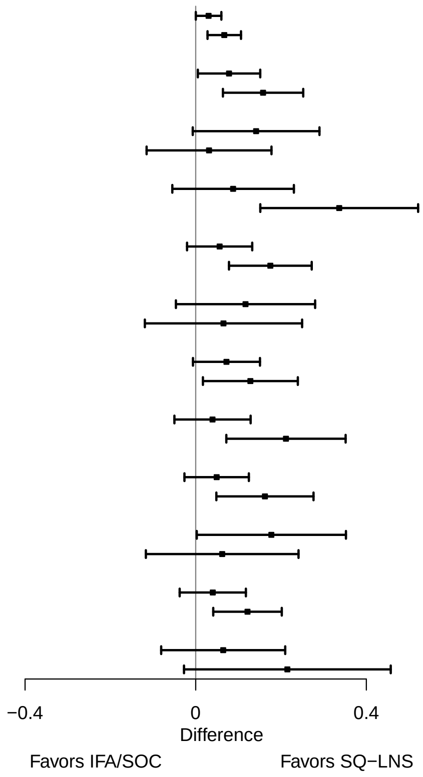

**Supplemental figure 3M: Household food security**

#### 3M2: Relative risks for birth outcomes

| Outcome (p−for−interaction) | SQ−LNS N | IFA/SOC N | Relative risk (95% CI) |
|---|---|---|---|
| Low birth weight (p=0.176) | | | |
| Mild to secure | 945 | 1884 | 0.96 (0.84, 1.10) |
| Moderate to severe | 643 | 1352 | 0.84 (0.71, 0.99) |
| Birth weight < 2 kg (p=0.189) | | | |
| Mild to secure | 945 | 1884 | 0.90 (0.66, 1.23) |
| Moderate to severe | 643 | 1352 | 0.63 (0.41, 0.99) |
| Small−for−gestational age (p=0.554) | | | |
| Mild to secure | 945 | 1884 | 0.98 (0.92, 1.05) |
| Moderate to severe | 643 | 1352 | 0.95 (0.86, 1.05) |
| Large−for−gestational age (p=0.279) | | | |
| Mild to secure | 354 | 331 | 0.70 (0.30, 1.65) |
| Moderate to severe | 336 | 354 | 1.10 (0.51, 2.36) |
| Newborn stunting (p=0.052) | | | |
| Mild to secure | 897 | 1844 | 0.93 (0.80, 1.08) |
| Moderate to severe | 556 | 1271 | 0.75 (0.61, 0.91) |
| Low LGAZ (p=0.514) | | | |
| Mild to secure | 897 | 1844 | 0.88 (0.73, 1.05) |
| Moderate to severe | 556 | 1271 | 0.96 (0.80, 1.17) |
| Low BMIZ (p=0.406) | | | |
| Mild to secure | 897 | 1841 | 0.93 (0.81, 1.06) |
| Moderate to severe | 552 | 1266 | 0.87 (0.75, 1.00) |
| Low HCZ (p=0.959) | | | |
| Mild to secure | 898 | 1844 | 0.86 (0.72, 1.03) |
| Moderate to severe | 555 | 1273 | 0.85 (0.70, 1.03) |
| Low HCGAZ (p=0.442) | | | |
| Mild to secure | 811 | 1762 | 0.84 (0.70, 1.00) |
| Moderate to severe | 392 | 1093 | 0.95 (0.74, 1.22) |
| Preterm birth (p=0.310) | | | |
| Mild to secure | 981 | 1919 | 1.03 (0.86, 1.22) |
| Moderate to severe | 678 | 1389 | 0.86 (0.66, 1.13) |

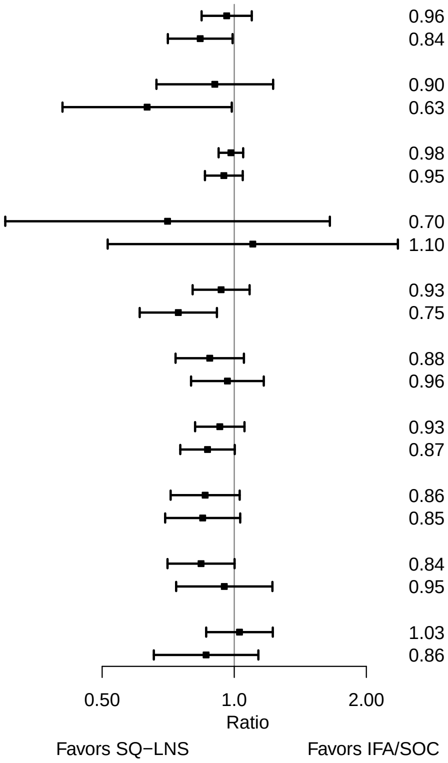

**Supplemental figure 3M: Household food security**

#### 3M3: Mean differences for 6 mo outcomes

| Outcome (p−for−interaction) | SQ−LNS N | IFA/SOC N | Mean difference (95% CI) |
|---|---|---|---|
| Weight−for−age z score (p=0.104) | | | |
| Mild to secure | 891 | 1798 | −0.03 (−0.14, 0.08) |
| Moderate to severe | 603 | 1298 | 0.07 (−0.04, 0.18) |
| Length−for−age z−score (p=0.044) | | | |
| Mild to secure | 891 | 1799 | 0.00 (−0.10, 0.09) |
| Moderate to severe | 604 | 1299 | 0.10 (0.01, 0.19) |
| Weight−for−length z−score (p=0.657) | | | |
| Mild to secure | 891 | 1797 | −0.03 (−0.13, 0.07) |
| Moderate to severe | 603 | 1298 | −0.01 (−0.12, 0.11) |
| Head circumference−for−age z−score (p=0.378) | | | |
| Mild to secure | 891 | 1800 | −0.01 (−0.09, 0.07) |
| Moderate to severe | 604 | 1297 | 0.02 (−0.07, 0.11) |
| MUAC−for−age z−score (p=0.215) | | | |
| Mild to secure | 891 | 1800 | −0.05 (−0.15, 0.05) |
| Moderate to severe | 604 | 1299 | 0.04 (−0.06, 0.14) |

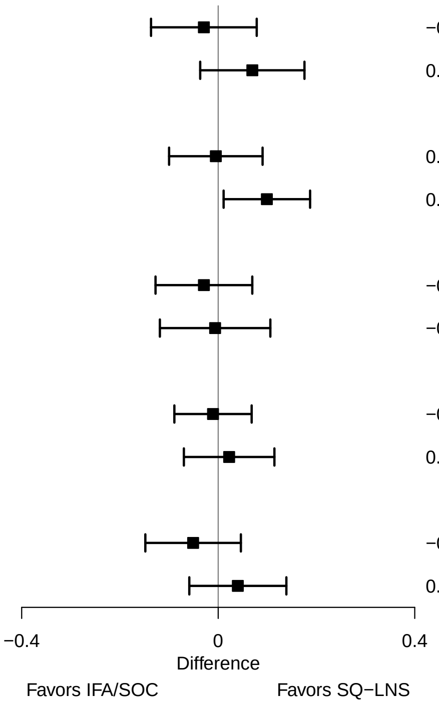

**Supplemental figure 3M: Household food security**

#### 3M4: Prevalence ratios for 6 mo outcomes

| Outcome (p−for−interaction) | SQ−LNS N | IFA/SOC N | Prevalence ratio (95% CI) |
|---|---|---|---|
| Underweight (p=0.053) | | | |
| Mild to secure | 891 | 1798 | 1.10 (0.86, 1.40) |
| Moderate to severe | 603 | 1298 | 0.84 (0.66, 1.07) |
| Stunted (p=0.173) | | | |
| Mild to secure | 891 | 1799 | 1.03 (0.84, 1.25) |
| Moderate to severe | 604 | 1299 | 0.85 (0.72, 1.00) |
| Wasted (p=0.965) | | | |
| Mild to secure | 891 | 1797 | 0.92 (0.59, 1.42) |
| Moderate to severe | 603 | 1298 | 1.04 (0.64, 1.68) |
| Low HCAZ (p=0.250) | | | |
| Mild to secure | 891 | 1800 | 0.97 (0.82, 1.14) |
| Moderate to severe | 604 | 1297 | 0.85 (0.71, 1.02) |
| Low MUAC (p=0.552) | | | |
| Mild to secure | 891 | 1800 | 1.14 (0.82, 1.58) |
| Moderate to severe | 604 | 1299 | 0.95 (0.68, 1.32) |
| Acute malnutrition (p=0.980) | | | |
| Mild to secure | 891 | 1797 | 1.03 (0.77, 1.38) |
| Moderate to severe | 603 | 1298 | 0.98 (0.71, 1.35) |

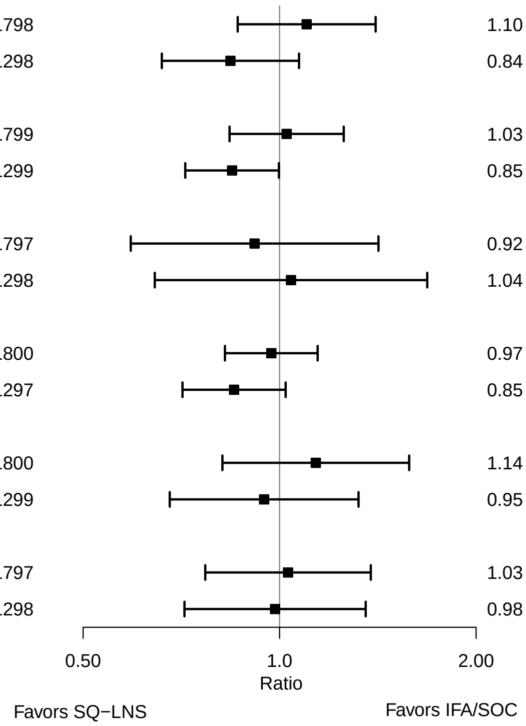

### Supplemental figure 3N: Sanitation

#### 3N1: Mean differences for birth outcomes

| Outcome (p−for−interaction) | SQ−LNS N | IFA/SOC N | Mean difference (95% CI) |
|---|---|---|---|
| Weight (kg) (p=0.194) | | | |
| Improved | 1074 | 2212 | 0.07 (0.04, 0.10) |
| Unimproved | 738 | 1238 | 0.03 (0.00, 0.06) |
| Weight−for−age z score (p=0.141) | | | |
| Improved | 1074 | 2212 | 0.17 (0.10, 0.24) |
| Unimproved | 738 | 1238 | 0.07 (−0.01, 0.15) |
| Weight−for−gestational age z−score (p=0.009) | | | |
| Improved | 113 | 111 | 0.36 (0.21, 0.52) |
| Unimproved | 454 | 437 | 0.00 (−0.11, 0.12) |
| Birth length (cm) (p=0.978) | | | |
| Improved | 1060 | 2199 | 0.19 (0.06, 0.32) |
| Unimproved | 618 | 1130 | 0.21 (0.02, 0.40) |
| Length−for−age z−score (p=0.720) | | | |
| Improved | 1060 | 2199 | 0.11 (0.04, 0.18) |
| Unimproved | 618 | 1130 | 0.11 (0.01, 0.21) |
| Length−for−gestational age z−score (p=0.278) | | | |
| Improved | 100 | 99 | 0.25 (0.05, 0.44) |
| Unimproved | 334 | 329 | 0.07 (−0.08, 0.21) |
| BMI z−score (p=0.073) | | | |
| Improved | 1060 | 2199 | 0.17 (0.09, 0.25) |
| Unimproved | 614 | 1122 | 0.01 (−0.09, 0.11) |
| Head circumference (cm) (p=0.323) | | | |
| Improved | 1059 | 2200 | 0.09 (0.00, 0.18) |
| Unimproved | 619 | 1131 | 0.13 (0.01, 0.25) |
| Head circumference−for−age z−score (p=0.511) | | | |
| Improved | 1059 | 2200 | 0.09 (0.01, 0.16) |
| Unimproved | 619 | 1131 | 0.10 (0.01, 0.20) |
| Head circumference−for−gestational age z score (p=0.985) | | | |
| Improved | 99 | 99 | 0.08 (−0.15, 0.31) |
| Unimproved | 335 | 330 | 0.06 (−0.08, 0.20) |
| Mid−upper arm circumference (cm) (p=0.873) | | | |
| Improved | 960 | 2105 | 0.08 (0.01, 0.15) |
| Unimproved | 499 | 1010 | 0.06 (−0.02, 0.14) |
| Duration (wk) (p=0.963) | | | |
| Improved | 1090 | 2227 | 0.11 (−0.04, 0.26) |
| Unimproved | 760 | 1257 | 0.15 (−0.05, 0.34) |

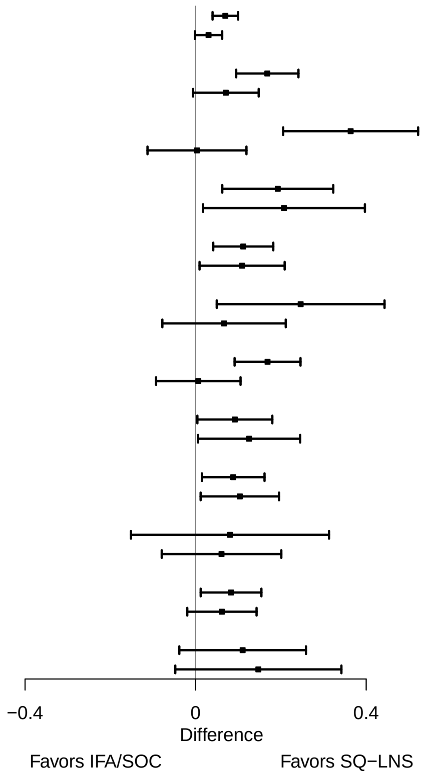

**Supplemental figure 3N: Sanitation**

#### 3N2: Relative risks for birth outcomes

| Outcome (p−for−interaction) | SQ−LNS N | IFA/SOC N | Relative risk (95% CI) |
|---|---|---|---|
| Low birth weight (p=0.324) | | | |
| Improved | 1074 | 2212 | 0.86 (0.75, 0.98) |
| Unimproved | 738 | 1238 | 0.95 (0.83, 1.09) |
| Birth weight < 2 kg (p=0.023) | | | |
| Improved | 676 | 1824 | 0.91 (0.66, 1.24) |
| Unimproved | 607 | 1106 | 0.61 (0.42, 0.87) |
| Small−for−gestational age (p=0.121) | | | |
| Improved | 1056 | 2194 | 0.94 (0.88, 1.00) |
| Unimproved | 714 | 1206 | 1.02 (0.94, 1.12) |
| Newborn stunting (p=0.252) | | | |
| Improved | 1043 | 2177 | 0.88 (0.76, 1.02) |
| Unimproved | 382 | 890 | 0.76 (0.62, 0.93) |
| Low LGAZ (p=0.740) | | | |
| Improved | 1042 | 2181 | 0.90 (0.77, 1.04) |
| Unimproved | 594 | 1098 | 0.93 (0.77, 1.14) |
| Low BMIZ (p=0.130) | | | |
| Improved | 763 | 1906 | 0.86 (0.77, 0.97) |
| Unimproved | 605 | 1111 | 1.01 (0.86, 1.17) |
| Low HCZ (p=0.678) | | | |
| Improved | 746 | 1885 | 0.87 (0.74, 1.03) |
| Unimproved | 373 | 879 | 0.87 (0.70, 1.09) |
| Low HCGAZ (p=0.528) | | | |
| Improved | 728 | 1867 | 0.91 (0.77, 1.09) |
| Unimproved | 349 | 847 | 0.86 (0.69, 1.06) |
| Preterm birth (p=0.727) | | | |
| Improved | 770 | 1912 | 0.98 (0.78, 1.24) |
| Unimproved | 750 | 1245 | 0.88 (0.70, 1.11) |

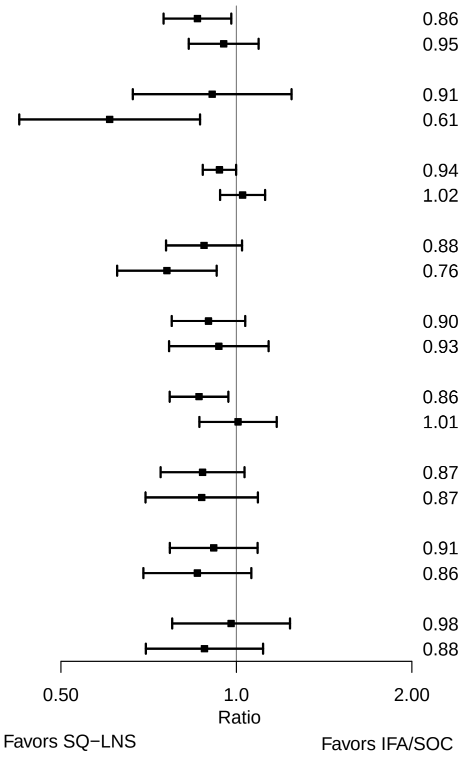

**Supplemental figure 3N: Sanitation**

#### 3N3: Mean differences for 6 mo outcomes

| Outcome (p−for−interaction) | SQ−LNS N | IFA/SOC N | Mean difference (95% CI) |
|---|---|---|---|
| Weight−for−age z score (p=0.097) | | | |
| Improved | 1018 | 2125 | 0.10 (0.01, 0.19) |
| Unimproved | 696 | 1183 | −0.02 (−0.13, 0.09) |
| Length−for−age z−score (p=0.601) | | | |
| Improved | 1019 | 2126 | 0.07 (−0.01, 0.15) |
| Unimproved | 696 | 1184 | 0.03 (−0.07, 0.14) |
| Weight−for−length z−score (p=0.044) | | | |
| Improved | 1018 | 2124 | 0.04 (−0.05, 0.13) |
| Unimproved | 696 | 1183 | −0.07 (−0.17, 0.03) |
| Head circumference−for−age z−score (p=0.252) | | | |
| Improved | 1019 | 2126 | 0.04 (−0.03, 0.11) |
| Unimproved | 696 | 1180 | −0.03 (−0.12, 0.06) |
| MUAC−for−age z−score (p=0.138) | | | |
| Improved | 919 | 2029 | 0.01 (−0.08, 0.09) |
| Unimproved | 577 | 1068 | −0.03 (−0.14, 0.08) |

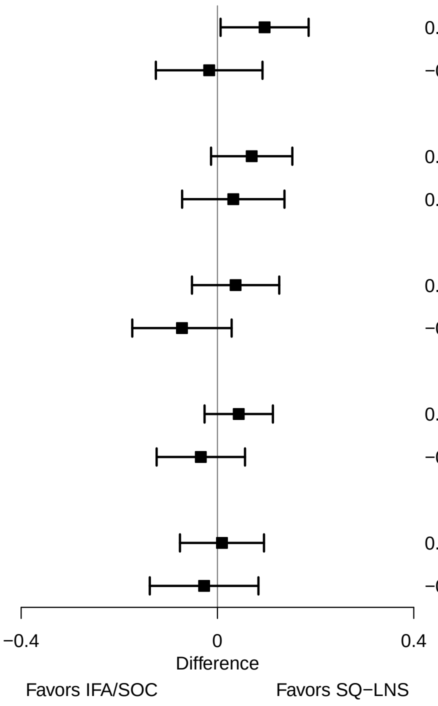

**Supplemental figure 3N: Sanitation**

#### 3N4: Prevalence ratios for 6 mo outcomes

| Outcome (p−for−interaction) | SQ−LNS N | IFA/SOC N | Prevalence ratio (95% CI) |
|---|---|---|---|
| Underweight (p=0.239) | | | |
| Improved | 747 | 1865 | 0.85 (0.66, 1.10) |
| Unimproved | 687 | 1172 | 1.04 (0.83, 1.31) |
| Stunted (p=0.122) | | | |
| Improved | 1019 | 2126 | 0.85 (0.72, 1.00) |
| Unimproved | 696 | 1184 | 0.97 (0.82, 1.16) |
| Low HCAZ (p=0.154) | | | |
| Improved | 1019 | 2126 | 0.85 (0.74, 0.98) |
| Unimproved | 696 | 1180 | 1.03 (0.83, 1.27) |

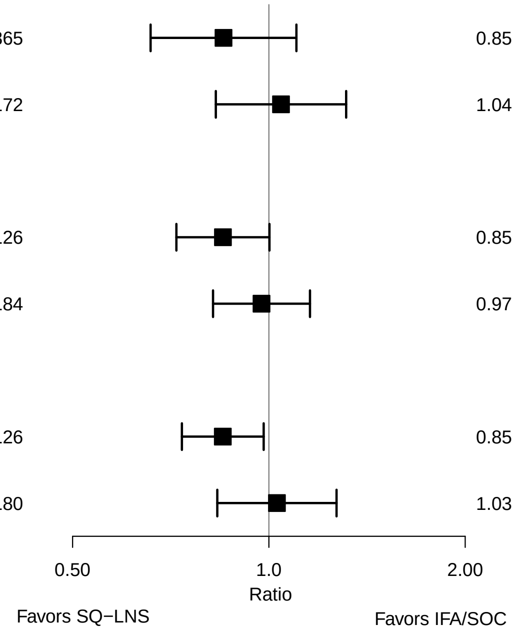

### Supplemental figure 4: Pooled plots for adverse outcomes stratified by potential effect modifiers, SQ-LNS vs IFA/SOC

#### Contents

**Supplemental figure 4A: Sex** 2

**Supplemental figure 4B: Birth order** 3

**Supplemental figure 4C: Maternal height** 4

**Supplemental figure 4D: Maternal BMI** 5

**Supplemental figure 4E: Maternal age** 6

**Supplemental figure 4F: Maternal education** 7

**Supplemental figure 4G: Baseline anemia status** 8

**Supplemental figure 4H: Baseline inflammation status** 9

**Supplemental figure 4I: Baseline malaria status** 10

**Supplemental figure 4J: Gestational age at supplementation** 11

**Supplemental figure 4K: Compliance with supplementation** 12

**Supplemental figure 4L: Household socio-economic status** 13

**Supplemental figure 4M: Household food security** 14

**Supplemental figure 4N: Sanitation** 15

These figures show pooled effects of SQ-LNS within study-level and individual-level characteristic subgroups along with the p-for-interaction. For definitions of effect modifiers, see Box 1 in the main paper. Individual study estimates were generated from log-binomial regression for dichotomous outcomes and simple linear regression for continuous outcomes with clustered observations using robust standard errors for cluster-randomized trials. Pooled sub-group estimates and statistical testing of the pooled interaction term were generated using inverse-variance weighting. For dichotomous outcomes analyzed via prevalence/risk ratios, the effect estimate is the prevalence/risk in the SQ-LNS group divided by the prevalence/risk in the IFA/SOC group. For dichotomous outcomes analyzed via prevalence/risk differences, the effect estimate is the prevalence/risk in the SQ-LNS group minus the prevalence/risk in the IFA/SOC group. The labels on the left y-axis correspond to the characteristic subgroups and their sample sizes. The values on the right indicate the pooled prevalence ratio and confidence interval within that subgroup.

LAZ, length-for-age z-score; WLZ, weight-for-length z-score; WAZ, weight for-age z-score; MUACZ, mid-upper arm circumference z-score; BMI, body mass index; HCZ, head circumference-for-age z-score; LGAZ, length-for-gestational-age z-score; HCGAZ, head circumference-for-gestational-age z-score; BMIZ, body mass index-for-age z-score; IFA/SOC, Iron and folic acid or standard of care; MD, mean difference; MMS, multiple micronutrient supplement; MUAC, mid-upper arm circumference; PR, prevalence ratio; PD, prevalence difference; RD, risk difference; RR, relative risk; SOC, standard of care; SQ-LNS, small-quantity lipid-based nutrient supplements; WGAZ, weight-for-gestational age z-score.

#### Supplemental figure 4A: Sex

| Outcome (p−for−interaction) | SQ−LNS N | IFA/SOC N | Relative risk (95% CI) |
|---|---|---|---|
| C−section (p=0.037) | | | |
| Male | 977 | 1874 | 1.01 (0.83, 1.22) |
| Female | 984 | 1840 | 1.11 (0.88, 1.39) |
| Neonatal mortality (p=0.306) | | | |
| Male | 662 | 1544 | 0.65 (0.33, 1.26) |
| Female | 672 | 1516 | 1.08 (0.60, 1.93) |
| Miscarriage or stillbirth or early neonatal mortality (p=0.405) | | | |
| Male | 980 | 1883 | 1.03 (0.71, 1.50) |
| Female | 989 | 1853 | 1.27 (0.85, 1.91) |
| Mortality 0−6 mo (p=0.579) | | | |
| Male | 778 | 1670 | 0.86 (0.52, 1.41) |
| Female | 798 | 1637 | 1.04 (0.65, 1.65) |

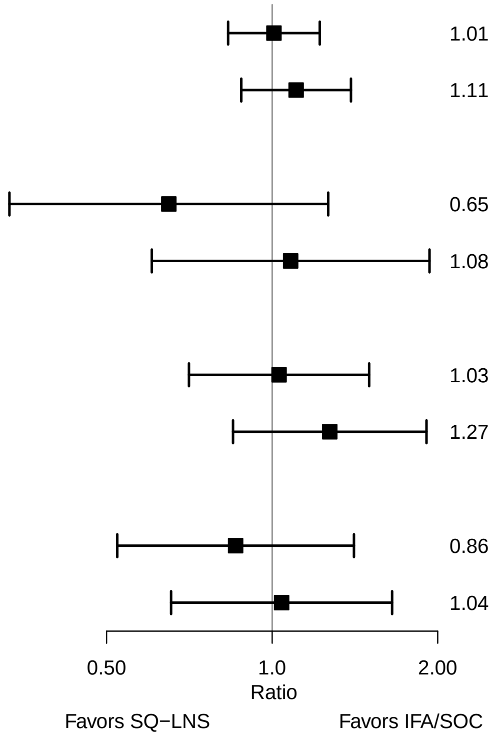

### Supplemental figure 4B: Birth order

| Outcome (p−for−interaction) | SQ−LNS N | IFA/SOC N | Relative risk (95% CI) |
|---|---|---|---|
| C−section (p=0.199) | | | |
| Later born | 1337 | 2418 | 1.01 (0.87, 1.17) |
| Firstborn | 630 | 1301 | 0.96 (0.75, 1.24) |
| Neonatal mortality (p=0.024) | | | |
| Later born | 1079 | 2138 | 1.08 (0.69, 1.70) |
| Firstborn | 594 | 1257 | 0.54 (0.27, 1.06) |
| Miscarriage or stillbirth or early neonatal mortality (p=0.657) | | | |
| Later born | 1421 | 2619 | 0.91 (0.73, 1.13) |
| Firstborn | 670 | 1381 | 0.97 (0.73, 1.30) |
| Mortality 0−6 mo (p=0.037) | | | |
| Later born | 1092 | 2156 | 1.22 (0.86, 1.73) |
| Firstborn | 598 | 1267 | 0.60 (0.32, 1.15) |

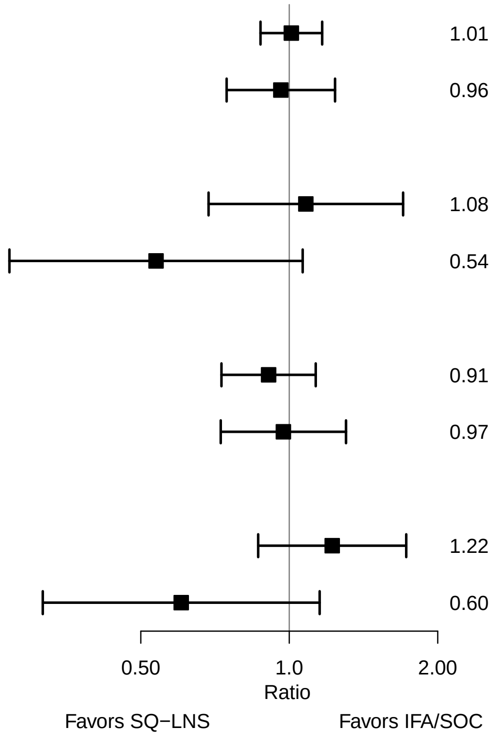

### Supplemental figure 4C: Maternal height

| Outcome (p−for−interaction) | SQ−LNS N | IFA/SOC N | Relative risk (95% CI) |
|---|---|---|---|
| C−section (p=0.288) | | | |
| At least 150.1 | 1244 | 2099 | 1.06 (0.87, 1.29) |
| Less than 150.1 | 679 | 1529 | 1.00 (0.83, 1.22) |
| Neonatal mortality (p=0.315) | | | |
| At least 150.1 | 900 | 1766 | 0.61 (0.36, 1.05) |
| Less than 150.1 | 630 | 1449 | 0.94 (0.47, 1.86) |
| Miscarriage or stillbirth or early neonatal mortality (p=0.086) | | | |
| At least 150.1 | 1308 | 2238 | 0.80 (0.63, 1.03) |
| Less than 150.1 | 735 | 1649 | 1.07 (0.81, 1.41) |
| Mortality 0−6 mo (p=0.523) | | | |
| At least 150.1 | 908 | 1771 | 0.80 (0.51, 1.24) |
| Less than 150.1 | 637 | 1466 | 0.99 (0.60, 1.61) |

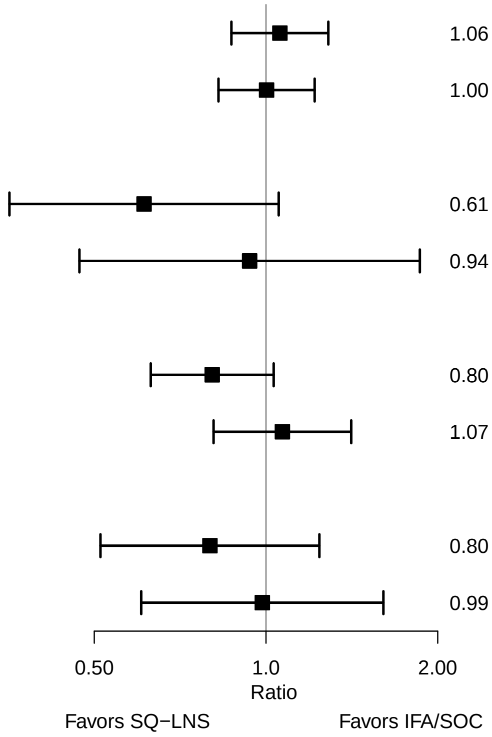

### Supplemental figure 4D: Maternal BMI

| Outcome (p−for−interaction) | SQ−LNS N | IFA/SOC N | Relative risk (95% CI) |
|---|---|---|---|
| C−section (p=0.987) | | | |
| At least 20 | 1123 | 1937 | 1.04 (0.91, 1.19) |
| Less than 20 | 799 | 1689 | 1.24 (0.86, 1.78) |
| Neonatal mortality (p=0.472) | | | |
| At least 20 | 611 | 1419 | 0.79 (0.42, 1.48) |
| Less than 20 | 693 | 1569 | 0.70 (0.40, 1.23) |
| Miscarriage or stillbirth or early neonatal mortality (p=0.879) | | | |
| At least 20 | 910 | 1804 | 0.94 (0.72, 1.23) |
| Less than 20 | 783 | 1739 | 0.98 (0.72, 1.34) |
| Mortality 0−6 mo (p=0.329) | | | |
| At least 20 | 620 | 1429 | 1.04 (0.66, 1.64) |
| Less than 20 | 701 | 1586 | 0.82 (0.52, 1.28) |

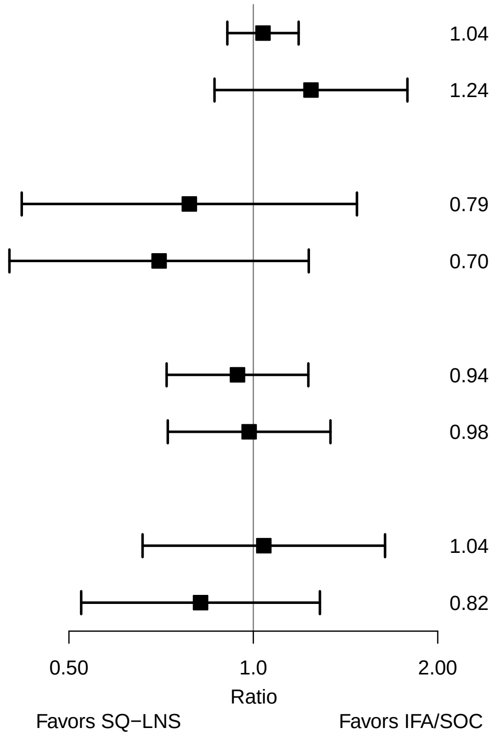

### Supplemental figure 4E: Maternal age

| Outcome (p−for−interaction) | SQ−LNS N | IFA/SOC N | Relative risk (95% CI) |
|---|---|---|---|
| C−section (p=0.994) | | | |
| At least 25 | 777 | 1262 | 1.02 (0.84, 1.24) |
| Less than 25 | 1191 | 2463 | 1.08 (0.88, 1.31) |
| Neonatal mortality (p=0.424) | | | |
| At least 25 | 546 | 1035 | 0.61 (0.33, 1.13) |
| Less than 25 | 1024 | 2267 | 0.97 (0.57, 1.65) |
| Miscarriage or stillbirth or early neonatal mortality (p=0.505) | | | |
| At least 25 | 827 | 1373 | 0.90 (0.71, 1.15) |
| Less than 25 | 1269 | 2633 | 1.00 (0.79, 1.26) |
| Mortality 0−6 mo (p=0.918) | | | |
| At least 25 | 555 | 1039 | 0.86 (0.54, 1.40) |
| Less than 25 | 1030 | 2286 | 0.96 (0.63, 1.46) |

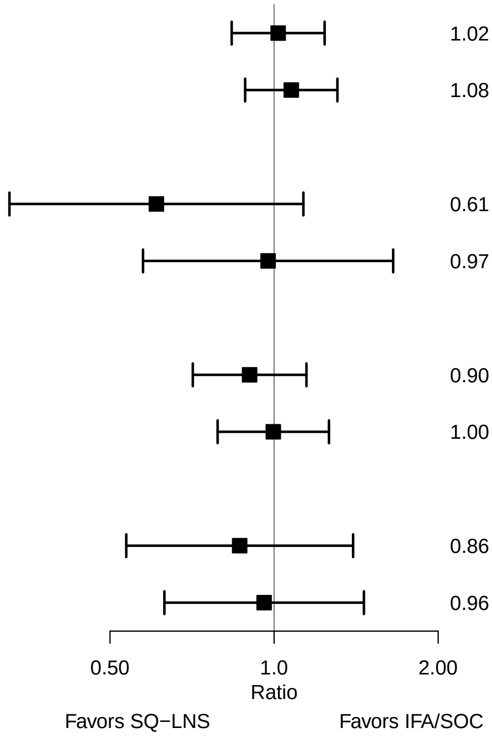

### Supplemental figure 4F: Maternal education

| Outcome (p−for−interaction) | SQ−LNS N | IFA/SOC N | Relative risk (95% CI) |
|---|---|---|---|
| C−section (p=0.919) | | | |
| Primary or greater | 1301 | 2503 | 1.01 (0.87, 1.16) |
| Incomplete or no formal | 663 | 1218 | 1.17 (0.86, 1.59) |
| Neonatal mortality (p=0.871) | | | |
| Primary or greater | 1056 | 2232 | 1.06 (0.56, 1.98) |
| Incomplete or no formal | 616 | 1165 | 0.80 (0.48, 1.33) |
| Miscarriage or stillbirth or early neonatal mortality (p=0.936) | | | |
| Primary or greater | 1393 | 2687 | 0.93 (0.74, 1.16) |
| Incomplete or no formal | 699 | 1314 | 0.95 (0.70, 1.30) |
| Mortality 0−6 mo (p=0.079) | | | |
| Primary or greater | 1059 | 2247 | 0.92 (0.54, 1.57) |
| Incomplete or no formal | 630 | 1178 | 1.36 (0.98, 1.88) |

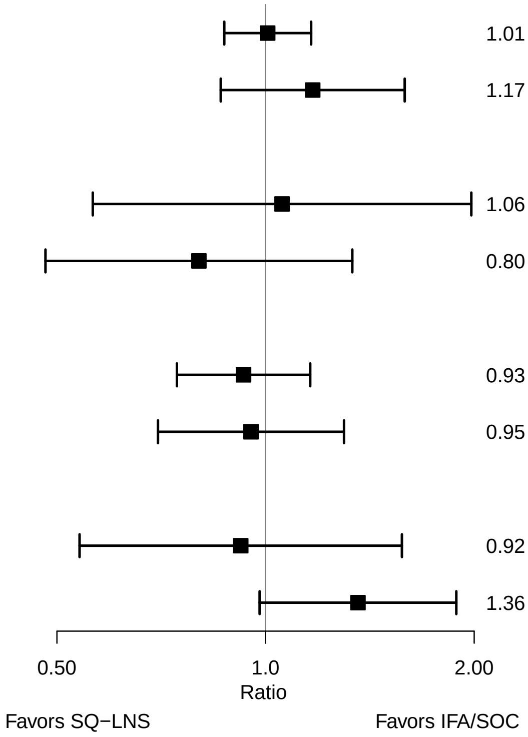

### Supplemental figure 4G: Baseline anemia status

| Outcome (p−for−interaction) | SQ−LNS N | IFA/SOC N | Relative risk (95% CI) |
|---|---|---|---|
| C−section (p=0.571) | | | |
| Anemia | 474 | 461 | 0.99 (0.68, 1.42) |
| No anemia | 867 | 746 | 1.12 (0.86, 1.46) |
| Neonatal mortality (p=0.084) | | | |
| Anemia | 458 | 459 | 1.24 (0.57, 2.67) |
| No anemia | 836 | 741 | 0.64 (0.34, 1.20) |
| Miscarriage or stillbirth or early neonatal mortality (p=0.540) | | | |
| Anemia | 492 | 490 | 1.05 (0.67, 1.67) |
| No anemia | 912 | 797 | 0.93 (0.68, 1.26) |
| Mortality 0−6 mo (p=0.438) | | | |
| Anemia | 460 | 462 | 1.15 (0.58, 2.31) |
| No anemia | 846 | 742 | 0.94 (0.55, 1.61) |

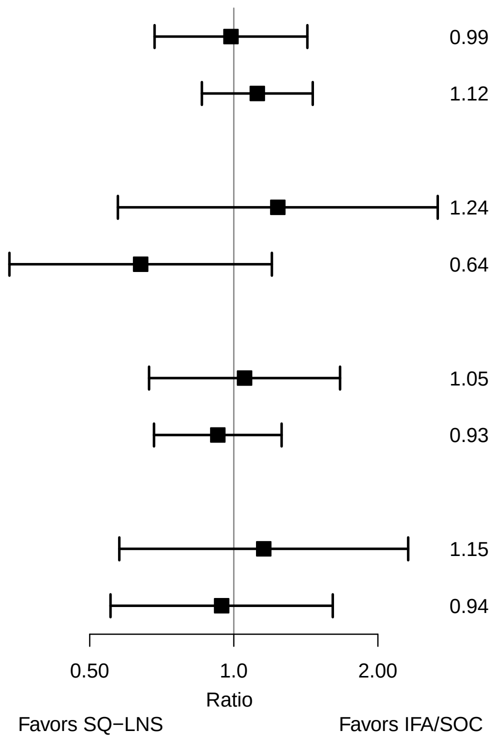

### Supplemental figure 4H: Baseline inflammation status

| Outcome (p−for−interaction) | SQ−LNS N | IFA/SOC N | Relative risk (95% CI) |
|---|---|---|---|
| C−section (p=0.921) | | | |
| Not | 892 | 817 | 1.04 (0.78, 1.37) |
| Inflamed | 433 | 371 | 1.06 (0.75, 1.51) |
| Neonatal mortality (p=0.317) | | | |
| Not | 856 | 809 | 0.91 (0.48, 1.72) |
| Inflamed | 423 | 372 | 0.50 (0.20, 1.23) |
| Miscarriage or stillbirth or early neonatal mortality (p=0.399) | | | |
| Not | 930 | 867 | 1.03 (0.73, 1.44) |
| Inflamed | 457 | 400 | 0.73 (0.44, 1.21) |
| Mortality 0−6 mo (p=0.111) | | | |
| Not | 866 | 811 | 1.24 (0.70, 2.19) |
| Inflamed | 425 | 374 | 0.53 (0.24, 1.17) |

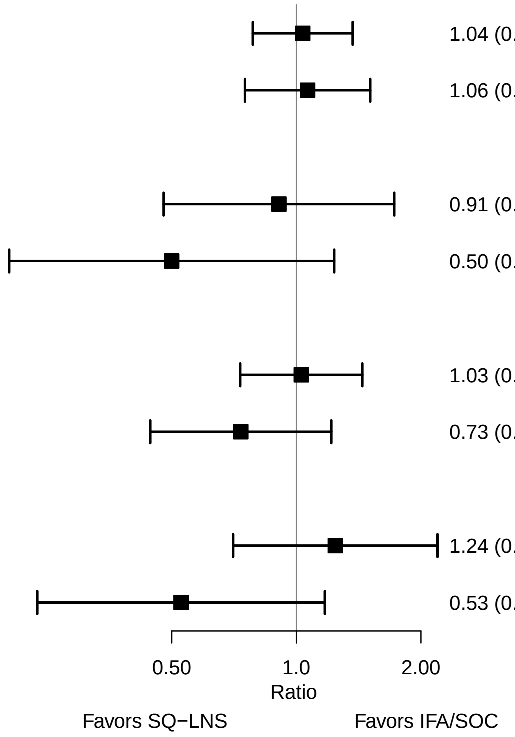

### Supplemental figure 4I: Baseline malaria status

| Outcome (p−for−interaction) | SQ−LNS N | IFA/SOC N | Relative risk (95% CI) |
|---|---|---|---|
| C−section (p=0.018) | | | |
| No malaria | 616 | 614 | 1.24 (0.92, 1.67) |
| Malaria | 132 | 118 | 0.53 (0.20, 1.41) |
| Miscarriage or stillbirth or early neonatal mortality (p=0.620) | | | |
| No malaria | 648 | 655 | 0.83 (0.54, 1.27) |
| Malaria | 135 | 127 | 1.18 (0.44, 3.16) |

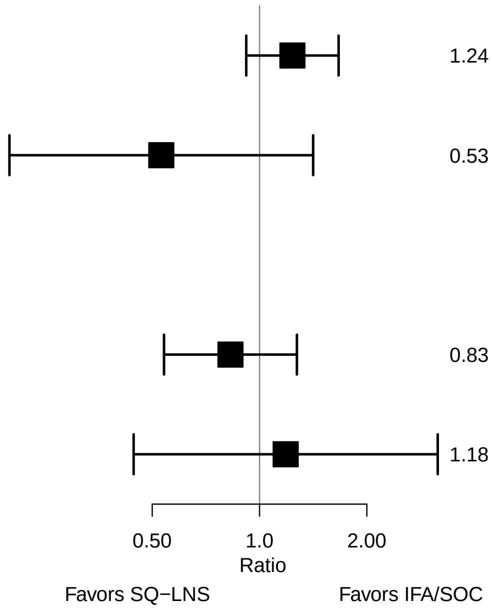

### Supplemental figure 4J: Gestational age at supplementation

| Outcome (p−for−interaction) | SQ−LNS N | IFA/SOC N | Relative risk (95% CI) |
|---|---|---|---|
| C−section (p=0.796) | | | |
| < 14 wks | 914 | 1971 | 1.02 (0.87, 1.20) |
| >= 14 wks | 1031 | 1715 | 1.10 (0.87, 1.39) |
| Neonatal mortality (p=0.877) | | | |
| < 14 wks | 621 | 1662 | 0.82 (0.44, 1.51) |
| >= 14 wks | 714 | 1404 | 0.71 (0.41, 1.23) |
| Miscarriage or stillbirth or early neonatal mortality (p=0.178) | | | |
| < 14 wks | 962 | 2109 | 1.07 (0.81, 1.41) |
| >= 14 wks | 1070 | 1799 | 0.83 (0.62, 1.11) |
| Mortality 0−6 mo (p=0.712) | | | |
| < 14 wks | 627 | 1675 | 0.91 (0.51, 1.64) |
| >= 14 wks | 725 | 1418 | 0.94 (0.64, 1.40) |

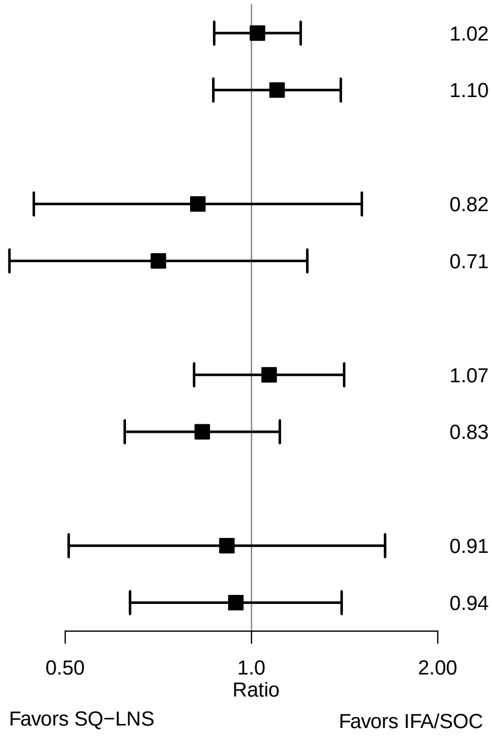

### Supplemental figure 4K: Compliance with supplementation

| Outcome (p−for−interaction) | SQ−LNS N | IFA/SOC N | Relative risk (95% CI) |
|---|---|---|---|
| C−section (p=0.247) | | | |
| >= 4/7 | 1269 | 3170 | 1.05 (0.84, 1.31) |
| Less than 4/7 | 450 | 286 | 1.26 (0.85, 1.86) |
| Miscarriage or stillbirth or early neonatal mortality (p=0.787) | | | |
| >= 4/7 | 890 | 2807 | 0.89 (0.58, 1.36) |
| Less than 4/7 | 434 | 268 | 1.07 (0.52, 2.18) |

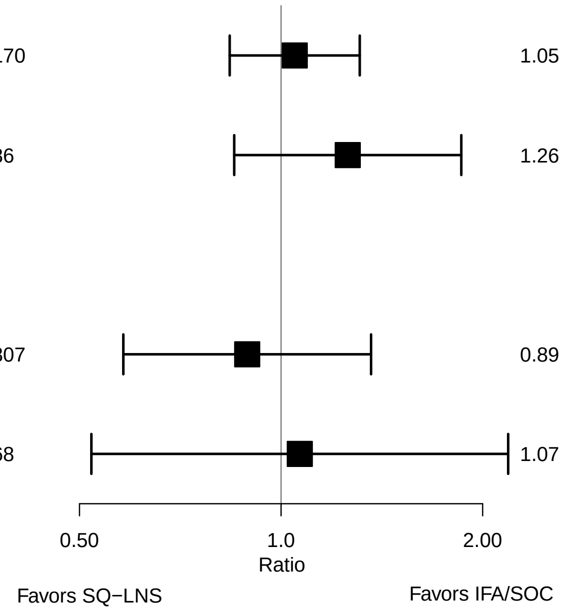

### Supplemental figure 4L: Household socio-economic status

| Outcome (p−for−interaction) | SQ−LNS N | IFA/SOC N | Relative risk (95% CI) |
|---|---|---|---|
| C−section (p=0.700) | | | |
| At least median | 932 | 1812 | 1.06 (0.87, 1.29) |
| Less than median | 998 | 1867 | 1.15 (0.89, 1.48) |
| Neonatal mortality (p=0.082) | | | |
| At least median | 812 | 1687 | 0.60 (0.32, 1.12) |
| Less than median | 860 | 1708 | 1.03 (0.65, 1.65) |
| Miscarriage or stillbirth or early neonatal mortality (p=0.131) | | | |
| At least median | 869 | 1828 | 0.82 (0.61, 1.10) |
| Less than median | 950 | 1895 | 1.08 (0.84, 1.39) |
| Mortality 0−6 mo (p=0.127) | | | |
| At least median | 816 | 1693 | 0.72 (0.41, 1.28) |
| Less than median | 873 | 1729 | 1.15 (0.81, 1.64) |

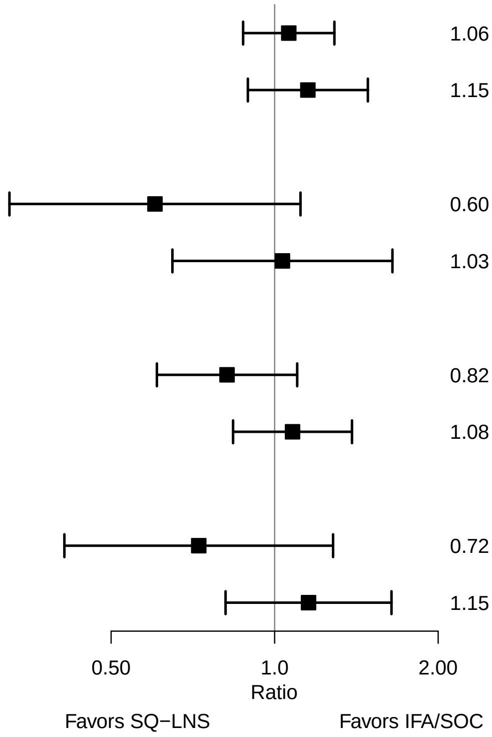

### Supplemental figure 4M: Household food security

| Outcome (p−for−interaction) | SQ−LNS N | IFA/SOC N | Relative risk (95% CI) |
|---|---|---|---|
| C−section (p=0.838) | | | |
| Mild to secure | 1031 | 2035 | 1.14 (0.90, 1.45) |
| Moderate to severe | 697 | 1453 | 1.08 (0.78, 1.51) |
| Neonatal mortality (p=0.895) | | | |
| Mild to secure | 997 | 1982 | 0.95 (0.58, 1.54) |
| Moderate to severe | 675 | 1413 | 0.78 (0.43, 1.41) |
| Miscarriage or stillbirth or early neonatal mortality (p=0.318) | | | |
| Mild to secure | 1076 | 2174 | 0.87 (0.66, 1.14) |
| Moderate to severe | 743 | 1551 | 1.08 (0.80, 1.45) |
| Mortality 0−6 mo (p=0.849) | | | |
| Mild to secure | 1005 | 1992 | 1.06 (0.73, 1.52) |
| Moderate to severe | 684 | 1431 | 0.92 (0.57, 1.49) |

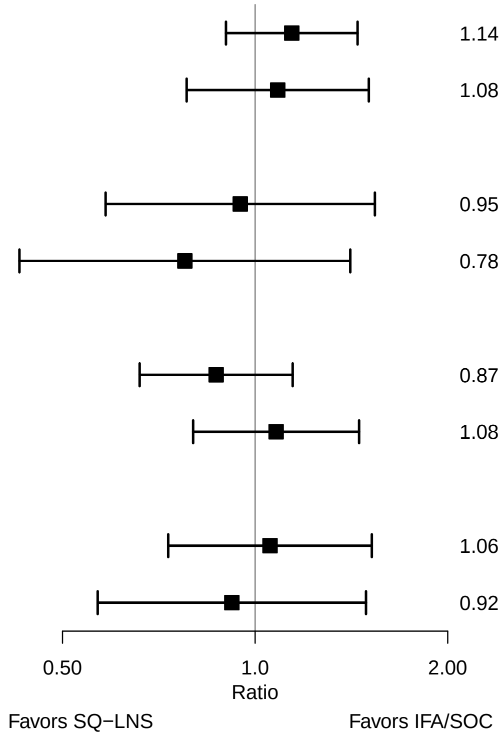

### Supplemental figure 4N: Sanitation

| **Outcome (p−for−interaction)** | **SQ−LNS N** | **IFA/SOC N** | **Relative risk (95% CI)** |
|---|---|---|---|
| C−section (p=0.483) | | | |
| Improved | 1166 | 2397 | 1.00 (0.82, 1.22) |
| Unimproved | 798 | 1320 | 1.19 (0.93, 1.53) |
| Miscarriage or stillbirth or early neonatal mortality (p=0.251) | | | |
| Improved | 865 | 2197 | 0.84 (0.64, 1.10) |
| Unimproved | 445 | 1024 | 1.00 (0.74, 1.35) |
| Mortality 0−6 mo (p=0.919) | | | |
| Improved | 786 | 1992 | 1.03 (0.67, 1.58) |
| Unimproved | 392 | 914 | 0.99 (0.50, 1.98) |

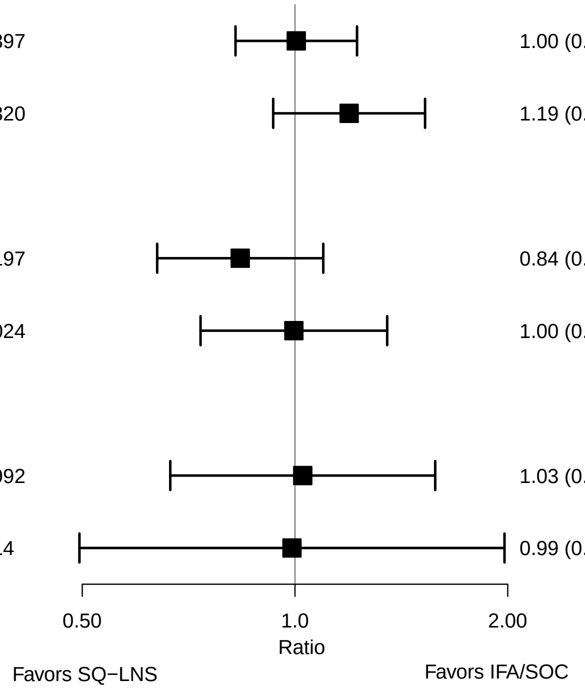

### Supplemental figure 5: Forest plots of main effects for all birth outcomes, SQ-LNS vs MMS

#### Contents

**Supplemental figure 5A: Mean difference in birth weight** **3**

**Supplemental figure 5B: Mean difference in weight-for-age z score** **4**

**Supplemental figure 5C: Mean difference in weight-for-gestational age z-score** **5**

**Supplemental figure 5D: Low birth weight relative risk** **6**

**Supplemental figure 5E: Low birth weight risk difference** **7**

**Supplemental figure 5F: Birth weight < 2 kg relative risk** **8**

**Supplemental figure 5G: Birth weight < 2 kg risk difference** **9**

**Supplemental figure 5H: Small-for-gestational age relative risk** **10**

**Supplemental figure 5I: Small-for-gestational age risk difference** **11**

**Supplemental figure 5J: Large-for-gestational age relative risk** **12**

**Supplemental figure 5K: Large-for-gestational age risk difference** **13**

**Supplemental figure 5L: Mean difference in birth length** **14**

**Supplemental figure 5M: Mean difference in length-for-age z score** **15**

**Supplemental figure 5N: Mean difference in length-for-gestational age z-score** **16**

**Supplemental figure 5O: Newborn stunting relative risk** **17**

**Supplemental figure 5P: Newborn stunting risk difference** **18**

**Supplemental figure 5Q: Low LGAZ relative risk** **19**

**Supplemental figure 5R: Low LGAZ risk difference** **20**

**Supplemental figure 5S: Mean difference in BMI-for-age z-score** **21**

**Supplemental figure 5T: Low BMIZ relative risk** **22**

**Supplemental figure 5U: Low BMIZ risk difference** **23**

**Supplemental figure 5V: Mean difference in head circumference** **24**

**Supplemental figure 5W: Mean difference in head circumference-for-age z score** **25**

**Supplemental figure 5X: Mean difference in head circumference-for-gestational age z score** **26**

**Supplemental figure 5Y: Low HCZ relative risk** **27**

**Supplemental figure 5Z: Low HCZ risk difference** **28**

**Supplemental figure 5AA: Low HCGAZ relative risk** **29**

**Supplemental figure 5AB: Low HCGAZ risk difference** **30**

**Supplemental figure 5AC: Mean difference in mid-upper arm circumference** **31**

**Supplemental figure 5AD: Mean difference in duration of gestation** **32**

**Supplemental figure 5AE: Preterm birth relative risk** **33**

**Supplemental figure 5AF: Preterm birth risk difference** **34**

These figures are forest plots showing the study-level estimates of intervention effect with the pooled estimate in the bottom summary rows. Individual study estimates were generated from log binomial regression for dichotomous outcomes and simple linear regression for continuous outcomes with clustered observations using robust standard errors for cluster-randomized trials. Pooled estimates were generated using inverse variance weighting in both fixed and random effects models. For continuous outcomes the intervention effect is measured by the difference in mean of the SQ-LNS group minus MMS. For dichotomous outcomes analyzed via prevalence/risk ratios the effect estimate is the prevalence/risk in the SQ-LNS group divided by the prevalence/risk in the MMS group. For dichotomous outcomes analyzed via prevalence/risk differences the effect estimate is the prevalence/risk in the SQ-LNS group minus the prevalence/risk in the MMS group. The labels on the left y-axis correspond to trial level information. The values on the right indicate the study level effect estimate, confidence interval, and weighting for deriving the pooled estimate.

LAZ, length-for-age z-score; WLZ, weight-for-length z-score; WAZ, weight for-age z-score; MUACZ, mid-upper arm circumference z-score; BMI, body mass index; HCZ, head circumference-for-age z-score; LGAZ, length-for-gestational-age z-score; HCGAZ, head circumference-for-gestational-age z-score; BMIZ, body mass index-for-age z-score; IFA/SOC, Iron and folic acid or standard of care; MD, mean difference; MMS, multiple micronutrient supplement; MUAC, mid-upper arm circumference; PR, prevalence ratio; PD, prevalence difference; RD, risk difference; RR, relative risk; SOC, standard of care; SQ-LNS, small-quantity lipid-based nutrient supplements; WGAZ, weight-for-gestational age z-score.

### Supplemental figure 5A: Mean difference in birth weight

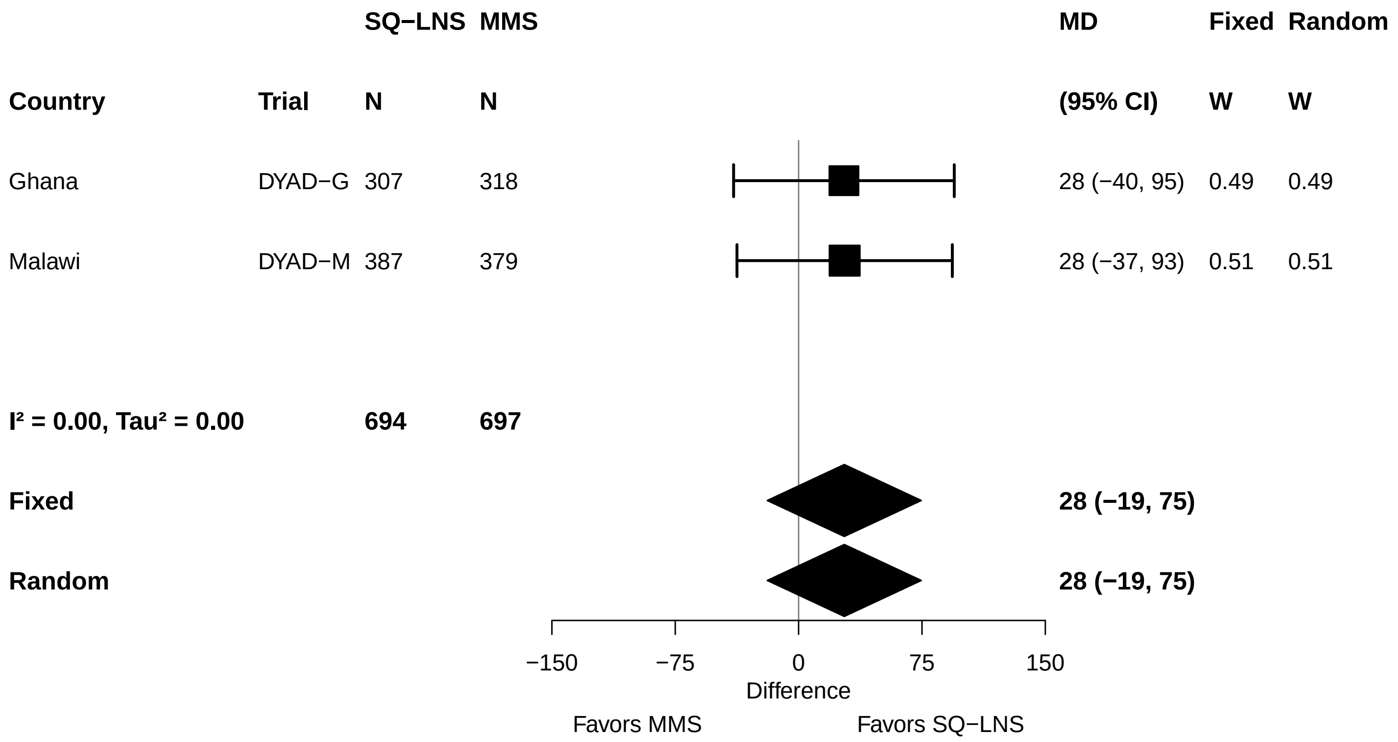

### Supplemental figure 5B: Mean difference in weight-for-age z score

| Country | Trial | SQ−LNS N | MMS N | MD (95% CI) | Fixed W | Random W |
|---|---|---|---|---|---|---|
| Ghana | DYAD−G | 307 | 318 | 0.06 (−0.09, 0.21) | 0.50 | 0.50 |
| Malawi | DYAD−M | 387 | 379 | 0.06 (−0.09, 0.21) | 0.50 | 0.50 |
| **I² = 0.00, Tau² = 0.00** | | **694** | **697** | | | |
| **Fixed** | | | | **0.06 (−0.05, 0.17)** | | |
| **Random** | | | | **0.06 (−0.05, 0.17)** | | |

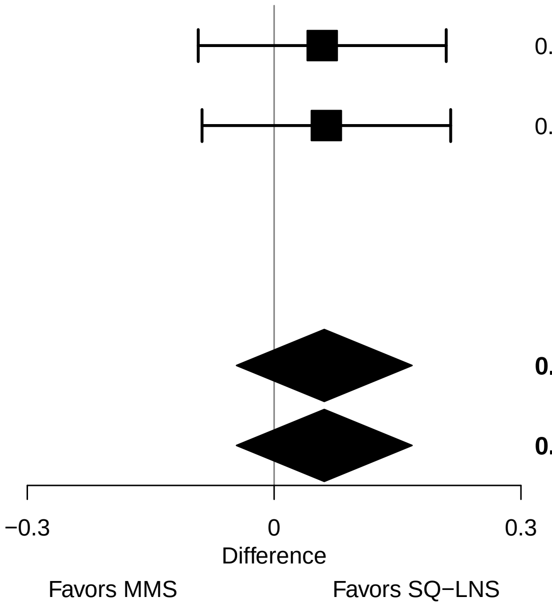

#### Supplemental figure 5C: Mean difference in weight-for-gestational age z-score

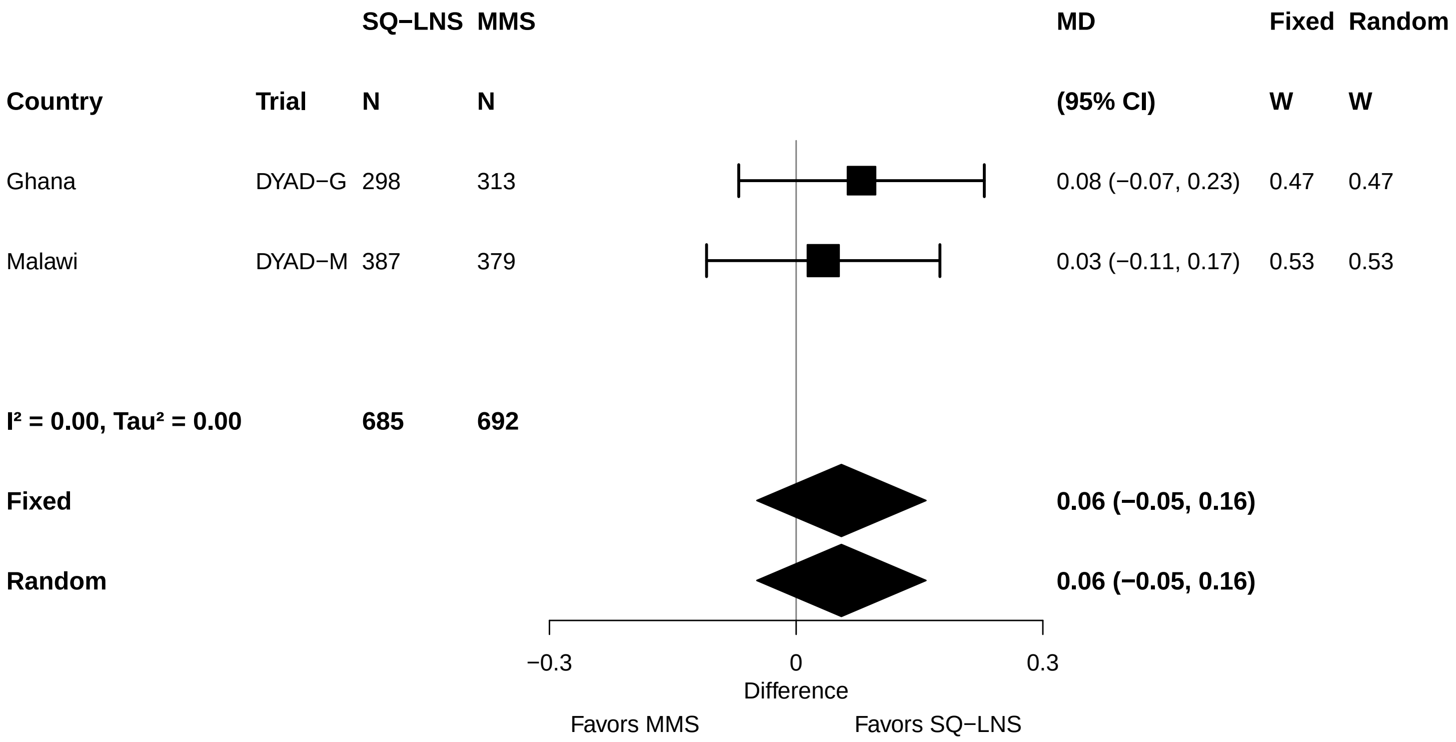

### Supplemental figure 5D: Low birth weight relative risk

| Country | Trial | SQ−LNS N | MMS N | RR (95% CI) | Fixed W | Random W |
|---|---|---|---|---|---|---|
| Ghana | DYAD−G | 307 | 318 | 0.87 (0.53, 1.43) | 0.34 | 0.34 |
| Malawi | DYAD−M | 387 | 379 | 0.89 (0.62, 1.27) | 0.66 | 0.66 |
| **I² = 0.00, Tau² = 0.00** | | **694** | **697** | | | |
| **Fixed** | | | | **0.88 (0.66, 1.18)** | | |
| **Random** | | | | **0.88 (0.66, 1.18)** | | |

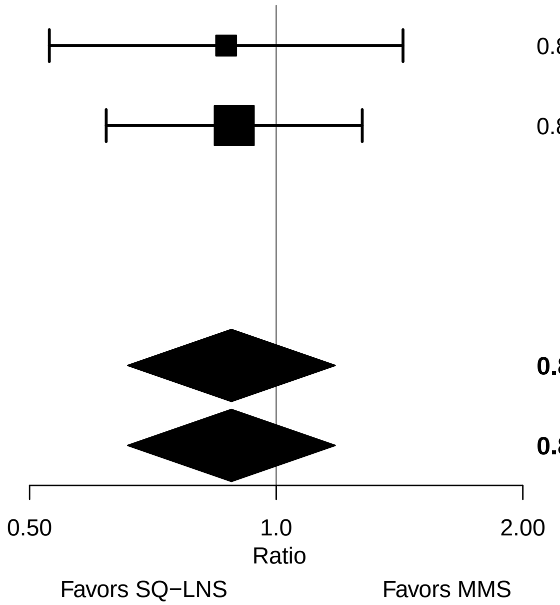

#### Supplemental figure 5E: Low birth weight risk difference

| Country | Trial | SQ−LNS N | MMS N | RD (95% CI) | Fixed W | Random W |
|---|---|---|---|---|---|---|
| Ghana | DYAD−G | 307 | 318 | −0.01 (−0.06, 0.03) | 0.53 | 0.53 |
| Malawi | DYAD−M | 387 | 379 | −0.02 (−0.06, 0.03) | 0.47 | 0.47 |
| **I² = 0.00, Tau² = 0.00** | | **694** | **697** | | | |
| **Fixed** | | | | **−0.01 (−0.05, 0.02)** | | |
| **Random** | | | | **−0.01 (−0.05, 0.02)** | | |

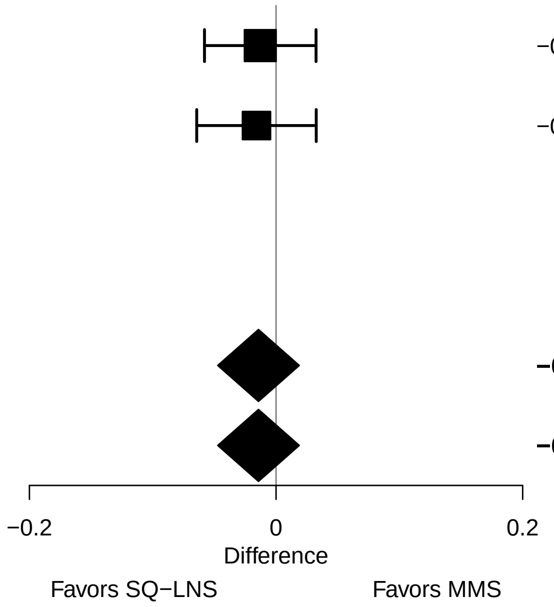

### Supplemental figure 5F: Birth weight < 2 kg relative risk

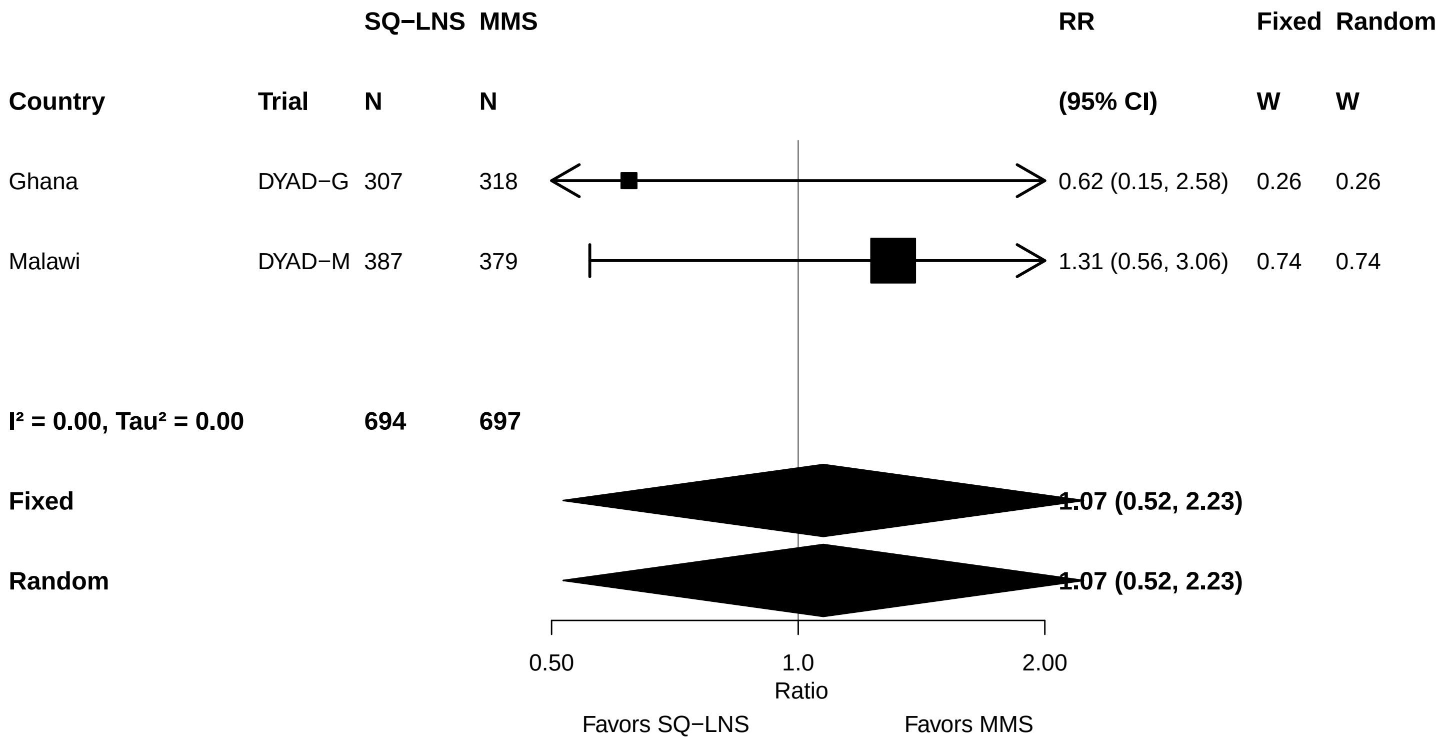

#### Supplemental figure 5G: Birth weight < 2 kg risk difference

| Country | Trial | SQ−LNS N | MMS N | RD (95% CI) | Fixed W | Random W |
|---|---|---|---|---|---|---|
| Ghana | DYAD−G | 307 | 318 | −0.01 (−0.02, 0.01) | 0.63 | 0.63 |
| Malawi | DYAD−M | 387 | 379 | 0.01 (−0.02, 0.03) | 0.37 | 0.37 |
| **I² = 0.00, Tau² = 0.00** | | **694** | **697** | | | |
| **Fixed** | | | | **0.00 (−0.02, 0.01)** | | |
| **Random** | | | | **0.00 (−0.02, 0.01)** | | |

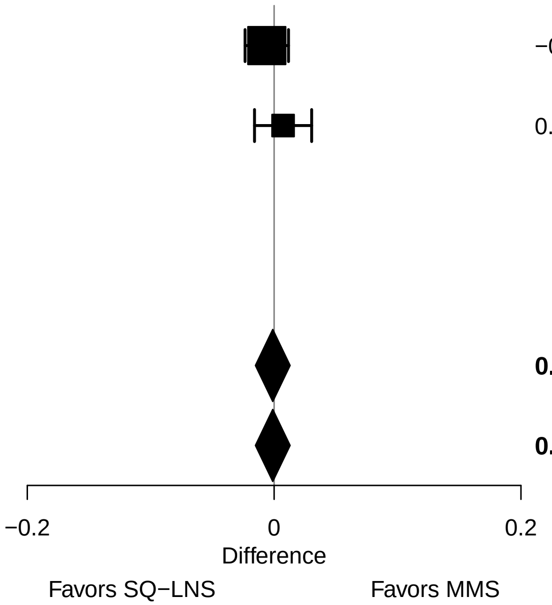

#### Supplemental figure 5H: Small-for-gestational age relative risk

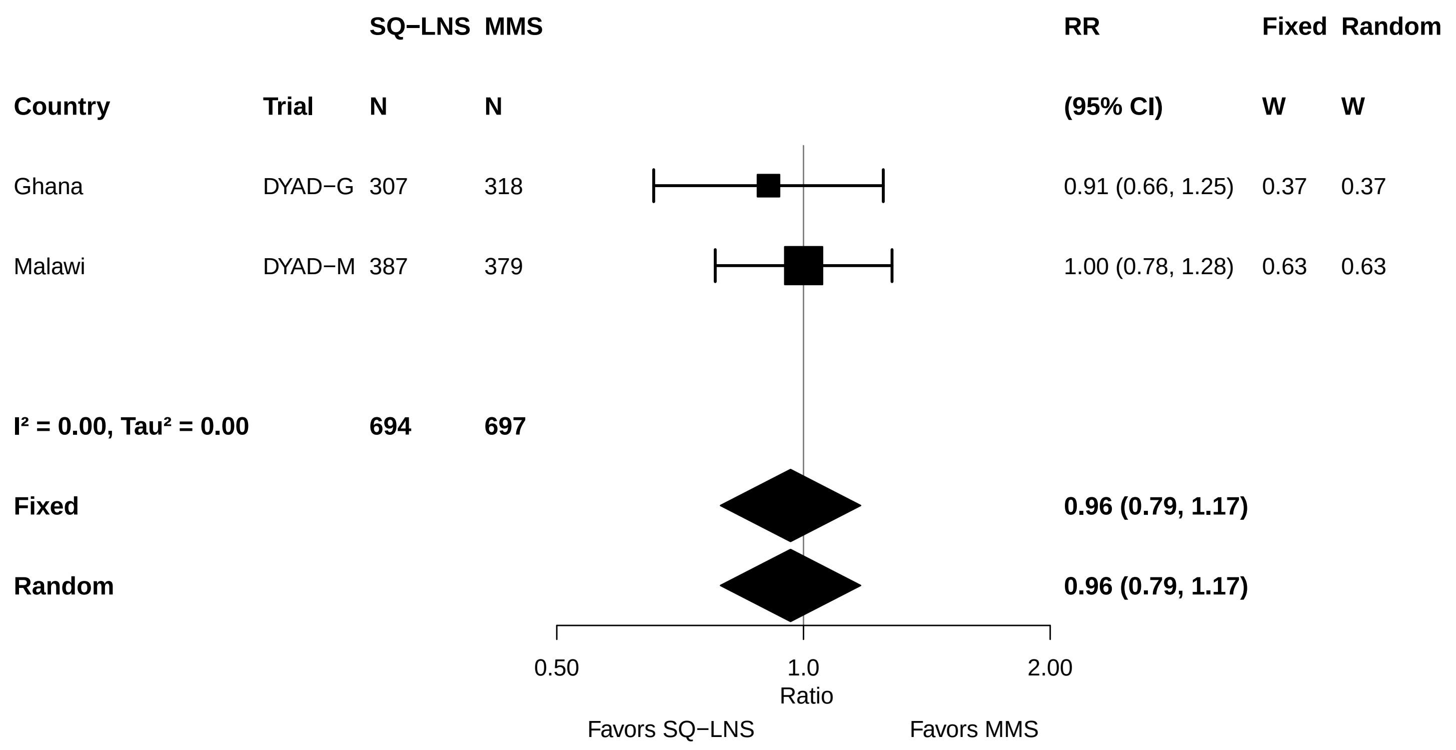

#### Supplemental figure 5I: Small-for-gestational age risk difference

| Country | Trial | SQ−LNS N | MMS N | RD (95% CI) | Fixed W | Random W |
|---|---|---|---|---|---|---|
| Ghana | DYAD−G | 307 | 318 | −0.02 (−0.08, 0.04) | 0.49 | 0.49 |
| Malawi | DYAD−M | 387 | 379 | 0.00 (−0.06, 0.06) | 0.51 | 0.51 |
| **I² = 0.00, Tau² = 0.00** | | **694** | **697** | | | |
| **Fixed** | | | | **−0.01 (−0.05, 0.03)** | | |
| **Random** | | | | **−0.01 (−0.05, 0.03)** | | |

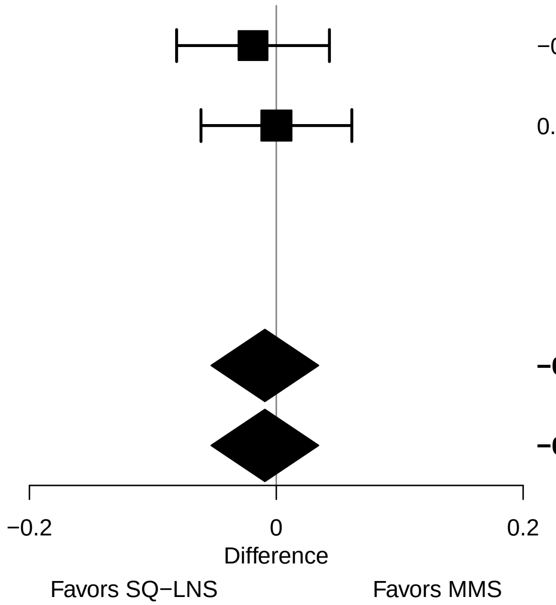

#### Supplemental figure 5J: Large-for-gestational age relative risk

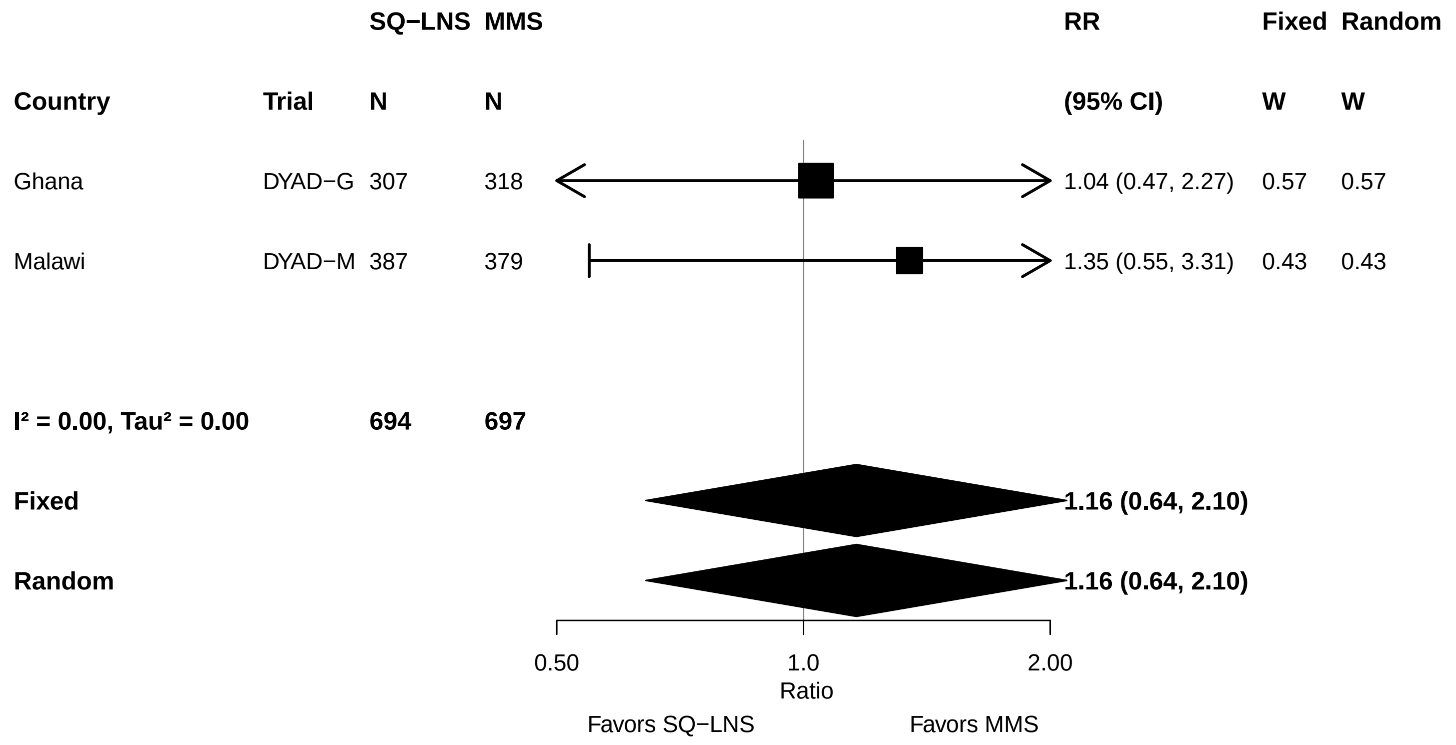

#### Supplemental figure 5K: Large-for-gestational age risk difference

| Country | Trial | SQ−LNS N | MMS N | RD (95% CI) | Fixed W | Random W |
|---|---|---|---|---|---|---|
| Ghana | DYAD−G | 307 | 318 | 0.00 (−0.03, 0.03) | 0.35 | 0.35 |
| Malawi | DYAD−M | 387 | 379 | 0.01 (−0.01, 0.03) | 0.65 | 0.65 |
| **I² = 0.00, Tau² = 0.00** | | **694** | **697** | | | |
| **Fixed** | | | | **0.01 (−0.01, 0.02)** | | |
| **Random** | | | | **0.01 (−0.01, 0.02)** | | |

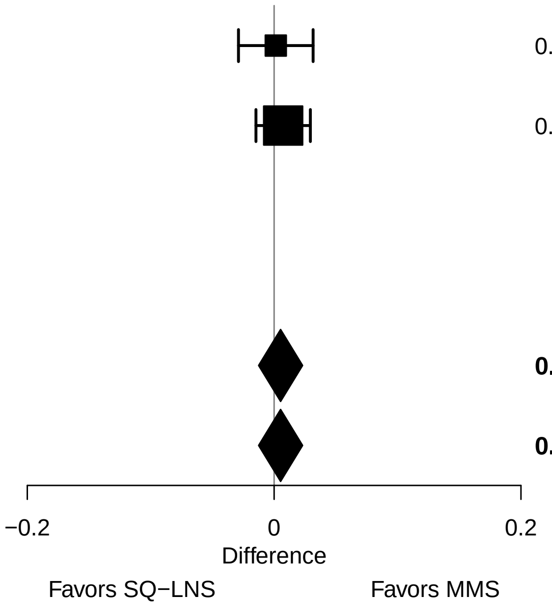

### Supplemental figure 5L: Mean difference in birth length

| Country | Trial | SQ−LNS N | MMS N | MD (95% CI) | Fixed W | Random W |
|---|---|---|---|---|---|---|
| Ghana | DYAD−G | 307 | 318 | 0.18 (−0.17, 0.52) | 0.50 | 0.50 |
| Malawi | DYAD−M | 253 | 259 | 0.13 (−0.21, 0.47) | 0.50 | 0.50 |
| **I² = 0.00, Tau² = 0.00** | | **560** | **577** | | | |
| **Fixed** | | | | **0.16 (−0.09, 0.40)** | | |
| **Random** | | | | **0.16 (−0.09, 0.40)** | | |

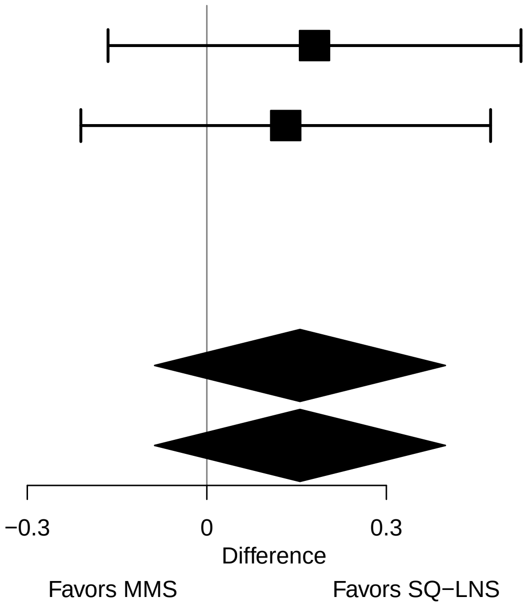

#### Supplemental figure 5M: Mean difference in length-for-age z score

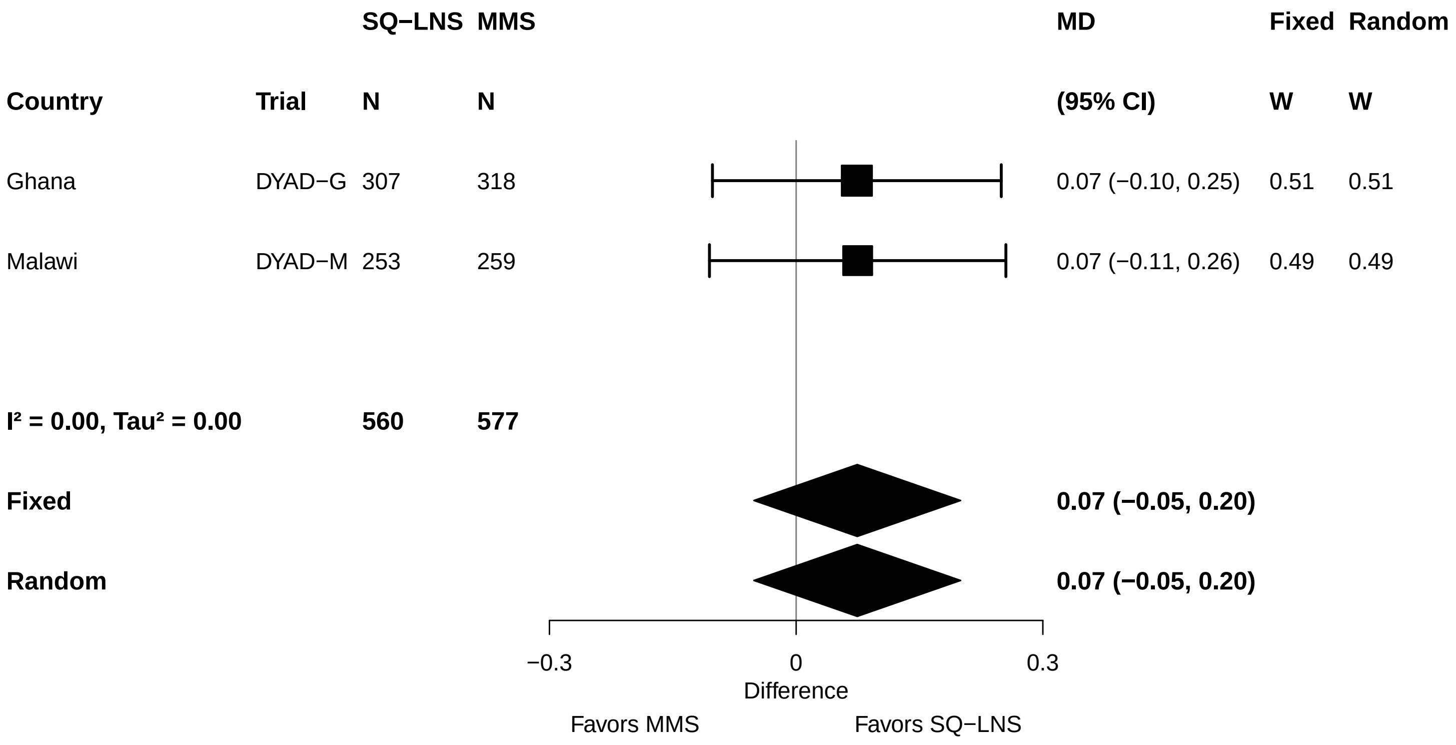

### Supplemental figure 5N: Mean difference in length-for-gestational age z-score

| Country | Trial | SQ−LNS N | MMS N | MD (95% CI) | Fixed W | Random W |
|---|---|---|---|---|---|---|
| Ghana | DYAD−G | 298 | 313 | 0.07 (−0.09, 0.23) | 0.55 | 0.55 |
| Malawi | DYAD−M | 253 | 259 | 0.09 (−0.09, 0.27) | 0.45 | 0.45 |
| **I² = 0.00, Tau² = 0.00** | | **551** | **572** | | | |
| **Fixed** | | | | **0.08 (−0.04, 0.20)** | | |
| **Random** | | | | **0.08 (−0.04, 0.20)** | | |

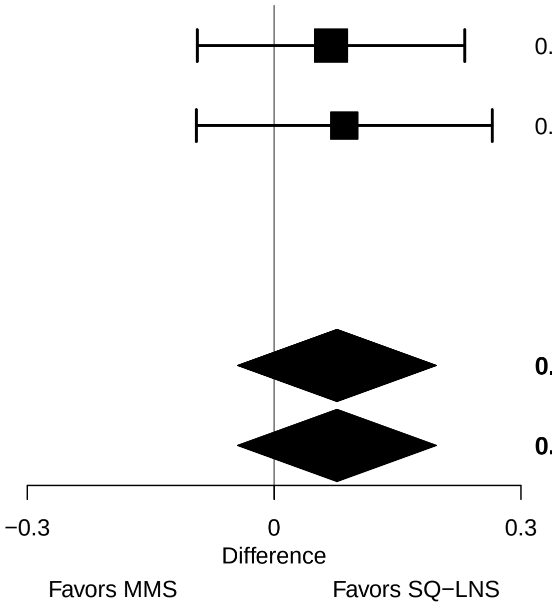

### Supplemental figure 5O: Newborn stunting relative risk

| Country | Trial | SQ−LNS N | MMS N | RR (95% CI) | Fixed W | Random W |
|---|---|---|---|---|---|---|
| Ghana | DYAD−G | 307 | 318 | 0.97 (0.61, 1.56) | 0.47 | 0.47 |
| Malawi | DYAD−M | 253 | 259 | 1.15 (0.74, 1.79) | 0.53 | 0.53 |
| **I² = 0.00, Tau² = 0.00** | | **560** | **577** | | | |
| **Fixed** | | | | **1.06 (0.77, 1.47)** | | |
| **Random** | | | | **1.06 (0.77, 1.47)** | | |

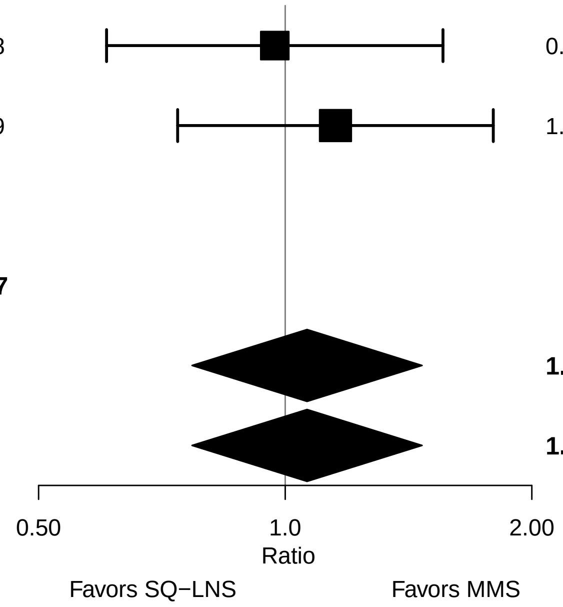

### Supplemental figure 5P: Newborn stunting risk difference

| Country | Trial | SQ−LNS N | MMS N | RD (95% CI) | Fixed W | Random W |
|---|---|---|---|---|---|---|
| Ghana | DYAD−G | 307 | 318 | 0.00 (−0.05, 0.04) | 0.61 | 0.61 |
| Malawi | DYAD−M | 253 | 259 | 0.02 (−0.04, 0.08) | 0.39 | 0.39 |
| **I² = 0.00, Tau² = 0.00** | | **560** | **577** | | | |
| **Fixed** | | | | **0.01 (−0.03, 0.04)** | | |
| **Random** | | | | **0.01 (−0.03, 0.04)** | | |

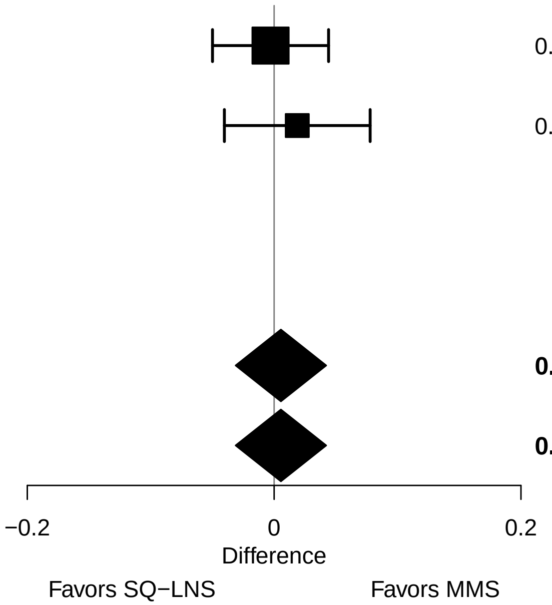

### Supplemental figure 5Q: Low LGAZ relative risk

| Country | Trial | SQ−LNS N | MMS N | RR (95% CI) | Fixed W | Random W |
|---|---|---|---|---|---|---|
| Ghana | DYAD−G | 307 | 318 | 1.04 (0.55, 1.95) | 0.31 | 0.31 |
| Malawi | DYAD−M | 253 | 259 | 1.31 (0.85, 2.01) | 0.69 | 0.69 |
| **I² = 0.00, Tau² = 0.00** | | **560** | **577** | | | |
| **Fixed** | | | | **1.22 (0.85, 1.74)** | | |
| **Random** | | | | **1.22 (0.85, 1.74)** | | |

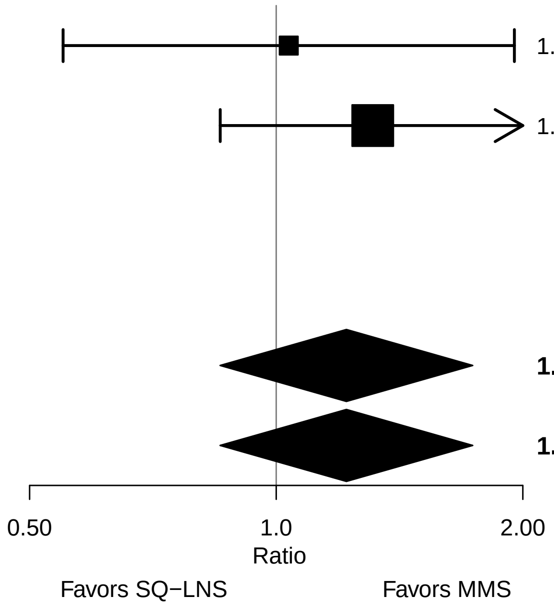

#### Supplemental figure 5R: Low LGAZ risk difference

| Country | Trial | SQ−LNS N | MMS N | RD (95% CI) | Fixed W | Random W |
|---|---|---|---|---|---|---|
| Ghana | DYAD−G | 307 | 318 | 0.00 (−0.03, 0.04) | 0.73 | 0.73 |
| Malawi | DYAD−M | 253 | 259 | 0.04 (−0.02, 0.10) | 0.27 | 0.27 |
| **I² = 0.01, Tau² = 0.00** | | **560** | **577** | | | |
| **Fixed** | | | | **0.01 (−0.02, 0.04)** | | |
| **Random** | | | | **0.01 (−0.02, 0.04)** | | |

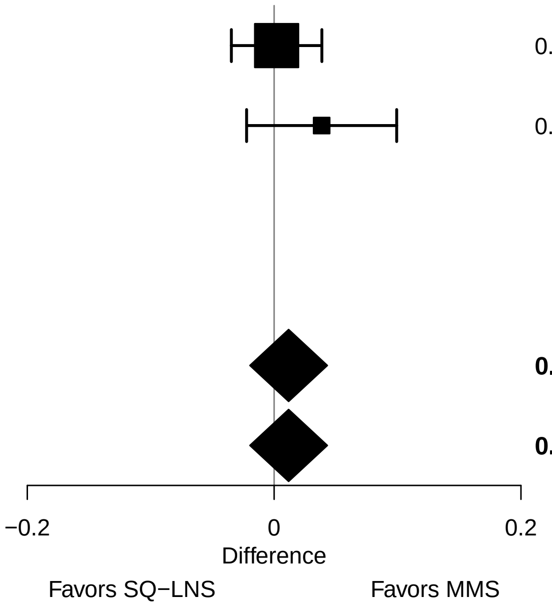

#### Supplemental figure 5S: Mean difference in BMI-for-age z-score

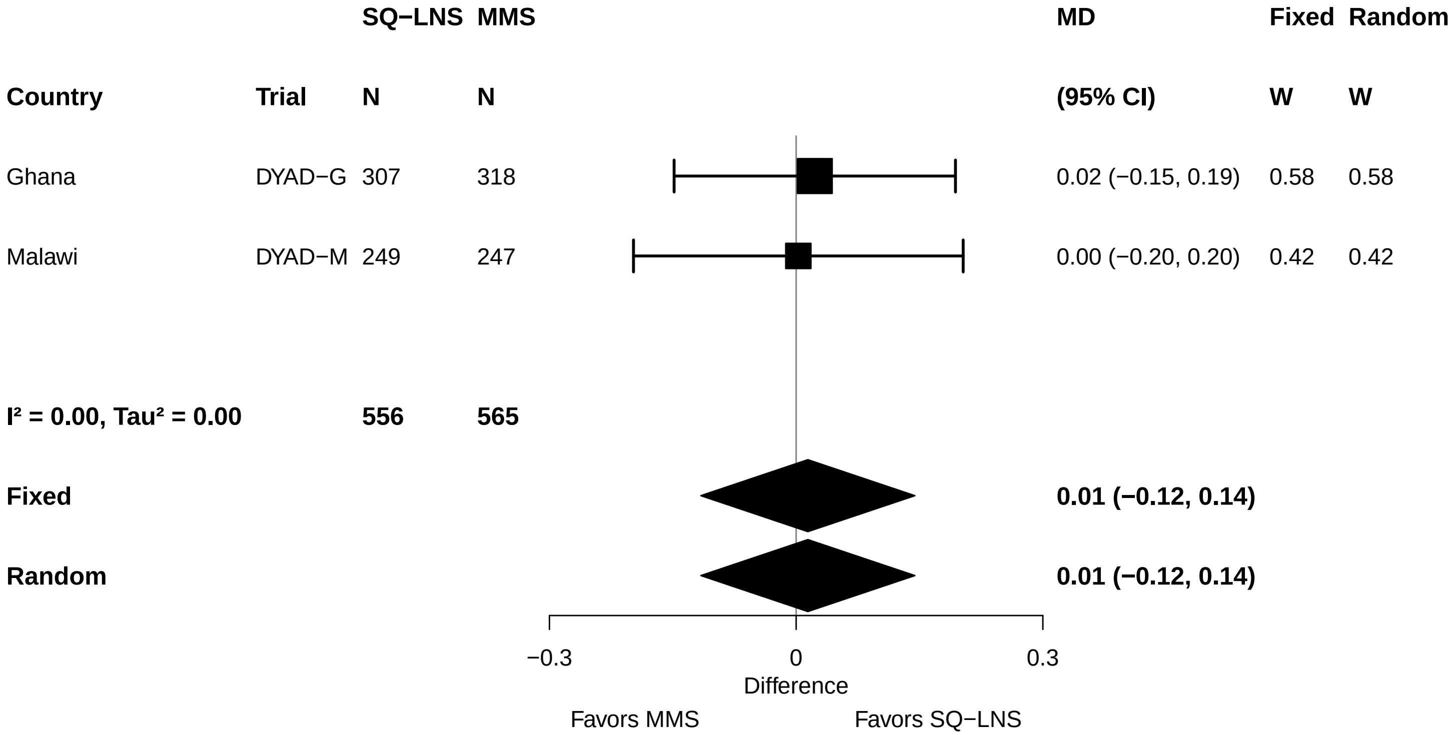

### Supplemental figure 5T: Low BMIZ relative risk

| Country | Trial | SQ−LNS N | MMS N | RR (95% CI) | Fixed W | Random W |
|---|---|---|---|---|---|---|
| Ghana | DYAD−G | 307 | 318 | 0.78 (0.43, 1.40) | 0.54 | 0.53 |
| Malawi | DYAD−M | 249 | 247 | 1.32 (0.69, 2.52) | 0.46 | 0.47 |
| **I² = 0.30, Tau² = 0.04** | | **556** | **565** | | | |
| **Fixed** | | | | **0.99 (0.64, 1.53)** | | |
| **Random** | | | | **1.00 (0.59, 1.68)** | | |

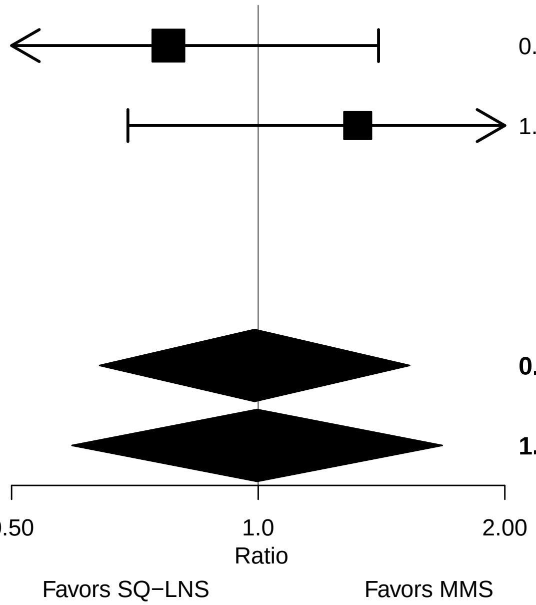

### Supplemental figure 5U: Low BMIZ risk difference

| Country | Trial | SQ−LNS N | MMS N | RD (95% CI) | Fixed W | Random W |
|---|---|---|---|---|---|---|
| Ghana | DYAD−G | 307 | 318 | −0.02 (−0.06, 0.02) | 0.57 | 0.55 |
| Malawi | DYAD−M | 249 | 247 | 0.02 (−0.03, 0.06) | 0.43 | 0.45 |
| **I² = 0.30, Tau² = 0.00** | | **556** | **565** | | | |
| **Fixed** | | | | **0.00 (−0.03, 0.03)** | | |
| **Random** | | | | **0.00 (−0.04, 0.03)** | | |

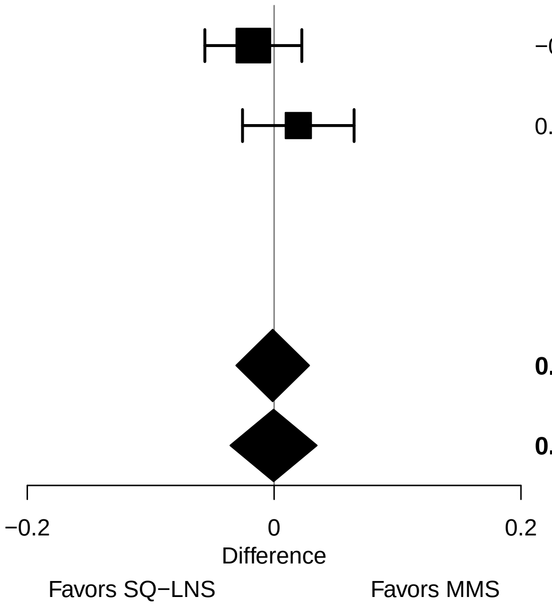

#### Supplemental figure 5V: Mean difference in head circumference

| Country | Trial | SQ−LNS N | MMS N | MD (95% CI) | Fixed W | Random W |
|---|---|---|---|---|---|---|
| Ghana | DYAD−G | 307 | 318 | 0.24 (0.00, 0.48) | 0.48 | 0.48 |
| Malawi | DYAD−M | 253 | 260 | 0.06 (−0.17, 0.29) | 0.52 | 0.52 |
| **I² = 0.08, Tau² = 0.00** | | **560** | **578** | | | |
| **Fixed** | | | | **0.14 (−0.02, 0.31)** | | |
| **Random** | | | | **0.14 (−0.03, 0.32)** | | |

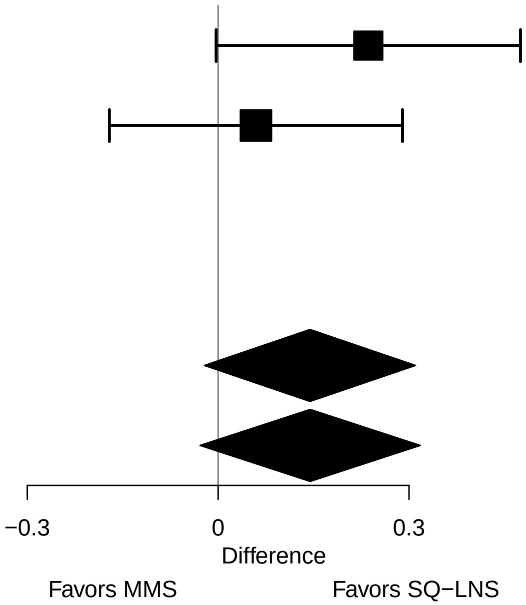

### Supplemental figure 5W: Mean difference in head circumference-for-age z score

| Country | Trial | SQ−LNS N | MMS N | MD (95% CI) | Fixed W | Random W |
|---|---|---|---|---|---|---|
| Ghana | DYAD−G | 307 | 318 | 0.16 (−0.03, 0.34) | 0.50 | 0.50 |
| Malawi | DYAD−M | 253 | 260 | 0.06 (−0.13, 0.24) | 0.50 | 0.50 |
| **I² = 0.00, Tau² = 0.00** | | **560** | **578** | | | |
| **Fixed** | | | | **0.11 (−0.02, 0.24)** | | |
| **Random** | | | | **0.11 (−0.02, 0.24)** | | |

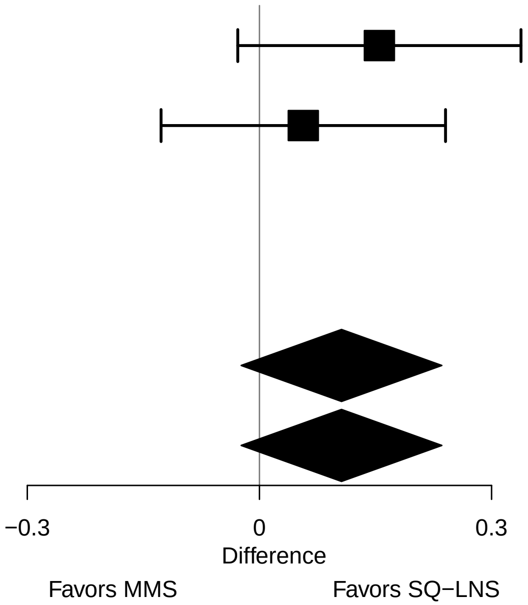

#### Supplemental figure 5X: Mean difference in head circumference-for-gestational age z score

| Country | Trial | SQ−LNS N | MMS N | MD (95% CI) | Fixed W | Random W |
|---|---|---|---|---|---|---|
| Ghana | DYAD−G | 298 | 313 | 0.17 (0.00, 0.34) | 0.51 | 0.51 |
| Malawi | DYAD−M | 253 | 260 | 0.06 (−0.12, 0.23) | 0.49 | 0.49 |
| **I² = 0.00, Tau² = 0.00** | | **551** | **573** | | | |
| **Fixed** | | | | **0.11 (−0.01, 0.23)** | | |
| **Random** | | | | **0.11 (−0.01, 0.23)** | | |

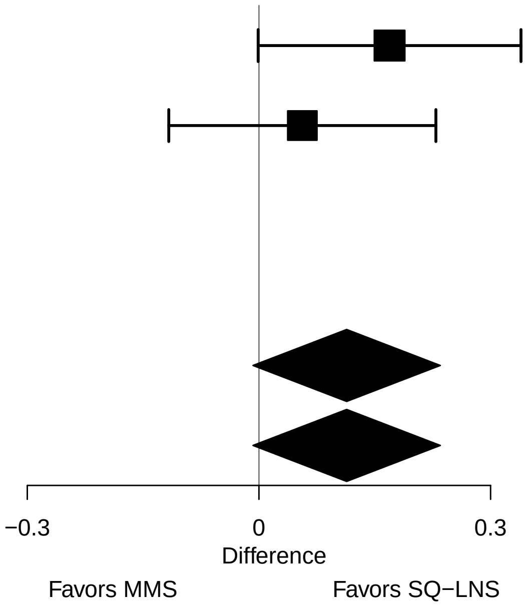

### Supplemental figure 5Y: Low HCZ relative risk

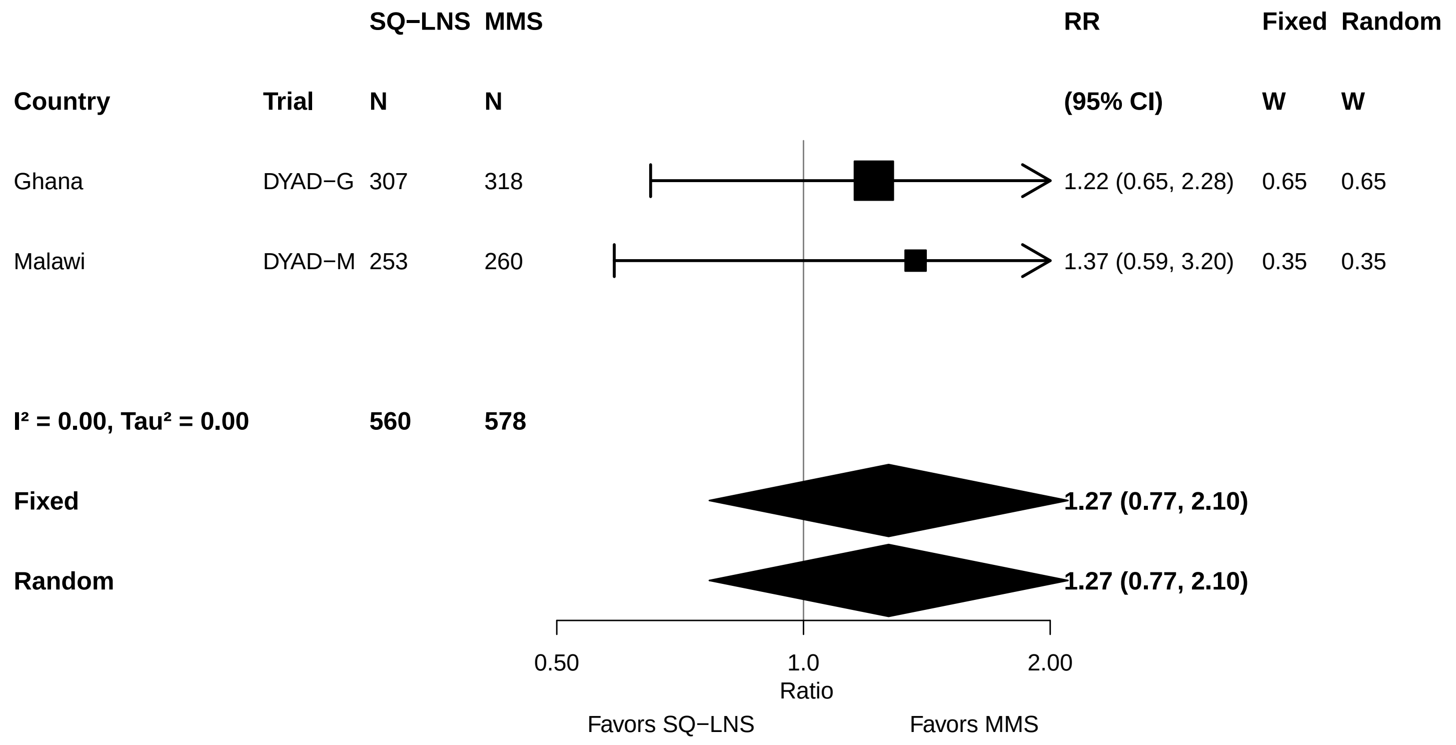

#### Supplemental figure 5Z: Low HCZ risk difference

| Country | Trial | SQ−LNS N | MMS N | RD (95% CI) | Fixed W | Random W |
|---|---|---|---|---|---|---|
| Ghana | DYAD−G | 307 | 318 | 0.01 (−0.03, 0.05) | 0.46 | 0.46 |
| Malawi | DYAD−M | 253 | 260 | 0.01 (−0.02, 0.05) | 0.54 | 0.54 |
| **I² = 0.00, Tau² = 0.00** | | **560** | **578** | | | |
| **Fixed** | | | | **0.01 (−0.01, 0.04)** | | |
| **Random** | | | | **0.01 (−0.01, 0.04)** | | |

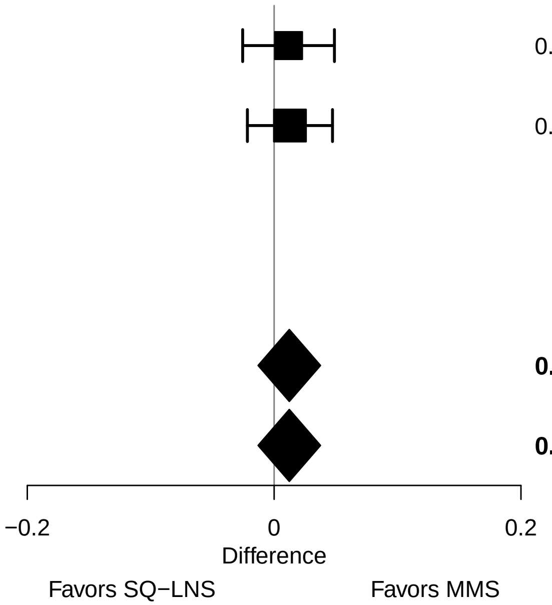

#### Supplemental figure 5AA: Low HCGAZ relative risk

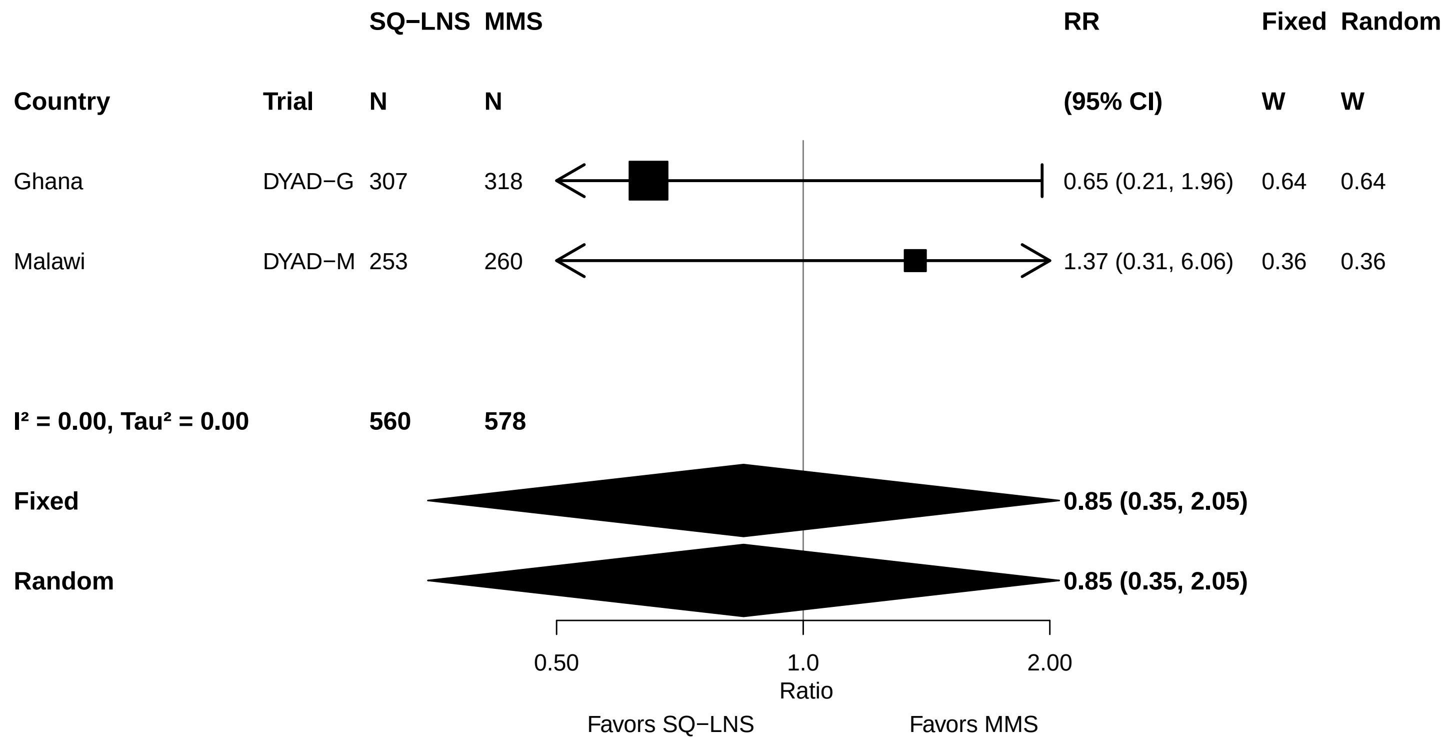

### Supplemental figure 5AB: Low HCGAZ risk difference

| Country | Trial | SQ−LNS N | MMS N | RD (95% CI) | Fixed W | Random W |
|---|---|---|---|---|---|---|
| Ghana | DYAD−G | 307 | 318 | −0.01 (−0.03, 0.01) | 0.45 | 0.45 |
| Malawi | DYAD−M | 253 | 260 | 0.00 (−0.02, 0.02) | 0.55 | 0.55 |
| **$I^2$ = 0.00, $Tau^2$ = 0.00** | | **560** | **578** | | | |
| **Fixed** | | | | **0.00 (−0.02, 0.01)** | | |
| **Random** | | | | **0.00 (−0.02, 0.01)** | | |

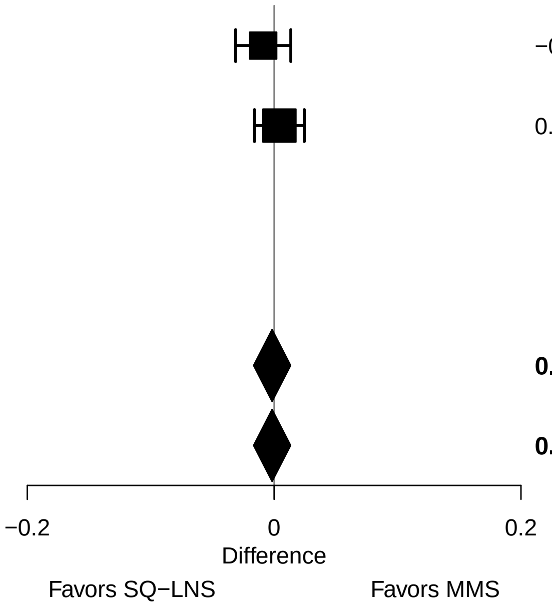

#### Supplemental figure 5AC: Mean difference in mid-upper arm circumference

| Country | Trial | SQ−LNS N | MMS N | MD (95% CI) | Fixed W | Random W |
|---|---|---|---|---|---|---|
| Ghana | DYAD−G | 307 | 318 | 0.10 (−0.05, 0.24) | 0.52 | 0.52 |
| Malawi | DYAD−M | 256 | 261 | 0.06 (−0.09, 0.21) | 0.48 | 0.48 |
| **I² = 0.00, Tau² = 0.00** | | **563** | **579** | | | |
| **Fixed** | | | | **0.08 (−0.02, 0.18)** | | |
| **Random** | | | | **0.08 (−0.02, 0.18)** | | |

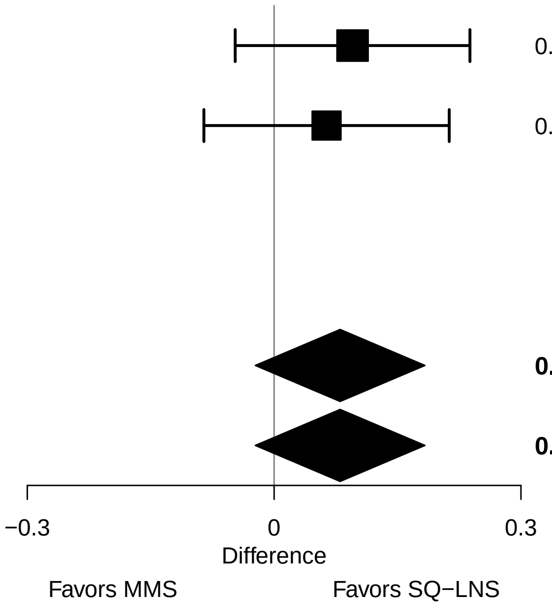

#### Supplemental figure 5AD: Mean difference in duration of gestation

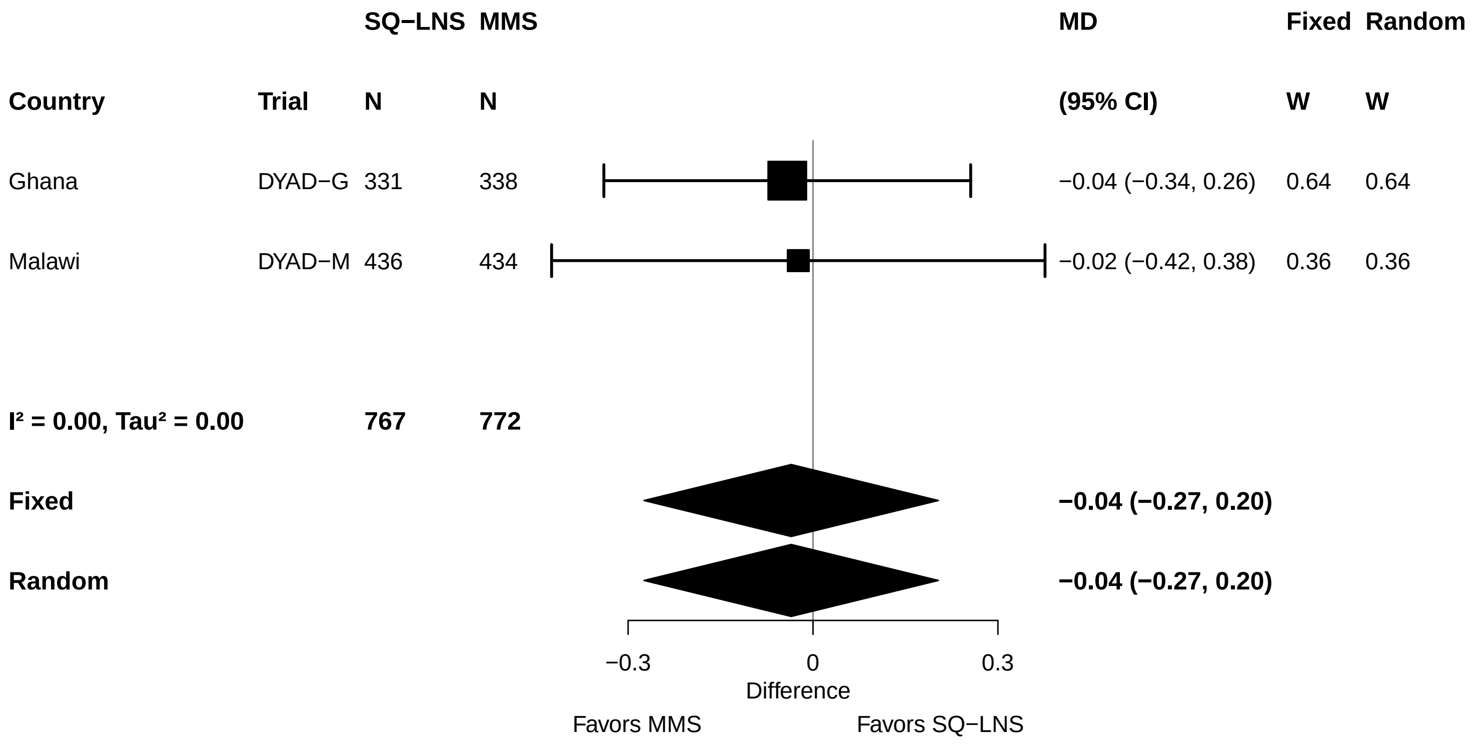

#### Supplemental figure 5AE: Preterm birth relative risk

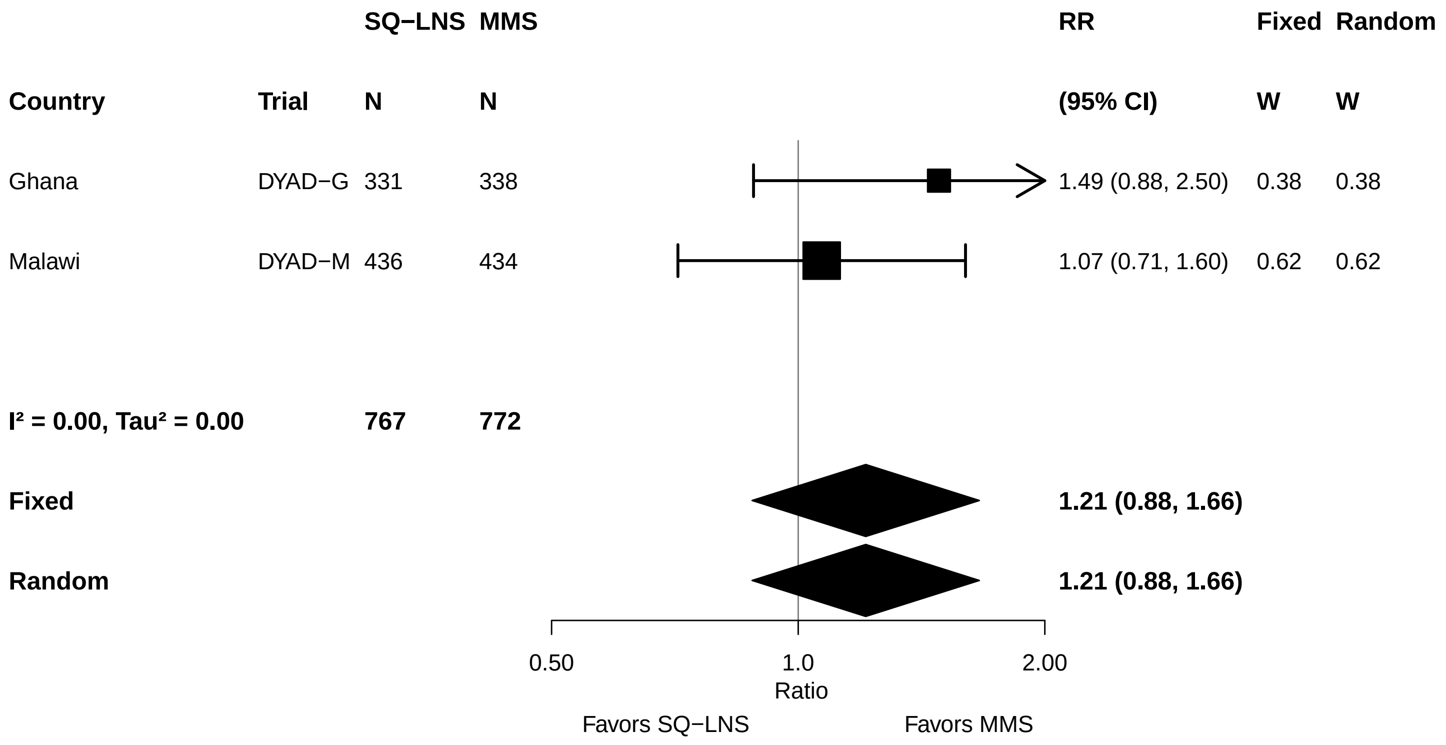

#### Supplemental figure 5AF: Preterm birth risk difference

| Country | Trial | SQ−LNS N | MMS N | RD (95% CI) | Fixed W | Random W |
|---|---|---|---|---|---|---|
| Ghana | DYAD−G | 331 | 338 | 0.03 (−0.01, 0.07) | 0.48 | 0.48 |
| Malawi | DYAD−M | 436 | 434 | 0.01 (−0.03, 0.05) | 0.52 | 0.52 |
| **I² = 0.00, Tau² = 0.00** | | **767** | **772** | | | |
| **Fixed** | | | | **0.02 (−0.01, 0.05)** | | |
| **Random** | | | | **0.02 (−0.01, 0.05)** | | |

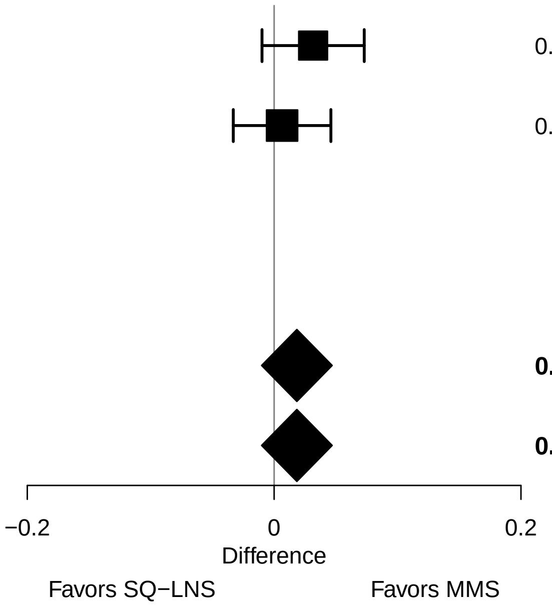

### Supplemental figure 6: Pooled plots for infant outcomes at birth and at 6 mo, stratified by potential effect modifiers, SQ-LNS vs MMS

These figures show pooled effects of SQ-LNS within study-level and individual-level characteristic subgroups along with the p-for-interaction. For definitions of effect modifiers, see Box 1 in the main paper. Individual study estimates were generated from log-binomial regression for dichotomous outcomes and simple linear regression for continuous outcomes with clustered observations using robust standard errors for cluster-randomized trials. Pooled sub-group estimates and statistical testing of the pooled interaction term were generated using inverse-variance weighting. For continuous outcomes the intervention effect is measured by the difference in mean of the SQ-LNS group minus MMS. For dichotomous outcomes analyzed via prevalence/risk ratios, the effect estimate is the prevalence/risk in the SQ-LNS group divided by the prevalence/risk in the MMS group. For dichotomous outcomes analyzed via prevalence/risk differences, the effect estimate is the prevalence/risk in the SQ-LNS group minus the prevalence/risk in the IMMS group. The labels on the left y-axis correspond to the characteristic subgroups and their sample sizes. The values on the right indicate the pooled prevalence ratio and confidence interval within that subgroup.

LAZ, length-for-age z-score; WLZ, weight-for-length z-score; WAZ, weight for-age z-score; MUACZ, mid-upper arm circumference z-score; BMI, body mass index; HCZ, head circumference-for-age z-score; LGAZ, length-for-gestational-age z-score; HCGAZ, head circumference-for-gestational-age z-score; BMIZ, body mass index-for-age z-score; IFA/SOC, Iron and folic acid or standard of care; MD, mean difference; MMS, multiple micronutrient supplement; MUAC, mid-upper arm circumference; PR, prevalence ratio; PD, prevalence difference; RD, risk difference; RR, relative risk; SOC, standard of care; SQ-LNS, small-quantity lipid-based nutrient supplements; WGAZ, weight-for-gestational age z-score.

### Supplemental figure 6A: Sex

#### 6A1: Mean differences for birth outcomes

| Outcome (p−for−interaction) | SQ−LNS N | MMS N | Mean difference (95% CI) |
|---|---|---|---|
| Weight (kg) (p=0.503) | | | |
| Male | 344 | 329 | 0.01 (−0.06, 0.08) |
| Female | 350 | 368 | 0.04 (−0.02, 0.10) |
| Weight−for−age z score (p=0.393) | | | |
| Male | 344 | 329 | 0.01 (−0.14, 0.17) |
| Female | 350 | 368 | 0.11 (−0.04, 0.26) |
| Weight−for−gestational age z−score (p=0.606) | | | |
| Male | 341 | 327 | 0.03 (−0.12, 0.18) |
| Female | 344 | 365 | 0.08 (−0.06, 0.23) |
| Birth length (cm) (p=0.116) | | | |
| Male | 274 | 273 | −0.06 (−0.39, 0.28) |
| Female | 286 | 304 | 0.33 (−0.01, 0.66) |
| Length−for−age z−score (p=0.120) | | | |
| Male | 274 | 273 | −0.03 (−0.21, 0.14) |
| Female | 286 | 304 | 0.17 (−0.01, 0.35) |
| Length−for−gestational age z−score (p=0.128) | | | |
| Male | 271 | 271 | −0.02 (−0.19, 0.15) |
| Female | 280 | 301 | 0.17 (0.00, 0.33) |
| BMI z−score (p=0.852) | | | |
| Male | 272 | 268 | 0.00 (−0.18, 0.18) |
| Female | 284 | 297 | 0.03 (−0.16, 0.21) |
| Head circumference (cm) (p=0.097) | | | |
| Male | 275 | 274 | −0.01 (−0.22, 0.21) |
| Female | 285 | 304 | 0.26 (0.03, 0.50) |
| Head circumference−for−age z−score (p=0.098) | | | |
| Male | 275 | 274 | −0.01 (−0.18, 0.16) |
| Female | 285 | 304 | 0.21 (0.01, 0.40) |
| Head circumference−for−gestational age z score (p=0.177) | | | |
| Male | 272 | 272 | 0.03 (−0.14, 0.20) |
| Female | 279 | 301 | 0.19 (0.02, 0.37) |
| Mid−upper arm circumference (cm) (p=0.758) | | | |
| Male | 277 | 274 | 0.05 (−0.09, 0.20) |
| Female | 286 | 305 | 0.09 (−0.06, 0.24) |
| Duration (wk) (p=0.195) | | | |
| Male | 376 | 363 | −0.16 (−0.46, 0.15) |
| Female | 376 | 394 | 0.13 (−0.16, 0.42) |

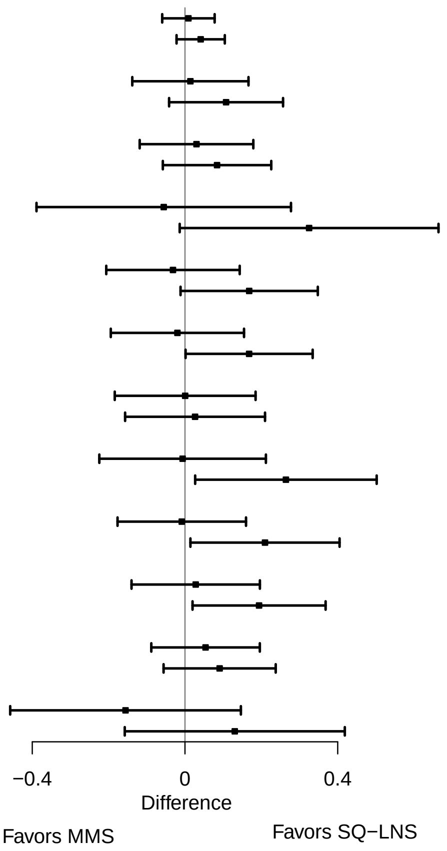

**Supplemental figure 6A: Sex**

#### 6A2: Relative risks for birth outcomes

| Outcome (p−for−interaction) | SQ−LNS N | MMS N | Relative risk (95% CI) |
|---|---|---|---|
| Low birth weight (p=0.301) | | | |
| Male | 344 | 329 | 1.10 (0.67, 1.82) |
| Female | 350 | 368 | 0.80 (0.56, 1.14) |
| Birth weight < 2 kg (p=0.079) | | | |
| Male | 344 | 329 | 2.10 (0.74, 5.95) |
| Female | 350 | 368 | 0.48 (0.15, 1.57) |
| Small−for−gestational age (p=0.094) | | | |
| Male | 344 | 329 | 1.13 (0.86, 1.47) |
| Female | 350 | 368 | 0.80 (0.60, 1.08) |
| Large−for−gestational age (p=0.204) | | | |
| Male | 344 | 329 | 1.73 (0.69, 4.34) |
| Female | 350 | 368 | 0.82 (0.36, 1.84) |
| Newborn stunting (p=0.128) | | | |
| Male | 274 | 273 | 1.46 (0.87, 2.44) |
| Female | 286 | 304 | 0.86 (0.56, 1.32) |
| Low LGAZ (p=0.414) | | | |
| Male | 274 | 273 | 1.44 (0.87, 2.39) |
| Female | 286 | 304 | 1.04 (0.63, 1.73) |
| Low BMIZ (p=0.006) | | | |
| Male | 272 | 268 | 2.04 (0.98, 4.26) |
| Female | 284 | 297 | 0.64 (0.35, 1.17) |
| Low HCZ (p=0.097) | | | |
| Male | 275 | 274 | 2.55 (0.91, 7.14) |
| Female | 285 | 304 | 0.93 (0.50, 1.70) |
| Preterm birth (p=0.159) | | | |
| Male | 376 | 363 | 1.50 (0.89, 2.55) |
| Female | 376 | 394 | 0.97 (0.61, 1.52) |

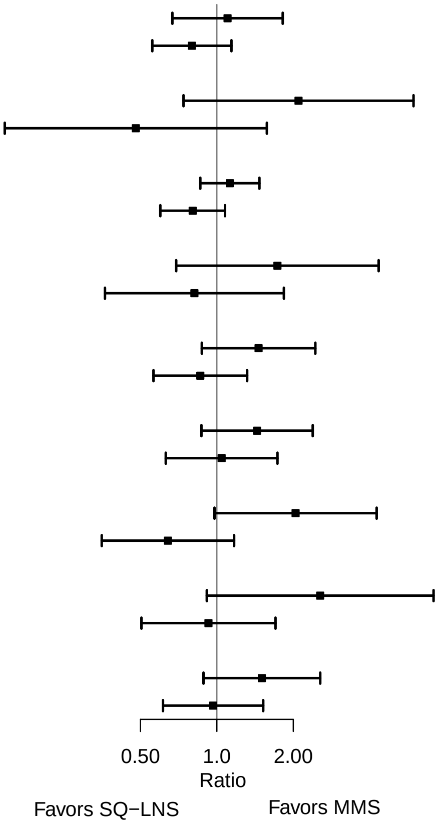

**Supplemental figure 6A: Sex**

#### 6A3: Mean differences for 6 mo outcomes

| Outcome (p−for−interaction) | SQ−LNS N | MMS N | Mean difference (95% CI) |
|---|---|---|---|
| Weight−for−age z score (p=0.185) | | | |
| Male | 313 | 310 | −0.10 (−0.29, 0.09) |
| Female | 318 | 351 | 0.07 (−0.10, 0.23) |
| Length−for−age z−score (p=0.451) | | | |
| Male | 313 | 309 | −0.02 (−0.20, 0.15) |
| Female | 319 | 348 | 0.07 (−0.09, 0.22) |
| Weight−for−length z−score (p=0.269) | | | |
| Male | 313 | 309 | −0.11 (−0.29, 0.08) |
| Female | 318 | 348 | 0.03 (−0.13, 0.20) |
| Head circumference−for−age z−score (p=0.912) | | | |
| Male | 313 | 310 | −0.02 (−0.17, 0.14) |
| Female | 319 | 351 | −0.03 (−0.17, 0.12) |
| MUAC−for−age z−score (p=0.549) | | | |
| Male | 313 | 310 | −0.06 (−0.23, 0.11) |
| Female | 319 | 351 | 0.02 (−0.14, 0.18) |

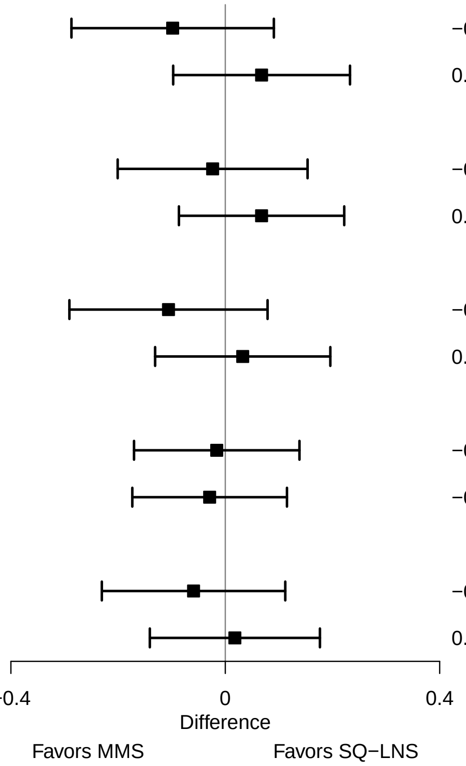

**Supplemental figure 6A: Sex**

#### 6A4: Prevalence ratios for 6 mo outcomes

| Outcome (p−for−interaction) | SQ−LNS N | MMS N | Prevalence ratio (95% CI) |
|---|---|---|---|
| Underweight (p=0.092) | | | |
| Male | 313 | 310 | 1.47 (0.91, 2.38) |
| Female | 318 | 351 | 0.75 (0.41, 1.36) |
| Stunted (p=0.869) | | | |
| Male | 313 | 309 | 1.02 (0.75, 1.38) |
| Female | 319 | 348 | 1.07 (0.72, 1.57) |
| Wasted (p=0.392) | | | |
| Male | 313 | 309 | 1.32 (0.46, 3.76) |
| Female | 318 | 348 | 0.82 (0.29, 2.34) |
| Low HCAZ (p=0.157) | | | |
| Male | 313 | 310 | 0.99 (0.59, 1.67) |
| Female | 319 | 351 | 0.55 (0.29, 1.03) |
| Low MUAC (p=0.626) | | | |
| Male | 313 | 310 | 0.76 (0.34, 1.71) |
| Female | 319 | 351 | 1.01 (0.65, 1.58) |
| Acute malnutrition (p=0.663) | | | |
| Male | 313 | 309 | 0.86 (0.41, 1.77) |
| Female | 318 | 348 | 1.07 (0.69, 1.65) |

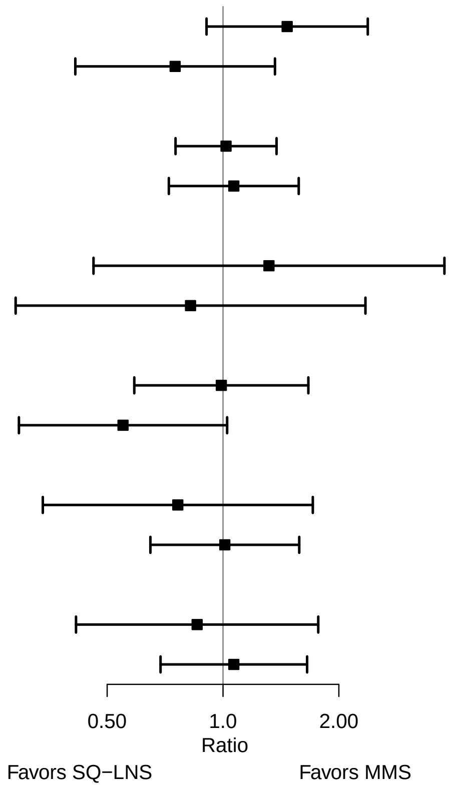

### Supplemental figure 6B: Birth order

#### 6B1: Mean differences for birth outcomes

| Outcome (p−for−interaction) | SQ−LNS N | MMS N | Mean difference (95% CI) |
|---|---|---|---|
| Weight (kg) (p=0.033) | | | |
| Later born | 511 | 521 | 0.00 (−0.05, 0.06) |
| Firstborn | 183 | 175 | 0.12 (0.04, 0.21) |
| Weight−for−age z score (p=0.022) | | | |
| Later born | 511 | 521 | 0.00 (−0.13, 0.12) |
| Firstborn | 183 | 175 | 0.28 (0.08, 0.48) |
| Weight−for−gestational age z−score (p=0.362) | | | |
| Later born | 508 | 516 | 0.03 (−0.09, 0.15) |
| Firstborn | 177 | 175 | 0.15 (−0.04, 0.34) |
| Birth length (cm) (p=0.209) | | | |
| Later born | 412 | 430 | 0.07 (−0.20, 0.34) |
| Firstborn | 148 | 146 | 0.39 (−0.10, 0.89) |
| Length−for−age z−score (p=0.182) | | | |
| Later born | 412 | 430 | 0.03 (−0.11, 0.17) |
| Firstborn | 148 | 146 | 0.21 (−0.04, 0.47) |
| Length−for−gestational age z−score (p=0.353) | | | |
| Later born | 409 | 425 | 0.05 (−0.09, 0.19) |
| Firstborn | 142 | 146 | 0.18 (−0.06, 0.41) |
| BMI z−score (p=0.260) | | | |
| Later born | 410 | 421 | −0.03 (−0.18, 0.12) |
| Firstborn | 146 | 143 | 0.17 (−0.09, 0.42) |
| Head circumference (cm) (p=0.138) | | | |
| Later born | 412 | 431 | 0.07 (−0.12, 0.26) |
| Firstborn | 148 | 146 | 0.39 (0.05, 0.74) |
| Head circumference−for−age z−score (p=0.109) | | | |
| Later born | 412 | 431 | 0.05 (−0.10, 0.19) |
| Firstborn | 148 | 146 | 0.31 (0.05, 0.58) |
| Head circumference−for−gestational age z score (p=0.253) | | | |
| Later born | 409 | 426 | 0.07 (−0.07, 0.21) |
| Firstborn | 142 | 146 | 0.26 (0.02, 0.50) |
| Mid−upper arm circumference (cm) (p=0.205) | | | |
| Later born | 415 | 432 | 0.04 (−0.08, 0.16) |
| Firstborn | 148 | 146 | 0.20 (0.01, 0.38) |
| Duration (wk) (p=0.140) | | | |
| Later born | 554 | 569 | −0.15 (−0.41, 0.12) |
| Firstborn | 213 | 202 | 0.24 (−0.27, 0.76) |

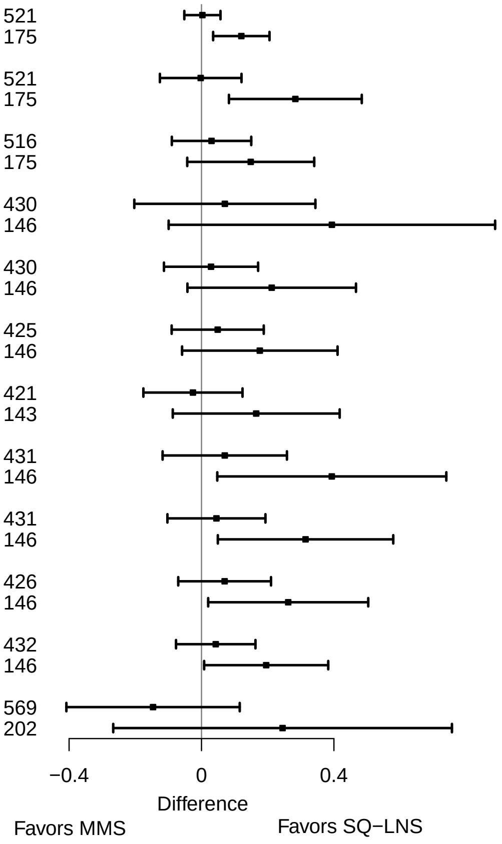

**Supplemental figure 6B: Birth order**

#### 6B2: Relative risks for birth outcomes

| Outcome (p−for−interaction) | SQ−LNS N | MMS N | Relative risk (95% CI) |
|---|---|---|---|
| Low birth weight (p=0.266) | | | |
| Later born | 511 | 521 | 1.00 (0.69, 1.45) |
| Firstborn | 183 | 175 | 0.71 (0.44, 1.15) |
| Birth weight < 2 kg (p=0.881) | | | |
| Later born | 511 | 521 | 1.12 (0.45, 2.76) |
| Firstborn | 183 | 175 | 0.98 (0.28, 3.42) |
| Small−for−gestational age (p=0.396) | | | |
| Later born | 511 | 521 | 1.03 (0.80, 1.31) |
| Firstborn | 183 | 175 | 0.84 (0.60, 1.17) |
| Newborn stunting (p=0.526) | | | |
| Later born | 412 | 430 | 1.16 (0.78, 1.74) |
| Firstborn | 148 | 146 | 0.90 (0.52, 1.57) |
| Low LGAZ (p=0.782) | | | |
| Later born | 412 | 430 | 1.19 (0.78, 1.81) |
| Firstborn | 148 | 146 | 1.26 (0.63, 2.52) |
| Low BMIZ (p=0.596) | | | |
| Later born | 410 | 421 | 1.12 (0.64, 1.98) |
| Firstborn | 146 | 143 | 0.84 (0.38, 1.89) |
| Low HCZ (p=0.181) | | | |
| Later born | 412 | 431 | 1.60 (0.84, 3.04) |
| Firstborn | 148 | 146 | 0.75 (0.31, 1.84) |
| Low HCGAZ (p=0.893) | | | |
| Later born | 412 | 431 | 0.90 (0.30, 2.74) |
| Firstborn | 148 | 146 | 0.74 (0.17, 3.27) |
| Preterm birth (p=0.814) | | | |
| Later born | 554 | 569 | 1.23 (0.82, 1.84) |
| Firstborn | 213 | 202 | 1.13 (0.67, 1.91) |

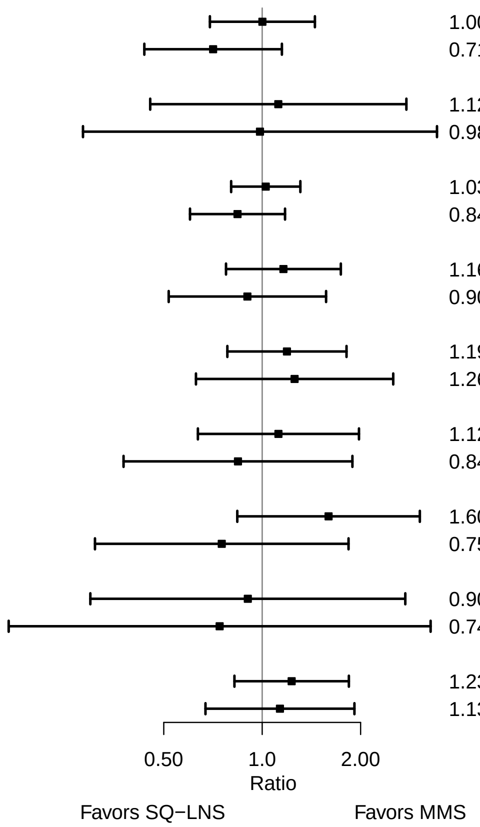

**Supplemental figure 6B: Birth order**

#### 6B3: Mean differences for 6 mo outcomes

| Outcome (p−for−interaction) | SQ−LNS N | MMS N | Mean difference (95% CI) |
|---|---|---|---|
| Weight−for−age z score (p=0.171) | | | |
| Later born | 463 | 500 | −0.07 (−0.21, 0.08) |
| Firstborn | 168 | 160 | 0.13 (−0.12, 0.38) |
| Length−for−age z−score (p=0.037) | | | |
| Later born | 463 | 496 | −0.05 (−0.19, 0.08) |
| Firstborn | 169 | 160 | 0.24 (0.01, 0.46) |
| Weight−for−length z−score (p=0.921) | | | |
| Later born | 463 | 496 | −0.05 (−0.19, 0.09) |
| Firstborn | 168 | 160 | −0.04 (−0.29, 0.22) |
| Head circumference−for−age z−score (p=0.835) | | | |
| Later born | 463 | 500 | −0.03 (−0.15, 0.10) |
| Firstborn | 169 | 160 | −0.05 (−0.25, 0.15) |
| MUAC−for−age z−score (p=0.656) | | | |
| Later born | 463 | 500 | −0.04 (−0.17, 0.09) |
| Firstborn | 169 | 160 | 0.01 (−0.24, 0.25) |

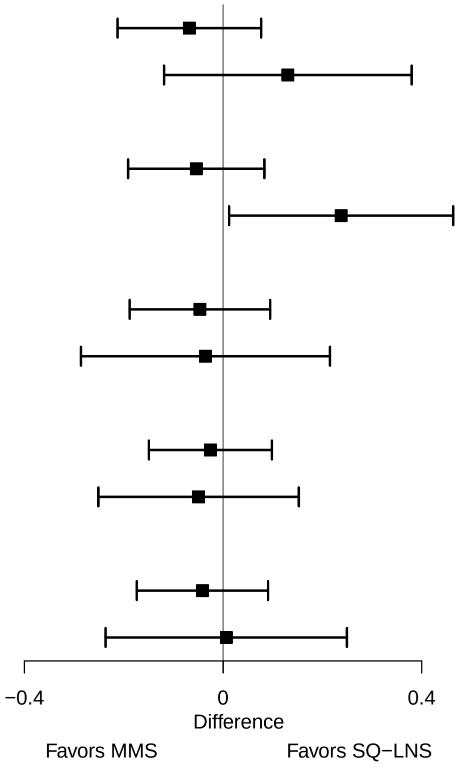

**Supplemental figure 6B: Birth order**

#### 6B4: Prevalence ratios for 6 mo outcomes

| Outcome (p−for−interaction) | SQ−LNS N | MMS N | Prevalence ratio (95% CI) |
|---|---|---|---|
| Underweight (p=0.660) | | | |
| Later born | 463 | 500 | 1.19 (0.79, 1.80) |
| Firstborn | 168 | 160 | 0.96 (0.43, 2.15) |
| Stunted (p=0.769) | | | |
| Later born | 463 | 496 | 1.08 (0.81, 1.42) |
| Firstborn | 169 | 160 | 0.99 (0.62, 1.59) |
| Wasted (p=0.898) | | | |
| Later born | 463 | 496 | 0.98 (0.44, 2.20) |
| Firstborn | 168 | 160 | 0.96 (0.20, 4.66) |
| Low HCAZ (p=0.043) | | | |
| Later born | 463 | 500 | 0.61 (0.39, 0.97) |
| Firstborn | 169 | 160 | 1.64 (0.70, 3.83) |
| Low MUAC (p=0.144) | | | |
| Later born | 463 | 500 | 0.74 (0.46, 1.20) |
| Firstborn | 169 | 160 | 1.44 (0.69, 3.03) |
| Acute malnutrition (p=0.319) | | | |
| Later born | 463 | 496 | 0.85 (0.54, 1.35) |
| Firstborn | 168 | 160 | 1.33 (0.64, 2.75) |

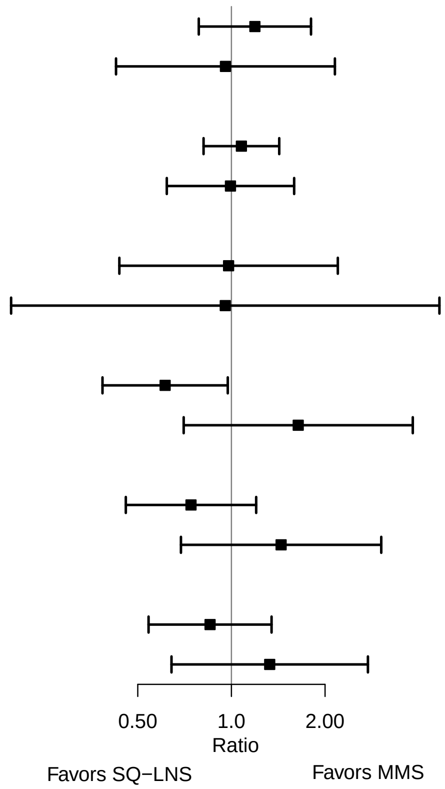

### Supplemental figure 6C: Maternal height

#### 6C1: Mean differences for birth outcomes

| Outcome (p−for−interaction) | SQ−LNS N | MMS N | Mean difference (95% CI) |
|---|---|---|---|
| Weight (kg) (p=0.704) | | | |
| At least 150.1 | 633 | 623 | 0.02 (−0.03, 0.07) |
| Less than 150.1 | 56 | 65 | 0.12 (−0.02, 0.26) |
| Weight−for−age z score (p=0.725) | | | |
| At least 150.1 | 633 | 623 | 0.05 (−0.06, 0.16) |
| Less than 150.1 | 56 | 65 | 0.26 (−0.07, 0.60) |
| Weight−for−gestational age z−score (p=0.850) | | | |
| At least 150.1 | 624 | 618 | 0.05 (−0.06, 0.16) |
| Less than 150.1 | 56 | 65 | 0.11 (−0.20, 0.42) |
| Birth length (cm) (p=0.530) | | | |
| At least 150.1 | 517 | 522 | 0.16 (−0.09, 0.41) |
| Less than 150.1 | 38 | 47 | 0.01 (−0.82, 0.83) |
| Length−for−age z−score (p=0.721) | | | |
| At least 150.1 | 517 | 522 | 0.07 (−0.06, 0.20) |
| Less than 150.1 | 38 | 47 | 0.02 (−0.42, 0.46) |
| Length−for−gestational age z−score (p=0.242) | | | |
| At least 150.1 | 508 | 517 | 0.09 (−0.03, 0.22) |
| Less than 150.1 | 38 | 47 | −0.18 (−0.57, 0.21) |
| BMI z−score (p=0.295) | | | |
| At least 150.1 | 514 | 512 | −0.01 (−0.14, 0.13) |
| Less than 150.1 | 37 | 45 | 0.42 (−0.03, 0.87) |
| Head circumference (cm) (p=0.484) | | | |
| At least 150.1 | 516 | 523 | 0.16 (−0.02, 0.33) |
| Less than 150.1 | 39 | 47 | 0.09 (−0.43, 0.62) |
| Head circumference−for−age z−score (p=0.701) | | | |
| At least 150.1 | 516 | 523 | 0.11 (−0.03, 0.25) |
| Less than 150.1 | 39 | 47 | 0.08 (−0.35, 0.51) |
| Head circumference−for−gestational age z score (p=0.338) | | | |
| At least 150.1 | 507 | 518 | 0.13 (0.00, 0.26) |
| Less than 150.1 | 39 | 47 | −0.11 (−0.48, 0.27) |
| Mid−upper arm circumference (cm) (p=0.548) | | | |
| At least 150.1 | 519 | 524 | 0.09 (−0.02, 0.20) |
| Less than 150.1 | 39 | 47 | 0.15 (−0.21, 0.51) |
| Duration (wk) (p=0.169) | | | |
| At least 150.1 | 696 | 683 | −0.10 (−0.35, 0.14) |
| Less than 150.1 | 66 | 80 | 0.79 (−0.16, 1.74) |

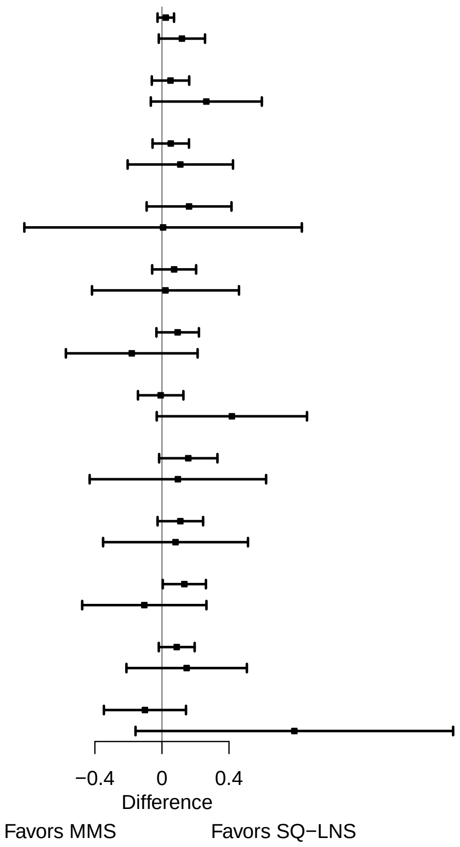

**Supplemental figure 6C: Maternal height**

#### 6C2: Relative risks for birth outcomes

| Outcome (p−for−interaction) | SQ−LNS N | MMS N | Relative risk (95% CI) |
|---|---|---|---|
| Small−for−gestational age (p=0.881) | | | |
| At least 150.1 | 633 | 623 | 0.97 (0.78, 1.19) |
| Less than 150.1 | 56 | 65 | 0.96 (0.54, 1.72) |
| Newborn stunting (p=0.583) | | | |
| At least 150.1 | 517 | 522 | 1.04 (0.74, 1.48) |
| Less than 150.1 | 38 | 47 | 1.42 (0.60, 3.36) |
| Low LGAZ (p=0.228) | | | |
| At least 150.1 | 517 | 522 | 1.14 (0.78, 1.66) |
| Less than 150.1 | 38 | 47 | 2.46 (0.79, 7.65) |
| Preterm birth (p=0.901) | | | |
| At least 150.1 | 696 | 683 | 1.25 (0.89, 1.76) |
| Less than 150.1 | 66 | 80 | 1.07 (0.44, 2.65) |

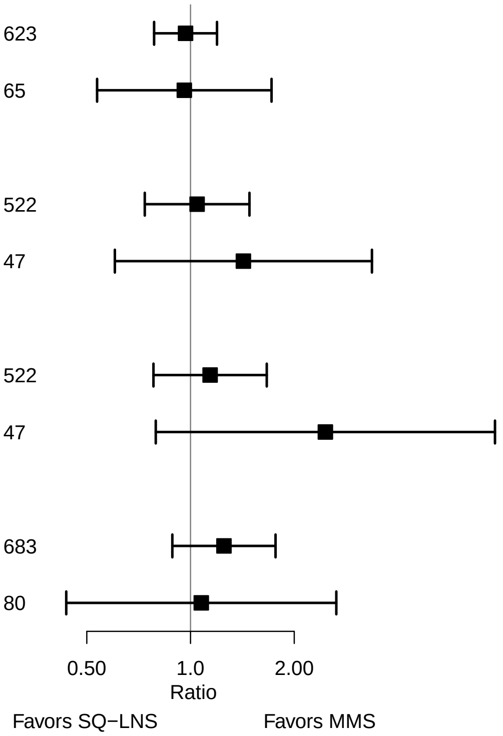

**Supplemental figure 6C: Maternal height**

#### 6C3: Mean differences for 6 mo outcomes

| Outcome (p−for−interaction) | SQ−LNS N | MMS N | Mean difference (95% CI) |
|---|---|---|---|
| Weight−for−age z score (p=0.171) | | | |
| At least 150.1 | 576 | 583 | −0.01 (−0.14, 0.13) |
| Less than 150.1 | 50 | 70 | −0.32 (−0.71, 0.07) |
| Length−for−age z−score (p=0.942) | | | |
| At least 150.1 | 577 | 580 | 0.00 (−0.12, 0.12) |
| Less than 150.1 | 50 | 69 | −0.03 (−0.42, 0.36) |
| Weight−for−length z−score (p=0.051) | | | |
| At least 150.1 | 576 | 580 | −0.01 (−0.14, 0.12) |
| Less than 150.1 | 50 | 69 | −0.44 (−0.83, −0.04) |
| Head circumference−for−age z−score (p=0.450) | | | |
| At least 150.1 | 577 | 583 | −0.01 (−0.12, 0.10) |
| Less than 150.1 | 50 | 70 | −0.21 (−0.53, 0.12) |
| MUAC−for−age z−score (p=0.056) | | | |
| At least 150.1 | 577 | 583 | 0.00 (−0.12, 0.12) |
| Less than 150.1 | 50 | 70 | −0.39 (−0.77, 0.00) |

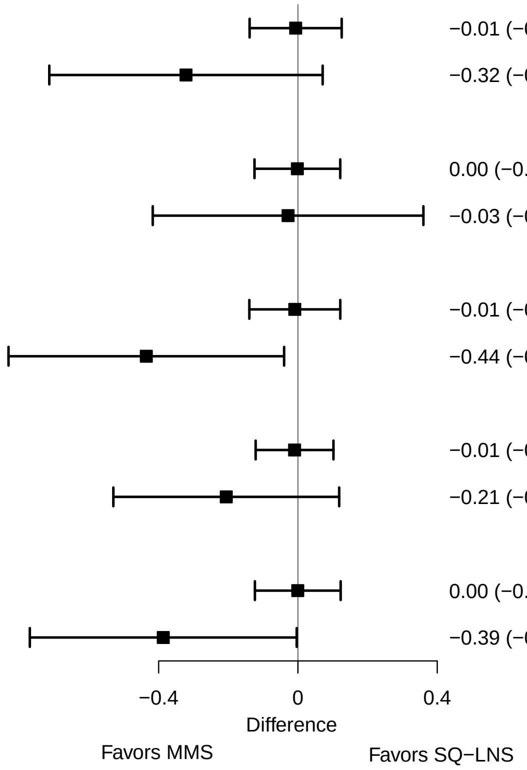

**Supplemental figure 6C: Maternal height**

#### 6C4: Prevalence ratios for 6 mo outcomes

| Outcome (p−for−interaction) | SQ−LNS N | MMS N | Prevalence ratio (95% CI) |
|---|---|---|---|
| Underweight (p=0.329) | | | |
| At least 150.1 | 576 | 583 | 1.06 (0.71, 1.56) |
| Less than 150.1 | 50 | 70 | 1.96 (0.62, 6.14) |
| Stunted (p=0.775) | | | |
| At least 150.1 | 577 | 580 | 1.08 (0.82, 1.43) |
| Less than 150.1 | 50 | 69 | 1.13 (0.73, 1.73) |
| Low MUAC (p=0.848) | | | |
| At least 150.1 | 577 | 583 | 0.94 (0.61, 1.44) |
| Less than 150.1 | 50 | 70 | 1.02 (0.33, 3.14) |
| Acute malnutrition (p=0.760) | | | |
| At least 150.1 | 576 | 580 | 0.99 (0.66, 1.48) |
| Less than 150.1 | 50 | 69 | 1.18 (0.36, 3.87) |

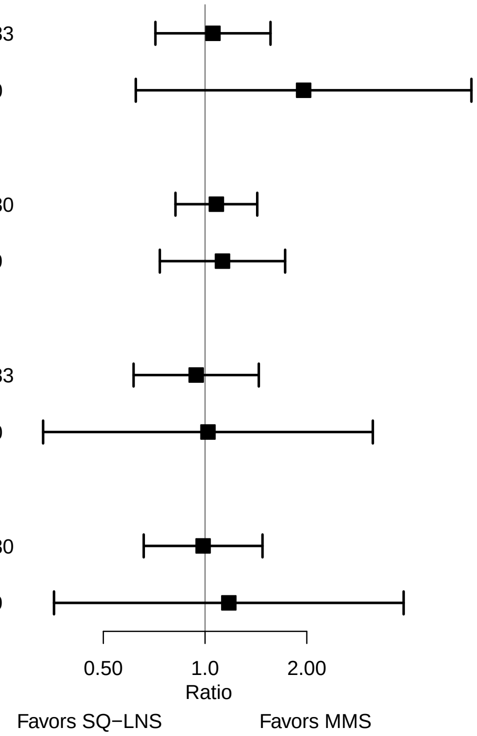

### Supplemental figure 6D: Maternal BMI

#### 6D1: Mean differences for birth outcomes

| Outcome (p−for−interaction) | SQ−LNS N | MMS N | Mean difference (95% CI) |
|---|---|---|---|
| Weight (kg) (p=0.779) | | | |
| At least 20 | 454 | 434 | 0.03 (−0.03, 0.09) |
| Less than 20 | 234 | 253 | 0.02 (−0.06, 0.09) |
| Weight−for−age z score (p=0.840) | | | |
| At least 20 | 454 | 434 | 0.06 (−0.07, 0.19) |
| Less than 20 | 234 | 253 | 0.05 (−0.13, 0.22) |
| Weight−for−gestational age z−score (p=0.686) | | | |
| At least 20 | 447 | 430 | 0.07 (−0.06, 0.20) |
| Less than 20 | 232 | 252 | 0.01 (−0.16, 0.18) |
| Birth length (cm) (p=0.656) | | | |
| At least 20 | 389 | 382 | 0.19 (−0.11, 0.50) |
| Less than 20 | 165 | 186 | 0.09 (−0.32, 0.49) |
| Length−for−age z−score (p=0.587) | | | |
| At least 20 | 389 | 382 | 0.09 (−0.06, 0.25) |
| Less than 20 | 165 | 186 | 0.04 (−0.18, 0.25) |
| Length−for−gestational age z−score (p=0.772) | | | |
| At least 20 | 382 | 378 | 0.08 (−0.06, 0.23) |
| Less than 20 | 163 | 185 | 0.05 (−0.17, 0.27) |
| BMI z−score (p=0.319) | | | |
| At least 20 | 388 | 377 | −0.03 (−0.19, 0.12) |
| Less than 20 | 162 | 179 | 0.11 (−0.12, 0.34) |
| Head circumference (cm) (p=0.730) | | | |
| At least 20 | 387 | 383 | 0.18 (−0.03, 0.39) |
| Less than 20 | 167 | 186 | 0.07 (−0.20, 0.33) |
| Head circumference−for−age z−score (p=0.688) | | | |
| At least 20 | 387 | 383 | 0.13 (−0.03, 0.30) |
| Less than 20 | 167 | 186 | 0.05 (−0.17, 0.26) |
| Head circumference−for−gestational age z score (p=0.744) | | | |
| At least 20 | 380 | 379 | 0.11 (−0.04, 0.26) |
| Less than 20 | 165 | 185 | 0.11 (−0.10, 0.32) |
| Mid−upper arm circumference (cm) (p=0.837) | | | |
| At least 20 | 390 | 383 | 0.09 (−0.04, 0.22) |
| Less than 20 | 167 | 187 | 0.06 (−0.11, 0.22) |
| Duration (wk) (p=0.610) | | | |
| At least 20 | 504 | 481 | −0.02 (−0.31, 0.27) |
| Less than 20 | 257 | 281 | −0.13 (−0.56, 0.30) |

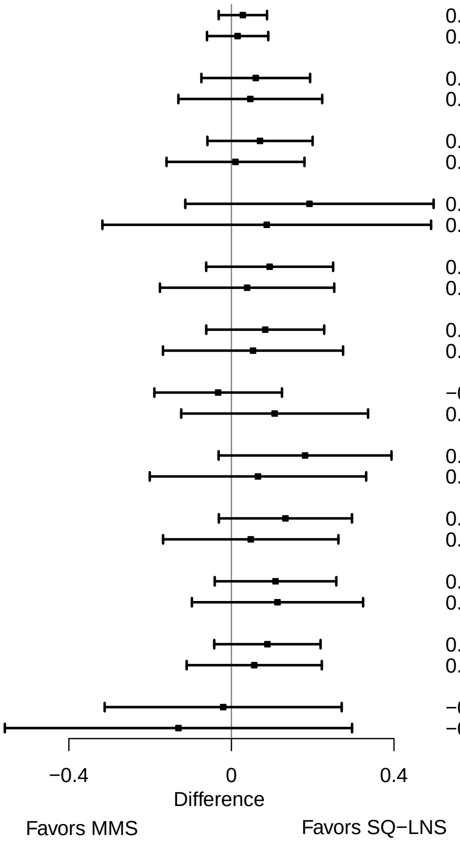

**Supplemental figure 6D: Maternal BMI**

#### 6D2: Relative risks for birth outcomes

| Outcome (p−for−interaction) | SQ−LNS N | MMS N | Relative risk (95% CI) |
|---|---|---|---|
| Low birth weight (p=0.635) | | | |
| At least 20 | 454 | 434 | 0.93 (0.64, 1.36) |
| Less than 20 | 234 | 253 | 0.80 (0.50, 1.28) |
| Small−for−gestational age (p=0.975) | | | |
| At least 20 | 454 | 434 | 0.94 (0.72, 1.23) |
| Less than 20 | 234 | 253 | 0.98 (0.73, 1.33) |
| Newborn stunting (p=0.481) | | | |
| At least 20 | 389 | 382 | 0.98 (0.66, 1.47) |
| Less than 20 | 165 | 186 | 1.28 (0.74, 2.23) |
| Low LGAZ (p=0.571) | | | |
| At least 20 | 389 | 382 | 1.12 (0.69, 1.82) |
| Less than 20 | 165 | 186 | 1.43 (0.84, 2.44) |
| Low BMIZ (p=0.143) | | | |
| At least 20 | 388 | 377 | 1.16 (0.67, 2.04) |
| Less than 20 | 162 | 179 | 0.69 (0.32, 1.49) |
| Low HCZ (p=0.549) | | | |
| At least 20 | 387 | 383 | 1.40 (0.76, 2.56) |
| Less than 20 | 167 | 186 | 1.02 (0.40, 2.63) |
| Low HCGAZ (p=0.764) | | | |
| At least 20 | 387 | 383 | 0.74 (0.26, 2.13) |
| Less than 20 | 167 | 186 | 1.16 (0.22, 6.22) |
| Preterm birth (p=0.471) | | | |
| At least 20 | 504 | 481 | 1.14 (0.77, 1.69) |
| Less than 20 | 257 | 281 | 1.39 (0.79, 2.44) |

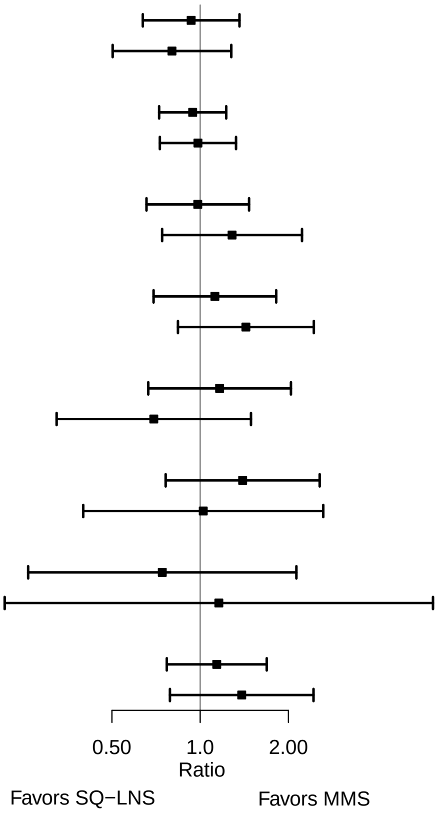

**Supplemental figure 6D: Maternal BMI**

#### 6D3: Mean differences for 6 mo outcomes

| Outcome (p−for−interaction) | SQ−LNS N | MMS N | Mean difference (95% CI) |
|---|---|---|---|
| Weight−for−age z score (p=0.529) | | | |
| At least 20 | 414 | 414 | 0.00 (−0.15, 0.15) |
| Less than 20 | 211 | 238 | −0.08 (−0.29, 0.13) |
| Length−for−age z−score (p=0.824) | | | |
| At least 20 | 414 | 411 | 0.00 (−0.14, 0.15) |
| Less than 20 | 212 | 237 | 0.02 (−0.19, 0.22) |
| Weight−for−length z−score (p=0.295) | | | |
| At least 20 | 414 | 411 | −0.01 (−0.16, 0.14) |
| Less than 20 | 211 | 237 | −0.14 (−0.35, 0.08) |
| Head circumference−for−age z−score (p=0.493) | | | |
| At least 20 | 414 | 414 | −0.03 (−0.16, 0.09) |
| Less than 20 | 212 | 238 | 0.01 (−0.17, 0.20) |
| MUAC−for−age z−score (p=0.070) | | | |
| At least 20 | 414 | 414 | 0.04 (−0.10, 0.18) |
| Less than 20 | 212 | 238 | −0.16 (−0.36, 0.04) |

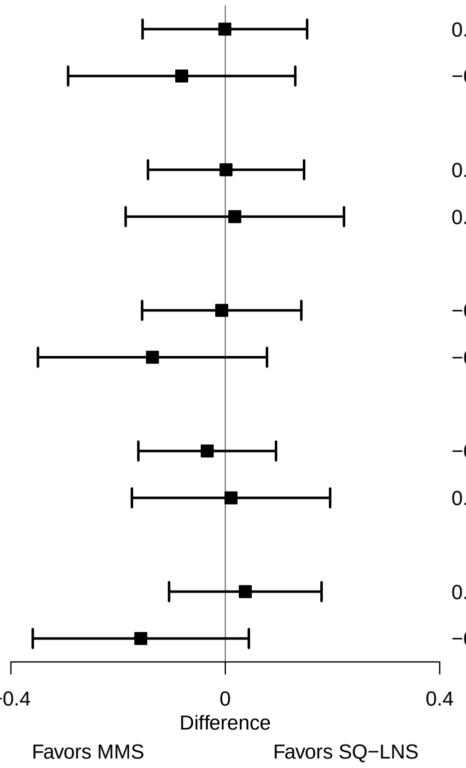

**Supplemental figure 6D: Maternal BMI**

#### 6D4: Prevalence ratios for 6 mo outcomes

| Outcome (p−for−interaction) | SQ−LNS N | MMS N | Prevalence ratio (95% CI) |
|---|---|---|---|
| Underweight (p=0.509) | | | |
| At least 20 | 414 | 414 | 1.01 (0.59, 1.76) |
| Less than 20 | 211 | 238 | 1.32 (0.78, 2.24) |
| Stunted (p=0.872) | | | |
| At least 20 | 414 | 411 | 1.09 (0.79, 1.52) |
| Less than 20 | 212 | 237 | 0.98 (0.68, 1.41) |
| Low HCAZ (p=0.737) | | | |
| At least 20 | 414 | 414 | 0.72 (0.42, 1.25) |
| Less than 20 | 212 | 238 | 0.83 (0.46, 1.52) |
| Low MUAC (p=0.369) | | | |
| At least 20 | 414 | 414 | 0.84 (0.47, 1.50) |
| Less than 20 | 212 | 238 | 1.08 (0.62, 1.89) |
| Acute malnutrition (p=0.327) | | | |
| At least 20 | 414 | 411 | 0.88 (0.49, 1.56) |
| Less than 20 | 211 | 237 | 1.18 (0.70, 1.98) |

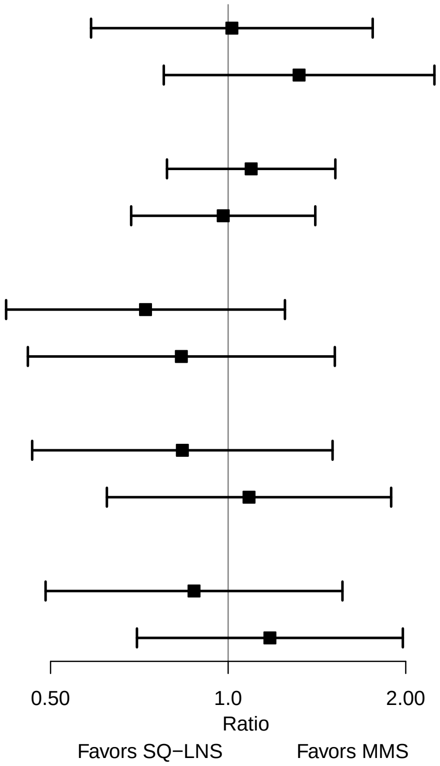

### Supplemental figure 6E: Maternal age

#### 6E1: Mean differences for birth outcomes

| Outcome (p−for−interaction) | SQ−LNS N | MMS N | Mean difference (95% CI) |
|---|---|---|---|
| Weight (kg) (p=0.143) | | | |
| At least 25 | 381 | 396 | 0.00 (−0.06, 0.06) |
| Less than 25 | 313 | 301 | 0.07 (0.00, 0.13) |
| Weight−for−age z score (p=0.130) | | | |
| At least 25 | 381 | 396 | −0.01 (−0.15, 0.14) |
| Less than 25 | 313 | 301 | 0.16 (0.01, 0.31) |
| Weight−for−gestational age z−score (p=0.795) | | | |
| At least 25 | 379 | 392 | 0.05 (−0.09, 0.19) |
| Less than 25 | 306 | 300 | 0.07 (−0.08, 0.22) |
| Birth length (cm) (p=0.222) | | | |
| At least 25 | 327 | 344 | 0.04 (−0.27, 0.35) |
| Less than 25 | 233 | 233 | 0.33 (−0.06, 0.71) |
| Length−for−age z−score (p=0.225) | | | |
| At least 25 | 327 | 344 | 0.01 (−0.15, 0.17) |
| Less than 25 | 233 | 233 | 0.16 (−0.04, 0.36) |
| Length−for−gestational age z−score (p=0.479) | | | |
| At least 25 | 325 | 340 | 0.04 (−0.11, 0.19) |
| Less than 25 | 226 | 232 | 0.13 (−0.06, 0.33) |
| BMI z−score (p=0.892) | | | |
| At least 25 | 326 | 336 | 0.01 (−0.16, 0.18) |
| Less than 25 | 230 | 229 | 0.03 (−0.18, 0.23) |
| Head circumference (cm) (p=0.173) | | | |
| At least 25 | 327 | 345 | 0.06 (−0.16, 0.27) |
| Less than 25 | 233 | 233 | 0.26 (0.00, 0.52) |
| Head circumference−for−age z−score (p=0.191) | | | |
| At least 25 | 327 | 345 | 0.04 (−0.13, 0.21) |
| Less than 25 | 233 | 233 | 0.20 (0.00, 0.40) |
| Head circumference−for−gestational age z score (p=0.347) | | | |
| At least 25 | 325 | 341 | 0.07 (−0.09, 0.23) |
| Less than 25 | 226 | 232 | 0.19 (−0.01, 0.38) |
| Mid−upper arm circumference (cm) (p=0.563) | | | |
| At least 25 | 330 | 345 | 0.06 (−0.08, 0.20) |
| Less than 25 | 233 | 234 | 0.12 (−0.03, 0.26) |
| Duration (wk) (p=0.206) | | | |
| At least 25 | 414 | 425 | −0.17 (−0.48, 0.14) |
| Less than 25 | 353 | 347 | 0.14 (−0.23, 0.52) |

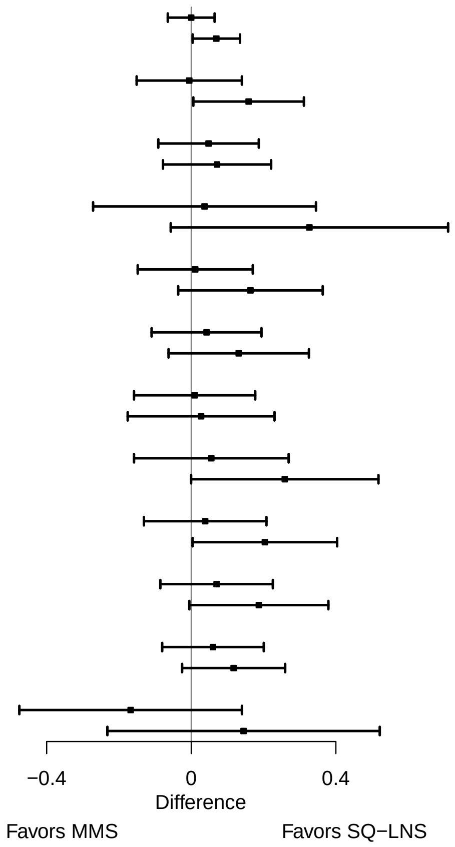

**Supplemental figure 6E: Maternal age**

#### 6E2: Relative risks for birth outcomes

| Outcome (p−for−interaction) | SQ−LNS N | MMS N | Relative risk (95% CI) |
|---|---|---|---|
| Low birth weight (p=0.272) | | | |
| At least 25 | 381 | 396 | 1.03 (0.68, 1.56) |
| Less than 25 | 313 | 301 | 0.75 (0.49, 1.13) |
| Birth weight < 2 kg (p=0.684) | | | |
| At least 25 | 381 | 396 | 1.18 (0.42, 3.28) |
| Less than 25 | 313 | 301 | 0.97 (0.34, 2.76) |
| Small−for−gestational age (p=0.039) | | | |
| At least 25 | 381 | 396 | 1.19 (0.89, 1.59) |
| Less than 25 | 313 | 301 | 0.79 (0.61, 1.03) |
| Large−for−gestational age (p=0.984) | | | |
| At least 25 | 381 | 396 | 1.12 (0.51, 2.46) |
| Less than 25 | 313 | 301 | 1.20 (0.48, 3.02) |
| Newborn stunting (p=0.828) | | | |
| At least 25 | 327 | 344 | 1.09 (0.67, 1.77) |
| Less than 25 | 233 | 233 | 1.04 (0.68, 1.59) |
| Low LGAZ (p=0.680) | | | |
| At least 25 | 327 | 344 | 1.12 (0.68, 1.86) |
| Less than 25 | 233 | 233 | 1.31 (0.80, 2.17) |
| Low BMIZ (p=0.268) | | | |
| At least 25 | 326 | 336 | 1.22 (0.65, 2.29) |
| Less than 25 | 230 | 229 | 0.86 (0.46, 1.60) |
| Low HCZ (p=0.296) | | | |
| At least 25 | 327 | 345 | 1.58 (0.82, 3.06) |
| Less than 25 | 233 | 233 | 0.91 (0.41, 2.02) |
| Preterm birth (p=0.313) | | | |
| At least 25 | 414 | 425 | 1.48 (0.92, 2.38) |
| Less than 25 | 353 | 347 | 0.99 (0.64, 1.54) |

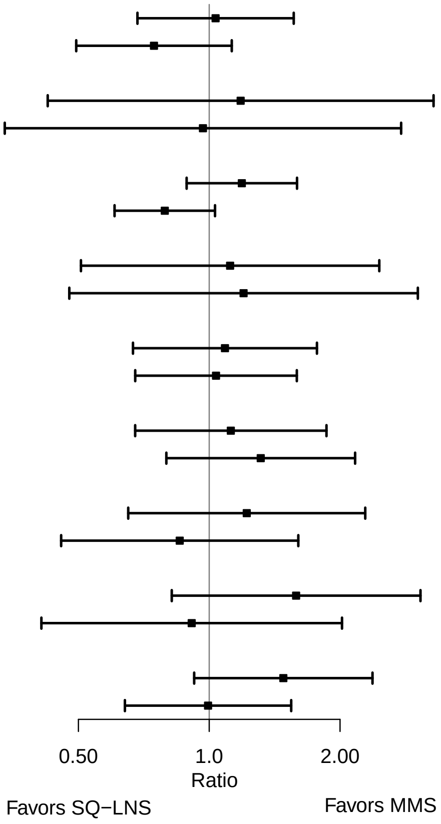

**Supplemental figure 6E: Maternal age**

#### 6E3: Mean differences for 6 mo outcomes

| Outcome (p−for−interaction) | SQ−LNS N | MMS N | Mean difference (95% CI) |
|---|---|---|---|
| Weight−for−age z score (p=0.397) | | | |
| At least 25 | 341 | 379 | −0.06 (−0.23, 0.11) |
| Less than 25 | 290 | 282 | 0.05 (−0.13, 0.23) |
| Length−for−age z−score (p=0.707) | | | |
| At least 25 | 342 | 376 | 0.00 (−0.16, 0.16) |
| Less than 25 | 290 | 281 | 0.04 (−0.13, 0.21) |
| Weight−for−length z−score (p=0.426) | | | |
| At least 25 | 341 | 376 | −0.08 (−0.25, 0.08) |
| Less than 25 | 290 | 281 | 0.02 (−0.16, 0.20) |
| Head circumference−for−age z−score (p=0.404) | | | |
| At least 25 | 342 | 379 | 0.02 (−0.12, 0.16) |
| Less than 25 | 290 | 282 | −0.08 (−0.24, 0.08) |
| MUAC−for−age z−score (p=0.341) | | | |
| At least 25 | 342 | 379 | −0.08 (−0.23, 0.08) |
| Less than 25 | 290 | 282 | 0.04 (−0.13, 0.22) |

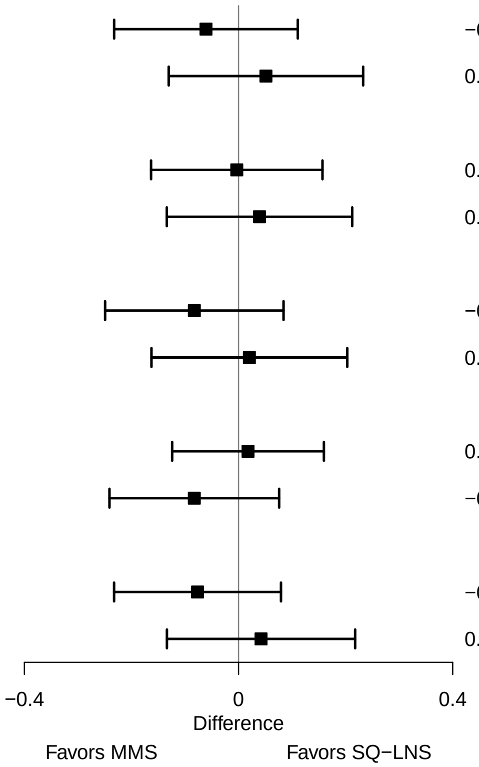

**Supplemental figure 6E: Maternal age**

#### 6E4: Prevalence ratios for 6 mo outcomes

| Outcome (p−for−interaction) | SQ−LNS N | MMS N | Prevalence ratio (95% CI) |
|---|---|---|---|
| Underweight (p=0.651) | | | |
| At least 25 | 341 | 379 | 1.22 (0.77, 1.92) |
| Less than 25 | 290 | 282 | 1.05 (0.56, 1.98) |
| Stunted (p=0.311) | | | |
| At least 25 | 342 | 376 | 1.18 (0.87, 1.60) |
| Less than 25 | 290 | 281 | 0.90 (0.60, 1.33) |
| Wasted (p=0.742) | | | |
| At least 25 | 341 | 376 | 1.00 (0.40, 2.53) |
| Less than 25 | 290 | 281 | 0.97 (0.32, 2.97) |
| Low HCAZ (p=0.044) | | | |
| At least 25 | 342 | 379 | 0.53 (0.30, 0.92) |
| Less than 25 | 290 | 282 | 1.23 (0.68, 2.25) |
| Low MUAC (p=0.432) | | | |
| At least 25 | 342 | 379 | 0.83 (0.50, 1.39) |
| Less than 25 | 290 | 282 | 1.09 (0.58, 2.05) |
| Acute malnutrition (p=0.482) | | | |
| At least 25 | 341 | 376 | 0.90 (0.55, 1.48) |
| Less than 25 | 290 | 281 | 1.12 (0.62, 2.03) |

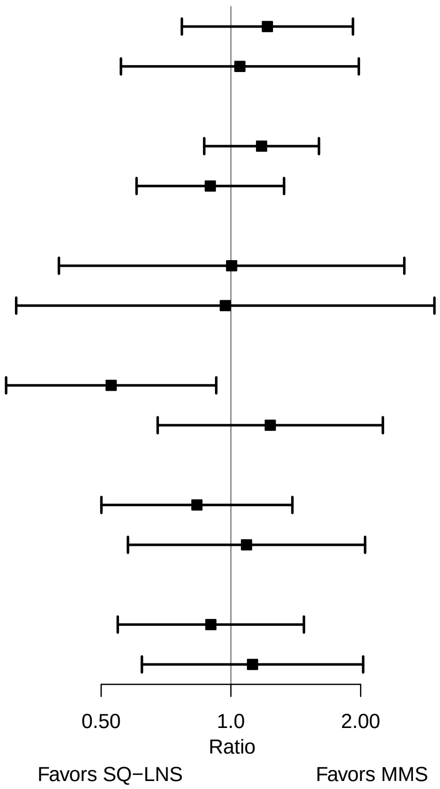

### Supplemental figure 6F: Maternal education

#### 6F1: Mean differences for birth outcomes

| Outcome (p−for−interaction) | SQ−LNS N | MMS N | Mean difference (95% CI) |
|---|---|---|---|
| Weight (kg) (p=0.391) | | | |
| Primary or greater | 305 | 318 | 0.01 (−0.06, 0.08) |
| Incomplete or no formal | 386 | 377 | 0.04 (−0.02, 0.11) |
| Weight−for−age z score (p=0.338) | | | |
| Primary or greater | 305 | 318 | 0.02 (−0.13, 0.18) |
| Incomplete or no formal | 386 | 377 | 0.10 (−0.04, 0.24) |
| Weight−for−gestational age z−score (p=0.729) | | | |
| Primary or greater | 299 | 314 | 0.06 (−0.09, 0.21) |
| Incomplete or no formal | 383 | 376 | 0.06 (−0.08, 0.20) |
| Birth length (cm) (p=0.637) | | | |
| Primary or greater | 287 | 295 | 0.22 (−0.14, 0.58) |
| Incomplete or no formal | 271 | 280 | 0.10 (−0.23, 0.43) |
| Length−for−age z−score (p=0.650) | | | |
| Primary or greater | 287 | 295 | 0.10 (−0.09, 0.28) |
| Incomplete or no formal | 271 | 280 | 0.05 (−0.12, 0.22) |
| Length−for−gestational age z−score (p=0.442) | | | |
| Primary or greater | 281 | 291 | 0.10 (−0.07, 0.27) |
| Incomplete or no formal | 268 | 279 | 0.05 (−0.12, 0.21) |
| BMI z−score (p=0.252) | | | |
| Primary or greater | 286 | 294 | −0.03 (−0.21, 0.16) |
| Incomplete or no formal | 268 | 270 | 0.08 (−0.10, 0.26) |
| Head circumference (cm) (p=0.927) | | | |
| Primary or greater | 287 | 295 | 0.20 (−0.04, 0.45) |
| Incomplete or no formal | 271 | 281 | 0.10 (−0.13, 0.33) |
| Head circumference−for−age z−score (p=0.991) | | | |
| Primary or greater | 287 | 295 | 0.14 (−0.05, 0.33) |
| Incomplete or no formal | 271 | 281 | 0.08 (−0.10, 0.25) |
| Head circumference−for−gestational age z score (p=0.816) | | | |
| Primary or greater | 281 | 291 | 0.17 (−0.01, 0.34) |
| Incomplete or no formal | 268 | 280 | 0.07 (−0.10, 0.24) |
| Mid−upper arm circumference (cm) (p=0.835) | | | |
| Primary or greater | 287 | 295 | 0.10 (−0.05, 0.25) |
| Incomplete or no formal | 274 | 282 | 0.07 (−0.07, 0.21) |
| Duration (wk) (p=0.978) | | | |
| Primary or greater | 329 | 343 | −0.04 (−0.35, 0.27) |
| Incomplete or no formal | 433 | 421 | −0.05 (−0.39, 0.30) |

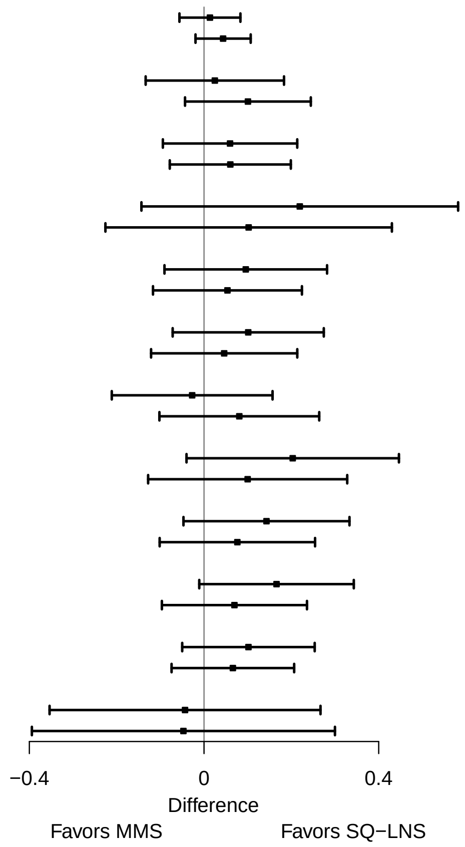

**Supplemental figure 6F: Maternal education**

#### 6F2: Relative risks for birth outcomes

| Outcome (p−for−interaction) | SQ−LNS N | MMS N | Relative risk (95% CI) |
|---|---|---|---|
| Low birth weight (p=0.791) | | | |
| Primary or greater | 305 | 318 | 0.91 (0.58, 1.43) |
| Incomplete or no formal | 386 | 377 | 0.87 (0.59, 1.27) |
| Small−for−gestational age (p=0.623) | | | |
| Primary or greater | 305 | 318 | 0.97 (0.72, 1.31) |
| Incomplete or no formal | 386 | 377 | 0.95 (0.73, 1.24) |
| Large−for−gestational age (p=0.842) | | | |
| Primary or greater | 305 | 318 | 1.04 (0.46, 2.36) |
| Incomplete or no formal | 386 | 377 | 1.32 (0.56, 3.11) |
| Newborn stunting (p=0.448) | | | |
| Primary or greater | 287 | 295 | 0.90 (0.56, 1.46) |
| Incomplete or no formal | 271 | 280 | 1.23 (0.79, 1.90) |
| Low LGAZ (p=0.381) | | | |
| Primary or greater | 287 | 295 | 1.31 (0.69, 2.45) |
| Incomplete or no formal | 271 | 280 | 1.11 (0.72, 1.73) |
| Low BMIZ (p=0.569) | | | |
| Primary or greater | 286 | 294 | 0.91 (0.50, 1.63) |
| Incomplete or no formal | 268 | 270 | 1.06 (0.55, 2.06) |
| Low HCZ (p=0.316) | | | |
| Primary or greater | 287 | 295 | 1.03 (0.52, 2.03) |
| Incomplete or no formal | 271 | 281 | 1.67 (0.77, 3.61) |
| Preterm birth (p=0.708) | | | |
| Primary or greater | 329 | 343 | 1.29 (0.78, 2.15) |
| Incomplete or no formal | 433 | 421 | 1.15 (0.76, 1.76) |

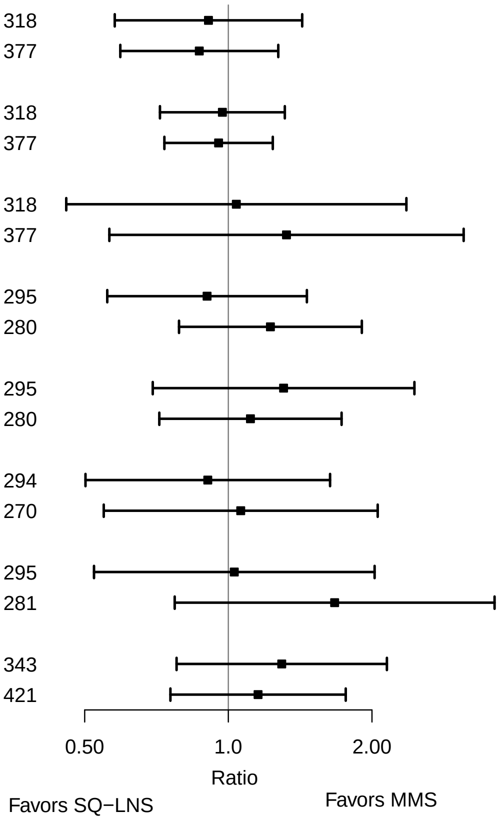

**Supplemental figure 6F: Maternal education**

#### 6F3: Mean differences for 6 mo outcomes

| Outcome (p−for−interaction) | SQ−LNS N | MMS N | Mean difference (95% CI) |
|---|---|---|---|
| Weight−for−age z score (p=0.127) | | | |
| Primary or greater | 277 | 292 | −0.09 (−0.27, 0.09) |
| Incomplete or no formal | 353 | 367 | 0.05 (−0.12, 0.22) |
| Length−for−age z−score (p=0.289) | | | |
| Primary or greater | 278 | 291 | −0.02 (−0.19, 0.15) |
| Incomplete or no formal | 353 | 364 | 0.05 (−0.11, 0.21) |
| Weight−for−length z−score (p=0.210) | | | |
| Primary or greater | 277 | 291 | −0.11 (−0.29, 0.07) |
| Incomplete or no formal | 353 | 364 | 0.03 (−0.14, 0.20) |
| Head circumference−for−age z−score (p=0.295) | | | |
| Primary or greater | 278 | 292 | −0.04 (−0.19, 0.11) |
| Incomplete or no formal | 353 | 367 | 0.00 (−0.14, 0.15) |
| MUAC−for−age z−score (p=0.105) | | | |
| Primary or greater | 278 | 292 | −0.12 (−0.28, 0.05) |
| Incomplete or no formal | 353 | 367 | 0.07 (−0.09, 0.23) |

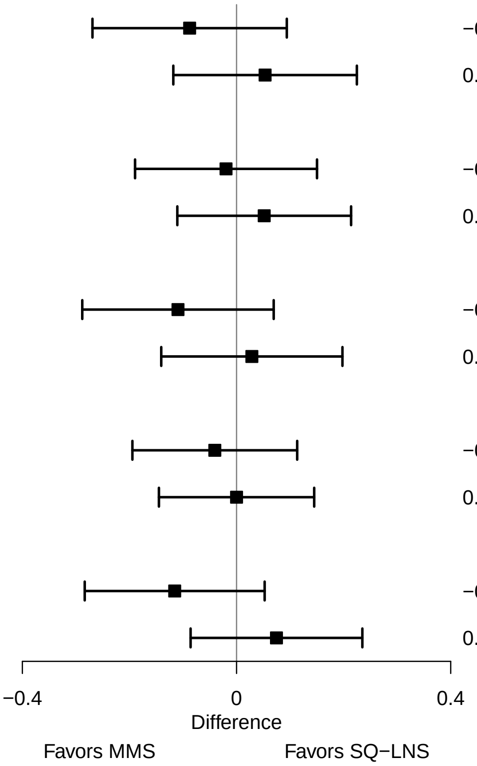

**Supplemental figure 6F: Maternal education**

#### 6F4: Prevalence ratios for 6 mo outcomes

| Outcome (p−for−interaction) | SQ−LNS N | MMS N | Prevalence ratio (95% CI) |
|---|---|---|---|
| Underweight (p=0.931) | | | |
| Primary or greater | 277 | 292 | 1.08 (0.56, 2.11) |
| Incomplete or no formal | 353 | 367 | 1.13 (0.73, 1.76) |
| Stunted (p=0.714) | | | |
| Primary or greater | 278 | 291 | 1.26 (0.78, 2.03) |
| Incomplete or no formal | 353 | 364 | 1.00 (0.75, 1.32) |
| Wasted (p=0.946) | | | |
| Primary or greater | 277 | 291 | 1.26 (0.39, 4.07) |
| Incomplete or no formal | 353 | 364 | 0.83 (0.34, 2.01) |
| Low HCAZ (p=0.703) | | | |
| Primary or greater | 278 | 292 | 0.82 (0.45, 1.50) |
| Incomplete or no formal | 353 | 367 | 0.75 (0.44, 1.27) |
| Acute malnutrition (p=0.709) | | | |
| Primary or greater | 277 | 291 | 0.97 (0.53, 1.79) |
| Incomplete or no formal | 353 | 364 | 0.97 (0.60, 1.58) |

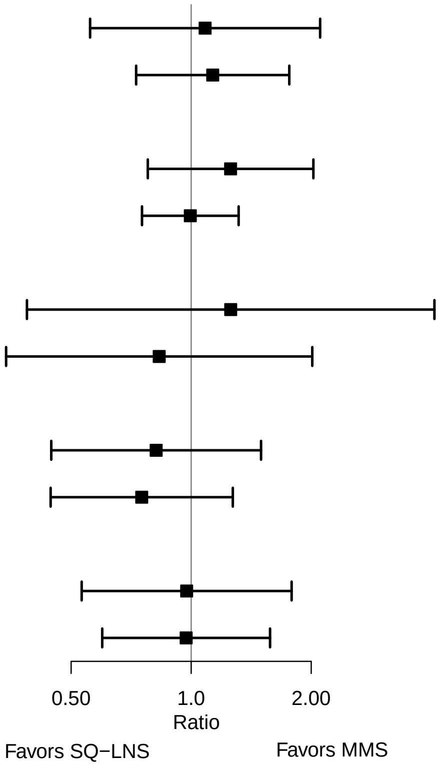

### Supplemental figure 6G: Baseline anemia status

#### 6G1: Mean differences for birth outcomes

| Outcome (p−for−interaction) | SQ−LNS N | MMS N | Mean difference (95% CI) |
|---|---|---|---|
| Weight (kg) (p=0.577) | | | |
| Anemia | 282 | 298 | 0.05 (−0.03, 0.12) |
| No anemia | 412 | 398 | 0.02 (−0.04, 0.08) |
| Weight−for−age z score (p=0.486) | | | |
| Anemia | 282 | 298 | 0.11 (−0.06, 0.29) |
| No anemia | 412 | 398 | 0.03 (−0.10, 0.17) |
| Weight−for−gestational age z−score (p=0.623) | | | |
| Anemia | 277 | 296 | 0.03 (−0.13, 0.19) |
| No anemia | 408 | 395 | 0.08 (−0.06, 0.21) |
| Birth length (cm) (p=0.497) | | | |
| Anemia | 221 | 244 | 0.07 (−0.32, 0.46) |
| No anemia | 339 | 332 | 0.23 (−0.08, 0.54) |
| Length−for−age z−score (p=0.601) | | | |
| Anemia | 221 | 244 | 0.04 (−0.16, 0.24) |
| No anemia | 339 | 332 | 0.10 (−0.06, 0.26) |
| Length−for−gestational age z−score (p=0.322) | | | |
| Anemia | 216 | 242 | 0.01 (−0.18, 0.21) |
| No anemia | 335 | 329 | 0.13 (−0.02, 0.29) |
| BMI z−score (p=0.207) | | | |
| Anemia | 218 | 236 | 0.12 (−0.10, 0.33) |
| No anemia | 338 | 328 | −0.06 (−0.22, 0.11) |
| Head circumference (cm) (p=0.798) | | | |
| Anemia | 221 | 244 | 0.14 (−0.11, 0.40) |
| No anemia | 339 | 333 | 0.16 (−0.06, 0.38) |
| Head circumference−for−age z−score (p=0.922) | | | |
| Anemia | 221 | 244 | 0.12 (−0.08, 0.32) |
| No anemia | 339 | 333 | 0.11 (−0.06, 0.28) |
| Head circumference−for−gestational age z score (p=0.560) | | | |
| Anemia | 216 | 242 | 0.08 (−0.11, 0.27) |
| No anemia | 335 | 330 | 0.14 (−0.02, 0.30) |
| Mid−upper arm circumference (cm) (p=0.747) | | | |
| Anemia | 222 | 244 | 0.07 (−0.09, 0.24) |
| No anemia | 341 | 334 | 0.10 (−0.03, 0.23) |
| Duration (wk) (p=0.660) | | | |
| Anemia | 318 | 334 | −0.11 (−0.50, 0.28) |
| No anemia | 449 | 437 | 0.00 (−0.30, 0.30) |

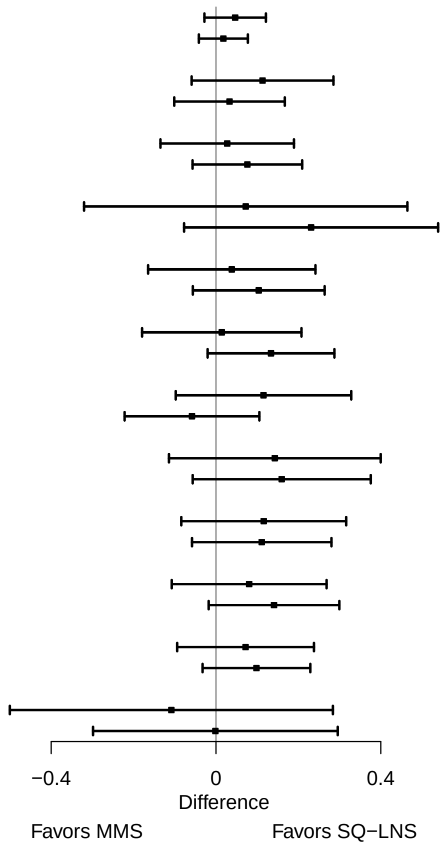

**Supplemental figure 6G: Baseline anemia status**

#### 6G2: Relative risks for birth outcomes

| Outcome (p−for−interaction) | SQ−LNS N | MMS N | Relative risk (95% CI) |
|---|---|---|---|
| Low birth weight (p=0.740) | | | |
| Anemia | 282 | 298 | 0.84 (0.55, 1.28) |
| No anemia | 412 | 398 | 0.92 (0.61, 1.39) |
| Birth weight < 2 kg (p=0.547) | | | |
| Anemia | 282 | 298 | 0.96 (0.38, 2.45) |
| No anemia | 412 | 398 | 1.47 (0.42, 5.18) |
| Small−for−gestational age (p=0.391) | | | |
| Anemia | 282 | 298 | 0.88 (0.65, 1.19) |
| No anemia | 412 | 398 | 1.05 (0.80, 1.36) |
| Large−for−gestational age (p=0.611) | | | |
| Anemia | 282 | 298 | 0.96 (0.38, 2.46) |
| No anemia | 412 | 398 | 1.29 (0.60, 2.78) |
| Newborn stunting (p=0.664) | | | |
| Anemia | 221 | 244 | 1.16 (0.73, 1.84) |
| No anemia | 339 | 332 | 0.98 (0.61, 1.58) |
| Low LGAZ (p=0.168) | | | |
| Anemia | 221 | 244 | 1.61 (0.91, 2.86) |
| No anemia | 339 | 332 | 0.97 (0.60, 1.57) |
| Low BMIZ (p=0.245) | | | |
| Anemia | 218 | 236 | 0.77 (0.38, 1.52) |
| No anemia | 338 | 328 | 1.31 (0.71, 2.42) |
| Low HCZ (p=0.235) | | | |
| Anemia | 221 | 244 | 0.89 (0.41, 1.95) |
| No anemia | 339 | 333 | 1.73 (0.82, 3.64) |
| Preterm birth (p=0.636) | | | |
| Anemia | 318 | 334 | 1.10 (0.73, 1.67) |
| No anemia | 449 | 437 | 1.36 (0.83, 2.23) |

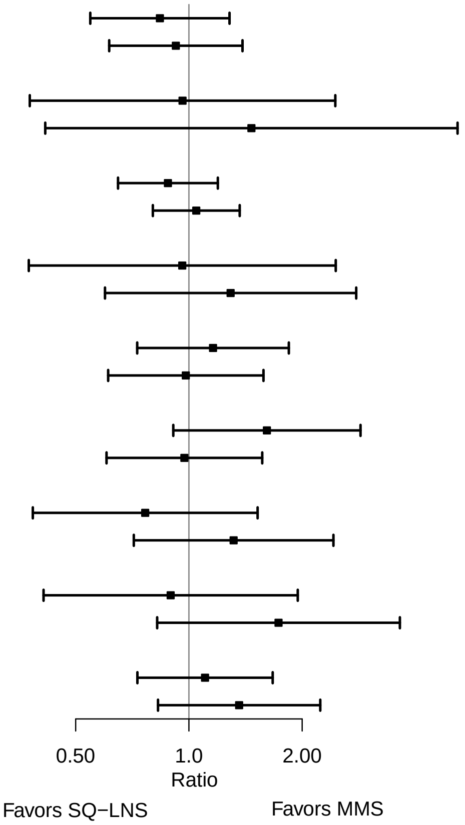

**Supplemental figure 6G: Baseline anemia status**

#### 6G3: Mean differences for 6 mo outcomes

| Outcome (p−for−interaction) | SQ−LNS N | MMS N | Mean difference (95% CI) |
|---|---|---|---|
| Weight−for−age z score (p=0.767) | | | |
| Anemia | 258 | 279 | 0.01 (−0.19, 0.21) |
| No anemia | 373 | 381 | −0.03 (−0.19, 0.13) |
| Length−for−age z−score (p=0.551) | | | |
| Anemia | 259 | 277 | 0.06 (−0.12, 0.25) |
| No anemia | 373 | 379 | −0.01 (−0.17, 0.14) |
| Weight−for−length z−score (p=0.952) | | | |
| Anemia | 258 | 277 | −0.04 (−0.24, 0.15) |
| No anemia | 373 | 379 | −0.04 (−0.20, 0.12) |
| Head circumference−for−age z−score (p=0.789) | | | |
| Anemia | 259 | 279 | −0.05 (−0.22, 0.13) |
| No anemia | 373 | 381 | −0.02 (−0.15, 0.11) |
| MUAC−for−age z−score (p=0.972) | | | |
| Anemia | 259 | 279 | −0.02 (−0.21, 0.17) |
| No anemia | 373 | 381 | −0.02 (−0.17, 0.13) |

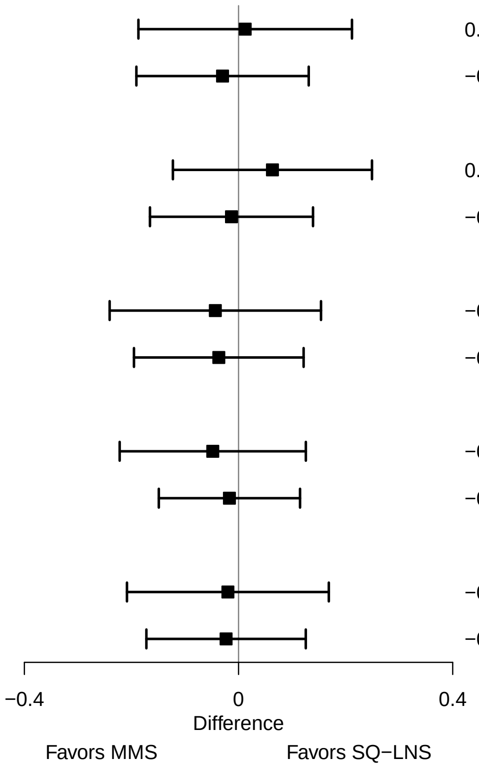

**Supplemental figure 6G: Baseline anemia status**

#### 6G4: Prevalence ratios for 6 mo outcomes

| Outcome (p−for−interaction) | SQ−LNS N | MMS N | Prevalence ratio (95% CI) |
|---|---|---|---|
| Underweight (p=0.216) | | | |
| Anemia | 258 | 279 | 0.89 (0.51, 1.55) |
| No anemia | 373 | 381 | 1.42 (0.84, 2.39) |
| Stunted (p=0.397) | | | |
| Anemia | 259 | 277 | 1.18 (0.83, 1.69) |
| No anemia | 373 | 379 | 0.95 (0.69, 1.33) |
| Low HCAZ (p=0.567) | | | |
| Anemia | 259 | 279 | 0.87 (0.54, 1.41) |
| No anemia | 373 | 381 | 0.68 (0.35, 1.32) |
| Low MUAC (p=0.632) | | | |
| Anemia | 259 | 279 | 1.00 (0.58, 1.72) |
| No anemia | 373 | 381 | 0.84 (0.47, 1.51) |
| Acute malnutrition (p=0.605) | | | |
| Anemia | 258 | 277 | 1.08 (0.64, 1.82) |
| No anemia | 373 | 379 | 0.89 (0.50, 1.58) |

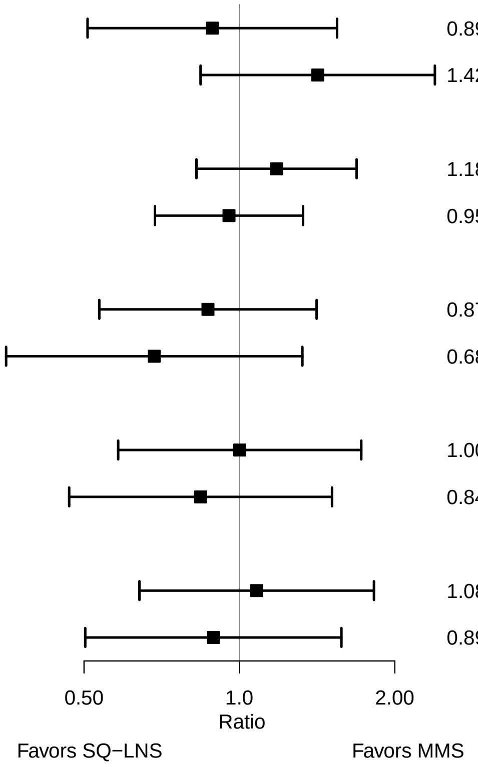

### Supplemental figure 6H: Baseline inflammation status

#### 6H1: Mean differences for birth outcomes

| Outcome (p−for−interaction) | SQ−LNS N | MMS N | Mean difference (95% CI) |
|---|---|---|---|
| Weight (kg) (p=0.080) | | | |
| Not | 384 | 421 | 0.00 (−0.05, 0.06) |
| Inflamed | 296 | 267 | 0.09 (0.01, 0.17) |
| Weight−for−age z score (p=0.106) | | | |
| Not | 384 | 421 | 0.01 (−0.12, 0.15) |
| Inflamed | 296 | 267 | 0.19 (0.02, 0.37) |
| Weight−for−gestational age z−score (p=0.704) | | | |
| Not | 377 | 420 | 0.05 (−0.08, 0.18) |
| Inflamed | 294 | 263 | 0.09 (−0.08, 0.25) |
| Birth length (cm) (p=0.758) | | | |
| Not | 317 | 353 | 0.17 (−0.12, 0.46) |
| Inflamed | 232 | 216 | 0.24 (−0.17, 0.66) |
| Length−for−age z−score (p=0.992) | | | |
| Not | 317 | 353 | 0.10 (−0.05, 0.25) |
| Inflamed | 232 | 216 | 0.10 (−0.11, 0.31) |
| Length−for−gestational age z−score (p=0.920) | | | |
| Not | 310 | 352 | 0.08 (−0.07, 0.23) |
| Inflamed | 230 | 212 | 0.07 (−0.13, 0.27) |
| BMI z−score (p=0.661) | | | |
| Not | 314 | 348 | 0.05 (−0.12, 0.21) |
| Inflamed | 231 | 209 | −0.01 (−0.23, 0.21) |
| Head circumference (cm) (p=0.173) | | | |
| Not | 316 | 354 | 0.26 (0.05, 0.48) |
| Inflamed | 233 | 216 | 0.04 (−0.21, 0.30) |
| Head circumference−for−age z−score (p=0.097) | | | |
| Not | 316 | 354 | 0.22 (0.05, 0.38) |
| Inflamed | 233 | 216 | 0.00 (−0.20, 0.20) |
| Head circumference−for−gestational age z score (p=0.052) | | | |
| Not | 309 | 353 | 0.22 (0.06, 0.38) |
| Inflamed | 231 | 212 | −0.02 (−0.21, 0.17) |
| Mid−upper arm circumference (cm) (p=0.841) | | | |
| Not | 317 | 355 | 0.11 (−0.01, 0.24) |
| Inflamed | 234 | 216 | 0.09 (−0.08, 0.26) |
| Duration (wk) (p=0.460) | | | |
| Not | 426 | 470 | −0.06 (−0.36, 0.24) |
| Inflamed | 325 | 292 | 0.12 (−0.27, 0.50) |

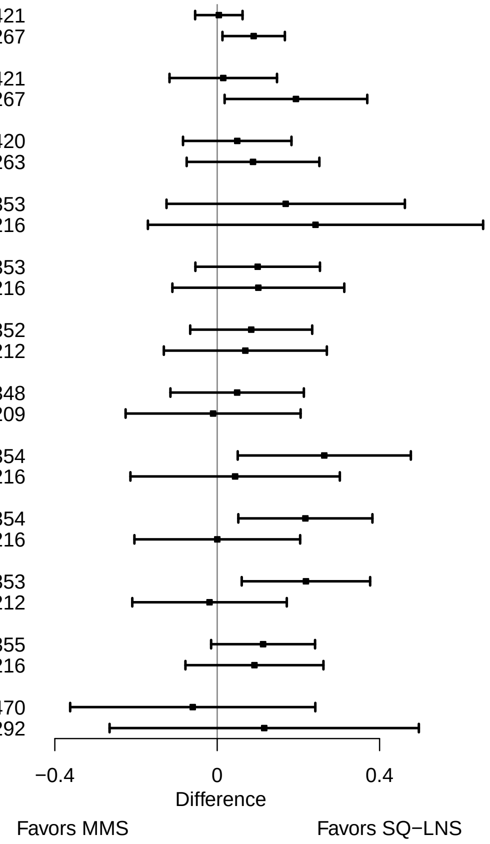

**Supplemental figure 6H: Baseline inflammation status**

#### 6H2: Relative risks for birth outcomes

| Outcome (p−for−interaction) | SQ−LNS N | MMS N | Relative risk (95% CI) |
|---|---|---|---|
| Low birth weight (p=0.220) | | | |
| Not | 384 | 421 | 0.97 (0.64, 1.47) |
| Inflamed | 296 | 267 | 0.65 (0.42, 1.02) |
| Small−for−gestational age (p=0.880) | | | |
| Not | 384 | 421 | 0.93 (0.71, 1.22) |
| Inflamed | 296 | 267 | 0.97 (0.71, 1.31) |
| Large−for−gestational age (p=0.287) | | | |
| Not | 384 | 421 | 0.93 (0.40, 2.17) |
| Inflamed | 296 | 267 | 1.54 (0.65, 3.65) |
| Newborn stunting (p=0.879) | | | |
| Not | 317 | 353 | 1.00 (0.64, 1.55) |
| Inflamed | 232 | 216 | 0.96 (0.57, 1.62) |
| Low LGAZ (p=0.999) | | | |
| Not | 317 | 353 | 1.17 (0.71, 1.94) |
| Inflamed | 232 | 216 | 1.19 (0.71, 1.99) |
| Low BMIZ (p=0.912) | | | |
| Not | 314 | 348 | 0.93 (0.53, 1.63) |
| Inflamed | 231 | 209 | 1.03 (0.50, 2.12) |
| Low HCZ (p=0.165) | | | |
| Not | 316 | 354 | 1.47 (0.70, 3.06) |
| Inflamed | 233 | 216 | 0.66 (0.29, 1.50) |
| Low HCGAZ (p=0.751) | | | |
| Not | 316 | 354 | 0.73 (0.21, 2.57) |
| Inflamed | 233 | 216 | 1.14 (0.31, 4.23) |
| Preterm birth (p=0.213) | | | |
| Not | 426 | 470 | 1.37 (0.88, 2.13) |
| Inflamed | 325 | 292 | 0.85 (0.53, 1.39) |

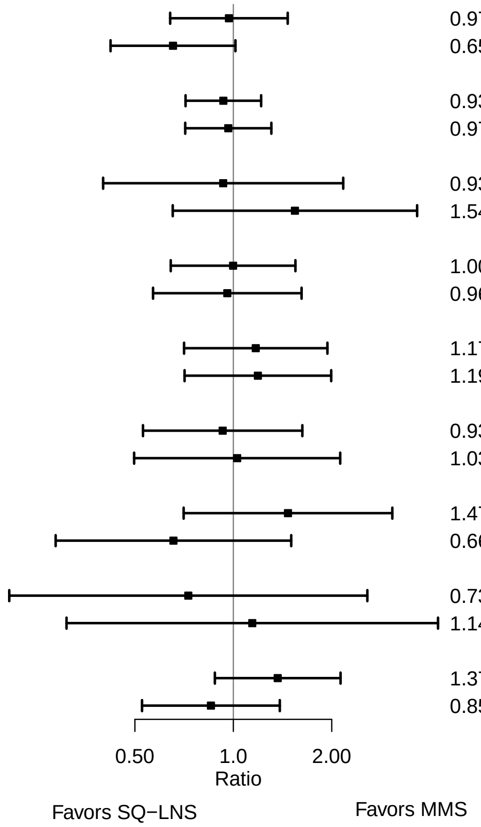

**Supplemental figure 6H: Baseline inflammation status**

#### 6H3: Mean differences for 6 mo outcomes

| **Outcome (p−for−interaction)** | **SQ−LNS N** | **MMS N** | **Mean difference (95% CI)** |
|---|---|---|---|
| Weight−for−age z score (p=0.686) | | | |
| Not | 349 | 401 | −0.01 (−0.17, 0.15) |
| Inflamed | 270 | 252 | 0.04 (−0.16, 0.24) |
| Length−for−age z−score (p=0.421) | | | |
| Not | 350 | 399 | 0.00 (−0.16, 0.15) |
| Inflamed | 270 | 250 | 0.09 (−0.09, 0.28) |
| Weight−for−length z−score (p=0.843) | | | |
| Not | 349 | 399 | −0.02 (−0.18, 0.15) |
| Inflamed | 270 | 250 | −0.05 (−0.24, 0.15) |
| Head circumference−for−age z−score (p=0.782) | | | |
| Not | 350 | 401 | 0.00 (−0.13, 0.14) |
| Inflamed | 270 | 252 | −0.03 (−0.20, 0.13) |
| MUAC−for−age z−score (p=0.838) | | | |
| Not | 350 | 401 | −0.01 (−0.16, 0.14) |
| Inflamed | 270 | 252 | 0.01 (−0.18, 0.20) |

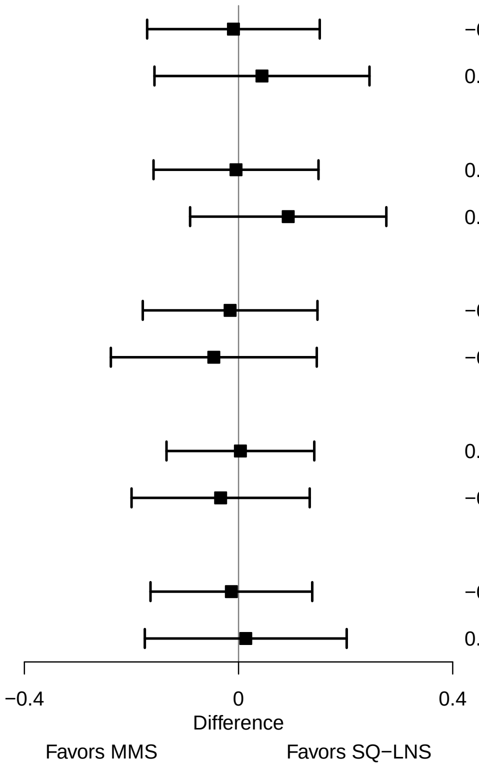

**Supplemental figure 6H: Baseline inflammation status**

#### 6H4: Prevalence ratios for 6 mo outcomes

| Outcome (p−for−interaction) | SQ−LNS N | MMS N | Prevalence ratio (95% CI) |
|---|---|---|---|
| Underweight (p=0.160) | | | |
| Not | 349 | 401 | 1.28 (0.75, 2.19) |
| Inflamed | 270 | 252 | 0.75 (0.42, 1.34) |
| Stunted (p=0.689) | | | |
| Not | 350 | 399 | 0.98 (0.71, 1.36) |
| Inflamed | 270 | 250 | 1.05 (0.72, 1.54) |
| Wasted (p=0.863) | | | |
| Not | 349 | 399 | 1.02 (0.39, 2.65) |
| Inflamed | 270 | 250 | 0.93 (0.30, 2.89) |
| Low HCAZ (p=0.984) | | | |
| Not | 350 | 401 | 0.75 (0.43, 1.31) |
| Inflamed | 270 | 252 | 0.75 (0.42, 1.34) |
| Low MUAC (p=0.092) | | | |
| Not | 350 | 401 | 1.32 (0.76, 2.30) |
| Inflamed | 270 | 252 | 0.62 (0.33, 1.17) |
| Acute malnutrition (p=0.097) | | | |
| Not | 349 | 399 | 1.36 (0.80, 2.28) |
| Inflamed | 270 | 250 | 0.68 (0.37, 1.24) |

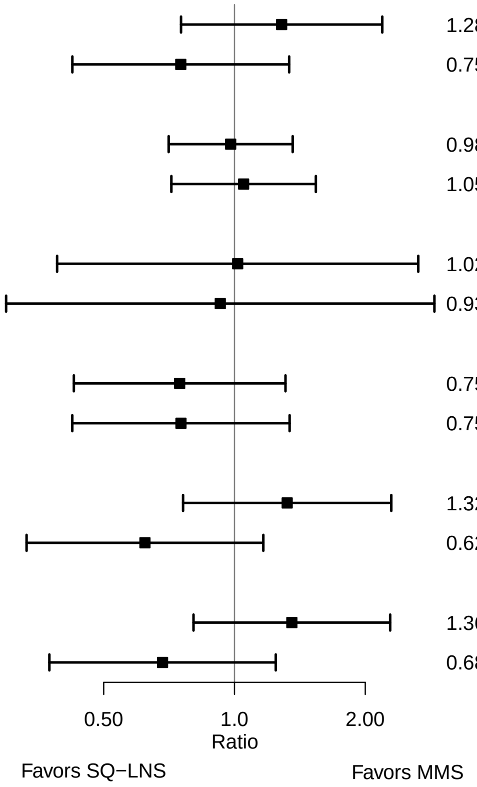

### Supplemental figure 6I: Baseline malaria status

#### 6I1: Mean differences for birth outcomes

| Outcome (p−for−interaction) | SQ−LNS N | MMS N | Mean difference (95% CI) |
|---|---|---|---|
| Weight (kg) (p=0.635) | | | |
| No malaria | 574 | 582 | 0.03 (−0.02, 0.08) |
| Malaria | 118 | 114 | −0.02 (−0.12, 0.09) |
| Weight−for−age z score (p=0.741) | | | |
| No malaria | 574 | 582 | 0.07 (−0.05, 0.18) |
| Malaria | 118 | 114 | −0.04 (−0.28, 0.20) |
| Weight−for−gestational age z−score (p=0.078) | | | |
| No malaria | 567 | 578 | 0.10 (−0.02, 0.21) |
| Malaria | 116 | 113 | −0.16 (−0.39, 0.07) |
| Birth length (cm) (p=0.021) | | | |
| No malaria | 470 | 493 | 0.28 (0.02, 0.54) |
| Malaria | 89 | 83 | −0.51 (−1.17, 0.15) |
| Length−for−age z−score (p=0.024) | | | |
| No malaria | 470 | 493 | 0.14 (0.00, 0.27) |
| Malaria | 89 | 83 | −0.26 (−0.60, 0.08) |
| Length−for−gestational age z−score (p=0.001) | | | |
| No malaria | 463 | 489 | 0.17 (0.04, 0.29) |
| Malaria | 87 | 82 | −0.39 (−0.72, −0.06) |
| BMI z−score (p=0.857) | | | |
| No malaria | 467 | 487 | 0.02 (−0.12, 0.16) |
| Malaria | 88 | 77 | −0.02 (−0.35, 0.30) |
| Head circumference (cm) (p=0.615) | | | |
| No malaria | 471 | 494 | 0.17 (−0.02, 0.35) |
| Malaria | 88 | 83 | −0.02 (−0.41, 0.36) |
| Head circumference−for−age z−score (p=0.752) | | | |
| No malaria | 471 | 494 | 0.12 (−0.02, 0.26) |
| Malaria | 88 | 83 | −0.01 (−0.31, 0.29) |
| Head circumference−for−gestational age z score (p=0.335) | | | |
| No malaria | 464 | 490 | 0.15 (0.01, 0.28) |
| Malaria | 86 | 82 | −0.05 (−0.35, 0.25) |
| Mid−upper arm circumference (cm) (p=0.195) | | | |
| No malaria | 473 | 495 | 0.11 (0.00, 0.22) |
| Malaria | 89 | 83 | −0.08 (−0.34, 0.17) |
| Duration (wk) (p=0.588) | | | |
| No malaria | 629 | 639 | −0.07 (−0.33, 0.19) |
| Malaria | 136 | 132 | 0.02 (−0.58, 0.62) |

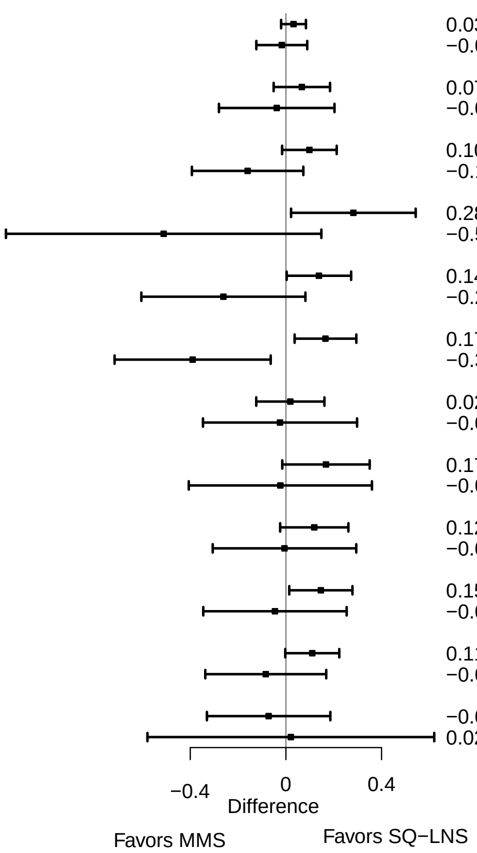

**Supplemental figure 6I: Baseline malaria status**

#### 6I2: Relative risks for birth outcomes

| Outcome (p−for−interaction) | SQ−LNS N | MMS N | Relative risk (95% CI) |
|---|---|---|---|
| Small−for−gestational age (p=0.717) | | | |
| No malaria | 574 | 582 | 0.94 (0.75, 1.17) |
| Malaria | 118 | 114 | 1.05 (0.66, 1.65) |
| Newborn stunting (p=0.005) | | | |
| No malaria | 470 | 493 | 0.83 (0.58, 1.20) |
| Malaria | 89 | 83 | 3.19 (1.35, 7.53) |
| Low LGAZ (p=0.066) | | | |
| No malaria | 470 | 493 | 0.99 (0.66, 1.49) |
| Malaria | 89 | 83 | 2.40 (1.06, 5.44) |

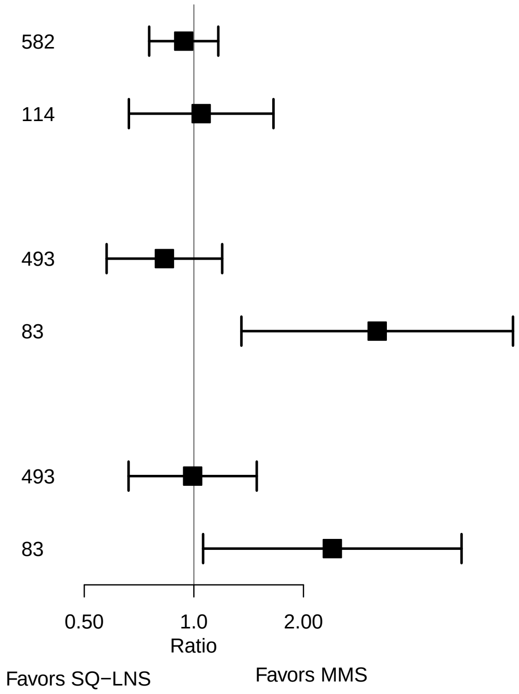

**Supplemental figure 6I: Baseline malaria status**

#### 6I3: Mean differences for 6 mo outcomes

| Outcome (p−for−interaction) | SQ−LNS N | MMS N | Mean difference (95% CI) |
|---|---|---|---|
| Weight−for−age z score (p=0.509) | | | |
| No malaria | 523 | 552 | 0.01 (−0.13, 0.14) |
| Malaria | 106 | 108 | −0.12 (−0.45, 0.20) |
| Length−for−age z−score (p=0.102) | | | |
| No malaria | 524 | 548 | 0.06 (−0.06, 0.19) |
| Malaria | 106 | 108 | −0.20 (−0.48, 0.09) |
| Weight−for−length z−score (p=0.768) | | | |
| No malaria | 523 | 548 | −0.05 (−0.18, 0.09) |
| Malaria | 106 | 108 | 0.00 (−0.33, 0.32) |
| Head circumference−for−age z−score (p=0.529) | | | |
| No malaria | 524 | 552 | −0.01 (−0.12, 0.11) |
| Malaria | 106 | 108 | −0.12 (−0.38, 0.14) |
| MUAC−for−age z−score (p=0.952) | | | |
| No malaria | 524 | 552 | −0.02 (−0.15, 0.11) |
| Malaria | 106 | 108 | −0.02 (−0.31, 0.27) |

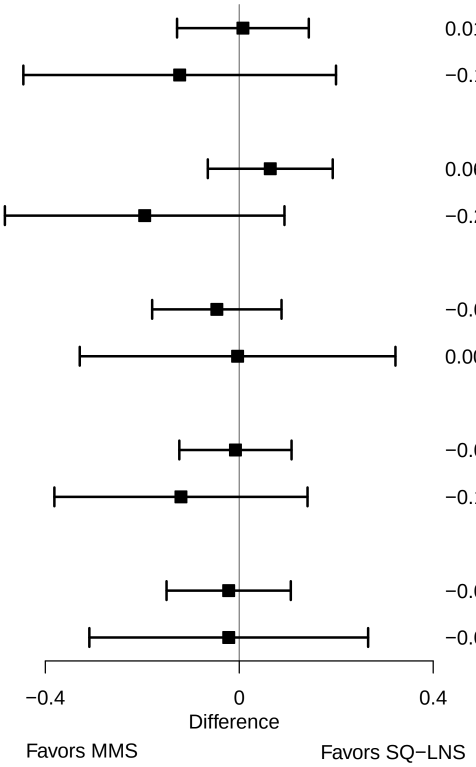

**Supplemental figure 6I: Baseline malaria status**

#### 6I4: Prevalence ratios for 6 mo outcomes

| Outcome (p−for−interaction) | SQ−LNS N | MMS N | Prevalence ratio (95% CI) |
|---|---|---|---|
| Stunted (p=0.116) | | | |
| No malaria | 524 | 548 | 0.95 (0.73, 1.24) |
| Malaria | 106 | 108 | 1.54 (0.85, 2.80) |
| Low HCAZ (p=0.801) | | | |
| No malaria | 524 | 552 | 0.79 (0.51, 1.23) |
| Malaria | 106 | 108 | 0.69 (0.27, 1.81) |

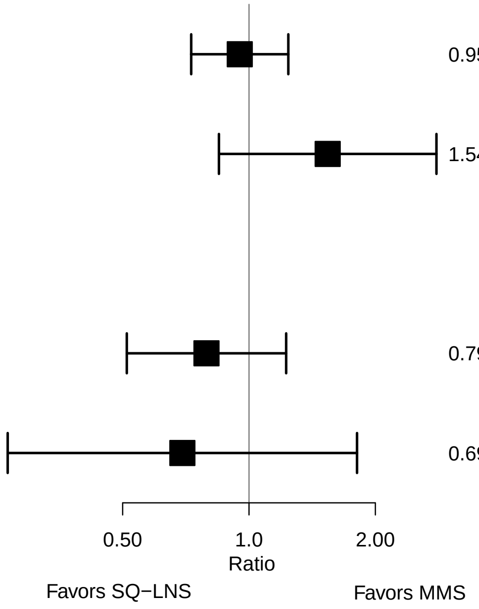

### Supplemental figure 6J: Gestational age at supplementation

#### 6J1: Mean differences for birth outcomes

| Outcome (p−for−interaction) | SQ−LNS N | MMS N | Mean difference (95% CI) |
|---|---|---|---|
| Weight (kg) (p=0.812) | | | |
| < 14 wks | 120 | 111 | 0.04 (−0.07, 0.14) |
| >= 14 wks | 574 | 586 | 0.03 (−0.03, 0.08) |
| Weight−for−age z score (p=0.755) | | | |
| < 14 wks | 120 | 111 | 0.10 (−0.14, 0.33) |
| >= 14 wks | 574 | 586 | 0.05 (−0.07, 0.17) |
| Weight−for−gestational age z−score (p=0.472) | | | |
| < 14 wks | 115 | 110 | 0.15 (−0.09, 0.38) |
| >= 14 wks | 570 | 582 | 0.04 (−0.08, 0.15) |
| Birth length (cm) (p=0.674) | | | |
| < 14 wks | 105 | 95 | 0.11 (−0.43, 0.65) |
| >= 14 wks | 455 | 482 | 0.18 (−0.09, 0.45) |
| Length−for−age z−score (p=0.785) | | | |
| < 14 wks | 105 | 95 | 0.07 (−0.21, 0.35) |
| >= 14 wks | 455 | 482 | 0.08 (−0.06, 0.22) |
| Length−for−gestational age z−score (p=0.898) | | | |
| < 14 wks | 100 | 94 | 0.08 (−0.20, 0.36) |
| >= 14 wks | 451 | 478 | 0.08 (−0.05, 0.22) |
| BMI z−score (p=0.436) | | | |
| < 14 wks | 105 | 95 | 0.13 (−0.17, 0.42) |
| >= 14 wks | 451 | 470 | −0.01 (−0.15, 0.14) |
| Head circumference (cm) (p=0.903) | | | |
| < 14 wks | 104 | 95 | 0.22 (−0.16, 0.59) |
| >= 14 wks | 456 | 483 | 0.14 (−0.04, 0.33) |
| Head circumference−for−age z−score (p=0.897) | | | |
| < 14 wks | 104 | 95 | 0.19 (−0.11, 0.48) |
| >= 14 wks | 456 | 483 | 0.10 (−0.04, 0.24) |
| Head circumference−for−gestational age z score (p=0.921) | | | |
| < 14 wks | 99 | 94 | 0.17 (−0.11, 0.46) |
| >= 14 wks | 452 | 479 | 0.12 (−0.02, 0.25) |
| Mid−upper arm circumference (cm) (p=0.419) | | | |
| < 14 wks | 105 | 95 | 0.18 (−0.06, 0.42) |
| >= 14 wks | 458 | 484 | 0.06 (−0.05, 0.17) |
| Duration (wk) (p=0.467) | | | |
| < 14 wks | 135 | 125 | 0.19 (−0.37, 0.75) |
| >= 14 wks | 632 | 647 | −0.08 (−0.34, 0.18) |

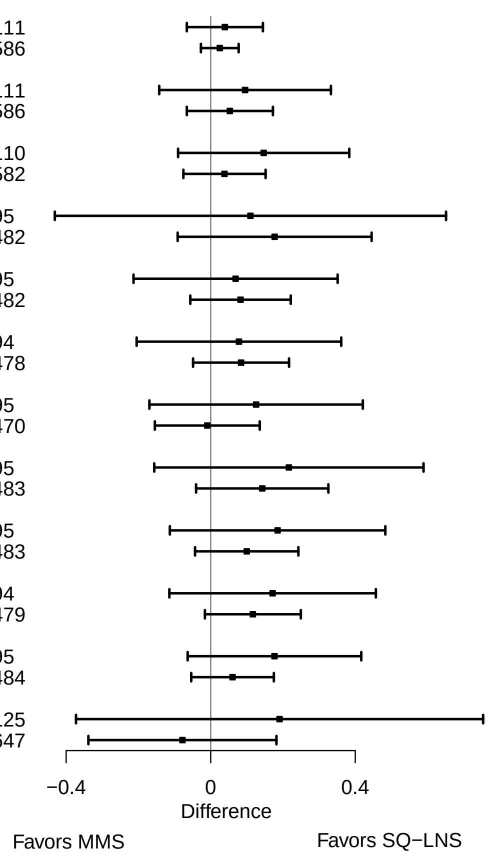

**Supplemental figure 6J: Gestational age at supplementation**

#### 6J2: Relative risks for birth outcomes

| Outcome (p−for−interaction) | SQ−LNS N | MMS N | Relative risk (95% CI) |
|---|---|---|---|
| Low birth weight (p=0.886) | | | |
| < 14 wks | 120 | 111 | 0.92 (0.43, 1.97) |
| >= 14 wks | 574 | 586 | 0.87 (0.64, 1.20) |
| Small−for−gestational age (p=0.830) | | | |
| < 14 wks | 120 | 111 | 1.01 (0.62, 1.65) |
| >= 14 wks | 574 | 586 | 0.96 (0.77, 1.19) |
| Newborn stunting (p=0.386) | | | |
| < 14 wks | 105 | 95 | 1.53 (0.62, 3.75) |
| >= 14 wks | 455 | 482 | 1.00 (0.71, 1.42) |
| Low LGAZ (p=0.341) | | | |
| < 14 wks | 105 | 95 | 1.77 (0.77, 4.08) |
| >= 14 wks | 455 | 482 | 1.11 (0.75, 1.65) |
| Low BMIZ (p=0.672) | | | |
| < 14 wks | 105 | 95 | 0.86 (0.30, 2.46) |
| >= 14 wks | 451 | 470 | 1.01 (0.61, 1.64) |
| Low HCZ (p=0.881) | | | |
| < 14 wks | 104 | 95 | 1.43 (0.34, 6.10) |
| >= 14 wks | 456 | 483 | 1.24 (0.72, 2.13) |
| Low HCGAZ (p=0.990) | | | |
| < 14 wks | 104 | 95 | 0.91 (0.17, 4.86) |
| >= 14 wks | 456 | 483 | 0.80 (0.28, 2.29) |
| Preterm birth (p=0.399) | | | |
| < 14 wks | 135 | 125 | 1.01 (0.47, 2.14) |
| >= 14 wks | 632 | 647 | 1.25 (0.87, 1.78) |

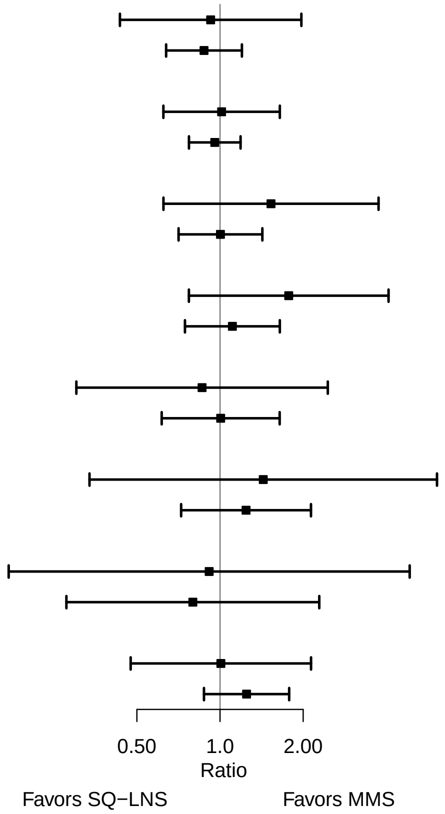

**Supplemental figure 6J: Gestational age at supplementation**

#### 6J3: Mean differences for 6 mo outcomes

| Outcome (p−for−interaction) | SQ−LNS N | MMS N | Mean difference (95% CI) |
|---|---|---|---|
| Weight−for−age z score (p=0.256) | | | |
| < 14 wks | 114 | 111 | 0.15 (−0.16, 0.46) |
| >= 14 wks | 517 | 550 | −0.05 (−0.18, 0.09) |
| Length−for−age z−score (p=0.581) | | | |
| < 14 wks | 114 | 111 | 0.09 (−0.20, 0.37) |
| >= 14 wks | 518 | 546 | 0.00 (−0.13, 0.13) |
| Weight−for−length z−score (p=0.262) | | | |
| < 14 wks | 114 | 111 | 0.11 (−0.19, 0.42) |
| >= 14 wks | 517 | 546 | −0.07 (−0.20, 0.07) |
| Head circumference−for−age z−score (p=0.123) | | | |
| < 14 wks | 114 | 111 | 0.17 (−0.09, 0.43) |
| >= 14 wks | 518 | 550 | −0.07 (−0.18, 0.05) |
| MUAC−for−age z−score (p=0.416) | | | |
| < 14 wks | 114 | 111 | 0.07 (−0.20, 0.35) |
| >= 14 wks | 518 | 550 | −0.04 (−0.17, 0.09) |

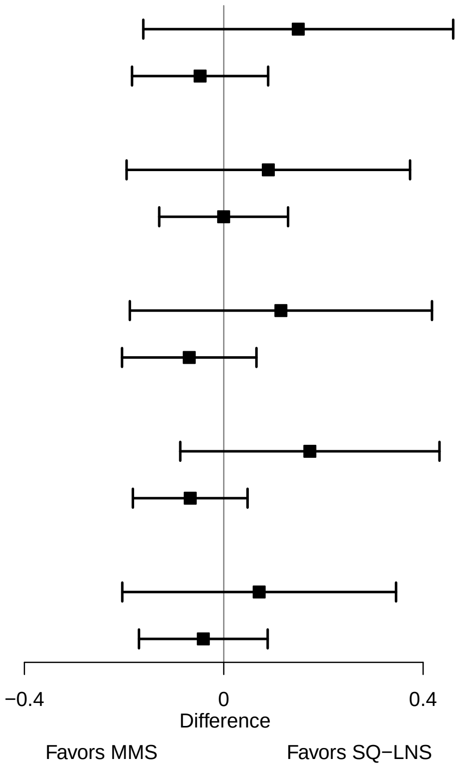

**Supplemental figure 6J: Gestational age at supplementation**

#### 6J4: Prevalence ratios for 6 mo outcomes

| Outcome (p−for−interaction) | SQ−LNS N | MMS N | Prevalence ratio (95% CI) |
|---|---|---|---|
| Underweight (p=0.561) | | | |
| < 14 wks | 114 | 111 | 0.88 (0.35, 2.22) |
| >= 14 wks | 517 | 550 | 1.19 (0.80, 1.79) |
| Stunted (p=0.221) | | | |
| < 14 wks | 114 | 111 | 0.76 (0.40, 1.44) |
| >= 14 wks | 518 | 546 | 1.11 (0.86, 1.44) |
| Wasted (p=0.396) | | | |
| < 14 wks | 114 | 111 | 0.50 (0.09, 2.84) |
| >= 14 wks | 517 | 546 | 1.14 (0.52, 2.49) |
| Low HCAZ (p=0.806) | | | |
| < 14 wks | 114 | 111 | 0.72 (0.24, 2.10) |
| >= 14 wks | 518 | 550 | 0.82 (0.53, 1.27) |
| Low MUAC (p=0.867) | | | |
| < 14 wks | 114 | 111 | 0.87 (0.35, 2.17) |
| >= 14 wks | 518 | 550 | 0.93 (0.60, 1.44) |
| Acute malnutrition (p=0.972) | | | |
| < 14 wks | 114 | 111 | 0.97 (0.40, 2.36) |
| >= 14 wks | 517 | 546 | 0.98 (0.65, 1.49) |

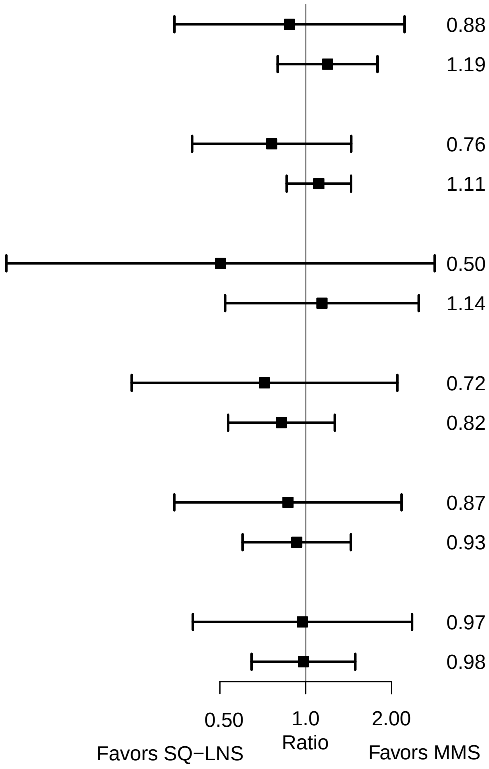

### Supplemental figure 6K: Compliance with supplementation

#### 6K1: Mean differences for birth outcomes

| Outcome (p−for−interaction) | SQ−LNS N | MMS N | Mean difference (95% CI) |
|---|---|---|---|
| Weight (kg) (p=0.677) | | | |
| >= 4/7 | 607 | 621 | 0.03 (−0.02, 0.08) |
| Less than 4/7 | 84 | 75 | 0.06 (−0.09, 0.21) |
| Weight−for−age z score (p=0.417) | | | |
| >= 4/7 | 607 | 621 | 0.05 (−0.06, 0.16) |
| Less than 4/7 | 84 | 75 | 0.19 (−0.16, 0.54) |
| Weight−for−gestational age z−score (p=0.297) | | | |
| >= 4/7 | 602 | 617 | 0.04 (−0.07, 0.15) |
| Less than 4/7 | 80 | 74 | 0.23 (−0.08, 0.55) |
| Birth length (cm) (p=0.800) | | | |
| >= 4/7 | 487 | 513 | 0.15 (−0.10, 0.40) |
| Less than 4/7 | 73 | 63 | 0.29 (−0.60, 1.18) |
| Length−for−age z−score (p=0.747) | | | |
| >= 4/7 | 487 | 513 | 0.07 (−0.06, 0.20) |
| Less than 4/7 | 73 | 63 | 0.14 (−0.30, 0.59) |
| Length−for−gestational age z−score (p=0.569) | | | |
| >= 4/7 | 482 | 509 | 0.09 (−0.04, 0.21) |
| Less than 4/7 | 69 | 62 | −0.01 (−0.39, 0.37) |
| BMI z−score (p=0.298) | | | |
| >= 4/7 | 483 | 501 | −0.01 (−0.14, 0.12) |
| Less than 4/7 | 73 | 63 | 0.21 (−0.25, 0.66) |
| Head circumference (cm) (p=0.285) | | | |
| >= 4/7 | 487 | 514 | 0.11 (−0.06, 0.29) |
| Less than 4/7 | 73 | 63 | 0.46 (−0.14, 1.05) |
| Head circumference−for−age z−score (p=0.219) | | | |
| >= 4/7 | 487 | 514 | 0.08 (−0.06, 0.21) |
| Less than 4/7 | 73 | 63 | 0.36 (−0.08, 0.80) |
| Head circumference−for−gestational age z score (p=0.363) | | | |
| >= 4/7 | 482 | 510 | 0.09 (−0.04, 0.22) |
| Less than 4/7 | 69 | 62 | 0.30 (−0.10, 0.70) |
| Mid−upper arm circumference (cm) (p=0.888) | | | |
| >= 4/7 | 490 | 515 | 0.08 (−0.03, 0.19) |
| Less than 4/7 | 73 | 63 | 0.12 (−0.16, 0.40) |
| Duration (wk) (p=0.358) | | | |
| >= 4/7 | 658 | 674 | 0.00 (−0.25, 0.26) |
| Less than 4/7 | 105 | 95 | −0.32 (−0.95, 0.31) |

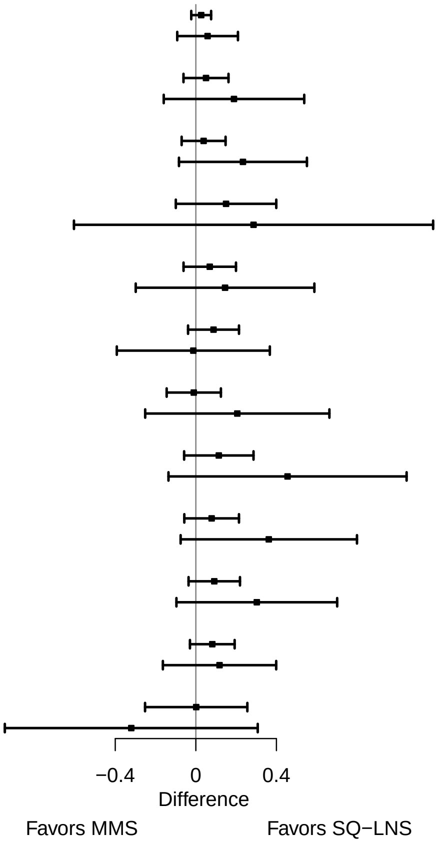

**Supplemental figure 6K: Compliance with supplementation**

#### 6K2: Relative risks for birth outcomes

| Outcome (p−for−interaction) | SQ−LNS N | MMS N | Relative risk (95% CI) |
|---|---|---|---|
| Low birth weight (p=0.460) | | | |
| >= 4/7 | 607 | 621 | 0.92 (0.67, 1.27) |
| Less than 4/7 | 84 | 75 | 0.68 (0.33, 1.41) |
| Small−for−gestational age (p=0.213) | | | |
| >= 4/7 | 607 | 621 | 1.01 (0.81, 1.24) |
| Less than 4/7 | 84 | 75 | 0.67 (0.38, 1.16) |
| Newborn stunting (p=0.589) | | | |
| >= 4/7 | 487 | 513 | 1.10 (0.77, 1.58) |
| Less than 4/7 | 73 | 63 | 0.82 (0.38, 1.74) |
| Low LGAZ (p=0.767) | | | |
| >= 4/7 | 487 | 513 | 1.25 (0.85, 1.84) |
| Less than 4/7 | 73 | 63 | 0.98 (0.38, 2.51) |
| Low BMIZ (p=0.148) | | | |
| >= 4/7 | 483 | 501 | 1.17 (0.72, 1.92) |
| Less than 4/7 | 73 | 63 | 0.44 (0.16, 1.23) |
| Low HCZ (p=0.113) | | | |
| >= 4/7 | 487 | 514 | 1.55 (0.89, 2.70) |
| Less than 4/7 | 73 | 63 | 0.48 (0.13, 1.83) |
| Preterm birth (p=0.861) | | | |
| >= 4/7 | 658 | 674 | 1.17 (0.82, 1.68) |
| Less than 4/7 | 105 | 95 | 1.34 (0.65, 2.74) |

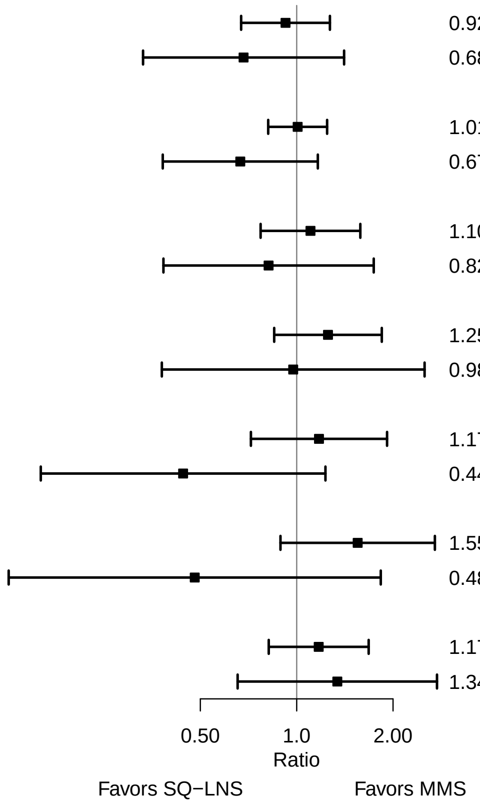

**Supplemental figure 6K: Compliance with supplementation**

#### 6K3: Mean differences for 6 mo outcomes

| Outcome (p−for−interaction) | SQ−LNS N | MMS N | Mean difference (95% CI) |
|---|---|---|---|
| Weight−for−age z score (p=0.454) | | | |
| >= 4/7 | 556 | 596 | −0.04 (−0.17, 0.09) |
| Less than 4/7 | 73 | 64 | 0.13 (−0.27, 0.53) |
| Length−for−age z−score (p=0.737) | | | |
| >= 4/7 | 557 | 592 | 0.02 (−0.10, 0.14) |
| Less than 4/7 | 73 | 64 | −0.06 (−0.43, 0.31) |
| Weight−for−length z−score (p=0.230) | | | |
| >= 4/7 | 556 | 592 | −0.07 (−0.20, 0.06) |
| Less than 4/7 | 73 | 64 | 0.16 (−0.21, 0.54) |
| Head circumference−for−age z−score (p=0.601) | | | |
| >= 4/7 | 557 | 596 | −0.04 (−0.15, 0.08) |
| Less than 4/7 | 73 | 64 | 0.05 (−0.25, 0.36) |
| MUAC−for−age z−score (p=0.985) | | | |
| >= 4/7 | 557 | 596 | −0.03 (−0.15, 0.09) |
| Less than 4/7 | 73 | 64 | 0.01 (−0.35, 0.37) |

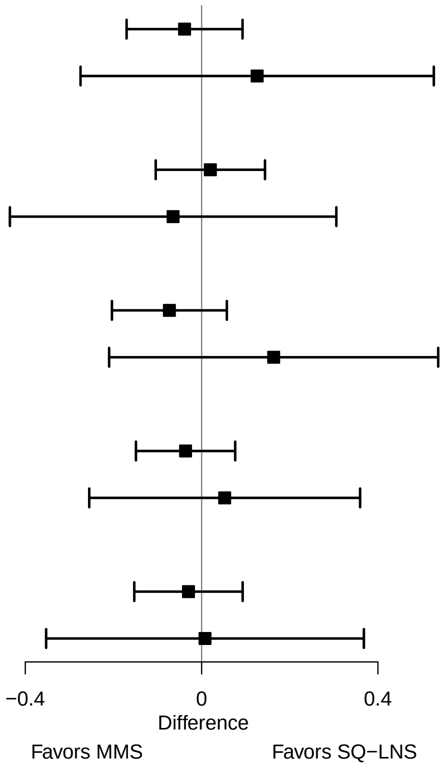

**Supplemental figure 6K: Compliance with supplementation**

#### 6K4: Prevalence ratios for 6 mo outcomes

| Outcome (p−for−interaction) | SQ−LNS N | MMS N | Prevalence ratio (95% CI) |
|---|---|---|---|
| Underweight (p=0.831) | | | |
| >= 4/7 | 556 | 596 | 1.14 (0.78, 1.66) |
| Less than 4/7 | 73 | 64 | 1.36 (0.23, 8.09) |
| Stunted (p=0.593) | | | |
| >= 4/7 | 557 | 592 | 1.07 (0.83, 1.37) |
| Less than 4/7 | 73 | 64 | 0.84 (0.28, 2.49) |
| Low HCAZ (p=0.454) | | | |
| >= 4/7 | 557 | 596 | 0.71 (0.46, 1.09) |
| Less than 4/7 | 73 | 64 | 1.29 (0.28, 5.87) |
| Low MUAC (p=0.600) | | | |
| >= 4/7 | 557 | 596 | 0.92 (0.61, 1.38) |
| Less than 4/7 | 73 | 64 | 1.34 (0.31, 5.81) |
| Acute malnutrition (p=0.675) | | | |
| >= 4/7 | 556 | 592 | 0.98 (0.66, 1.45) |
| Less than 4/7 | 73 | 64 | 1.34 (0.31, 5.81) |

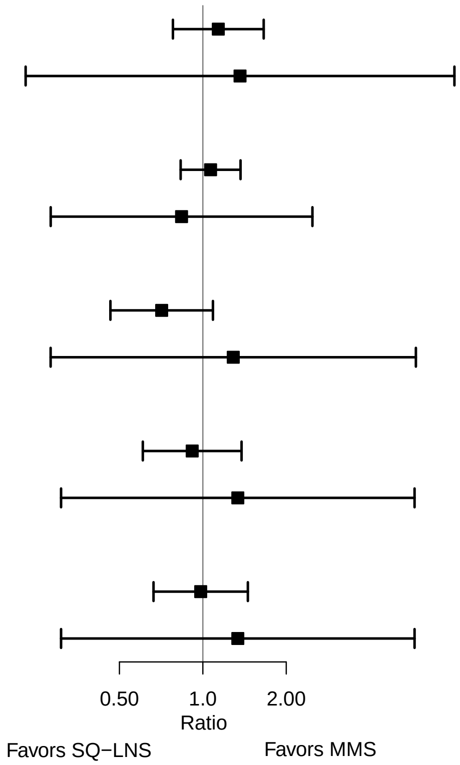

### Supplemental figure 6L: Household socio-economic status

#### 6L1: Mean differences for birth outcomes

| Outcome (p−for−interaction) | SQ−LNS N | MMS N | Mean difference (95% CI) |
|---|---|---|---|
| Weight (kg) (p=0.982) | | | |
| At least median | 305 | 343 | 0.03 (−0.04, 0.10) |
| Less than median | 386 | 350 | 0.03 (−0.03, 0.10) |
| Weight−for−age z score (p=0.875) | | | |
| At least median | 305 | 343 | 0.06 (−0.09, 0.22) |
| Less than median | 386 | 350 | 0.07 (−0.07, 0.22) |
| Weight−for−gestational age z−score (p=0.864) | | | |
| At least median | 303 | 339 | 0.05 (−0.10, 0.20) |
| Less than median | 379 | 349 | 0.07 (−0.08, 0.21) |
| Birth length (cm) (p=0.748) | | | |
| At least median | 248 | 288 | 0.21 (−0.11, 0.53) |
| Less than median | 310 | 287 | 0.12 (−0.23, 0.48) |
| Length−for−age z−score (p=0.806) | | | |
| At least median | 248 | 288 | 0.10 (−0.07, 0.26) |
| Less than median | 310 | 287 | 0.06 (−0.12, 0.25) |
| Length−for−gestational age z−score (p=0.556) | | | |
| At least median | 246 | 284 | 0.12 (−0.05, 0.28) |
| Less than median | 303 | 286 | 0.05 (−0.13, 0.22) |
| BMI z−score (p=0.927) | | | |
| At least median | 246 | 284 | 0.03 (−0.15, 0.21) |
| Less than median | 308 | 280 | 0.04 (−0.14, 0.22) |
| Head circumference (cm) (p=0.728) | | | |
| At least median | 246 | 288 | 0.20 (−0.02, 0.42) |
| Less than median | 312 | 288 | 0.10 (−0.14, 0.34) |
| Head circumference−for−age z−score (p=0.688) | | | |
| At least median | 246 | 288 | 0.15 (−0.03, 0.33) |
| Less than median | 312 | 288 | 0.07 (−0.11, 0.26) |
| Head circumference−for−gestational age z score (p=0.582) | | | |
| At least median | 244 | 284 | 0.16 (−0.01, 0.33) |
| Less than median | 305 | 287 | 0.07 (−0.10, 0.24) |
| Mid−upper arm circumference (cm) (p=0.934) | | | |
| At least median | 248 | 288 | 0.11 (−0.04, 0.26) |
| Less than median | 313 | 289 | 0.09 (−0.05, 0.23) |
| Duration (wk) (p=0.251) | | | |
| At least median | 335 | 379 | 0.11 (−0.19, 0.41) |
| Less than median | 427 | 383 | −0.17 (−0.52, 0.19) |

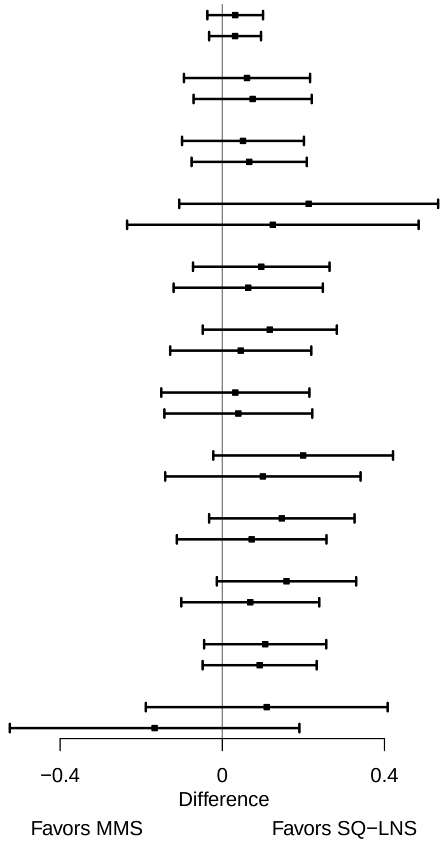

**Supplemental figure 6L: Household socio-economic status**

#### 6L2: Relative risks for birth outcomes

| Outcome (p−for−interaction) | SQ−LNS N | MMS N | Relative risk (95% CI) |
|---|---|---|---|
| Low birth weight (p=0.822) | | | |
| At least median | 305 | 343 | 0.85 (0.54, 1.34) |
| Less than median | 386 | 350 | 0.91 (0.61, 1.34) |
| Birth weight < 2 kg (p=0.333) | | | |
| At least median | 305 | 343 | 1.71 (0.56, 5.18) |
| Less than median | 386 | 350 | 0.72 (0.27, 1.93) |
| Small−for−gestational age (p=0.884) | | | |
| At least median | 305 | 343 | 0.97 (0.71, 1.32) |
| Less than median | 386 | 350 | 0.94 (0.73, 1.22) |
| Large−for−gestational age (p=0.804) | | | |
| At least median | 305 | 343 | 1.24 (0.52, 2.96) |
| Less than median | 386 | 350 | 1.08 (0.47, 2.48) |
| Newborn stunting (p=0.250) | | | |
| At least median | 248 | 288 | 0.82 (0.48, 1.40) |
| Less than median | 310 | 287 | 1.19 (0.79, 1.80) |
| Low LGAZ (p=0.928) | | | |
| At least median | 248 | 288 | 1.18 (0.65, 2.15) |
| Less than median | 310 | 287 | 1.20 (0.77, 1.86) |
| Low BMIZ (p=0.127) | | | |
| At least median | 246 | 284 | 1.39 (0.69, 2.82) |
| Less than median | 308 | 280 | 0.68 (0.38, 1.23) |
| Low HCZ (p=0.675) | | | |
| At least median | 246 | 288 | 1.05 (0.43, 2.55) |
| Less than median | 312 | 288 | 1.30 (0.70, 2.41) |
| Preterm birth (p=0.257) | | | |
| At least median | 335 | 379 | 0.97 (0.59, 1.59) |
| Less than median | 427 | 383 | 1.47 (0.94, 2.30) |

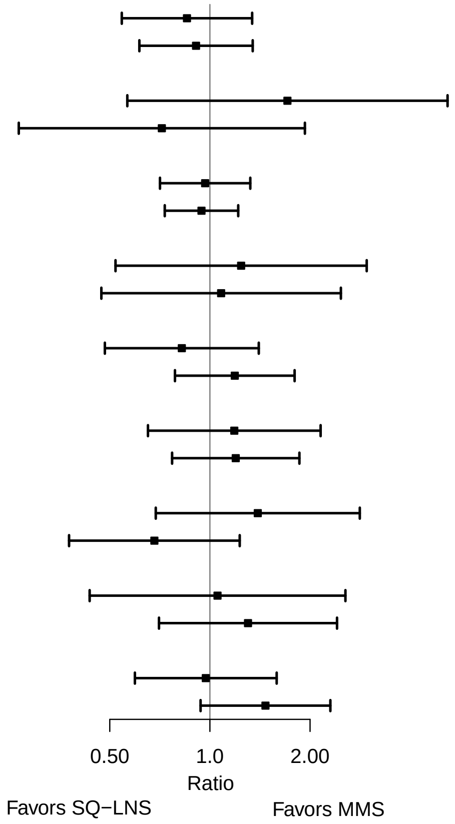

**Supplemental figure 6L: Household socio-economic status**

#### 6L3: Mean differences for 6 mo outcomes

| Outcome (p−for−interaction) | SQ−LNS N | MMS N | Mean difference (95% CI) |
|---|---|---|---|
| Weight−for−age z score (p=0.717) | | | |
| At least median | 267 | 325 | −0.03 (−0.22, 0.15) |
| Less than median | 361 | 332 | 0.01 (−0.16, 0.19) |
| Length−for−age z−score (p=0.463) | | | |
| At least median | 268 | 322 | −0.03 (−0.20, 0.14) |
| Less than median | 361 | 331 | 0.06 (−0.11, 0.22) |
| Weight−for−length z−score (p=0.949) | | | |
| At least median | 267 | 322 | −0.04 (−0.22, 0.14) |
| Less than median | 361 | 331 | −0.03 (−0.20, 0.14) |
| Head circumference−for−age z−score (p=0.914) | | | |
| At least median | 268 | 325 | −0.02 (−0.17, 0.14) |
| Less than median | 361 | 332 | −0.04 (−0.18, 0.11) |
| MUAC−for−age z−score (p=0.857) | | | |
| At least median | 268 | 325 | −0.03 (−0.19, 0.14) |
| Less than median | 361 | 332 | −0.01 (−0.17, 0.16) |

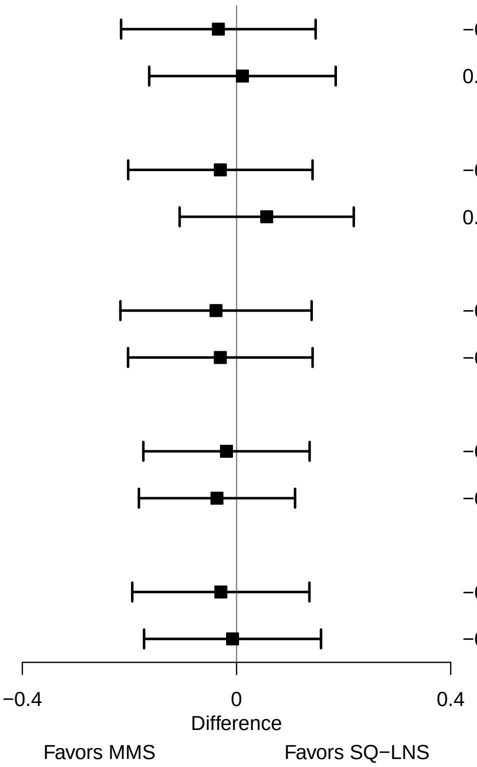

**Supplemental figure 6L: Household socio-economic status**

#### 6L4: Prevalence ratios for 6 mo outcomes

| Outcome (p−for−interaction) | SQ−LNS N | MMS N | Prevalence ratio (95% CI) |
|---|---|---|---|
| Underweight (p=0.986) | | | |
| At least median | 267 | 325 | 1.11 (0.62, 1.99) |
| Less than median | 361 | 332 | 1.12 (0.70, 1.81) |
| Stunted (p=0.924) | | | |
| At least median | 268 | 322 | 1.09 (0.75, 1.59) |
| Less than median | 361 | 331 | 1.03 (0.75, 1.42) |
| Wasted (p=0.792) | | | |
| At least median | 267 | 322 | 1.04 (0.35, 3.07) |
| Less than median | 361 | 331 | 0.89 (0.35, 2.29) |
| Low HCAZ (p=0.308) | | | |
| At least median | 268 | 325 | 0.61 (0.33, 1.13) |
| Less than median | 361 | 332 | 0.91 (0.54, 1.54) |
| Low MUAC (p=0.940) | | | |
| At least median | 268 | 325 | 0.93 (0.49, 1.75) |
| Less than median | 361 | 332 | 0.88 (0.53, 1.47) |
| Acute malnutrition (p=0.744) | | | |
| At least median | 267 | 322 | 0.90 (0.49, 1.65) |
| Less than median | 361 | 331 | 1.01 (0.62, 1.65) |

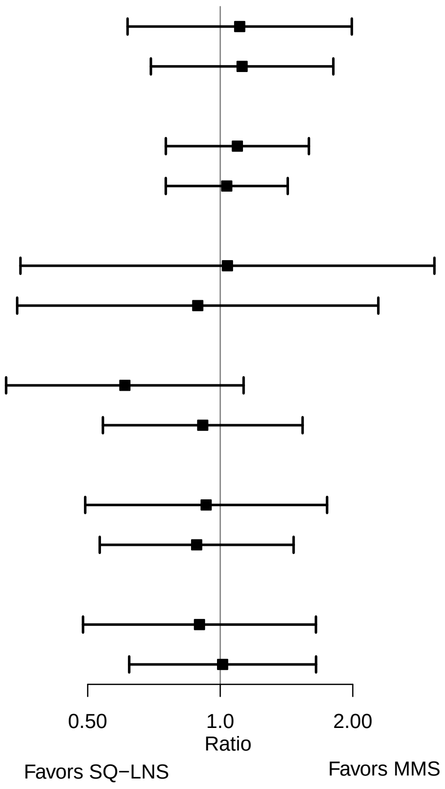

### Supplemental figure 6M: Household food security

#### 6M1: Mean differences for birth outcomes

| Outcome (p−for−interaction) | SQ−LNS N | MMS N | Mean difference (95% CI) |
|---|---|---|---|
| Weight (kg) (p=0.715) | | | |
| Mild to secure | 354 | 334 | 0.02 (−0.05, 0.08) |
| Moderate to severe | 336 | 354 | 0.04 (−0.03, 0.10) |
| Weight−for−age z score (p=0.731) | | | |
| Mild to secure | 354 | 334 | 0.04 (−0.11, 0.19) |
| Moderate to severe | 336 | 354 | 0.08 (−0.07, 0.23) |
| Weight−for−gestational age z−score (p=0.727) | | | |
| Mild to secure | 347 | 330 | 0.04 (−0.10, 0.19) |
| Moderate to severe | 334 | 353 | 0.06 (−0.09, 0.21) |
| Birth length (cm) (p=0.559) | | | |
| Mild to secure | 307 | 298 | 0.11 (−0.21, 0.44) |
| Moderate to severe | 249 | 272 | 0.23 (−0.13, 0.60) |
| Length−for−age z−score (p=0.608) | | | |
| Mild to secure | 307 | 298 | 0.05 (−0.12, 0.22) |
| Moderate to severe | 249 | 272 | 0.12 (−0.07, 0.31) |
| Length−for−gestational age z−score (p=0.597) | | | |
| Mild to secure | 300 | 294 | 0.05 (−0.10, 0.21) |
| Moderate to severe | 247 | 271 | 0.12 (−0.07, 0.31) |
| BMI z−score (p=0.810) | | | |
| Mild to secure | 307 | 298 | 0.00 (−0.18, 0.17) |
| Moderate to severe | 245 | 261 | 0.03 (−0.17, 0.22) |
| Head circumference (cm) (p=0.373) | | | |
| Mild to secure | 308 | 299 | 0.25 (0.00, 0.49) |
| Moderate to severe | 248 | 272 | 0.03 (−0.19, 0.26) |
| Head circumference−for−age z−score (p=0.400) | | | |
| Mild to secure | 308 | 299 | 0.18 (−0.01, 0.37) |
| Moderate to severe | 248 | 272 | 0.03 (−0.14, 0.21) |
| Head circumference−for−gestational age z score (p=0.293) | | | |
| Mild to secure | 301 | 295 | 0.19 (0.02, 0.37) |
| Moderate to severe | 246 | 271 | 0.03 (−0.14, 0.20) |
| Mid−upper arm circumference (cm) (p=0.274) | | | |
| Mild to secure | 309 | 299 | 0.03 (−0.11, 0.17) |
| Moderate to severe | 250 | 273 | 0.13 (−0.02, 0.28) |
| Duration (wk) (p=0.383) | | | |
| Mild to secure | 390 | 365 | 0.03 (−0.28, 0.33) |
| Moderate to severe | 371 | 393 | −0.16 (−0.53, 0.21) |

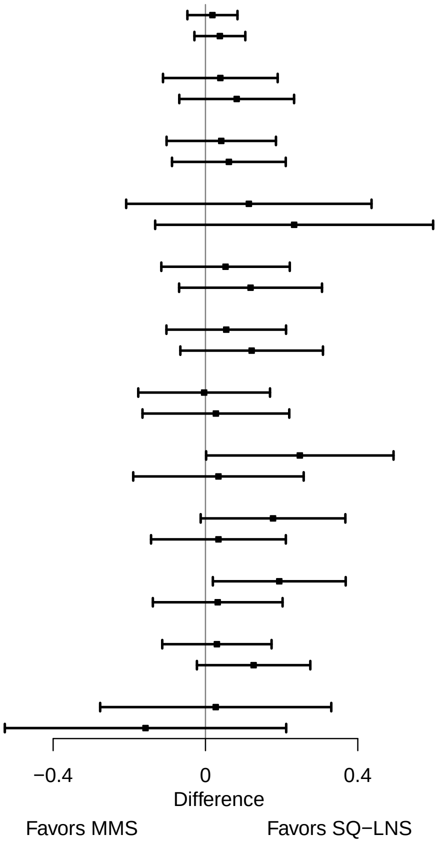

**Supplemental figure 6M: Household food security**

#### 6M2: Relative risks for birth outcomes

| Outcome (p−for−interaction) | SQ−LNS N | MMS N | Relative risk (95% CI) |
|---|---|---|---|
| Low birth weight (p=0.539) | | | |
| Mild to secure | 354 | 334 | 0.98 (0.64, 1.49) |
| Moderate to severe | 336 | 354 | 0.81 (0.54, 1.22) |
| Birth weight < 2 kg (p=0.194) | | | |
| Mild to secure | 354 | 334 | 1.62 (0.57, 4.58) |
| Moderate to severe | 336 | 354 | 0.60 (0.18, 2.06) |
| Small−for−gestational age (p=0.515) | | | |
| Mild to secure | 354 | 334 | 1.02 (0.77, 1.36) |
| Moderate to severe | 336 | 354 | 0.92 (0.70, 1.22) |
| Large−for−gestational age (p=0.445) | | | |
| Mild to secure | 354 | 334 | 0.87 (0.36, 2.11) |
| Moderate to severe | 336 | 354 | 1.49 (0.67, 3.30) |
| Newborn stunting (p=0.446) | | | |
| Mild to secure | 307 | 298 | 0.93 (0.57, 1.51) |
| Moderate to severe | 249 | 272 | 1.22 (0.79, 1.90) |
| Low LGAZ (p=0.287) | | | |
| Mild to secure | 307 | 298 | 1.50 (0.84, 2.68) |
| Moderate to severe | 249 | 272 | 1.07 (0.68, 1.69) |
| Low BMIZ (p=0.312) | | | |
| Mild to secure | 307 | 298 | 1.17 (0.66, 2.07) |
| Moderate to severe | 245 | 261 | 0.85 (0.41, 1.78) |
| Low HCZ (p=0.627) | | | |
| Mild to secure | 308 | 299 | 1.16 (0.57, 2.34) |
| Moderate to severe | 248 | 272 | 1.50 (0.69, 3.25) |
| Low HCGAZ (p=0.692) | | | |
| Mild to secure | 308 | 299 | 0.92 (0.28, 3.06) |
| Moderate to severe | 248 | 272 | 0.66 (0.16, 2.74) |
| Preterm birth (p=0.982) | | | |
| Mild to secure | 390 | 365 | 1.27 (0.78, 2.06) |
| Moderate to severe | 371 | 393 | 1.15 (0.74, 1.79) |

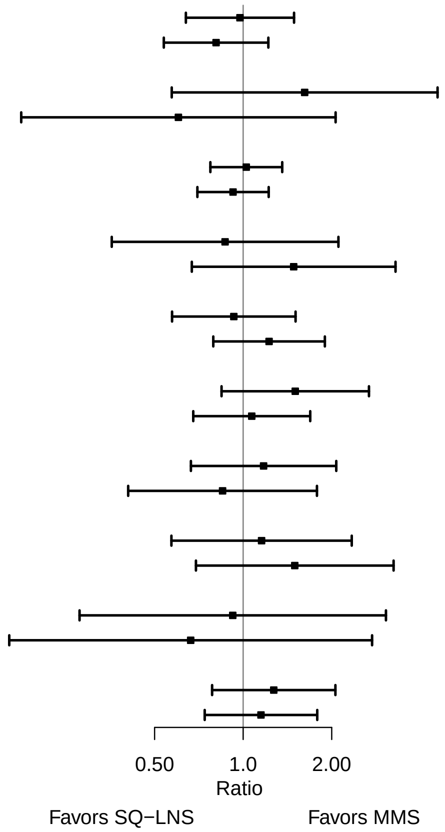

**Supplemental figure 6M: Household food security**

#### 6M3: Mean differences for 6 mo outcomes

| Outcome (p−for−interaction) | SQ−LNS N | MMS N | Mean difference (95% CI) |
|---|---|---|---|
| Weight−for−age z score (p=0.628) | | | |
| Mild to secure | 319 | 314 | 0.02 (−0.15, 0.20) |
| Moderate to severe | 309 | 339 | −0.05 (−0.23, 0.13) |
| Length−for−age z−score (p=0.803) | | | |
| Mild to secure | 319 | 312 | 0.05 (−0.11, 0.21) |
| Moderate to severe | 310 | 338 | 0.00 (−0.17, 0.17) |
| Weight−for−length z−score (p=0.566) | | | |
| Mild to secure | 319 | 312 | −0.01 (−0.18, 0.17) |
| Moderate to severe | 309 | 338 | −0.08 (−0.25, 0.10) |
| Head circumference−for−age z−score (p=0.243) | | | |
| Mild to secure | 319 | 314 | 0.05 (−0.10, 0.20) |
| Moderate to severe | 310 | 339 | −0.10 (−0.25, 0.05) |
| MUAC−for−age z−score (p=0.551) | | | |
| Mild to secure | 319 | 314 | 0.00 (−0.16, 0.16) |
| Moderate to severe | 310 | 339 | −0.04 (−0.22, 0.13) |

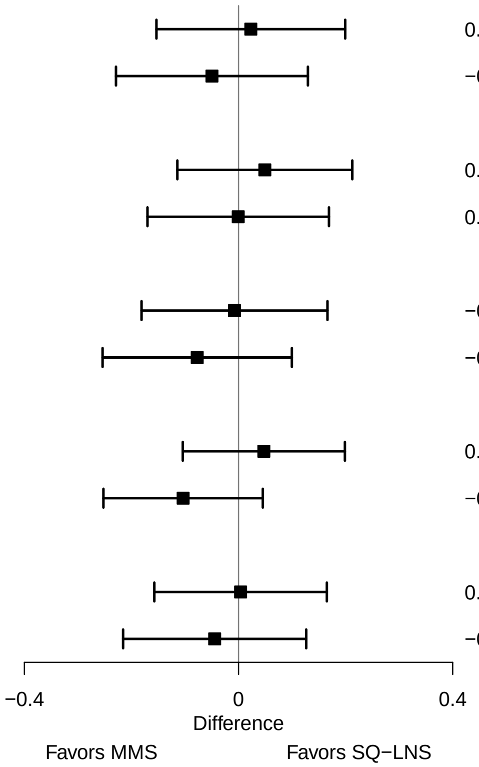

**Supplemental figure 6M: Household food security**

#### 6M4: Prevalence ratios for 6 mo outcomes

| Outcome (p−for−interaction) | SQ−LNS N | MMS N | Prevalence ratio (95% CI) |
|---|---|---|---|
| Underweight (p=0.859) | | | |
| Mild to secure | 319 | 314 | 1.16 (0.67, 2.01) |
| Moderate to severe | 309 | 339 | 1.14 (0.69, 1.89) |
| Stunted (p=0.327) | | | |
| Mild to secure | 319 | 312 | 1.25 (0.85, 1.85) |
| Moderate to severe | 310 | 338 | 0.94 (0.68, 1.28) |
| Wasted (p=0.644) | | | |
| Mild to secure | 319 | 312 | 1.03 (0.31, 3.42) |
| Moderate to severe | 309 | 338 | 1.09 (0.44, 2.71) |
| Low HCAZ (p=0.253) | | | |
| Mild to secure | 319 | 314 | 0.63 (0.37, 1.06) |
| Moderate to severe | 310 | 339 | 1.00 (0.55, 1.82) |
| Low MUAC (p=0.921) | | | |
| Mild to secure | 319 | 314 | 0.94 (0.54, 1.66) |
| Moderate to severe | 310 | 339 | 0.88 (0.50, 1.55) |
| Acute malnutrition (p=0.819) | | | |
| Mild to secure | 319 | 312 | 1.02 (0.60, 1.75) |
| Moderate to severe | 309 | 338 | 0.93 (0.54, 1.61) |

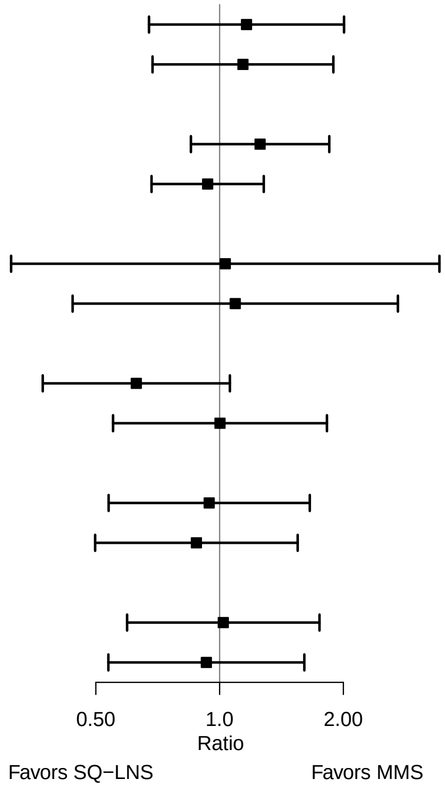
